# Circulating lipid profiles are associated with cross-sectional and longitudinal changes of central biomarkers for Alzheimer’s disease

**DOI:** 10.1101/2023.06.12.23291054

**Authors:** Jun Pyo Kim, Kwangsik Nho, Tingting Wang, Kevin Huynh, Matthias Arnold, Shannon L. Risacher, Paula J. Bice, Xianlin Han, Bruce S. Kristal, Colette Blach, Rebecca Baillie, Gabi Kastenmüller, Peter J. Meikle, Andrew J. Saykin, Rima Kaddurah-Daouk, Alzheimer’s Disease Neuroimaging Initiative, Alzheimer’s Disease Metabolomics Consortium

**Author notes:** Equal corresponding Authors: Rima Kaddurah-Daouk, Ph.D. Duke University Medical Center Durham, NC, USA, and Andrew J Saykin, Psy.D. Indiana University School of Medicine Indianapolis, IN, USA. Equal first author (Jun Pyo Kim, Kwangsik Nho). Data used in preparation of this article were obtained from the Alzheimer’s Disease Neuroimaging Initiative (ADNI) database (adni.loni.usc.edu). As such, the investigators within the ADNI contributed to the design and implementation of ADNI and/or provided data but did not participate in analysis or writing of this report. A complete listing of ADNI investigators can be found at: http://adni.loni.usc.edu/wp-content/uploads/how_to_apply/ADNI_Acknowledgement_List.pdf. Data used in preparation of this article were generated by the Alzheimer’s Disease Metabolomics Consortium (ADMC), a part of the Accelerating Medicines Partnership for Alzheimer’s Disease (AMP-AD). A complete listing of ADMC investigators can be found at: https://sites.duke.edu/adnimetab/team/.

## Abstract

Investigating the association of lipidome profiles with central Alzheimer’s disease (AD) biomarkers, including amyloid/tau/neurodegeneration (A/T/N), can provide a holistic view between the lipidome and AD. We performed cross-sectional and longitudinal association analysis of serum lipidome profiles with AD biomarkers in the Alzheimer’s Disease Neuroimaging Initiative cohort (N=1,395). We identified lipid species, classes, and network modules that were significantly associated with cross-sectional and longitudinal changes of A/T/N biomarkers for AD. Notably, we identified the lysoalkylphosphatidylcholine (LPC(O)) as associated with “A/N” biomarkers at baseline at lipid species, class, and module levels. Also, G_M3_ ganglioside showed significant association with baseline levels and longitudinal changes of the “N” biomarkers at species and class levels. Our study of circulating lipids and central AD biomarkers enabled identification of lipids that play potential roles in the cascade of AD pathogenesis. Our results suggest dysregulation of lipid metabolic pathways as precursors to AD development and progression.

## INTRODUCTION

Alzheimer’s disease (AD) is a neurodegenerative disorder characterized by a cascade of pathological processes, from the accumulation of misfolded proteins such as β-amyloid (Aβ) and hyperphosphorylated tau (p-Tau) to the eventual development of neurodegeneration.^1^ Despite decades of research efforts, a significant portion of AD pathogenesis remains uncertain. To understand the complex nature of AD, multiple levels of omics characteristics of the disease have been investigated,^2^ lipidomics being one of them. Lipidomics is the systems-level analysis of lipids and factors that interact with lipids.^3^ The involvement of lipids in AD pathogenesis has been suggested in many previous studies. In particular, alterations of phospholipid, plasmalogens, ceramide, ganglioside, and sulfatide levels in the brain have been observed.^4–10^ Also, several recent studies have demonstrated alterations of blood lipidome profiles in AD.^11–15^ Furthermore, in a recent lipidomics study using two large cohorts, a total of 218 lipid species were identified as associated with prevalent or incident AD.^16^

Advances in AD biomarker research have led to the shift of AD diagnosis from clinical syndrome to biological process. AD-related biomarkers can be grouped into those of β-amyloid, hyperphosphorylated tau, and neurodegeneration (A/T/N).^17^ In this regard, the concept of an A/T/N classification system was included in the 2018 National Institute of Aging and Alzheimer’s Association Research Framework^18^ and has been widely used in AD research since then. A few existing studies examining the associations between A/T/N biomarkers for AD and blood lipidome have employed techniques that have limited resolution of lipid species^19–21^ and have focused primarily on cross-sectional associations.

Here, we investigated associations of circulating lipidome with cross-sectional and longitudinal central A/T/N biomarkers for AD using a recently developed lipidomics platform covering 749 lipid species across 46 classes that focuses on lipid and lipid-like compounds utilizing chromatographic separation and quantitation. We examined the main effect and interactions of sex and *APOE* ε4 carrier status on circulating lipids at the lipid species and lipid class levels. Finally, we identified network modules of correlated lipids and performed association analyses between the modules and cross-sectional and longitudinal A/T/N biomarkers for AD.

## METHODS

### 1. Participants

Data used in the preparation of this article were obtained from the Alzheimer’s Disease Neuroimaging Initiative (ADNI) database (adni.loni.usc.edu). The ADNI was launched in 2003 as a public-private partnership, led by Principal Investigator Michael W. Weiner, MD. The primary goal of ADNI has been to test whether serial magnetic resonance imaging (MRI), positron emission tomography (PET), other biological markers, and clinical and neuropsychological assessment can be combined to measure the progression of mild cognitive impairment (MCI) and early Alzheimer’s disease (AD). Inclusion and exclusion criteria, clinical and neuroimaging protocols, and other information about ADNI can be found at www.adni-info.org. Demographic information, apolipoprotein E (APOE) and clinical information are available and were downloaded from the ADNI data repository (www.loni.usc.edu/ADNI/). Written informed consent was obtained according to the Declaration of Helsinki at the time of enrollment, and consent forms were approved by each participating sites’ Institutional Review Board.

### 2. Amyloid/Tau/Neurodegeneration biomarkers for AD

Baseline and longitudinal CSF biomarker, FDG PET scan, and MRI scan data were downloaded from the LONI database (https://adni.loni.usc.edu/). For CSF biomarker data, we used the data set generated using the validated and highly automated Roche Elecsys electrochemiluminescence immunoassays.^22^ CSF p-tau values were additionally log-transformed. For FDG-PET phenotypes, we used an ROI-based measure of average uptake across the left and right temporal regions derived from preprocessed scans (co-registered, averaged, standardized image and voxel size, uniform resolution) and intensity-normalized using the pons/vermis region to obtain standard uptake value ratio means.^23^ T1-weighted brain MRI scans were downloaded from the ADNI database. As detailed in previous studies,^24^ FreeSurfer software was used to process T1-weighted brain MRI scans and extract region of interest (ROI)-based imaging phenotypes. We used a global cortical amyloid deposition measured from amyloid PET scans as biomarkers of β-amyloid (“A”). We used CSF phosphorylated tau (p-tau) levels as the biomarker of fibrillary tau (“T”). Both hippocampal volume and temporal lobar FDG uptake were used as a biomarker of neurodegeneration (“N”).

### 3. Measurement of circulating lipid profiles

We acquired lipidomic profiles (749 lipid species from 46 lipid classes, Supplementary Table 1.) of all plasma samples using our recently expanded, targeted lipidomic profiling strategy based on reverse phase liquid chromatography coupled to an Agilent 6495C QqQ mass spectrometer. In terms of the lipid extraction and LC-MS/MS methodology, we used scheduled multiple reaction monitoring (MRM), as previously described^25^, with the addition of approximately 200 novel lipid species from 17 lipid classes^26^. Further details about our latest lipid profiling methodology are described on our laboratory website (https://metabolomics.baker.edu.au/method/).

### 4. Statistical analysis

We used linear regression models to examine the associations of individual lipids, lipid classes, and lipid modules with A/T/N biomarkers at baseline. For longitudinal analysis of A/T/N biomarkers, we used linear mixed effects models with a random intercept and slope. For class-level analysis, we used the first principal component of each pre-defined class (Supplementary Table 1) to represent the class. The principal component was obtained using the psych R package. To detect network-based correlation modules in lipidome data, we used the WGCNA R package that utilizes the hierarchical clustering and dynamic tree cut algorithm. The biweight midcorrelation method was used to calculate the correlation between lipids with a soft-thresholding power of 7. The minimum number of lipids within modules was set to 5. The levels of lipids in a module were represented by the module eigen value (ME), which is defined as the first principal component of the lipid matrix of the corresponding module. We examined the interactions of sex and *APOE* ε4 carrier status on circulating lipids, lipid classes and lipid molecules showing significant associations.

All linear models and linear mixed effects models were adjusted for age, sex, the number of *APOE* L4 alleles, body mass index (BMI), fasting status, levels of clinical lipids (triglyceride, HDL cholesterol, total cholesterol), statin use, and omega-3 use. For association analysis with hippocampal volume, educational attainment, intracranial volume, and magnetic field strength were additionally included as covariates. Taking multiple testing into account, all *p*-values were adjusted using a false discovery rate (FDR) correction with the Benjamini-Hochberg procedure.

For whole brain imaging analysis, the processed amyloid PET and FDG PET images were used to perform a voxel-wise statistical analysis of the effect of lipid network modules on brain amyloid-β deposition and brain glucose metabolism, respectively, across the whole brain using SPM12 (www.fil.ion.ucl.ac.uk/spm/). For surface-based whole brain analysis, the SurfStat software package (www.math.mcgill.ca/keith/surfstat/) was used to perform a multivariable analysis of cortical thickness to examine the effect of lipid network modules on brain structural atrophy on a vertex-by-vertex basis using a general linear model (GLM) approach.^27^ We performed a multivariable regression analysis using the same covariates included in the linear models for cross-sectional analysis. For cortical thickness, MRI magnetic field strength and intracranial volume (ICV) were added as additional covariates. In the voxel-wise whole brain analysis, the significant statistical parameters were selected to correspond to a threshold of *p* < 0.05 (FDR-corrected). In the surface-based whole brain analysis, an adjustment for multiple comparisons was performed using the random field theory correction method with p<0.05 adjusted as the level for significance.^28, 29^

## RESULTS

### 1. Study sample

A total of 1,395 individuals were included in the analysis. The characteristics of individuals used in the cross-sectional and longitudinal analysis are shown in **Table 1**.

**Table 1.**
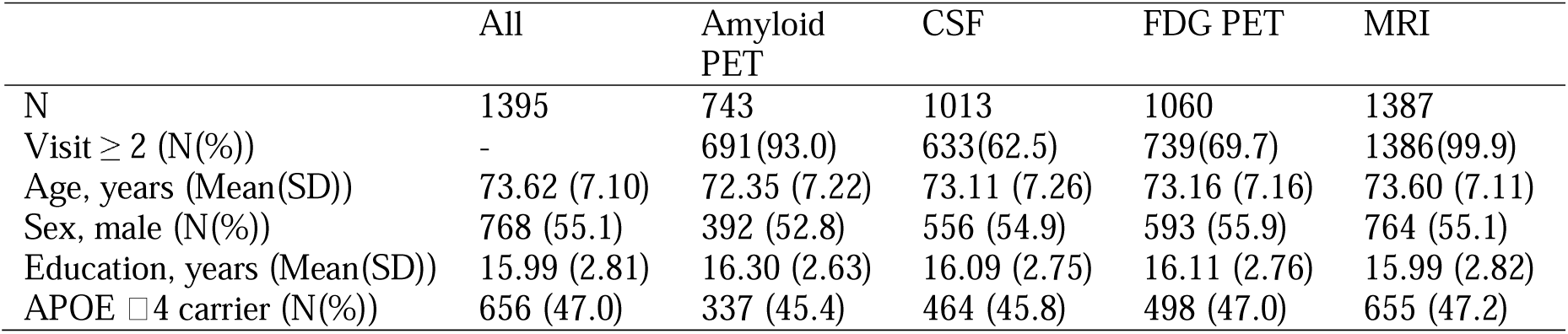
Clinical characteristics. PET positron emission tomography, CSF cerebrospinal fluid, FDG fluorodeoxyglycose, MRI magnetic resonance imaging, SD standard deviation, APOE Apolipoprotein E

### 2. Cross-sectional association analysis between lipids and A/T/N biomarkers at baseline

#### 1) Individual lipid species associated with A/T/N biomarkers at baseline

The results of cross-sectional analysis at baseline between lipid levels and A/T/N biomarkers for AD after FDR correction are shown in **Fig. 1 (Supplementary Table 2)**. A total of nine lipids across four classes (sphingomyelin [SM], lysophosphatidylcholine [LPC], alkylphosphatidylethanolamine [PE(O)], and lysoalkylphosphatidylcholine [LPC(O)]) were identified as significantly associated with the “A” biomarker. Also, 36 lipids across 12 classes (SM, PC, alkylphosphatidylcholine [PC(O)], alkenylphosphatidylcholine [PC(P)], LPC, phosphatidylethanolamine [PE], PE(O), alkenylphosphatidylethanolamines [PE(P)], phosphatidylinositol [PI], dehydrocholesteryl ester [DE], diacylglycerol [DG], and triacylglycerol [TG]) showed significant associations with the “T” biomarker. For the “N” biomarker, 94 lipids across 26 classes were significantly associated with hippocampal volume or brain glucose metabolism. The classes included Cer(d), trihexosylceramide [Hex3Cer], G_M3_ gangliosides [GM3], sulfatide [SHexCer], SM, PC, PC(O), PC(P), LPC, LPC(O), lysoalkenylphosphatidylcholine [LPC(P)], PE(O), PE(P), LPE, PI, lysophosphatidylinositol [LPI], CE, dehydrodesmosteryl ester [deDE], dimethyl-cholesteryl ester [dimethyl-CE], free fatty acid [FFA], acylcarnitine [AC], hydroxylated acylcarnitine [AC-OH], DG, TG, alkyldiacylglycerol [TG(O)], and oxidized lipids.

**Figure 1.**
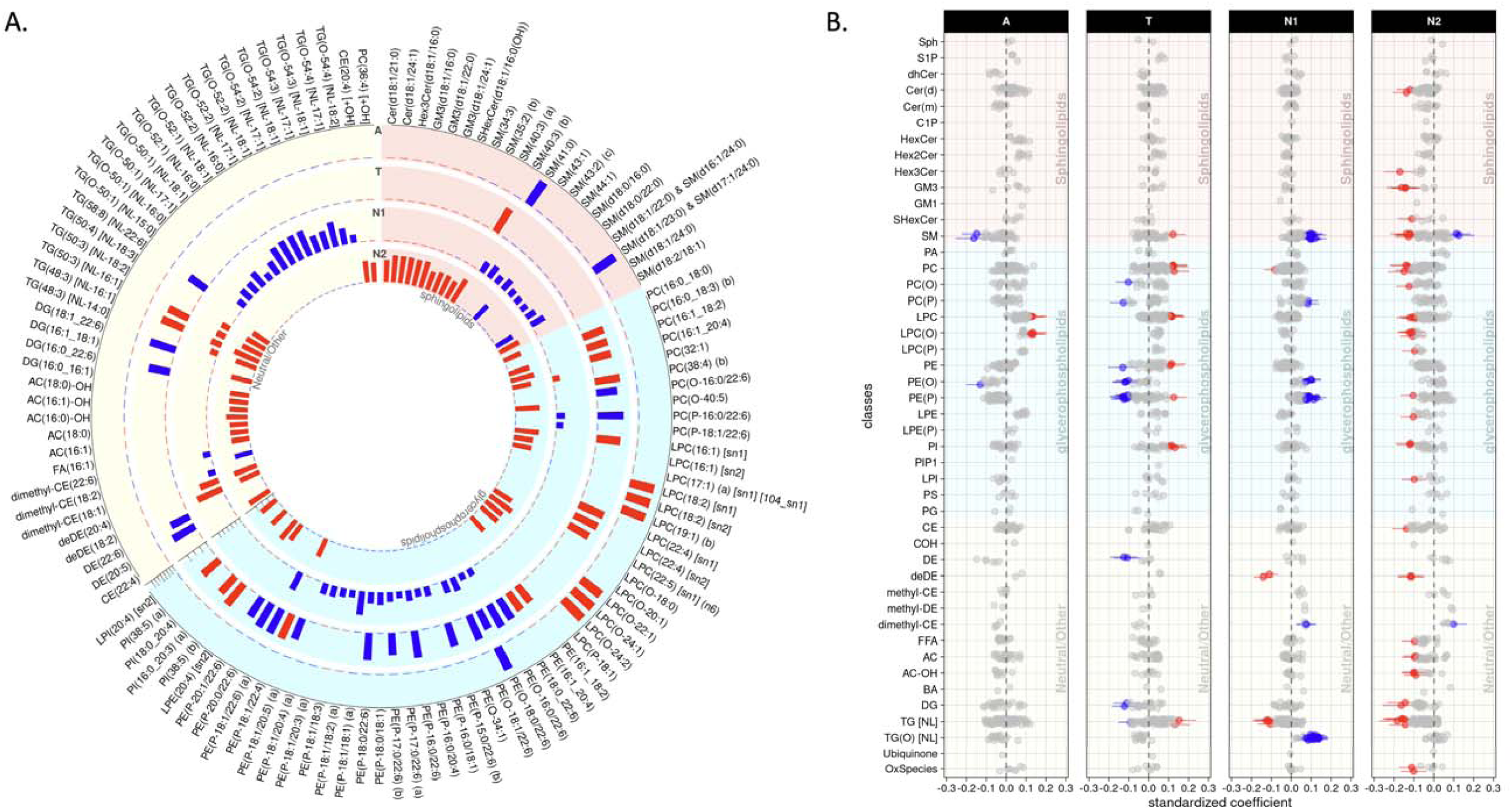
Lipid species levels: association results of lipid species with A/T/N biomarkers at baseline. **(A)** The bar plots from the outermost to innermost circles showed association results between lipid species and A(Amyloid PET)/T(CSF pTau)/N1(MRI)/N2(FDG PET) biomarkers at baseline, respectively. The height of the bars represents the log-transformed FDR corrected *p* values. **(B)** The forest plots showed lipid species-wise cross-sectional association results arranged by lipid classes. Red colors represent that higher levels of the red colored lipid species were significantly associated with worse AD biomarkers, and blue colors represent that lower levels of the blue colored lipid species were significantly associated with worse AD biomarkers.

We identified a significant interaction of *APOE* L4-carrier status with LPC(O-24:1) levels in association with the “A” biomarker after FDR correction (β_interaction_(95% confidence interval[CI]) 0.134(0.065 – 0.202), p_FDR_ = 0.034). LPC(O-24:1) showed a significant association with the “A” biomarker in the *APOE* L4-carrier group (β_interaction_(95%CI) 0.242(0.133 – 0.352), p_FDR_ = 0.007), but the association was not significant in the *APOE* L4 non-carriers (β_interaction_(95%CI) 0.059(−0.045 – 0.164), p_FDR_ = 0.924). However, we did not identify any significant interactions of sex with lipids in the association with A/T/N biomarkers.

#### 2) Lipid classes associated with A/T/N biomarkers at baseline

In the class-level association analysis, six classes (Hex2Cer, LPC, LPC(O), LPC(P), LPE, and DE) were identified as significantly associated with the “A” biomarker (**Fig. 2, Supplementary Table 3**). For the “T” biomarker, none of the classes showed any significant associations at the class level. For the “N” biomarker, five classes (PE(P), deDE, methyl-DE, dimethyl-CE, and TG(O)) were significantly associated with hippocampal volume, and three classes (GM3, LPC(O), and deDE) were associated with brain glucose metabolism. Although deDE level was significantly associated with N biomarkers, it needs to be noted that levels of deDE species were strongly associated with the use of acetylcholinesterase inhibitors, which are commonly used as treatment in patients with dementia. Lipid classes did not show any significant interactions with sex or *APOE* ε4-carrier status in the association with A/T/N biomarkers for AD.

**Figure 2.**
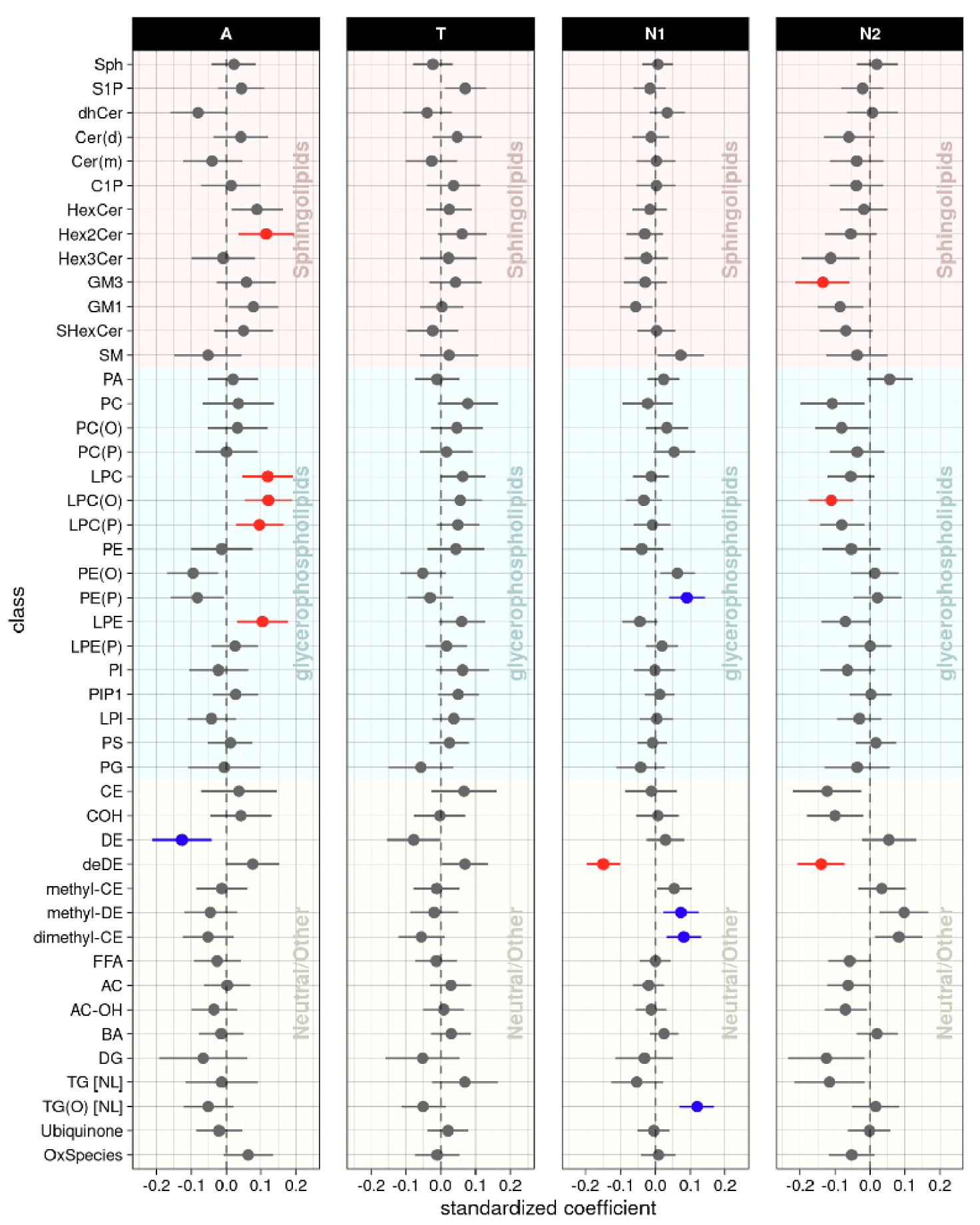
Lipid class levels: association results of lipid classes with A/T/N biomarkers at baseline. The forest plots showed cross-sectional association results between lipid classes and A(Amyloid PET)/T(CSF pTau)/N1(MRI)/N2(FDG PET) biomarkers at baseline after FDR correction. Red colors represent that higher levels of the red colored lipid classes were significantly associated with worse AD biomarkers, and blue colors represent that lower levels of the blue colored lipid classes were significantly associated with worse AD biomarkers.

#### 3) Lipid network modules associated with A/T/N biomarkers at baseline

Lipid correlation network analysis identified 46 network modules, and the number of lipids in each module ranged from 5 to 62 (excluding the M0 module, which comprises unassigned lipids (**Supplementary Table 4**). Among these 46 modules, the M39 (lyso-ether lipids), M34 (PUFA-containing lysoglycerophospholipids dominant), M38 (glycosphingolipids (Hex2Cer)), and M3 (LPCs containing odd/branched chain fatty acid) modules were identified as significantly associated with the “A” biomarker, and the M21 (glycerophospholipids with omega-6 fatty acids) module showed a significant association with the “T” biomarker (**Fig. 3, Supplementary Table 5**). For the “N” biomarker, the M39 (lyso-ether lipids) module was significantly associated with brain glucose metabolism and five modules (M18 (cholesteryl/dehydrocholesteryl esters), M16 (TG(O)s), M15 (ethanolamine ether lipids), M12 (methyl sterol esters), M9 (SM dominant), and M8 (phosphatidylethanolamine plasmalogens containing arachidonic acid)) were significantly associated with hippocampal volume.

**Figure 3.**
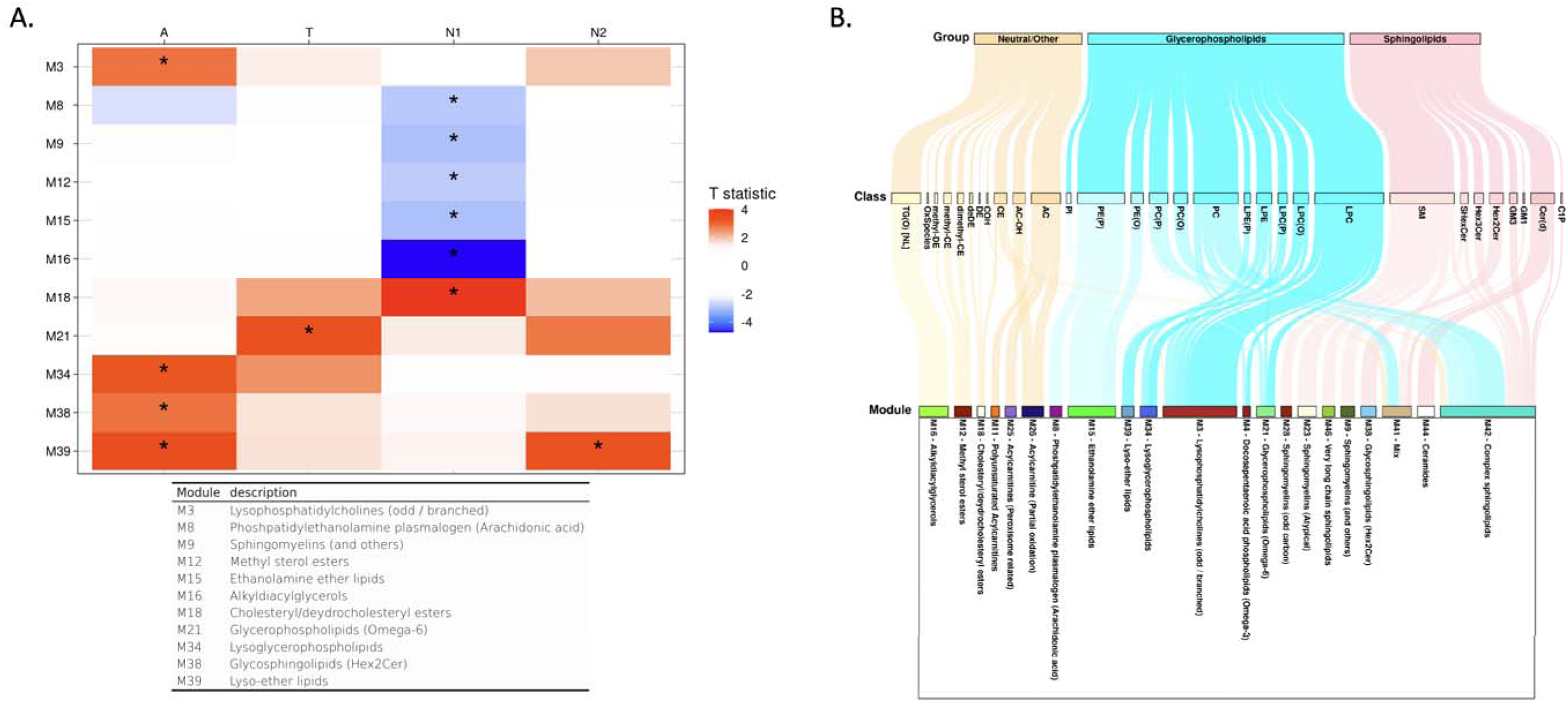
Lipid network module levels: association results of lipid network modules with A/T/N biomarkers at baseline. (A) The tabl showed association results between lipid network modules and A/T/N biomarkers at baseline. Asterisks indicate significant associations after FDR correction. Red colors indicate that higher ME values of the red-colored module were associated with worse AD biomarkers, and blue colors indicate that lower ME values of the blue-colored module were significantly associated with worse AD biomarkers. T statistic values were derived from linear regression analysis and positive T statistic values indicate that higher ME values are associated with worse AD biomarkers. (B) A Sankey diagram was used to visualize clustering of lipid species in the lipid network modules identified as significantly associated with cross-sectional or longitudinal changes of A/T/N biomarkers.

The M39 (lyso-ether lipids) module displayed a significant interaction with *APOE* ε4-carrier status in the association with the “A” biomarker after FDR correction (β_interaction_(95%CI) 0.203(0.077 – 0.328), p_FDR_ = 0.006). The M39 (lyso-ether lipids) module was significantly associated with the “A” biomarker in the *APOE* ε4-carrier group(β_interaction_(95%CI) 0.242(0.130 – 0.354), p_FDR_ = 0.001), but the association was not significant in the *APOE* ε4-non-carrier group (β_interaction_(95%CI) 0.038(−0.068 – 0.143), p_FDR_ = 0.879). No significant interactions of sex with network modules were identified in the association with A/T/N biomarkers.

In addition, we performed detailed whole-brain association analysis to determine the effect of lipid correlation network modules on brain amyloid-β deposition and brain glucose metabolism on a voxel-wise level and brain structural atrophy on a vertex-wise level (**Fig. 4**). For amyloid-β deposition, we identified significant associations of four modules (M39 (lyso-ether lipids), M34 (PUFA-containing lysoglycerophospholipids dominant), M38 (Hex2Cer), and M3 (LPCs containing odd/branched chain fatty acid)) with amyloid-β deposition (**Fig 4A**).

**Figure 4.**
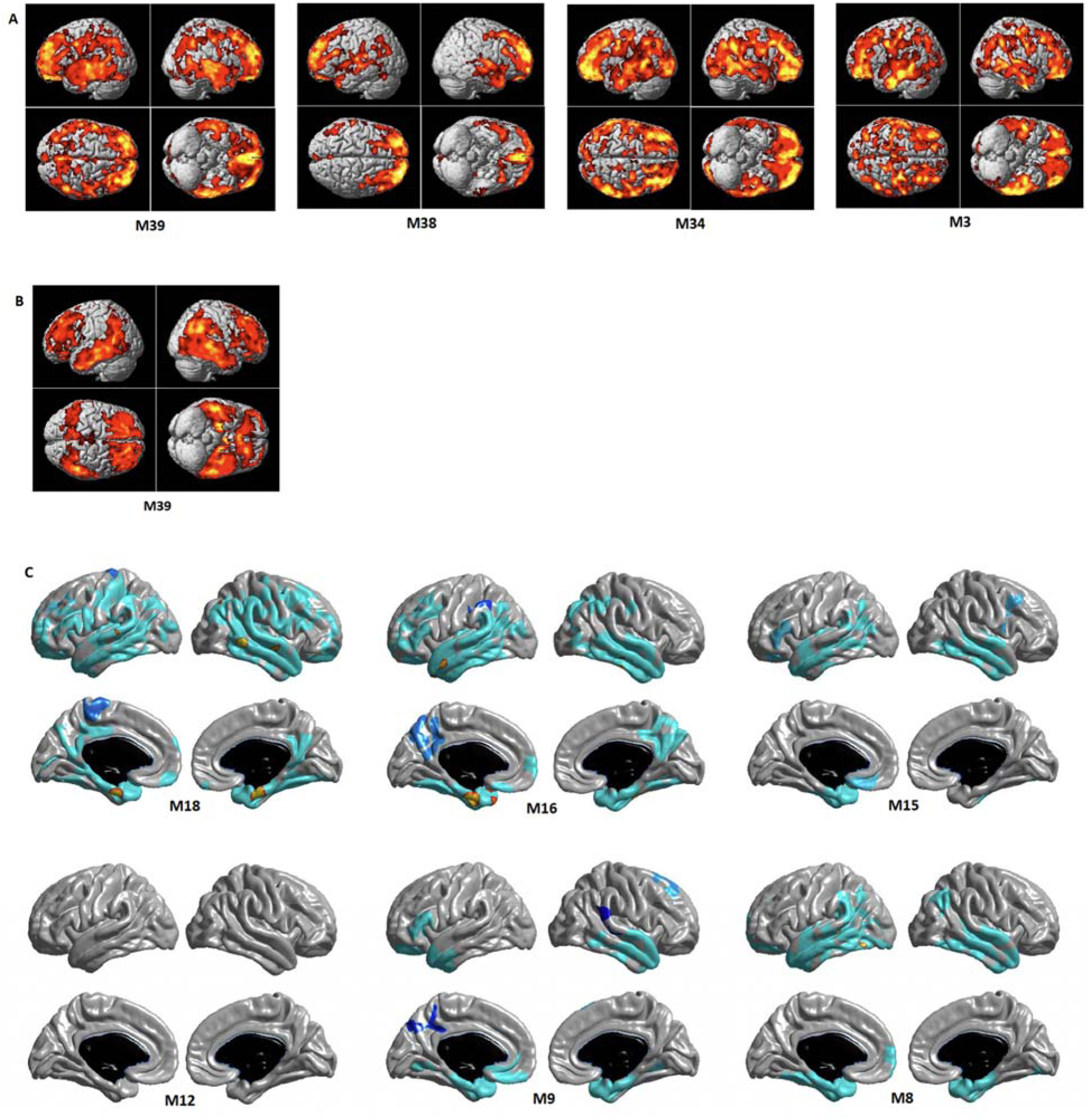
Detailed whole brain association analysis for lipid correlation network module levels: association results of network modules with neuroimaging (PET and MRI) biomarkers for AD at baseline for (A) brain amyloid deposition using Amyloid PET scans, (B) brain glucose metabolism using FDG PET scans, and (C) brain structural atrophy using MRI scans. In a voxel-based whole brain analysis (A) and (B), colored regions represented significant associations (cluster wise threshold of FDR-corrected *p* < 0.05). In a surface-based whole brain analysis (C), statistical maps were thresholded using a random field theory for a multiple testing adjustment to a corrected significance level of 0.05. The *p-*value for clusters indicates significant *p* values with the lightest blue color.

Higher ME values of the four modules were significantly associated with increased amyloid-β deposition in a widespread pattern, especially in the bilateral frontal, parietal, and temporal lobes. For brain glucose metabolism, we identified significant associations for the M39 (lyso-ether lipids) module. Higher ME values of the M39 module were significantly associated with reduced brain glucose metabolism in a widespread pattern, especially in the bilateral frontal, parietal, and temporal lobes including the hippocampus (**Fig 4B**). For brain structural atrophy, the surface-based whole brain analysis identified significant associations of five modules (M18 (cholesteryl/dehydrocholesteryl esters), M16 (TG(O)s), M15 (ethanolamine ether lipids), M9 (SM dominant), and M8 (phosphatidylethanolamine plasmalogens containing arachidonic acid)) with cortical thickness thinning (**Fig 4C**). Lower ME values of the M8, M9, and M15 modules were significantly associated with decreased cortical thickness in the bilateral temporal lobe including the entorhinal cortex. Lower ME values of the M16 module were significantly associated with decreased cortical thickness in a widespread pattern, especially in the bilateral parietal and temporal lobes including the entorhinal cortex. In contrast, higher ME values of the M18 module were significantly associated with decreased cortical thickness in a widespread pattern, especially in the bilateral frontal, parietal and temporal lobes including the entorhinal cortex.

### 3. Longitudinal association analysis between lipids at baseline and longitudinal changes of A/T/N biomarkers

#### 1) Individual lipid species associated with longitudinal changes of A/T/N biomarkers

In the association analysis between baseline lipid levels and longitudinal changes of the “A” and “T” biomarkers, lipids showed no significant associations after FDR correction (**Fig. 5, Supplementary Table 6**). However, in terms of “N”biomarkers, 15 lipids across five classes (HexCer, dihexocylceramide [Hex2Cer], GM3, PC, LPC(O), PI, deDE, and AC) showed significant associations with longitudinal changes of hippocampal volume. In addition, a total of 113 lipids across 22 classes (Cer(d), HexCer, Hex2Cer, Hex3Cer, GM1, GM3, SM, PC, PC(O), PC(P), LPC, LPC(O), PE(P), LPE, PI, CE, COH, DE, deDE, AC, AC-OH, and TG) were significantly associated with longitudinal changes of brain glucose metabolism. Of note, among 113 lipids, 26 lipids were also significantly associated with brain glucose metabolism at baseline. The interactions of sex and *APOE* L4 carrier status with lipids were not significant in the association with longitudinal changes of the A/T/N biomarkers.

**Figure 5.**
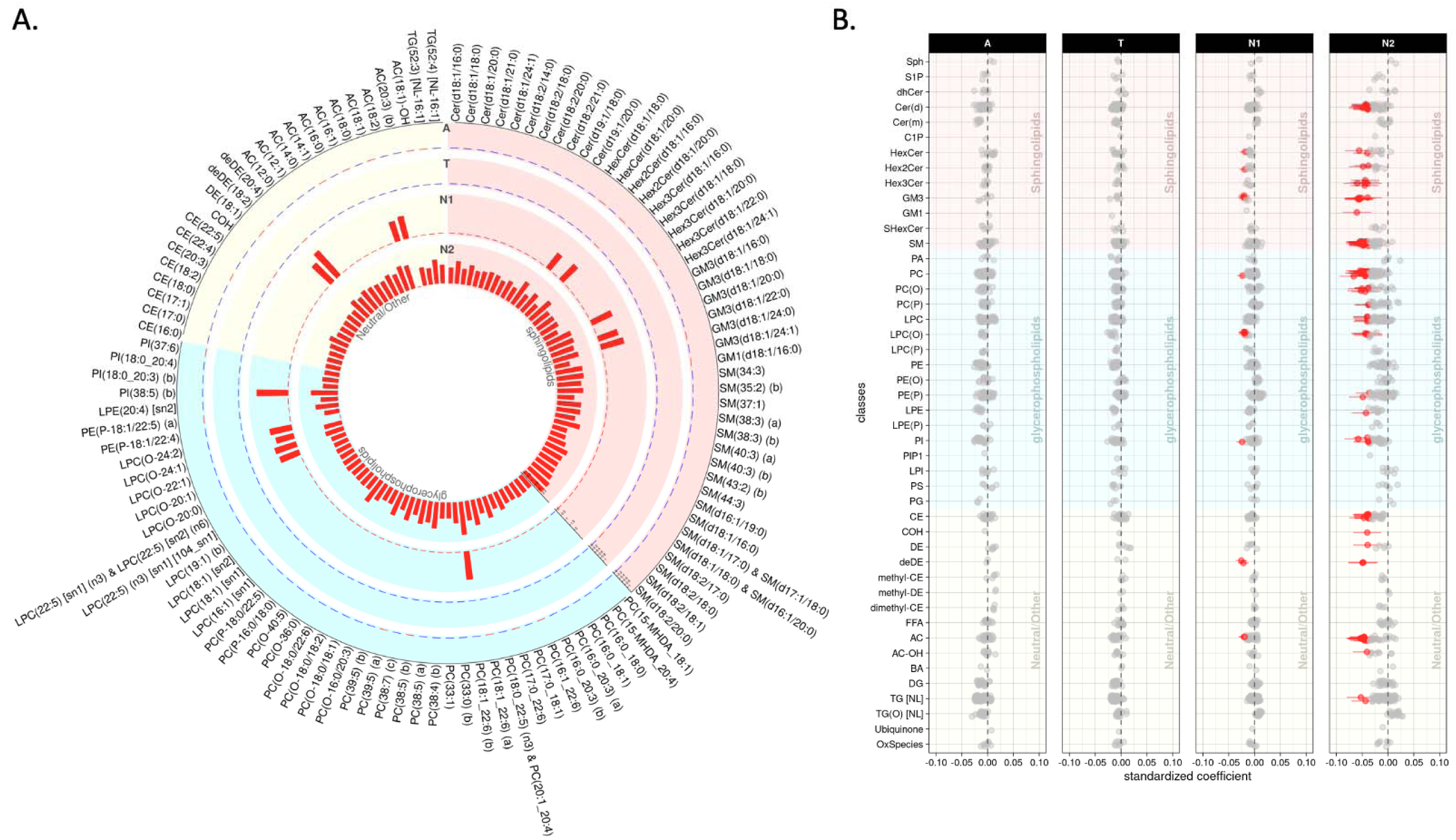
Lipid species levels: association results of lipids with longitudinal change rates of A/T/N biomarkers for AD. **(A)** The bar plots from the outermost to innermost circles showed longitudinal association results between lipid species and longitudinal change rates of A(Amyloid PET)/T(CSF pTau)/N1(MRI)/N2(FDG PET) biomarkers, respectively. The height of the bars represents the log-transformed FDR corrected *p* values. **(B)** The forest plots showed lipid species-wise longitudinal association results arranged by lipid classes. Red colors represent that higher levels of the red colored lipid species were significantly associated with worse progression of AD biomarkers.

#### 2) Lipid classes associated with longitudinal changes of A/T/N biomarkers

In the class-level longitudinal association analysis, none of the classes were significantly associated with the “A” and “T” biomarkers. However, three classes (GM3, LPC(O), and deDE) were significantly associated with longitudinal changes of hippocampal volume, and 12 classes (Hex3Cer, deDE, PC(O), GM3, GM1, CE, SM, LPC(O), COH, PIP1, PC, and AC) were associated with longitudinal changes of brain glucose metabolism. (**Fig. 6, Supplementary Table 7**). The interactions of sex and *APOE* L4-carrier status with lipid classes were not significant in the association with longitudinal changes of the A/T/N biomarkers.

**Figure 6.**
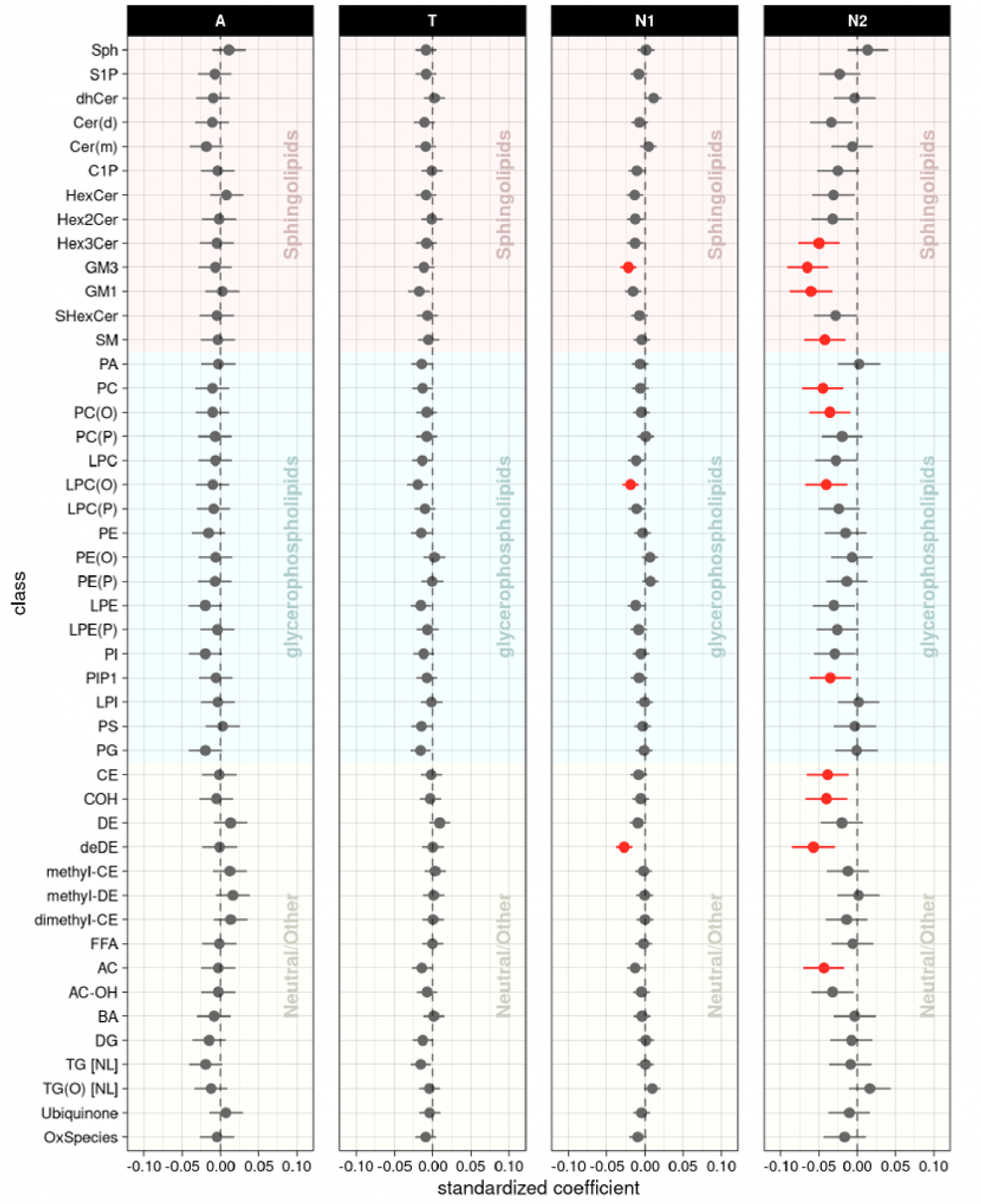
Lipid class levels: longitudinal association results of lipid classes with longitudinal change rates of A/T/N biomarkers. The forest plots showed longitudinal association results between lipid classes and longitudinal change rates of A(Amyloid PET)/T(CSF pTau)/N1(MRI)/N2(FDG PET) biomarkers after FDR correction. Red colors represent that higher levels of the red colored lipid classes were significantly associated with worse progression of AD biomarkers.

#### 3) Lipid network modules associated with longitudinal changes of A/T/N biomarkers

There were no significant associations of lipid network modules with longitudinal changes of the “A” and “T” biomarkers, but 13 modules were significantly associated with longitudinal changes of the “N” biomarkers (**Fig. 7, Supplementary Table 8)**). Three modules (M39 (lyso-ether lipids), M18 (cholesteryl/deshydrocholesteryl esters), and M11 (polyunsaturated ACs)) were associated with longitudinal changes of hippocampal volume, and 12 modules (M26 (ACs (partial oxidation)), M25 (ACs (peroxisome-related)), M44 (Cer(d)s), M18 (cholesteryl/deshydrocholesteryl esters), M42 (Complex sphingolipids), M46 (Very long chain sphingolipids), M23 (Atypical SMs), M38 (glycosphingolipids (HEX2CER)), M28 (odd-numbered SMs), M4 (docosapentaenoic acid phospholipids (Omega-3)), M41(mixed), and M39 (lyso-ether lipids)) were identified as significantly associated with longitudinal changes of brain glucose metabolism. For the 13 modules, higher ME values were associated with faster neurodegeneration. Of note, the M39 (lyso-ether lipids) module was significantly associated with the “N” and “A” biomarkers at baseline, and the M18 module was also significantly associated with the “N” biomarker at baseline. The interactions of sex and *APOE* L4 carrier status with network modules were not significant in association with longitudinal changes of the A/T/N biomarkers.

**Figure 7.**
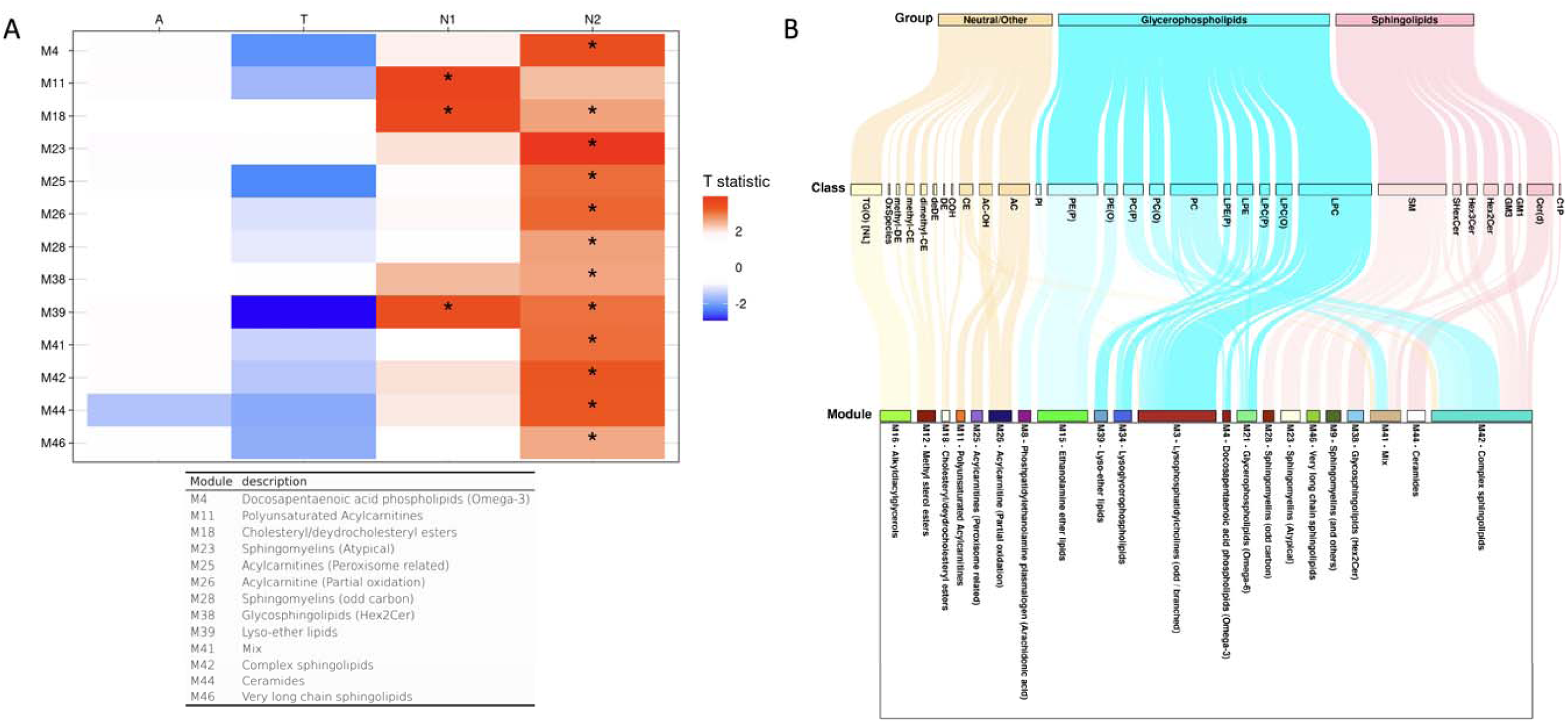
Lipid network module levels: longitudinal association results of lipid network modules with longitudinal change rates of A/T/N biomarkers. (A) The table showed longitudinal association results between lipid network modules and longitudinal change rates of A/T/N biomarkers. Asterisks indicate significant associations after FDR correction. Red colors indicate that higher ME values of the red-colored module were associated with worse progression of AD biomarkers, and blue colors indicate that lower ME values of the blue-colored module were significantly associated with worse progression of AD biomarkers. T statistic values were derived from linear mixed effect analysis and positive T statistic values indicate that higher ME values are associated with worse progression of AD biomarkers. (B) A Sankey diagram was used to visualize clustering of lipid species in the lipid network modules identified as significantly associated with cross-sectional or longitudinal changes of A/T/N biomarkers.

## DISCUSSIONS

Using an up-to-date lipidomics platform capable of separating a larger number of lipids compared to the platforms used in previous studies, we identified lipid species, lipid classes, and lipid correlation network modules as significantly associated with cross-sectional and longitudinal A/T/N biomarkers for AD. The investigation of the relationship between circulating lipids and central AD biomarkers enabled us to identify lipids having potential roles in the cascade of AD pathogenesis. Of note, we identified lipid species, lipid classes, and lipid network modules as associated with cross-sectional and longitudinal changes of multiple AD biomarkers, implying their substantial roles in AD pathogenesis and potential roles of lipid profiles as diagnostic and prognostic biomarkers for AD.

We identified the LPC(O) class as associated with baseline amyloid-β deposition (“A”) and neurodegeneration (“N”) at the individual lipid, lipid class, and lipid correlation network module levels. In particular, higher levels of LPC(O) species were associated with higher cortical Aβ deposition and lower glucose metabolism at baseline. Also, the LPC(O) lipids were significantly associated with longitudinal changes of the “N” biomarkers in all three levels. Similar to the cross-sectional association results, higher levels of LPC(O) were associated with more rapid progression of neurodegeneration. These findings are in line with previous studies reporting increases in brain and blood LPC(O) levels in AD patients,^30,31^and a recent study that showed the association between LPC(O) levels and CSF pTau/A42 ratio.^32^ The overall increase of LPC, LPC(O), LPC(P) in subjects with more severe amyloid biomarker in our baseline analysis indicates phospholipase A2 (PLA2) activation, since PLA2 hydrolyzes PC and PC ether species into LPC or LPC ether species. In fact, studies have shown that Aβ_1-42_ activates PLA2,^33–35^ which can drive neuroinflammation and oxidative stress in the brain.^36, 37^

Another finding was that PE ethers (PE(O), PE(P)) were associated with baseline AD biomarkers in the species level, most of them being associated with more favorable (or less severe) A/T/N biomarker status. Also, in the class and module levels, PE(P) class and two modules related to PE ethers were associated with greater baseline hippocampal volume. However, no significant associations were identified between PE ethers and longitudinal changes of the A/T/N biomarkers in the class or module levels, and only two PE(P) species were associated with faster decline of brain glucose metabolism. PE(P) species (referred to as ethanolamine plasmalogens) are abundant in brain myelin^38^ and are known to be decreased in AD, showing negative association with disease severity and CSF Tau.^39, 40^ Previous studies have shown that PE(P) species function as endogenous antioxidants through their vinyl ether bonds and protect cells against oxidative stress.^41, 42^ The PE(P) species also affect biosynthesis and intracellular transport of cholesterol.^43, 44^ The protective effect of PE(P) against AD may have been contributed by these actions, as oxidative stress and intracellular cholesterol trafficking are both known to have roles in AD.^45, 46^ PE(O), also known as plasmanylethanolamine, is a precursor of PE(P).^47^ Of note, PE(O-18:0/22:6) showed a significant association with all three A/T/N biomarkers at baseline in our analysis. Unlike PE(P), biological functions of the PE(O) species are not well understood. However, a recent study showed that not only PE(P) but also PE(O) species were negatively associated with prevalent and incident AD.^16^ Our results are in line with this study and extend the associations to the AD biomarker level.

It is notable that 20 lipid species across 7 classes out of all 57 species identified as significantly associated with “less severe” AD biomarker status at baseline contained docosahexaenoic acid (DHA). In particular, the associations of those DHA-containing lipids with the T/N biomarkers were prominent. This type of association cannot be detected at the class level since DHA-containing species exist across the classes. At the module level, one of the modules (M45), in which most members contain DHA, showed marginally significant associations with lower CSF pTau (p_FDR_ = 0.053) and larger hippocampal volume (p_FDR_ = 0.054). DHA is mainly distributed throughout pyramidal cell-rich regions (cerebral cortex and hippocampus),^48^ and is involved in multiple pathways related to neuronal biology, from neuronal development to regulation of synaptic function, neuroprotection and modulation of apoptosis.^49, 50^ Studies have shown that individuals with AD have low serum DHA levels, and higher red blood cell DHA is associated with lower risk of incident AD.^51, 52^ In line with these studies, there are studies suggesting protective effect of DHA on incident AD or AD-related cognitive decline. ^53–55^ Our analysis results provide additional evidence of association between DHA and AD on the AD biomarker level. The negative association between DHA and AD biomarkers can be attributed to the anti-inflammatory action of DHA.^56^ Also, it has both a direct and indirect antioxidant effect, which can be a potential protective mechanism of DHA against AD-related damage.^50, 57^

There were other lipids showing significant associations with the “N” biomarkers. For instance, lipid species in the TG(O) and G_M3_ ganglioside classes were significantly associated with baseline “N” biomarkers at the lipid species and lipid class levels. TG(O) was negatively associated with neurodegeneration, while G_M3_ ganglioside was positively associated with neurodegeneration. TG(O) has been identified as having a protective effect on prevalent and incident AD in a recent multicohort study.^16^ Although the direct biological functions of TG(O) are not yet clearly known, TG(O)s are precursors of plasmalogens (PE(P), PC(P)) that play an important role as antioxidants (discussed above).^40, 58^ From a cellular functional perspective, higher levels of peroxisome-derived ether lipids (PC(P), PE(P), PE(O), TG(O)) and related modules (M34, M39) being associated with less neurodegeneration may implicate the role of peroxisomal function in neuroprotection. Peroxisomal alteration in neurodegenerative disease and AD have been suggested in previous studies,^59, 60^ and peroxisome proliferator-activated receptor (PPAR) has been proposed as a potential treatment target and a link between diabetes and AD.^61, 62^

In addition to the significant results in baseline analysis, G_M3_ ganglioside showed significant association with longitudinal changes of the “N” biomarkers at the lipid species and class levels. In line with the cross-sectional analysis results, higher G_M3_ ganglioside levels were associated with more rapid progression of neurodegeneration. The G_M3_ ganglioside is the first ganglioside in the biosynthetic pathway of the major brain gangliosides. In a mouse model study, the reduction of G_M3_ ganglioside by inhibition of glucosylceramide synthase resulted in stabilized remote memory and lower soluble Aβ in the brain.^63^ A previous human study showed that increased levels of the G_M3_ ganglioside species were associated with incident or prevalent AD.^8^ Thus, our findings regarding these classes are in line with these reports, supporting the evidence for their potential roles in AD pathogenesis.

Several limitations in the study should be mentioned. First, our findings are observational, and mechanistic studies are needed to identify causal relationships. Second, our findings need to be replicated in larger independent cohorts to consolidate the findings. Nevertheless, it is noteworthy that we identified lipids as significantly associated with cross-sectional and longitudinal changes of the A/T/N biomarkers in our comprehensive association analysis.

In conclusion, our study investigating the relationship between circulating lipid profiles and central A/T/N biomarkers for AD has implicated several lipid species, lipid classes, and lipid correlation network modules as potential blood-based AD biomarkers that point to dysregulation of specific lipid metabolic pathways as precursors to AD and linked to the progression of the disease. In particular, we identified lipids showing significant associations across multiple A/T/N biomarkers cross-sectionally and longitudinally, including LPC(O) and PE ether species. We also showed that the previously reported beneficial effects of DHA on AD are significant at the biomarker level. Lastly, our findings using AD endophenotypes strengthen evidence from previous studies that were performed using only an AD diagnosis by linking peripheral metabolic changes with brain metabolic and structural states.

## DATA AVAILABILITY

The results published here are in whole or in part based on data obtained from the AD Knowledge Portal and the Alzheimer’s Disease Neuroimaging Initiative (ADNI) database (adni.loni.usc.edu).

## Data Availability

All data produced in the present work are contained in the manuscript.

https://www.loni.usc.edu/ADNI/

## ACKNOWLEDGEMENT

The data available in the AD Knowledge Portal would not be possible without the participation of research volunteers and the contribution of data by collaborating researchers. Metabolomics data as well as data preprocessing, analysis, and interpretation is provided by the Alzheimer’s Disease Metabolomics Consortium (ADMC), part of the Accelerating Medicines Partnership for Alzhiemer’s Disease (AMP-AD), and funded wholly or in part by the following grants and supplements thereto: NIA Alzheimer’s Disease Neuroimaging Initiative (ADNI) database (adni.loni.usc.edu), RF1AG051550, RF1AG057452, R01AG059093, RF1AG058942, U01AG061359, U19AG063744 and FNIH: #DAOU16AMPA awarded to Dr. Kaddurah-Daouk at Duke University in partnership with a large number of academic institutions. A complete listing of ADMC investigators can be found at: https://sites.duke.edu/adnimetab/team/. Data on lipids was generated at Baker Heart and Diabetes Institute, a member of ADMC. Details on the lipid profiling technologies are described at: https://metabolomics.baker.edu.au/method/.

Data collection and sharing for this project was funded by the Alzheimer’s Disease Neuroimaging Initiative (ADNI) (National Institutes of Health Grant U01 AG024904) and DOD ADNI (Department of Defense award number W81XWH-12-2-0012). ADNI is funded by the National Institute on Aging, the National Institute of Biomedical Imaging and Bioengineering, and through generous contributions from the following: AbbVie, Alzheimer’s Association; Alzheimer’s Drug Discovery Foundation; Araclon Biotech; BioClinica, Inc.; Biogen; Bristol-Myers Squibb Company; CereSpir, Inc.; Cogstate; Eisai Inc.; Elan Pharmaceuticals, Inc.; Eli Lilly and Company; EuroImmun; F. Hoffmann-La Roche Ltd and its affiliated company Genentech, Inc.; Fujirebio; GE Healthcare; IXICO Ltd.; Janssen Alzheimer Immunotherapy Research & Development, LLC.; Johnson & Johnson Pharmaceutical Research & Development LLC.; Lumosity; Lundbeck; Merck & Co., Inc.; Meso Scale Diagnostics, LLC.; NeuroRx Research; Neurotrack Technologies; Novartis Pharmaceuticals Corporation; Pfizer Inc.; Piramal Imaging; Servier; Takeda Pharmaceutical Company; and Transition Therapeutics. The Canadian Institutes of Health Research is providing funds to support ADNI clinical sites in Canada. Private sector contributions are facilitated by the Foundation for the National Institutes of Health (www.fnih.org). The grantee organization is the Northern California Institute for Research and Education, and the study is coordinated by the Alzheimer’s Therapeutic Research Institute at the University of Southern California. ADNI data are disseminated by the Laboratory for Neuro Imaging at the University of Southern California.

