## Supplementary Table 1 for "Circulating lipid profiles are associated with cross-sectional and longitudinal changes of central biomarkers for Alzheimer’s disease"

**Supplementary Table 1. Lipid species and classes**

| Lipid Species | Group | Class (Full name) | Class (Abbreviation) |
|---|---|---|---|
| Sph(d18:1) | sphingolipids | Sphingosine | Sph |
| Sph(d18:2) | sphingolipids | Sphingosine | Sph |
| S1P(d16:1) | sphingolipids | Sphingosine-1-phosphate | S1P |
| S1P(d18:0) | sphingolipids | Sphingosine-1-phosphate | S1P |
| S1P(d18:1) | sphingolipids | Sphingosine-1-phosphate | S1P |
| S1P(d18:2) | sphingolipids | Sphingosine-1-phosphate | S1P |
| dhCer(d18:0/16:0) | sphingolipids | Dihydroceramide | dhCer |
| dhCer(d18:0/18:0) | sphingolipids | Dihydroceramide | dhCer |
| dhCer(d18:0/20:0) | sphingolipids | Dihydroceramide | dhCer |
| dhCer(d18:0/22:0) | sphingolipids | Dihydroceramide | dhCer |
| dhCer(d18:0/24:0) | sphingolipids | Dihydroceramide | dhCer |
| dhCer(d18:0/24:1) | sphingolipids | Dihydroceramide | dhCer |
| Cer(d16:1/16:0) | sphingolipids | Ceramide | Cer(d) |
| Cer(d16:1/18:0) | sphingolipids | Ceramide | Cer(d) |
| Cer(d16:1/20:0) | sphingolipids | Ceramide | Cer(d) |
| Cer(d16:1/22:0) | sphingolipids | Ceramide | Cer(d) |
| Cer(d16:1/23:0) | sphingolipids | Ceramide | Cer(d) |
| Cer(d16:1/24:0) | sphingolipids | Ceramide | Cer(d) |
| Cer(d16:1/24:1) | sphingolipids | Ceramide | Cer(d) |
| Cer(d17:1/16:0) | sphingolipids | Ceramide | Cer(d) |
| Cer(d17:1/18:0) | sphingolipids | Ceramide | Cer(d) |
| Cer(d17:1/20:0) | sphingolipids | Ceramide | Cer(d) |
| Cer(d17:1/22:0) | sphingolipids | Ceramide | Cer(d) |
| Cer(d17:1/23:0) | sphingolipids | Ceramide | Cer(d) |
| Cer(d17:1/24:0) | sphingolipids | Ceramide | Cer(d) |
| Cer(d17:1/24:1) | sphingolipids | Ceramide | Cer(d) |
| Cer(d18:1/14:0) | sphingolipids | Ceramide | Cer(d) |
| Cer(d18:1/16:0) | sphingolipids | Ceramide | Cer(d) |
| Cer(d18:1/18:0) | sphingolipids | Ceramide | Cer(d) |
| Cer(d18:1/19:0) | sphingolipids | Ceramide | Cer(d) |
| Cer(d18:1/20:0) | sphingolipids | Ceramide | Cer(d) |
| Cer(d18:1/21:0) | sphingolipids | Ceramide | Cer(d) |
| Cer(d18:1/22:0) | sphingolipids | Ceramide | Cer(d) |
| Cer(d18:1/23:0) | sphingolipids | Ceramide | Cer(d) |
| Cer(d18:1/24:0) | sphingolipids | Ceramide | Cer(d) |
| Cer(d18:1/24:1) | sphingolipids | Ceramide | Cer(d) |
| Cer(d18:1/26:0) | sphingolipids | Ceramide | Cer(d) |
| Cer(d18:2/14:0) | sphingolipids | Ceramide | Cer(d) |
| Cer(d18:2/16:0) | sphingolipids | Ceramide | Cer(d) |
| Cer(d18:2/18:0) | sphingolipids | Ceramide | Cer(d) |
| Cer(d18:2/20:0) | sphingolipids | Ceramide | Cer(d) |
| Cer(d18:2/21:0) | sphingolipids | Ceramide | Cer(d) |
| Cer(d18:2/22:0) | sphingolipids | Ceramide | Cer(d) |
| Cer(d18:2/23:0) | sphingolipids | Ceramide | Cer(d) |
| Cer(d18:2/24:0) | sphingolipids | Ceramide | Cer(d) |
| Cer(d18:2/24:1) | sphingolipids | Ceramide | Cer(d) |
| Cer(d18:2/26:0) | sphingolipids | Ceramide | Cer(d) |
| Cer(d19:1/16:0) | sphingolipids | Ceramide | Cer(d) |
| Cer(d19:1/18:0) | sphingolipids | Ceramide | Cer(d) |
| Cer(d19:1/20:0) | sphingolipids | Ceramide | Cer(d) |
| Cer(d19:1/22:0) | sphingolipids | Ceramide | Cer(d) |
| Cer(d19:1/23:0) | sphingolipids | Ceramide | Cer(d) |
| Cer(d19:1/24:0) | sphingolipids | Ceramide | Cer(d) |
| Cer(d19:1/24:1) | sphingolipids | Ceramide | Cer(d) |

| | | | |
|---|---|---|---|
| Cer(d19:1/26:0) | sphingolipids | Ceramide | Cer(d) |
| Cer(d20:1/22:0) | sphingolipids | Ceramide | Cer(d) |
| Cer(d20:1/23:0) | sphingolipids | Ceramide | Cer(d) |
| Cer(d20:1/24:0) | sphingolipids | Ceramide | Cer(d) |
| Cer(d20:1/24:1) | sphingolipids | Ceramide | Cer(d) |
| Cer(m18:0/20:0) | sphingolipids | Deoxyceramide | Cer(m) |
| Cer(m18:0/22:0) | sphingolipids | Deoxyceramide | Cer(m) |
| Cer(m18:0/23:0) | sphingolipids | Deoxyceramide | Cer(m) |
| Cer(m18:0/24:0) | sphingolipids | Deoxyceramide | Cer(m) |
| Cer(m18:0/24:1) | sphingolipids | Deoxyceramide | Cer(m) |
| Cer(m18:1/18:0) | sphingolipids | Deoxyceramide | Cer(m) |
| Cer(m18:1/20:0) | sphingolipids | Deoxyceramide | Cer(m) |
| Cer(m18:1/22:0) | sphingolipids | Deoxyceramide | Cer(m) |
| Cer(m18:1/23:0) | sphingolipids | Deoxyceramide | Cer(m) |
| Cer(m18:1/24:0) | sphingolipids | Deoxyceramide | Cer(m) |
| Cer(m18:1/24:1) | sphingolipids | Deoxyceramide | Cer(m) |
| Cer1P(d18:1/16:0) | sphingolipids | Ceramide-1-phosphate | C1P |
| HexCer(d16:1/18:0) | sphingolipids | Monohexosylceramide | HexCer |
| HexCer(d16:1/20:0) | sphingolipids | Monohexosylceramide | HexCer |
| HexCer(d16:1/22:0) | sphingolipids | Monohexosylceramide | HexCer |
| HexCer(d16:1/24:0) | sphingolipids | Monohexosylceramide | HexCer |
| HexCer(d18:1/16:0) | sphingolipids | Monohexosylceramide | HexCer |
| HexCer(d18:1/18:0) | sphingolipids | Monohexosylceramide | HexCer |
| HexCer(d18:1/20:0) | sphingolipids | Monohexosylceramide | HexCer |
| HexCer(d18:1/22:0) | sphingolipids | Monohexosylceramide | HexCer |
| HexCer(d18:1/24:0) | sphingolipids | Monohexosylceramide | HexCer |
| HexCer(d18:1/24:1) | sphingolipids | Monohexosylceramide | HexCer |
| HexCer(d18:2/18:0) | sphingolipids | Monohexosylceramide | HexCer |
| HexCer(d18:2/20:0) | sphingolipids | Monohexosylceramide | HexCer |
| HexCer(d18:2/22:0) | sphingolipids | Monohexosylceramide | HexCer |
| HexCer(d18:2/24:0) | sphingolipids | Monohexosylceramide | HexCer |
| Hex2Cer(d16:1/16:0) | sphingolipids | Dihexosylceramide | Hex2Cer |
| Hex2Cer(d16:1/24:1) | sphingolipids | Dihexosylceramide | Hex2Cer |
| Hex2Cer(d18:1/16:0) | sphingolipids | Dihexosylceramide | Hex2Cer |
| Hex2Cer(d18:1/20:0) | sphingolipids | Dihexosylceramide | Hex2Cer |
| Hex2Cer(d18:1/22:0) | sphingolipids | Dihexosylceramide | Hex2Cer |
| Hex2Cer(d18:1/24:0) | sphingolipids | Dihexosylceramide | Hex2Cer |
| Hex2Cer(d18:1/24:1) | sphingolipids | Dihexosylceramide | Hex2Cer |
| Hex2Cer(d18:2/16:0) | sphingolipids | Dihexosylceramide | Hex2Cer |
| Hex2Cer(d18:2/24:1) | sphingolipids | Dihexosylceramide | Hex2Cer |
| Hex3Cer(d18:1/16:0) | sphingolipids | Trihexosylcermide | Hex3Cer |
| Hex3Cer(d18:1/18:0) | sphingolipids | Trihexosylcermide | Hex3Cer |
| Hex3Cer(d18:1/20:0) | sphingolipids | Trihexosylcermide | Hex3Cer |
| Hex3Cer(d18:1/22:0) | sphingolipids | Trihexosylcermide | Hex3Cer |
| Hex3Cer(d18:1/24:0) | sphingolipids | Trihexosylcermide | Hex3Cer |
| Hex3Cer(d18:1/24:1) | sphingolipids | Trihexosylcermide | Hex3Cer |
| GM3(d18:1/16:0) | sphingolipids | GM3 ganglioside | GM3 |
| GM3(d18:1/18:0) | sphingolipids | GM3 ganglioside | GM3 |
| GM3(d18:1/20:0) | sphingolipids | GM3 ganglioside | GM3 |
| GM3(d18:1/22:0) | sphingolipids | GM3 ganglioside | GM3 |
| GM3(d18:1/24:0) | sphingolipids | GM3 ganglioside | GM3 |
| GM3(d18:1/24:1) | sphingolipids | GM3 ganglioside | GM3 |
| GM1(d18:1/16:0) | sphingolipids | GM1 ganglioside | GM1 |
| SHexCer(d18:1/16:0(OH)) | sphingolipids | Sulfatide | SHexCer |
| SHexCer(d18:1/16:0) | sphingolipids | Sulfatide | SHexCer |
| SHexCer(d18:1/24:0(OH)) | sphingolipids | Sulfatide | SHexCer |
| SHexCer(d18:1/24:0) | sphingolipids | Sulfatide | SHexCer |

| | | | |
|---|---|---|---|
| SHexCer(d18:1/24:1(OH)) | sphingolipids | Sulfatide | SHexCer |
| SHexCer(d18:1/24:1) | sphingolipids | Sulfatide | SHexCer |
| SM(34:3) | sphingolipids | Sphingomyelin | SM |
| SM(35:2) (b) | sphingolipids | Sphingomyelin | SM |
| SM(37:1) | sphingolipids | Sphingomyelin | SM |
| SM(37:2) | sphingolipids | Sphingomyelin | SM |
| SM(38:3) (a) | sphingolipids | Sphingomyelin | SM |
| SM(38:3) (b) | sphingolipids | Sphingomyelin | SM |
| SM(40:3) (a) | sphingolipids | Sphingomyelin | SM |
| SM(40:3) (b) | sphingolipids | Sphingomyelin | SM |
| SM(41:0) | sphingolipids | Sphingomyelin | SM |
| SM(41:1) (a) | sphingolipids | Sphingomyelin | SM |
| SM(43:1) | sphingolipids | Sphingomyelin | SM |
| SM(43:2) (b) | sphingolipids | Sphingomyelin | SM |
| SM(43:2) (c) | sphingolipids | Sphingomyelin | SM |
| SM(44:1) | sphingolipids | Sphingomyelin | SM |
| SM(44:2) | sphingolipids | Sphingomyelin | SM |
| SM(44:3) | sphingolipids | Sphingomyelin | SM |
| SM(d16:1/19:0) | sphingolipids | Sphingomyelin | SM |
| SM(d16:1/23:0) & SM(d17:1/22:0) | sphingolipids | Sphingomyelin | SM |
| SM(d16:1/24:1) | sphingolipids | Sphingomyelin | SM |
| SM(d17:1/14:0) | sphingolipids | Sphingomyelin | SM |
| SM(d17:1/16:0) | sphingolipids | Sphingomyelin | SM |
| SM(d17:1/24:1) | sphingolipids | Sphingomyelin | SM |
| SM(d18:0/14:0) | sphingolipids | Sphingomyelin | SM |
| SM(d18:0/16:0) | sphingolipids | Sphingomyelin | SM |
| SM(d18:0/22:0) | sphingolipids | Sphingomyelin | SM |
| SM(d18:1/14:0) & SM(d16:1/16:0) | sphingolipids | Sphingomyelin | SM |
| SM(d18:1/16:0) | sphingolipids | Sphingomyelin | SM |
| SM(d18:1/17:0) & SM(d17:1/18:0) | sphingolipids | Sphingomyelin | SM |
| SM(d18:1/18:0) & SM(d16:1/20:0) | sphingolipids | Sphingomyelin | SM |
| SM(d18:1/20:0) & SM(d16:1/22:0) | sphingolipids | Sphingomyelin | SM |
| SM(d18:1/22:0) & SM(d16:1/24:0) | sphingolipids | Sphingomyelin | SM |
| SM(d18:1/23:0) & SM(d17:1/24:0) | sphingolipids | Sphingomyelin | SM |
| SM(d18:1/24:0) | sphingolipids | Sphingomyelin | SM |
| SM(d18:1/24:1) | sphingolipids | Sphingomyelin | SM |
| SM(d18:2/14:0) | sphingolipids | Sphingomyelin | SM |
| SM(d18:2/16:0) | sphingolipids | Sphingomyelin | SM |
| SM(d18:2/17:0) | sphingolipids | Sphingomyelin | SM |
| SM(d18:2/18:0) | sphingolipids | Sphingomyelin | SM |
| SM(d18:2/18:1) | sphingolipids | Sphingomyelin | SM |
| SM(d18:2/20:0) | sphingolipids | Sphingomyelin | SM |
| SM(d18:2/22:0) | sphingolipids | Sphingomyelin | SM |
| SM(d18:2/23:0) | sphingolipids | Sphingomyelin | SM |
| SM(d18:2/24:0) | sphingolipids | Sphingomyelin | SM |
| SM(d19:1/24:1) | sphingolipids | Sphingomyelin | SM |
| PA(34:1) | glycerophospholipids | Phosphatidic acid | PA |
| PA(36:1) | glycerophospholipids | Phosphatidic acid | PA |
| PA(36:2) | glycerophospholipids | Phosphatidic acid | PA |
| PA(36:3) | glycerophospholipids | Phosphatidic acid | PA |
| PA(36:4) | glycerophospholipids | Phosphatidic acid | PA |
| PA(40:6) | glycerophospholipids | Phosphatidic acid | PA |
| PC(14:0_16:0) | glycerophospholipids | Phosphatidylcholine | PC |
| PC(14:0_20:4) | glycerophospholipids | Phosphatidylcholine | PC |
| PC(14:0_22:6) | glycerophospholipids | Phosphatidylcholine | PC |
| PC(15-MHDA_18:1) | glycerophospholipids | Phosphatidylcholine | PC |
| PC(15-MHDA_18:2) | glycerophospholipids | Phosphatidylcholine | PC |

| | | | |
|---|---|---|---|
| PC(15-MHDA_20:4) | glycerophospholipids | Phosphatidylcholine | PC |
| PC(15-MHDA_22:6) | glycerophospholipids | Phosphatidylcholine | PC |
| PC(15:0_20:3) | glycerophospholipids | Phosphatidylcholine | PC |
| PC(15:0_20:4) | glycerophospholipids | Phosphatidylcholine | PC |
| PC(15:0_22:6) | glycerophospholipids | Phosphatidylcholine | PC |
| PC(16:0_16:0) | glycerophospholipids | Phosphatidylcholine | PC |
| PC(16:0_18:0) | glycerophospholipids | Phosphatidylcholine | PC |
| PC(16:0_18:1) | glycerophospholipids | Phosphatidylcholine | PC |
| PC(16:0_18:2) | glycerophospholipids | Phosphatidylcholine | PC |
| PC(16:0_18:3) (a) | glycerophospholipids | Phosphatidylcholine | PC |
| PC(16:0_18:3) (b) | glycerophospholipids | Phosphatidylcholine | PC |
| PC(16:0_20:3) (a) | glycerophospholipids | Phosphatidylcholine | PC |
| PC(16:0_20:3) (b) | glycerophospholipids | Phosphatidylcholine | PC |
| PC(16:0_20:4) | glycerophospholipids | Phosphatidylcholine | PC |
| PC(16:0_20:5) | glycerophospholipids | Phosphatidylcholine | PC |
| PC(16:0_22:6) | glycerophospholipids | Phosphatidylcholine | PC |
| PC(16:1_18:2) | glycerophospholipids | Phosphatidylcholine | PC |
| PC(16:1_20:4) | glycerophospholipids | Phosphatidylcholine | PC |
| PC(16:1_22:6) | glycerophospholipids | Phosphatidylcholine | PC |
| PC(17:0_18:1) | glycerophospholipids | Phosphatidylcholine | PC |
| PC(17:0_18:2) | glycerophospholipids | Phosphatidylcholine | PC |
| PC(17:0_20:4) | glycerophospholipids | Phosphatidylcholine | PC |
| PC(17:0_22:6) | glycerophospholipids | Phosphatidylcholine | PC |
| PC(17:1_18:2) | glycerophospholipids | Phosphatidylcholine | PC |
| PC(18:0_18:1) | glycerophospholipids | Phosphatidylcholine | PC |
| PC(18:0_18:2) | glycerophospholipids | Phosphatidylcholine | PC |
| PC(18:0_20:3) | glycerophospholipids | Phosphatidylcholine | PC |
| PC(18:0_20:4) | glycerophospholipids | Phosphatidylcholine | PC |
| PC(18:0_22:4) | glycerophospholipids | Phosphatidylcholine | PC |
| PC(18:0_22:5) (n3) & PC(20:1_20:4) | glycerophospholipids | Phosphatidylcholine | PC |
| PC(18:0_22:5) (n6) | glycerophospholipids | Phosphatidylcholine | PC |
| PC(18:0_22:6) | glycerophospholipids | Phosphatidylcholine | PC |
| PC(18:1_18:1) | glycerophospholipids | Phosphatidylcholine | PC |
| PC(18:1_18:2) | glycerophospholipids | Phosphatidylcholine | PC |
| PC(18:1_20:3) | glycerophospholipids | Phosphatidylcholine | PC |
| PC(18:1_22:6) (a) | glycerophospholipids | Phosphatidylcholine | PC |
| PC(18:1_22:6) (b) | glycerophospholipids | Phosphatidylcholine | PC |
| PC(18:2_18:2) | glycerophospholipids | Phosphatidylcholine | PC |
| PC(18:2_20:5) | glycerophospholipids | Phosphatidylcholine | PC |
| PC(20:0_20:4) | glycerophospholipids | Phosphatidylcholine | PC |
| PC(28:0) | glycerophospholipids | Phosphatidylcholine | PC |
| PC(31:0) (a) | glycerophospholipids | Phosphatidylcholine | PC |
| PC(31:0) (b) | glycerophospholipids | Phosphatidylcholine | PC |
| PC(32:1) | glycerophospholipids | Phosphatidylcholine | PC |
| PC(32:2) | glycerophospholipids | Phosphatidylcholine | PC |
| PC(33:0) (a) | glycerophospholipids | Phosphatidylcholine | PC |
| PC(33:0) (b) | glycerophospholipids | Phosphatidylcholine | PC |
| PC(33:1) | glycerophospholipids | Phosphatidylcholine | PC |
| PC(33:2) | glycerophospholipids | Phosphatidylcholine | PC |
| PC(34:5) | glycerophospholipids | Phosphatidylcholine | PC |
| PC(35:5) | glycerophospholipids | Phosphatidylcholine | PC |
| PC(36:6) (a) | glycerophospholipids | Phosphatidylcholine | PC |
| PC(38:2) | glycerophospholipids | Phosphatidylcholine | PC |
| PC(38:4) (b) | glycerophospholipids | Phosphatidylcholine | PC |
| PC(38:5) (a) | glycerophospholipids | Phosphatidylcholine | PC |
| PC(38:5) (b) | glycerophospholipids | Phosphatidylcholine | PC |
| PC(38:6) (a) | glycerophospholipids | Phosphatidylcholine | PC |

| | | | |
|---|---|---|---|
| PC(38:7) (c) | glycerophospholipids | Phosphatidylcholine | PC |
| PC(39:5) (a) | glycerophospholipids | Phosphatidylcholine | PC |
| PC(39:5) (b) | glycerophospholipids | Phosphatidylcholine | PC |
| PC(40:7) (a) | glycerophospholipids | Phosphatidylcholine | PC |
| PC(40:8) | glycerophospholipids | Phosphatidylcholine | PC |
| PC(44:12) | glycerophospholipids | Phosphatidylcholine | PC |
| PC(O-16:0/16:0) | glycerophospholipids | Alkylphosphatidylcholine | PC(O) |
| PC(O-16:0/20:3) | glycerophospholipids | Alkylphosphatidylcholine | PC(O) |
| PC(O-16:0/20:4) | glycerophospholipids | Alkylphosphatidylcholine | PC(O) |
| PC(O-16:0/22:6) | glycerophospholipids | Alkylphosphatidylcholine | PC(O) |
| PC(O-18:0/18:1) | glycerophospholipids | Alkylphosphatidylcholine | PC(O) |
| PC(O-18:0/18:2) | glycerophospholipids | Alkylphosphatidylcholine | PC(O) |
| PC(O-18:0/20:4) | glycerophospholipids | Alkylphosphatidylcholine | PC(O) |
| PC(O-18:0/22:6) | glycerophospholipids | Alkylphosphatidylcholine | PC(O) |
| PC(O-18:1/18:1) | glycerophospholipids | Alkylphosphatidylcholine | PC(O) |
| PC(O-18:1/18:2) | glycerophospholipids | Alkylphosphatidylcholine | PC(O) |
| PC(O-32:1) | glycerophospholipids | Alkylphosphatidylcholine | PC(O) |
| PC(O-32:2) | glycerophospholipids | Alkylphosphatidylcholine | PC(O) |
| PC(O-34:1) | glycerophospholipids | Alkylphosphatidylcholine | PC(O) |
| PC(O-34:2) | glycerophospholipids | Alkylphosphatidylcholine | PC(O) |
| PC(O-34:4) | glycerophospholipids | Alkylphosphatidylcholine | PC(O) |
| PC(O-35:4) | glycerophospholipids | Alkylphosphatidylcholine | PC(O) |
| PC(O-36:0) | glycerophospholipids | Alkylphosphatidylcholine | PC(O) |
| PC(O-36:5) | glycerophospholipids | Alkylphosphatidylcholine | PC(O) |
| PC(O-38:5) | glycerophospholipids | Alkylphosphatidylcholine | PC(O) |
| PC(O-40:5) | glycerophospholipids | Alkylphosphatidylcholine | PC(O) |
| PC(P-15:0/20:4) (a) | glycerophospholipids | Alkenylphosphatidylcholine (plasmalogen) | PC(P) |
| PC(P-15:0/20:4) (b) | glycerophospholipids | Alkenylphosphatidylcholine (plasmalogen) | PC(P) |
| PC(P-16:0/14:0) | glycerophospholipids | Alkenylphosphatidylcholine (plasmalogen) | PC(P) |
| PC(P-16:0/16:0) | glycerophospholipids | Alkenylphosphatidylcholine (plasmalogen) | PC(P) |
| PC(P-16:0/16:1) | glycerophospholipids | Alkenylphosphatidylcholine (plasmalogen) | PC(P) |
| PC(P-16:0/18:0) | glycerophospholipids | Alkenylphosphatidylcholine (plasmalogen) | PC(P) |
| PC(P-16:0/18:1) | glycerophospholipids | Alkenylphosphatidylcholine (plasmalogen) | PC(P) |
| PC(P-16:0/18:2) | glycerophospholipids | Alkenylphosphatidylcholine (plasmalogen) | PC(P) |
| PC(P-16:0/18:3) | glycerophospholipids | Alkenylphosphatidylcholine (plasmalogen) | PC(P) |
| PC(P-16:0/20:4) | glycerophospholipids | Alkenylphosphatidylcholine (plasmalogen) | PC(P) |
| PC(P-16:0/20:5) | glycerophospholipids | Alkenylphosphatidylcholine (plasmalogen) | PC(P) |
| PC(P-16:0/22:6) | glycerophospholipids | Alkenylphosphatidylcholine (plasmalogen) | PC(P) |
| PC(P-17:0/20:4) (a) | glycerophospholipids | Alkenylphosphatidylcholine (plasmalogen) | PC(P) |
| PC(P-17:0/20:4) (b) | glycerophospholipids | Alkenylphosphatidylcholine (plasmalogen) | PC(P) |
| PC(P-18:0/18:2) | glycerophospholipids | Alkenylphosphatidylcholine (plasmalogen) | PC(P) |

| | | | |
|---|---|---|---|
| PC(P-18:0/20:4) | glycerophospholipids | Alkenylphosphatidylcholine (plasmalogen) | PC(P) |
| PC(P-18:0/22:5) | glycerophospholipids | Alkenylphosphatidylcholine (plasmalogen) | PC(P) |
| PC(P-18:0/22:6) | glycerophospholipids | Alkenylphosphatidylcholine (plasmalogen) | PC(P) |
| PC(P-18:1/18:1) | glycerophospholipids | Alkenylphosphatidylcholine (plasmalogen) | PC(P) |
| PC(P-18:1/22:6) | glycerophospholipids | Alkenylphosphatidylcholine (plasmalogen) | PC(P) |
| PC(P-20:0/20:4) | glycerophospholipids | Alkenylphosphatidylcholine (plasmalogen) | PC(P) |
| PC(P-35:2) (a) | glycerophospholipids | Alkenylphosphatidylcholine (plasmalogen) | PC(P) |
| PC(P-35:2) (b) | glycerophospholipids | Alkenylphosphatidylcholine (plasmalogen) | PC(P) |
| PC(P-36:3) | glycerophospholipids | Alkenylphosphatidylcholine (plasmalogen) | PC(P) |
| PC(P-38:5) (a) | glycerophospholipids | Alkenylphosphatidylcholine (plasmalogen) | PC(P) |
| PC(P-38:5) (b) | glycerophospholipids | Alkenylphosphatidylcholine (plasmalogen) | PC(P) |
| LPC(14:0) [sn1] | glycerophospholipids | Lysophosphatidylcholine | LPC |
| LPC(14:0) [sn2] | glycerophospholipids | Lysophosphatidylcholine | LPC |
| LPC(15-MHDA) [sn1] [104_sn1] | glycerophospholipids | Lysophosphatidylcholine | LPC |
| LPC(15-MHDA) [sn1] & LPC(17:0) [sn2] | glycerophospholipids | Lysophosphatidylcholine | LPC |
| LPC(15-MHDA) [sn2] | glycerophospholipids | Lysophosphatidylcholine | LPC |
| LPC(15:0) [sn1] | glycerophospholipids | Lysophosphatidylcholine | LPC |
| LPC(15:0) [sn2] | glycerophospholipids | Lysophosphatidylcholine | LPC |
| LPC(16:0) [sn1] | glycerophospholipids | Lysophosphatidylcholine | LPC |
| LPC(16:0) [sn2] | glycerophospholipids | Lysophosphatidylcholine | LPC |
| LPC(16:1) [sn1] | glycerophospholipids | Lysophosphatidylcholine | LPC |
| LPC(16:1) [sn2] | glycerophospholipids | Lysophosphatidylcholine | LPC |
| LPC(17:0) [sn1] | glycerophospholipids | Lysophosphatidylcholine | LPC |
| LPC(17:1) (a) [sn1] [104_sn1] | glycerophospholipids | Lysophosphatidylcholine | LPC |
| LPC(17:1) [sn1] (a) & LPC(17:1) [sn2] (b) | glycerophospholipids | Lysophosphatidylcholine | LPC |
| LPC(17:1) [sn1] (b) | glycerophospholipids | Lysophosphatidylcholine | LPC |
| LPC(17:1) [sn2] (a) | glycerophospholipids | Lysophosphatidylcholine | LPC |
| LPC(18:0) [sn1] | glycerophospholipids | Lysophosphatidylcholine | LPC |
| LPC(18:0) [sn2] | glycerophospholipids | Lysophosphatidylcholine | LPC |
| LPC(18:1) [sn1] | glycerophospholipids | Lysophosphatidylcholine | LPC |
| LPC(18:1) [sn2] | glycerophospholipids | Lysophosphatidylcholine | LPC |
| LPC(18:2) [sn1] | glycerophospholipids | Lysophosphatidylcholine | LPC |
| LPC(18:2) [sn2] | glycerophospholipids | Lysophosphatidylcholine | LPC |
| LPC(18:3) (a) [sn1] [104_sn1] | glycerophospholipids | Lysophosphatidylcholine | LPC |
| LPC(18:3) [sn1] (a) & LPC(18:3) [sn2] (b) | glycerophospholipids | Lysophosphatidylcholine | LPC |
| LPC(18:3) [sn1] (b) | glycerophospholipids | Lysophosphatidylcholine | LPC |
| LPC(18:3) [sn2] (a) | glycerophospholipids | Lysophosphatidylcholine | LPC |
| LPC(19:0) (a) [sn1] [104_sn1] | glycerophospholipids | Lysophosphatidylcholine | LPC |
| LPC(19:0) [sn1] (a) & LPC(19:0) [sn2] (b) | glycerophospholipids | Lysophosphatidylcholine | LPC |
| LPC(19:0) [sn1] (b) | glycerophospholipids | Lysophosphatidylcholine | LPC |
| LPC(19:0) [sn2] (a) | glycerophospholipids | Lysophosphatidylcholine | LPC |
| LPC(19:1) (a) | glycerophospholipids | Lysophosphatidylcholine | LPC |
| LPC(19:1) (b) | glycerophospholipids | Lysophosphatidylcholine | LPC |
| LPC(19:1) (c) | glycerophospholipids | Lysophosphatidylcholine | LPC |
| LPC(20:0) [sn1] | glycerophospholipids | Lysophosphatidylcholine | LPC |
| LPC(20:0) [sn2] | glycerophospholipids | Lysophosphatidylcholine | LPC |

| | | | |
|---|---|---|---|
| LPC(20:1) [sn1] | glycerophospholipids | Lysophosphatidylcholine | LPC |
| LPC(20:1) [sn2] | glycerophospholipids | Lysophosphatidylcholine | LPC |
| LPC(20:2) [sn1] | glycerophospholipids | Lysophosphatidylcholine | LPC |
| LPC(20:2) [sn2] | glycerophospholipids | Lysophosphatidylcholine | LPC |
| LPC(20:3) [sn1] | glycerophospholipids | Lysophosphatidylcholine | LPC |
| LPC(20:3) [sn2] | glycerophospholipids | Lysophosphatidylcholine | LPC |
| LPC(20:4) [sn1] | glycerophospholipids | Lysophosphatidylcholine | LPC |
| LPC(20:4) [sn2] | glycerophospholipids | Lysophosphatidylcholine | LPC |
| LPC(20:5) [sn1] | glycerophospholipids | Lysophosphatidylcholine | LPC |
| LPC(20:5) [sn2] | glycerophospholipids | Lysophosphatidylcholine | LPC |
| LPC(22:0) [sn1] | glycerophospholipids | Lysophosphatidylcholine | LPC |
| LPC(22:0) [sn2] | glycerophospholipids | Lysophosphatidylcholine | LPC |
| LPC(22:1) [sn1] | glycerophospholipids | Lysophosphatidylcholine | LPC |
| LPC(22:1) [sn2] | glycerophospholipids | Lysophosphatidylcholine | LPC |
| LPC(22:4) [sn1] | glycerophospholipids | Lysophosphatidylcholine | LPC |
| LPC(22:4) [sn2] | glycerophospholipids | Lysophosphatidylcholine | LPC |
| LPC(22:5) (n3) [sn1] [104_sn1] | glycerophospholipids | Lysophosphatidylcholine | LPC |
| LPC(22:5) [sn1] (n3) & LPC(22:5) [sn2] (n6) | glycerophospholipids | Lysophosphatidylcholine | LPC |
| LPC(22:5) [sn1] (n6) | glycerophospholipids | Lysophosphatidylcholine | LPC |
| LPC(22:5) [sn2] (n3) | glycerophospholipids | Lysophosphatidylcholine | LPC |
| LPC(22:6) [sn1] | glycerophospholipids | Lysophosphatidylcholine | LPC |
| LPC(22:6) [sn2] | glycerophospholipids | Lysophosphatidylcholine | LPC |
| LPC(24:0) [sn1] | glycerophospholipids | Lysophosphatidylcholine | LPC |
| LPC(24:0) [sn2] | glycerophospholipids | Lysophosphatidylcholine | LPC |
| LPC(26:0) [sn1] | glycerophospholipids | Lysophosphatidylcholine | LPC |
| LPC(26:0) [sn2] | glycerophospholipids | Lysophosphatidylcholine | LPC |
| LPC(O-16:0) | glycerophospholipids | Lysoalkylphosphatidylcholine (lysoplatelet activating factor) | LPC(O) |
| LPC(O-18:0) | glycerophospholipids | Lysoalkylphosphatidylcholine (lysoplatelet activating factor) | LPC(O) |
| LPC(O-18:1) | glycerophospholipids | Lysoalkylphosphatidylcholine (lysoplatelet activating factor) | LPC(O) |
| LPC(O-20:0) | glycerophospholipids | Lysoalkylphosphatidylcholine (lysoplatelet activating factor) | LPC(O) |
| LPC(O-20:1) | glycerophospholipids | Lysoalkylphosphatidylcholine (lysoplatelet activating factor) | LPC(O) |
| LPC(O-22:0) | glycerophospholipids | Lysoalkylphosphatidylcholine (lysoplatelet activating factor) | LPC(O) |
| LPC(O-22:1) | glycerophospholipids | Lysoalkylphosphatidylcholine (lysoplatelet activating factor) | LPC(O) |
| LPC(O-24:0) | glycerophospholipids | Lysoalkylphosphatidylcholine (lysoplatelet activating factor) | LPC(O) |
| LPC(O-24:1) | glycerophospholipids | Lysoalkylphosphatidylcholine (lysoplatelet activating factor) | LPC(O) |
| LPC(O-24:2) | glycerophospholipids | Lysoalkylphosphatidylcholine (lysoplatelet activating factor) | LPC(O) |
| LPC(P-16:0) | glycerophospholipids | Lysoalkenylphosphatidylcholine (plasmalogen) | LPC(P) |
| LPC(P-17:0) (a) | glycerophospholipids | Lysoalkenylphosphatidylcholine (plasmalogen) | LPC(P) |
| LPC(P-17:0) (b) | glycerophospholipids | Lysoalkenylphosphatidylcholine (plasmalogen) | LPC(P) |
| LPC(P-18:0) | glycerophospholipids | Lysoalkenylphosphatidylcholine (plasmalogen) | LPC(P) |
| LPC(P-18:1) | glycerophospholipids | Lysoalkenylphosphatidylcholine (plasmalogen) | LPC(P) |

| | | | |
|---|---|---|---|
| LPC(P-20:0) | glycerophospholipids | Lysoalkenylphosphatidylcholine (plasmalogen) | LPC(P) |
| PE(15-MHDA_18:1) | glycerophospholipids | Phosphatidylethanolamine | PE |
| PE(15-MHDA_18:2) | glycerophospholipids | Phosphatidylethanolamine | PE |
| PE(15-MHDA_20:4) | glycerophospholipids | Phosphatidylethanolamine | PE |
| PE(15-MHDA_22:6) | glycerophospholipids | Phosphatidylethanolamine | PE |
| PE(16:0_16:0) | glycerophospholipids | Phosphatidylethanolamine | PE |
| PE(16:0_16:1) | glycerophospholipids | Phosphatidylethanolamine | PE |
| PE(16:0_18:1) | glycerophospholipids | Phosphatidylethanolamine | PE |
| PE(16:0_18:2) | glycerophospholipids | Phosphatidylethanolamine | PE |
| PE(16:0_18:3) (a) | glycerophospholipids | Phosphatidylethanolamine | PE |
| PE(16:0_18:3) (b) | glycerophospholipids | Phosphatidylethanolamine | PE |
| PE(16:0_20:3) | glycerophospholipids | Phosphatidylethanolamine | PE |
| PE(16:0_20:4) | glycerophospholipids | Phosphatidylethanolamine | PE |
| PE(16:0_20:5) | glycerophospholipids | Phosphatidylethanolamine | PE |
| PE(16:0_22:6) | glycerophospholipids | Phosphatidylethanolamine | PE |
| PE(16:1_18:2) | glycerophospholipids | Phosphatidylethanolamine | PE |
| PE(16:1_20:4) | glycerophospholipids | Phosphatidylethanolamine | PE |
| PE(17:0_18:1) | glycerophospholipids | Phosphatidylethanolamine | PE |
| PE(17:0_18:2) | glycerophospholipids | Phosphatidylethanolamine | PE |
| PE(17:0_20:4) | glycerophospholipids | Phosphatidylethanolamine | PE |
| PE(17:0_22:6) | glycerophospholipids | Phosphatidylethanolamine | PE |
| PE(18:0_18:1) | glycerophospholipids | Phosphatidylethanolamine | PE |
| PE(18:0_18:2) | glycerophospholipids | Phosphatidylethanolamine | PE |
| PE(18:0_20:3) (a) | glycerophospholipids | Phosphatidylethanolamine | PE |
| PE(18:0_20:3) (b) | glycerophospholipids | Phosphatidylethanolamine | PE |
| PE(18:0_20:4) | glycerophospholipids | Phosphatidylethanolamine | PE |
| PE(18:0_22:4) | glycerophospholipids | Phosphatidylethanolamine | PE |
| PE(18:0_22:5) (n3) | glycerophospholipids | Phosphatidylethanolamine | PE |
| PE(18:0_22:5) (n6) | glycerophospholipids | Phosphatidylethanolamine | PE |
| PE(18:0_22:6) | glycerophospholipids | Phosphatidylethanolamine | PE |
| PE(18:1_18:1) | glycerophospholipids | Phosphatidylethanolamine | PE |
| PE(18:1_18:2) | glycerophospholipids | Phosphatidylethanolamine | PE |
| PE(18:1_22:6) (a) | glycerophospholipids | Phosphatidylethanolamine | PE |
| PE(18:1_22:6) (b) | glycerophospholipids | Phosphatidylethanolamine | PE |
| PE(20:0_20:4) | glycerophospholipids | Phosphatidylethanolamine | PE |
| PE(36:0) | glycerophospholipids | Phosphatidylethanolamine | PE |
| PE(38:5) (a) | glycerophospholipids | Phosphatidylethanolamine | PE |
| PE(38:5) (b) | glycerophospholipids | Phosphatidylethanolamine | PE |
| PE(O-16:0/18:2) | glycerophospholipids | Alkylphosphatidylethanolamine | PE(O) |
| PE(O-16:0/20:3) | glycerophospholipids | Alkylphosphatidylethanolamine | PE(O) |
| PE(O-16:0/20:4) | glycerophospholipids | Alkylphosphatidylethanolamine | PE(O) |
| PE(O-16:0/22:4) | glycerophospholipids | Alkylphosphatidylethanolamine | PE(O) |
| PE(O-16:0/22:6) | glycerophospholipids | Alkylphosphatidylethanolamine | PE(O) |
| PE(O-18:0/20:4) | glycerophospholipids | Alkylphosphatidylethanolamine | PE(O) |
| PE(O-18:0/22:5) | glycerophospholipids | Alkylphosphatidylethanolamine | PE(O) |
| PE(O-18:0/22:6) | glycerophospholipids | Alkylphosphatidylethanolamine | PE(O) |
| PE(O-18:1/18:2) | glycerophospholipids | Alkylphosphatidylethanolamine | PE(O) |
| PE(O-18:1/22:6) | glycerophospholipids | Alkylphosphatidylethanolamine | PE(O) |
| PE(O-34:1) | glycerophospholipids | Alkylphosphatidylethanolamine | PE(O) |
| PE(O-36:5) | glycerophospholipids | Alkylphosphatidylethanolamine | PE(O) |
| PE(O-38:5) (a) | glycerophospholipids | Alkylphosphatidylethanolamine | PE(O) |
| PE(O-38:5) (b) | glycerophospholipids | Alkylphosphatidylethanolamine | PE(O) |
| PE(P-15:0/20:4) (a) | glycerophospholipids | Alkenylphosphatidylethanolamine (plasmalogen) | PE(P) |
| PE(P-15:0/20:4) (b) | glycerophospholipids | Alkenylphosphatidylethanolamine (plasmalogen) | PE(P) |

| | | | |
|---|---|---|---|
| PE(P-15:0/22:6) (a) | glycerophospholipids | Alkenylphosphatidylethanolamine (plasmalogen) | PE(P) |
| PE(P-15:0/22:6) (b) | glycerophospholipids | Alkenylphosphatidylethanolamine (plasmalogen) | PE(P) |
| PE(P-16:0/18:1) | glycerophospholipids | Alkenylphosphatidylethanolamine (plasmalogen) | PE(P) |
| PE(P-16:0/18:2) | glycerophospholipids | Alkenylphosphatidylethanolamine (plasmalogen) | PE(P) |
| PE(P-16:0/18:3) | glycerophospholipids | Alkenylphosphatidylethanolamine (plasmalogen) | PE(P) |
| PE(P-16:0/20:3) (a) | glycerophospholipids | Alkenylphosphatidylethanolamine (plasmalogen) | PE(P) |
| PE(P-16:0/20:3) (b) | glycerophospholipids | Alkenylphosphatidylethanolamine (plasmalogen) | PE(P) |
| PE(P-16:0/20:4) | glycerophospholipids | Alkenylphosphatidylethanolamine (plasmalogen) | PE(P) |
| PE(P-16:0/20:5) | glycerophospholipids | Alkenylphosphatidylethanolamine (plasmalogen) | PE(P) |
| PE(P-16:0/22:4) | glycerophospholipids | Alkenylphosphatidylethanolamine (plasmalogen) | PE(P) |
| PE(P-16:0/22:5) (n3) | glycerophospholipids | Alkenylphosphatidylethanolamine (plasmalogen) | PE(P) |
| PE(P-16:0/22:5) (n6) | glycerophospholipids | Alkenylphosphatidylethanolamine (plasmalogen) | PE(P) |
| PE(P-16:0/22:6) | glycerophospholipids | Alkenylphosphatidylethanolamine (plasmalogen) | PE(P) |
| PE(P-17:0/20:4) (a) | glycerophospholipids | Alkenylphosphatidylethanolamine (plasmalogen) | PE(P) |
| PE(P-17:0/20:4) (b) | glycerophospholipids | Alkenylphosphatidylethanolamine (plasmalogen) | PE(P) |
| PE(P-17:0/22:6) (a) | glycerophospholipids | Alkenylphosphatidylethanolamine (plasmalogen) | PE(P) |
| PE(P-17:0/22:6) (b) | glycerophospholipids | Alkenylphosphatidylethanolamine (plasmalogen) | PE(P) |
| PE(P-18:0/18:1) | glycerophospholipids | Alkenylphosphatidylethanolamine (plasmalogen) | PE(P) |
| PE(P-18:0/18:2) | glycerophospholipids | Alkenylphosphatidylethanolamine (plasmalogen) | PE(P) |
| PE(P-18:0/18:3) | glycerophospholipids | Alkenylphosphatidylethanolamine (plasmalogen) | PE(P) |
| PE(P-18:0/20:3) (a) | glycerophospholipids | Alkenylphosphatidylethanolamine (plasmalogen) | PE(P) |
| PE(P-18:0/20:3) (b) | glycerophospholipids | Alkenylphosphatidylethanolamine (plasmalogen) | PE(P) |
| PE(P-18:0/20:4) | glycerophospholipids | Alkenylphosphatidylethanolamine (plasmalogen) | PE(P) |
| PE(P-18:0/20:5) | glycerophospholipids | Alkenylphosphatidylethanolamine (plasmalogen) | PE(P) |
| PE(P-18:0/22:4) | glycerophospholipids | Alkenylphosphatidylethanolamine (plasmalogen) | PE(P) |
| PE(P-18:0/22:5) (n3) | glycerophospholipids | Alkenylphosphatidylethanolamine (plasmalogen) | PE(P) |
| PE(P-18:0/22:5) (n6) | glycerophospholipids | Alkenylphosphatidylethanolamine (plasmalogen) | PE(P) |
| PE(P-18:0/22:6) | glycerophospholipids | Alkenylphosphatidylethanolamine (plasmalogen) | PE(P) |

| | | | |
|---|---|---|---|
| PE(P-18:1/18:1) (a) | glycerophospholipids | Alkenylphosphatidylethanolamine (plasmalogen) | PE(P) |
| PE(P-18:1/18:1) (b) | glycerophospholipids | Alkenylphosphatidylethanolamine (plasmalogen) | PE(P) |
| PE(P-18:1/18:2) (a) | glycerophospholipids | Alkenylphosphatidylethanolamine (plasmalogen) | PE(P) |
| PE(P-18:1/18:2) (b) | glycerophospholipids | Alkenylphosphatidylethanolamine (plasmalogen) | PE(P) |
| PE(P-18:1/18:3) | glycerophospholipids | Alkenylphosphatidylethanolamine (plasmalogen) | PE(P) |
| PE(P-18:1/20:3) (a) | glycerophospholipids | Alkenylphosphatidylethanolamine (plasmalogen) | PE(P) |
| PE(P-18:1/20:3) (b) | glycerophospholipids | Alkenylphosphatidylethanolamine (plasmalogen) | PE(P) |
| PE(P-18:1/20:4) (a) | glycerophospholipids | Alkenylphosphatidylethanolamine (plasmalogen) | PE(P) |
| PE(P-18:1/20:4) (b) | glycerophospholipids | Alkenylphosphatidylethanolamine (plasmalogen) | PE(P) |
| PE(P-18:1/20:5) (a) | glycerophospholipids | Alkenylphosphatidylethanolamine (plasmalogen) | PE(P) |
| PE(P-18:1/20:5) (b) | glycerophospholipids | Alkenylphosphatidylethanolamine (plasmalogen) | PE(P) |
| PE(P-18:1/22:4) | glycerophospholipids | Alkenylphosphatidylethanolamine (plasmalogen) | PE(P) |
| PE(P-18:1/22:5) (a) | glycerophospholipids | Alkenylphosphatidylethanolamine (plasmalogen) | PE(P) |
| PE(P-18:1/22:5) (b) | glycerophospholipids | Alkenylphosphatidylethanolamine (plasmalogen) | PE(P) |
| PE(P-18:1/22:6) (a) | glycerophospholipids | Alkenylphosphatidylethanolamine (plasmalogen) | PE(P) |
| PE(P-18:1/22:6) (b) | glycerophospholipids | Alkenylphosphatidylethanolamine (plasmalogen) | PE(P) |
| PE(P-19:0/20:4) (a) | glycerophospholipids | Alkenylphosphatidylethanolamine (plasmalogen) | PE(P) |
| PE(P-19:0/20:4) (b) | glycerophospholipids | Alkenylphosphatidylethanolamine (plasmalogen) | PE(P) |
| PE(P-20:0/18:1) | glycerophospholipids | Alkenylphosphatidylethanolamine (plasmalogen) | PE(P) |
| PE(P-20:0/18:2) | glycerophospholipids | Alkenylphosphatidylethanolamine (plasmalogen) | PE(P) |
| PE(P-20:0/20:4) | glycerophospholipids | Alkenylphosphatidylethanolamine (plasmalogen) | PE(P) |
| PE(P-20:0/22:6) | glycerophospholipids | Alkenylphosphatidylethanolamine (plasmalogen) | PE(P) |
| PE(P-20:1/20:4) | glycerophospholipids | Alkenylphosphatidylethanolamine (plasmalogen) | PE(P) |
| PE(P-20:1/22:6) | glycerophospholipids | Alkenylphosphatidylethanolamine (plasmalogen) | PE(P) |
| LPE(16:0) [sn1] | glycerophospholipids | Lysophosphatidylethanolamine | LPE |
| LPE(16:0) [sn2] | glycerophospholipids | Lysophosphatidylethanolamine | LPE |
| LPE(18:0) [sn1] | glycerophospholipids | Lysophosphatidylethanolamine | LPE |
| LPE(18:0) [sn2] | glycerophospholipids | Lysophosphatidylethanolamine | LPE |
| LPE(18:1) [sn1] | glycerophospholipids | Lysophosphatidylethanolamine | LPE |
| LPE(18:1) [sn2] | glycerophospholipids | Lysophosphatidylethanolamine | LPE |
| LPE(18:2) [sn1] | glycerophospholipids | Lysophosphatidylethanolamine | LPE |
| LPE(18:2) [sn2] | glycerophospholipids | Lysophosphatidylethanolamine | LPE |
| LPE(20:4) [sn1] | glycerophospholipids | Lysophosphatidylethanolamine | LPE |

| | | | |
|---|---|---|---|
| LPE(20:4) [sn2] | glycerophospholipids | Lysophosphatidylethanolamine | LPE |
| LPE(22:6) [sn1] | glycerophospholipids | Lysophosphatidylethanolamine | LPE |
| LPE(22:6) [sn2] | glycerophospholipids | Lysophosphatidylethanolamine | LPE |
| LPE(P-16:0) | glycerophospholipids | Lysoalkenylphosphatidylethanolamine (plasmalogen) | LPE(P) |
| LPE(P-18:0) | glycerophospholipids | Lysoalkenylphosphatidylethanolamine (plasmalogen) | LPE(P) |
| LPE(P-18:1) | glycerophospholipids | Lysoalkenylphosphatidylethanolamine (plasmalogen) | LPE(P) |
| LPE(P-20:0) | glycerophospholipids | Lysoalkenylphosphatidylethanolamine (plasmalogen) | LPE(P) |
| PI(38:5) (b) | glycerophospholipids | Phosphatidylinositol | PI |
| PI(15-MHDA_18:1) & PI(17:0_18:1) | glycerophospholipids | Phosphatidylinositol | PI |
| PI(15-MHDA_18:2) & PI(17:0_18:2) | glycerophospholipids | Phosphatidylinositol | PI |
| PI(15-MHDA_20:4) & PI(17:0_20:4) | glycerophospholipids | Phosphatidylinositol | PI |
| PI(16:0_16:1) | glycerophospholipids | Phosphatidylinositol | PI |
| PI(16:0_20:3) (a) | glycerophospholipids | Phosphatidylinositol | PI |
| PI(16:0_20:3) (b) | glycerophospholipids | Phosphatidylinositol | PI |
| PI(16:0_20:4) | glycerophospholipids | Phosphatidylinositol | PI |
| PI(16:0/16:0) | glycerophospholipids | Phosphatidylinositol | PI |
| PI(18:0_18:1) | glycerophospholipids | Phosphatidylinositol | PI |
| PI(18:0_20:2) | glycerophospholipids | Phosphatidylinositol | PI |
| PI(18:0_20:3) (a) | glycerophospholipids | Phosphatidylinositol | PI |
| PI(18:0_20:3) (b) | glycerophospholipids | Phosphatidylinositol | PI |
| PI(18:0_20:4) | glycerophospholipids | Phosphatidylinositol | PI |
| PI(18:0_22:4) | glycerophospholipids | Phosphatidylinositol | PI |
| PI(18:0_22:5) (n3) | glycerophospholipids | Phosphatidylinositol | PI |
| PI(18:0_22:5) (n6) | glycerophospholipids | Phosphatidylinositol | PI |
| PI(18:0_22:6) | glycerophospholipids | Phosphatidylinositol | PI |
| PI(18:1_18:2) | glycerophospholipids | Phosphatidylinositol | PI |
| PI(20:0_20:4) | glycerophospholipids | Phosphatidylinositol | PI |
| PI(34:0) | glycerophospholipids | Phosphatidylinositol | PI |
| PI(34:1) | glycerophospholipids | Phosphatidylinositol | PI |
| PI(36:2) | glycerophospholipids | Phosphatidylinositol | PI |
| PI(37:6) | glycerophospholipids | Phosphatidylinositol | PI |
| PI(38:5) (a) | glycerophospholipids | Phosphatidylinositol | PI |
| PI(38:6) | glycerophospholipids | Phosphatidylinositol | PI |
| PI(39:6) | glycerophospholipids | Phosphatidylinositol | PI |
| PIP1(38:4) | glycerophospholipids | Phosphatidylinositol monophosphate | PIP1 |
| LPI(18:0) [sn1] | glycerophospholipids | Lysophosphatidylinositol | LPI |
| LPI(18:0) [sn2] | glycerophospholipids | Lysophosphatidylinositol | LPI |
| LPI(18:1) [sn1] | glycerophospholipids | Lysophosphatidylinositol | LPI |
| LPI(18:1) [sn2] | glycerophospholipids | Lysophosphatidylinositol | LPI |
| LPI(18:2) [sn1] | glycerophospholipids | Lysophosphatidylinositol | LPI |
| LPI(18:2) [sn2] | glycerophospholipids | Lysophosphatidylinositol | LPI |
| LPI(20:4) [sn1] | glycerophospholipids | Lysophosphatidylinositol | LPI |
| LPI(20:4) [sn2] | glycerophospholipids | Lysophosphatidylinositol | LPI |
| PS(36:1) | glycerophospholipids | Phosphatidylserine | PS |
| PS(36:2) | glycerophospholipids | Phosphatidylserine | PS |
| PS(38:3) | glycerophospholipids | Phosphatidylserine | PS |
| PS(38:4) | glycerophospholipids | Phosphatidylserine | PS |
| PS(40:5) | glycerophospholipids | Phosphatidylserine | PS |
| PS(40:6) | glycerophospholipids | Phosphatidylserine | PS |
| PG(34:1) | glycerophospholipids | Phosphatidylglycerol | PG |
| PG(36:1) | glycerophospholipids | Phosphatidylglycerol | PG |
| PG(36:2) | glycerophospholipids | Phosphatidylglycerol | PG |

| | | | |
|---|---|---|---|
| CE(14:0) | Neutral/Other | Cholesteryl ester | CE |
| CE(15:0) | Neutral/Other | Cholesteryl ester | CE |
| CE(16:0) | Neutral/Other | Cholesteryl ester | CE |
| CE(16:1) | Neutral/Other | Cholesteryl ester | CE |
| CE(16:2) | Neutral/Other | Cholesteryl ester | CE |
| CE(17:0) | Neutral/Other | Cholesteryl ester | CE |
| CE(17:1) | Neutral/Other | Cholesteryl ester | CE |
| CE(18:0) | Neutral/Other | Cholesteryl ester | CE |
| CE(18:1) | Neutral/Other | Cholesteryl ester | CE |
| CE(18:2) | Neutral/Other | Cholesteryl ester | CE |
| CE(18:3) | Neutral/Other | Cholesteryl ester | CE |
| CE(20:0) | Neutral/Other | Cholesteryl ester | CE |
| CE(20:1) | Neutral/Other | Cholesteryl ester | CE |
| CE(20:2) | Neutral/Other | Cholesteryl ester | CE |
| CE(20:3) | Neutral/Other | Cholesteryl ester | CE |
| CE(20:4) | Neutral/Other | Cholesteryl ester | CE |
| CE(20:5) | Neutral/Other | Cholesteryl ester | CE |
| CE(22:0) | Neutral/Other | Cholesteryl ester | CE |
| CE(22:1) | Neutral/Other | Cholesteryl ester | CE |
| CE(22:4) | Neutral/Other | Cholesteryl ester | CE |
| CE(22:5) | Neutral/Other | Cholesteryl ester | CE |
| CE(22:6) | Neutral/Other | Cholesteryl ester | CE |
| CE(24:0) | Neutral/Other | Cholesteryl ester | CE |
| CE(24:1) | Neutral/Other | Cholesteryl ester | CE |
| CE(24:4) | Neutral/Other | Cholesteryl ester | CE |
| CE(24:5) | Neutral/Other | Cholesteryl ester | CE |
| CE(24:6) | Neutral/Other | Cholesteryl ester | CE |
| COH | Neutral/Other | Free Cholesterol | COH |
| DE(16:0) | Neutral/Other | Dehydrocholesterol ester | DE |
| DE(18:1) | Neutral/Other | Dehydrocholesterol ester | DE |
| DE(18:2) | Neutral/Other | Dehydrocholesterol ester | DE |
| DE(20:4) | Neutral/Other | Dehydrocholesterol ester | DE |
| DE(20:5) | Neutral/Other | Dehydrocholesterol ester | DE |
| DE(22:6) | Neutral/Other | Dehydrocholesterol ester | DE |
| deDE(18:2) | Neutral/Other | Dehydrodemosterol ester | deDE |
| deDE(20:4) | Neutral/Other | Dehydrodemosterol ester | deDE |
| methyl-CE(18:0) | Neutral/Other | Methyl-cholesteryl ester | methyl-CE |
| methyl-CE(18:1) | Neutral/Other | Methyl-cholesteryl ester | methyl-CE |
| methyl-CE(18:2) | Neutral/Other | Methyl-cholesteryl ester | methyl-CE |
| methyl-CE(20:4) | Neutral/Other | Methyl-cholesteryl ester | methyl-CE |
| methyl-CE(22:6) | Neutral/Other | Methyl-cholesteryl ester | methyl-CE |
| methyl-DE(18:1) | Neutral/Other | Methyl-dehydrocholesteryl ester | methyl-DE |
| methyl-DE(18:2) | Neutral/Other | Methyl-dehydrocholesteryl ester | methyl-DE |
| dimethyl-CE(18:1) | Neutral/Other | Dimethyl-cholesteryl ester | dimethyl-CE |
| dimethyl-CE(18:2) | Neutral/Other | Dimethyl-cholesteryl ester | dimethyl-CE |
| dimethyl-CE(20:4) | Neutral/Other | Dimethyl-cholesteryl ester | dimethyl-CE |
| dimethyl-CE(22:6) | Neutral/Other | Dimethyl-cholesteryl ester | dimethyl-CE |
| FA(14:0) | Neutral/Other | Free fatty acid | FFA |
| FA(16:0) | Neutral/Other | Free fatty acid | FFA |
| FA(16:1) | Neutral/Other | Free fatty acid | FFA |
| FA(17:0) | Neutral/Other | Free fatty acid | FFA |
| FA(17:1) | Neutral/Other | Free fatty acid | FFA |
| FA(18:0) | Neutral/Other | Free fatty acid | FFA |
| FA(18:1) | Neutral/Other | Free fatty acid | FFA |
| FA(18:2) | Neutral/Other | Free fatty acid | FFA |
| FA(18:3) | Neutral/Other | Free fatty acid | FFA |
| FA(20:2) | Neutral/Other | Free fatty acid | FFA |

| | | | |
|---|---|---|---|
| FA(20:3) | Neutral/Other | Free fatty acid | FFA |
| FA(20:4) | Neutral/Other | Free fatty acid | FFA |
| FA(20:5) | Neutral/Other | Free fatty acid | FFA |
| FA(22:4) | Neutral/Other | Free fatty acid | FFA |
| FA(22:5) | Neutral/Other | Free fatty acid | FFA |
| FA(22:6) | Neutral/Other | Free fatty acid | FFA |
| AC(12:0) | Neutral/Other | Acylcarnitine | AC |
| AC(12:1) | Neutral/Other | Acylcarnitine | AC |
| AC(13:0) | Neutral/Other | Acylcarnitine | AC |
| AC(14:0) | Neutral/Other | Acylcarnitine | AC |
| AC(14:1) | Neutral/Other | Acylcarnitine | AC |
| AC(14:2) | Neutral/Other | Acylcarnitine | AC |
| AC(15:0) (a) | Neutral/Other | Acylcarnitine | AC |
| AC(15:0) (b) | Neutral/Other | Acylcarnitine | AC |
| AC(16:0) | Neutral/Other | Acylcarnitine | AC |
| AC(16:1) | Neutral/Other | Acylcarnitine | AC |
| AC(17:0) (a) | Neutral/Other | Acylcarnitine | AC |
| AC(17:0) (b) | Neutral/Other | Acylcarnitine | AC |
| AC(18:0) | Neutral/Other | Acylcarnitine | AC |
| AC(18:1) | Neutral/Other | Acylcarnitine | AC |
| AC(18:2) | Neutral/Other | Acylcarnitine | AC |
| AC(18:3) | Neutral/Other | Acylcarnitine | AC |
| AC(20:3) (a) | Neutral/Other | Acylcarnitine | AC |
| AC(20:3) (b) | Neutral/Other | Acylcarnitine | AC |
| AC(20:4) | Neutral/Other | Acylcarnitine | AC |
| AC(20:5) | Neutral/Other | Acylcarnitine | AC |
| AC(22:5) | Neutral/Other | Acylcarnitine | AC |
| AC(22:6) | Neutral/Other | Acylcarnitine | AC |
| AC(24:0) | Neutral/Other | Acylcarnitine | AC |
| AC(24:1) | Neutral/Other | Acylcarnitine | AC |
| AC(26:0) | Neutral/Other | Acylcarnitine | AC |
| AC(26:1) | Neutral/Other | Acylcarnitine | AC |
| AC(14:0)-OH | Neutral/Other | Hydroxylated acylcarnitine | AC-OH |
| AC(14:1)-OH | Neutral/Other | Hydroxylated acylcarnitine | AC-OH |
| AC(16:0)-OH | Neutral/Other | Hydroxylated acylcarnitine | AC-OH |
| AC(16:1)-OH | Neutral/Other | Hydroxylated acylcarnitine | AC-OH |
| AC(18:0)-OH | Neutral/Other | Hydroxylated acylcarnitine | AC-OH |
| AC(18:1)-OH | Neutral/Other | Hydroxylated acylcarnitine | AC-OH |
| AC(20:3)-OH | Neutral/Other | Hydroxylated acylcarnitine | AC-OH |
| AC(22:5)-OH | Neutral/Other | Hydroxylated acylcarnitine | AC-OH |
| AC(24:1)-OH | Neutral/Other | Hydroxylated acylcarnitine | AC-OH |
| CA | Neutral/Other | Bile acid | BA |
| dxCA | Neutral/Other | Bile acid | BA |
| DG(14:0_16:0) | Neutral/Other | Diacylglycerol | DG |
| DG(14:0_18:2) | Neutral/Other | Diacylglycerol | DG |
| DG(16:0_16:0) | Neutral/Other | Diacylglycerol | DG |
| DG(16:0_16:1) | Neutral/Other | Diacylglycerol | DG |
| DG(16:0_18:1) | Neutral/Other | Diacylglycerol | DG |
| DG(16:0_18:2) | Neutral/Other | Diacylglycerol | DG |
| DG(16:0_20:4) | Neutral/Other | Diacylglycerol | DG |
| DG(16:0_22:5) | Neutral/Other | Diacylglycerol | DG |
| DG(16:0_22:6) | Neutral/Other | Diacylglycerol | DG |
| DG(16:1_18:1) | Neutral/Other | Diacylglycerol | DG |
| DG(18:0_18:1) | Neutral/Other | Diacylglycerol | DG |
| DG(18:0_18:2) | Neutral/Other | Diacylglycerol | DG |
| DG(18:0_20:4) | Neutral/Other | Diacylglycerol | DG |
| DG(18:1_18:1) | Neutral/Other | Diacylglycerol | DG |

| | | | |
|---|---|---|---|
| DG(18:1_18:2) | Neutral/Other | Diacylglycerol | DG |
| DG(18:1_18:3) | Neutral/Other | Diacylglycerol | DG |
| DG(18:1_20:3) | Neutral/Other | Diacylglycerol | DG |
| DG(18:1_20:4) | Neutral/Other | Diacylglycerol | DG |
| DG(18:1_20:5) | Neutral/Other | Diacylglycerol | DG |
| DG(18:1_22:5) | Neutral/Other | Diacylglycerol | DG |
| DG(18:1_22:6) | Neutral/Other | Diacylglycerol | DG |
| DG(18:2_18:2) | Neutral/Other | Diacylglycerol | DG |
| DG(18:2_20:4) | Neutral/Other | Diacylglycerol | DG |
| DG(18:2_22:6) | Neutral/Other | Diacylglycerol | DG |
| TG(48:0) [NL-16:0] | Neutral/Other | Triacylglycerol (neutral loss, for associations) | TG [NL] |
| TG(48:0) [NL-18:0] | Neutral/Other | Triacylglycerol (neutral loss, for associations) | TG [NL] |
| TG(48:1) [NL-16:1] | Neutral/Other | Triacylglycerol (neutral loss, for associations) | TG [NL] |
| TG(48:1) [NL-18:1] | Neutral/Other | Triacylglycerol (neutral loss, for associations) | TG [NL] |
| TG(48:2) [NL-14:0] | Neutral/Other | Triacylglycerol (neutral loss, for associations) | TG [NL] |
| TG(48:2) [NL-14:1] | Neutral/Other | Triacylglycerol (neutral loss, for associations) | TG [NL] |
| TG(48:2) [NL-16:1] | Neutral/Other | Triacylglycerol (neutral loss, for associations) | TG [NL] |
| TG(48:2) [NL-18:2] | Neutral/Other | Triacylglycerol (neutral loss, for associations) | TG [NL] |
| TG(48:3) [NL-14:0] | Neutral/Other | Triacylglycerol (neutral loss, for associations) | TG [NL] |
| TG(48:3) [NL-16:1] | Neutral/Other | Triacylglycerol (neutral loss, for associations) | TG [NL] |
| TG(48:3) [NL-18:3] | Neutral/Other | Triacylglycerol (neutral loss, for associations) | TG [NL] |
| TG(49:1) [NL-16:1] | Neutral/Other | Triacylglycerol (neutral loss, for associations) | TG [NL] |
| TG(49:1) [NL-17:1] | Neutral/Other | Triacylglycerol (neutral loss, for associations) | TG [NL] |
| TG(50:0) [NL-18:0] | Neutral/Other | Triacylglycerol (neutral loss, for associations) | TG [NL] |
| TG(50:1) [NL-14:0] | Neutral/Other | Triacylglycerol (neutral loss, for associations) | TG [NL] |
| TG(50:1) [NL-16:0] | Neutral/Other | Triacylglycerol (neutral loss, for associations) | TG [NL] |
| TG(50:1) [NL-18:1] | Neutral/Other | Triacylglycerol (neutral loss, for associations) | TG [NL] |
| TG(50:2) [NL-14:0] | Neutral/Other | Triacylglycerol (neutral loss, for associations) | TG [NL] |
| TG(50:2) [NL-16:1] | Neutral/Other | Triacylglycerol (neutral loss, for associations) | TG [NL] |
| TG(50:2) [NL-18:1] | Neutral/Other | Triacylglycerol (neutral loss, for associations) | TG [NL] |
| TG(50:2) [NL-18:2] | Neutral/Other | Triacylglycerol (neutral loss, for associations) | TG [NL] |
| TG(50:3) [NL-14:0] | Neutral/Other | Triacylglycerol (neutral loss, for associations) | TG [NL] |
| TG(50:3) [NL-14:1] | Neutral/Other | Triacylglycerol (neutral loss, for associations) | TG [NL] |

| | | | |
|---|---|---|---|
| TG(50:3) [NL-16:1] | Neutral/Other | Triacylglycerol (neutral loss, for associations) | TG [NL] |
| TG(50:3) [NL-18:2] | Neutral/Other | Triacylglycerol (neutral loss, for associations) | TG [NL] |
| TG(50:3) [NL-18:3] | Neutral/Other | Triacylglycerol (neutral loss, for associations) | TG [NL] |
| TG(50:4) [NL-14:0] | Neutral/Other | Triacylglycerol (neutral loss, for associations) | TG [NL] |
| TG(50:4) [NL-18:3] | Neutral/Other | Triacylglycerol (neutral loss, for associations) | TG [NL] |
| TG(50:4) [NL-20:4] | Neutral/Other | Triacylglycerol (neutral loss, for associations) | TG [NL] |
| TG(51:0) [NL-16:0] | Neutral/Other | Triacylglycerol (neutral loss, for associations) | TG [NL] |
| TG(51:1) [NL-17:0] | Neutral/Other | Triacylglycerol (neutral loss, for associations) | TG [NL] |
| TG(51:2) [NL-15:0] | Neutral/Other | Triacylglycerol (neutral loss, for associations) | TG [NL] |
| TG(51:2) [NL-17:0] | Neutral/Other | Triacylglycerol (neutral loss, for associations) | TG [NL] |
| TG(51:2) [NL-17:1] | Neutral/Other | Triacylglycerol (neutral loss, for associations) | TG [NL] |
| TG(52:1) [NL-18:0] | Neutral/Other | Triacylglycerol (neutral loss, for associations) | TG [NL] |
| TG(52:1) [NL-18:1] | Neutral/Other | Triacylglycerol (neutral loss, for associations) | TG [NL] |
| TG(52:2) [NL-16:0] | Neutral/Other | Triacylglycerol (neutral loss, for associations) | TG [NL] |
| TG(52:2) [NL-18:2] | Neutral/Other | Triacylglycerol (neutral loss, for associations) | TG [NL] |
| TG(52:3) [NL-16:1] | Neutral/Other | Triacylglycerol (neutral loss, for associations) | TG [NL] |
| TG(52:3) [NL-18:2] | Neutral/Other | Triacylglycerol (neutral loss, for associations) | TG [NL] |
| TG(52:4) [NL-16:1] | Neutral/Other | Triacylglycerol (neutral loss, for associations) | TG [NL] |
| TG(52:4) [NL-18:2] | Neutral/Other | Triacylglycerol (neutral loss, for associations) | TG [NL] |
| TG(52:4) [NL-18:3] | Neutral/Other | Triacylglycerol (neutral loss, for associations) | TG [NL] |
| TG(52:5) [NL-18:3] | Neutral/Other | Triacylglycerol (neutral loss, for associations) | TG [NL] |
| TG(52:5) [NL-20:4] | Neutral/Other | Triacylglycerol (neutral loss, for associations) | TG [NL] |
| TG(52:5) [NL-20:5] | Neutral/Other | Triacylglycerol (neutral loss, for associations) | TG [NL] |
| TG(53:2) [NL-17:1] | Neutral/Other | Triacylglycerol (neutral loss, for associations) | TG [NL] |
| TG(53:2) [NL-18:1] | Neutral/Other | Triacylglycerol (neutral loss, for associations) | TG [NL] |
| TG(54:0) [NL-18:0] | Neutral/Other | Triacylglycerol (neutral loss, for associations) | TG [NL] |
| TG(54:1) [NL-18:1] | Neutral/Other | Triacylglycerol (neutral loss, for associations) | TG [NL] |
| TG(54:2) [NL-18:0] | Neutral/Other | Triacylglycerol (neutral loss, for associations) | TG [NL] |

| | | | |
|---|---|---|---|
| TG(54:2) [NL-20:1] | Neutral/Other | Triacylglycerol (neutral loss, for associations) | TG [NL] |
| TG(54:3) [NL-18:1] | Neutral/Other | Triacylglycerol (neutral loss, for associations) | TG [NL] |
| TG(54:3) [NL-18:2] | Neutral/Other | Triacylglycerol (neutral loss, for associations) | TG [NL] |
| TG(54:4) [NL-18:2] | Neutral/Other | Triacylglycerol (neutral loss, for associations) | TG [NL] |
| TG(54:4) [NL-20:3] | Neutral/Other | Triacylglycerol (neutral loss, for associations) | TG [NL] |
| TG(54:5) [NL-18:3] | Neutral/Other | Triacylglycerol (neutral loss, for associations) | TG [NL] |
| TG(54:5) [NL-20:4] | Neutral/Other | Triacylglycerol (neutral loss, for associations) | TG [NL] |
| TG(54:6) [NL-18:3] | Neutral/Other | Triacylglycerol (neutral loss, for associations) | TG [NL] |
| TG(54:6) [NL-20:4] | Neutral/Other | Triacylglycerol (neutral loss, for associations) | TG [NL] |
| TG(54:6) [NL-20:5] | Neutral/Other | Triacylglycerol (neutral loss, for associations) | TG [NL] |
| TG(54:6) [NL-22:6] | Neutral/Other | Triacylglycerol (neutral loss, for associations) | TG [NL] |
| TG(54:7) [NL-20:5] | Neutral/Other | Triacylglycerol (neutral loss, for associations) | TG [NL] |
| TG(54:7) [NL-22:6] | Neutral/Other | Triacylglycerol (neutral loss, for associations) | TG [NL] |
| TG(56:6) [NL-20:4] | Neutral/Other | Triacylglycerol (neutral loss, for associations) | TG [NL] |
| TG(56:6) [NL-22:5] | Neutral/Other | Triacylglycerol (neutral loss, for associations) | TG [NL] |
| TG(56:7) [NL-20:4] | Neutral/Other | Triacylglycerol (neutral loss, for associations) | TG [NL] |
| TG(56:7) [NL-20:5] | Neutral/Other | Triacylglycerol (neutral loss, for associations) | TG [NL] |
| TG(56:7) [NL-22:5] | Neutral/Other | Triacylglycerol (neutral loss, for associations) | TG [NL] |
| TG(56:7) [NL-22:6] | Neutral/Other | Triacylglycerol (neutral loss, for associations) | TG [NL] |
| TG(56:8) [NL-20:4] | Neutral/Other | Triacylglycerol (neutral loss, for associations) | TG [NL] |
| TG(56:8) [NL-20:5] | Neutral/Other | Triacylglycerol (neutral loss, for associations) | TG [NL] |
| TG(56:8) [NL-22:6] | Neutral/Other | Triacylglycerol (neutral loss, for associations) | TG [NL] |
| TG(56:9) [NL-22:6] | Neutral/Other | Triacylglycerol (neutral loss, for associations) | TG [NL] |
| TG(58:10) [NL-22:6] | Neutral/Other | Triacylglycerol (neutral loss, for associations) | TG [NL] |
| TG(58:8) [NL-22:6] | Neutral/Other | Triacylglycerol (neutral loss, for associations) | TG [NL] |
| TG(58:9) [NL-22:6] | Neutral/Other | Triacylglycerol (neutral loss, for associations) | TG [NL] |
| TG(O-50:1) [NL-15:0] | Neutral/Other | Alkyldiacylglycerol (neutral loss, for associations) | TG(O) [NL] |
| TG(O-50:1) [NL-16:0] | Neutral/Other | Alkyldiacylglycerol (neutral loss, for associations) | TG(O) [NL] |

| | | | |
|---|---|---|---|
| TG(O-50:1) [NL-17:1] | Neutral/Other | Alkyldiacylglycerol (neutral loss, for associations) | TG(O) [NL] |
| TG(O-50:1) [NL-18:1] | Neutral/Other | Alkyldiacylglycerol (neutral loss, for associations) | TG(O) [NL] |
| TG(O-50:2) [NL-18:1] | Neutral/Other | Alkyldiacylglycerol (neutral loss, for associations) | TG(O) [NL] |
| TG(O-50:2) [NL-18:2] | Neutral/Other | Alkyldiacylglycerol (neutral loss, for associations) | TG(O) [NL] |
| TG(O-50:3) [NL-18:2] | Neutral/Other | Alkyldiacylglycerol (neutral loss, for associations) | TG(O) [NL] |
| TG(O-52:0) [NL-16:0] | Neutral/Other | Alkyldiacylglycerol (neutral loss, for associations) | TG(O) [NL] |
| TG(O-52:1) [NL-16:0] | Neutral/Other | Alkyldiacylglycerol (neutral loss, for associations) | TG(O) [NL] |
| TG(O-52:1) [NL-18:1] | Neutral/Other | Alkyldiacylglycerol (neutral loss, for associations) | TG(O) [NL] |
| TG(O-52:2) [NL-16:0] | Neutral/Other | Alkyldiacylglycerol (neutral loss, for associations) | TG(O) [NL] |
| TG(O-52:2) [NL-17:1] | Neutral/Other | Alkyldiacylglycerol (neutral loss, for associations) | TG(O) [NL] |
| TG(O-52:2) [NL-18:1] | Neutral/Other | Alkyldiacylglycerol (neutral loss, for associations) | TG(O) [NL] |
| TG(O-54:2) [NL-17:1] | Neutral/Other | Alkyldiacylglycerol (neutral loss, for associations) | TG(O) [NL] |
| TG(O-54:2) [NL-18:1] | Neutral/Other | Alkyldiacylglycerol (neutral loss, for associations) | TG(O) [NL] |
| TG(O-54:3) [NL-17:1] | Neutral/Other | Alkyldiacylglycerol (neutral loss, for associations) | TG(O) [NL] |
| TG(O-54:3) [NL-18:1] | Neutral/Other | Alkyldiacylglycerol (neutral loss, for associations) | TG(O) [NL] |
| TG(O-54:4) [NL-17:1] | Neutral/Other | Alkyldiacylglycerol (neutral loss, for associations) | TG(O) [NL] |
| TG(O-54:4) [NL-18:2] | Neutral/Other | Alkyldiacylglycerol (neutral loss, for associations) | TG(O) [NL] |
| Ubiquinone | Neutral/Other | Ubiquinone | Ubiquinone |
| CE(18:2) [+OH] | Neutral/Other | Oxidised lipids | OxSpecies |
| CE(20:4) [+OH] | Neutral/Other | Oxidised lipids | OxSpecies |
| CE(22:6) [+OH] | Neutral/Other | Oxidised lipids | OxSpecies |
| LPC(18:2) [+OH] | Neutral/Other | Oxidised lipids | OxSpecies |
| LPC(20:4) [+OH] | Neutral/Other | Oxidised lipids | OxSpecies |
| LPC(22:6) [+OH] | Neutral/Other | Oxidised lipids | OxSpecies |
| PC(34:2) [+OH] | Neutral/Other | Oxidised lipids | OxSpecies |
| PC(36:4) [+OH] | Neutral/Other | Oxidised lipids | OxSpecies |
