## Supplementary Table 3 for "Circulating lipid profiles are associated with cross-sectional and longitudinal changes of central biomarkers for Alzheimer’s disease"

**Supplementary Table 3. Associations of lipid classes with baseline A/T/N biomarkers**

| | A: Amyloid PET (AV45) uptake | | | | | T: CSF pTau | | | | | N1: Hippocampal volume | | | | | N2: FDG uptake | | | | |
|---|---|---|---|---|---|---|---|---|---|---|---|---|---|---|---|---|---|---|---|---|
| Lipid classes | Beta | 95% CI (Lower) | 95% CI (Upper) | Pvalue | Pvalue(BH) | Beta | 95% CI (Lower) | 95% CI (Upper) | Pvalue | Pvalue(BH) | Beta | 95% CI (Lower) | 95% CI (Upper) | Pvalue | Pvalue(BH) | Beta | 95% CI (Lower) | 95% CI (Upper) | Pvalue | Pvalue(BH) |
| LPC(O) | 0.121 | 0.053 | 0.189 | 0.001 | 0.024 | 0.056 | -0.006 | 0.118 | 0.076 | 0.414 | -0.033 | -0.085 | 0.019 | 0.211 | 0.649 | -0.111 | -0.175 | -0.047 | 0.001 | 0.011 |
| LPC | 0.119 | 0.047 | 0.192 | 0.001 | 0.030 | 0.064 | -0.001 | 0.128 | 0.054 | 0.414 | -0.012 | -0.064 | 0.039 | 0.638 | 0.889 | -0.055 | -0.122 | 0.012 | 0.110 | 0.224 |
| LPC(P) | 0.096 | 0.027 | 0.165 | 0.006 | 0.047 | 0.049 | -0.012 | 0.111 | 0.115 | 0.414 | -0.009 | -0.061 | 0.043 | 0.734 | 0.916 | -0.080 | -0.144 | -0.017 | 0.013 | 0.071 |
| LPE | 0.104 | 0.030 | 0.178 | 0.006 | 0.047 | 0.060 | -0.006 | 0.127 | 0.074 | 0.414 | -0.044 | -0.095 | 0.006 | 0.086 | 0.360 | -0.070 | -0.139 | 0.000 | 0.048 | 0.137 |
| Hex2Cer | 0.115 | 0.035 | 0.194 | 0.005 | 0.047 | 0.062 | -0.009 | 0.132 | 0.085 | 0.414 | -0.030 | -0.082 | 0.022 | 0.254 | 0.649 | -0.055 | -0.129 | 0.019 | 0.146 | 0.269 |
| DE | -0.127 | -0.213 | -0.041 | 0.004 | 0.047 | -0.077 | -0.153 | -0.002 | 0.045 | 0.414 | 0.029 | -0.026 | 0.084 | 0.299 | 0.687 | 0.055 | -0.023 | 0.133 | 0.170 | 0.300 |
| PE(O) | -0.095 | -0.169 | -0.022 | 0.011 | 0.073 | -0.051 | -0.117 | 0.014 | 0.126 | 0.414 | 0.064 | 0.014 | 0.113 | 0.012 | 0.094 | 0.014 | -0.054 | 0.082 | 0.689 | 0.754 |
| HexCer | 0.089 | 0.015 | 0.163 | 0.018 | 0.106 | 0.024 | -0.041 | 0.090 | 0.465 | 0.684 | -0.016 | -0.065 | 0.033 | 0.515 | 0.845 | -0.018 | -0.086 | 0.051 | 0.609 | 0.700 |
| GM1 | 0.079 | 0.007 | 0.150 | 0.030 | 0.148 | 0.003 | -0.059 | 0.065 | 0.921 | 0.935 | -0.056 | -0.103 | -0.009 | 0.020 | 0.135 | -0.085 | -0.151 | -0.020 | 0.011 | 0.071 |
| PE(P) | -0.082 | -0.158 | -0.007 | 0.032 | 0.148 | -0.031 | -0.096 | 0.035 | 0.361 | 0.594 | 0.091 | 0.040 | 0.142 | 0.001 | 0.008 | 0.022 | -0.046 | 0.090 | 0.528 | 0.639 |
| deDE | 0.077 | 0.002 | 0.152 | 0.045 | 0.174 | 0.070 | 0.004 | 0.136 | 0.038 | 0.414 | -0.149 | -0.197 | -0.102 | 0.000 | 0.000 | -0.140 | -0.208 | -0.073 | 0.000 | 0.002 |
| dhCer | -0.081 | -0.160 | -0.002 | 0.045 | 0.174 | -0.038 | -0.108 | 0.031 | 0.279 | 0.584 | 0.034 | -0.018 | 0.086 | 0.199 | 0.649 | 0.007 | -0.065 | 0.080 | 0.848 | 0.907 |
| OxSpecies | 0.064 | -0.008 | 0.135 | 0.082 | 0.291 | -0.010 | -0.074 | 0.053 | 0.752 | 0.805 | 0.009 | -0.040 | 0.058 | 0.725 | 0.916 | -0.052 | -0.117 | 0.013 | 0.117 | 0.224 |
| GM3 | 0.058 | -0.026 | 0.143 | 0.174 | 0.500 | 0.043 | -0.032 | 0.117 | 0.262 | 0.574 | -0.029 | -0.091 | 0.033 | 0.355 | 0.743 | -0.136 | -0.213 | -0.058 | 0.001 | 0.011 |
| dimethyl-CE | -0.052 | -0.126 | 0.022 | 0.167 | 0.500 | -0.055 | -0.122 | 0.011 | 0.105 | 0.414 | 0.082 | 0.032 | 0.131 | 0.001 | 0.014 | 0.083 | 0.015 | 0.151 | 0.017 | 0.078 |
| TG(O) [NL] | -0.051 | -0.123 | 0.020 | 0.157 | 0.500 | -0.050 | -0.114 | 0.014 | 0.125 | 0.414 | 0.119 | 0.070 | 0.169 | 0.000 | 0.000 | 0.017 | -0.050 | 0.083 | 0.626 | 0.703 |
| S1P | 0.044 | -0.022 | 0.110 | 0.194 | 0.525 | 0.071 | 0.012 | 0.130 | 0.018 | 0.414 | -0.016 | -0.062 | 0.030 | 0.497 | 0.845 | -0.022 | -0.083 | 0.040 | 0.490 | 0.633 |
| methyl-DE | -0.045 | -0.121 | 0.031 | 0.244 | 0.565 | -0.019 | -0.088 | 0.050 | 0.591 | 0.718 | 0.074 | 0.023 | 0.124 | 0.004 | 0.039 | 0.097 | 0.027 | 0.168 | 0.007 | 0.071 |
| SHexCer | 0.050 | -0.034 | 0.134 | 0.246 | 0.565 | -0.023 | -0.097 | 0.051 | 0.543 | 0.718 | 0.003 | -0.051 | 0.058 | 0.904 | 0.970 | -0.069 | -0.145 | 0.008 | 0.079 | 0.191 |
| LPI | -0.042 | -0.111 | 0.028 | 0.240 | 0.565 | 0.038 | -0.023 | 0.098 | 0.224 | 0.531 | 0.004 | -0.044 | 0.051 | 0.878 | 0.970 | -0.030 | -0.094 | 0.034 | 0.350 | 0.520 |
| DG | -0.066 | -0.193 | 0.061 | 0.309 | 0.592 | -0.051 | -0.158 | 0.055 | 0.343 | 0.585 | -0.031 | -0.114 | 0.052 | 0.465 | 0.822 | -0.125 | -0.235 | -0.015 | 0.026 | 0.085 |
| AC-OH | -0.035 | -0.100 | 0.031 | 0.298 | 0.592 | 0.008 | -0.050 | 0.067 | 0.781 | 0.817 | -0.012 | -0.056 | 0.032 | 0.590 | 0.876 | -0.070 | -0.130 | -0.009 | 0.024 | 0.085 |
| Cer(d) | 0.042 | -0.036 | 0.120 | 0.292 | 0.592 | 0.047 | -0.023 | 0.117 | 0.184 | 0.498 | -0.013 | -0.066 | 0.040 | 0.638 | 0.889 | -0.060 | -0.132 | 0.013 | 0.105 | 0.224 |
| SM | -0.052 | -0.147 | 0.044 | 0.291 | 0.592 | 0.024 | -0.060 | 0.107 | 0.577 | 0.718 | 0.073 | 0.006 | 0.140 | 0.032 | 0.164 | -0.037 | -0.126 | 0.052 | 0.414 | 0.577 |
| COH | 0.042 | -0.045 | 0.129 | 0.344 | 0.633 | -0.003 | -0.077 | 0.071 | 0.935 | 0.935 | 0.007 | -0.054 | 0.068 | 0.823 | 0.970 | -0.100 | -0.180 | -0.020 | 0.014 | 0.071 |
| Cer(m) | -0.040 | -0.125 | 0.046 | 0.361 | 0.639 | -0.026 | -0.100 | 0.048 | 0.490 | 0.684 | 0.003 | -0.053 | 0.059 | 0.925 | 0.970 | -0.038 | -0.115 | 0.039 | 0.337 | 0.517 |
| PC(O) | 0.032 | -0.053 | 0.118 | 0.460 | 0.707 | 0.046 | -0.028 | 0.120 | 0.220 | 0.531 | 0.033 | -0.027 | 0.093 | 0.285 | 0.687 | -0.081 | -0.158 | -0.004 | 0.040 | 0.124 |
| FFA | -0.025 | -0.093 | 0.042 | 0.461 | 0.707 | -0.013 | -0.073 | 0.048 | 0.683 | 0.786 | 0.000 | -0.044 | 0.045 | 0.995 | 0.995 | -0.058 | -0.120 | 0.004 | 0.068 | 0.173 |
| PIP1 | 0.027 | -0.038 | 0.092 | 0.416 | 0.707 | 0.050 | -0.008 | 0.108 | 0.093 | 0.414 | 0.012 | -0.031 | 0.056 | 0.578 | 0.876 | 0.003 | -0.057 | 0.063 | 0.930 | 0.972 |
| LPE(P) | 0.025 | -0.042 | 0.092 | 0.458 | 0.707 | 0.016 | -0.043 | 0.076 | 0.587 | 0.718 | 0.019 | -0.028 | 0.066 | 0.424 | 0.780 | 0.001 | -0.061 | 0.062 | 0.986 | 0.986 |
| CE | 0.037 | -0.072 | 0.145 | 0.510 | 0.711 | 0.067 | -0.026 | 0.160 | 0.156 | 0.447 | -0.012 | -0.086 | 0.063 | 0.757 | 0.916 | -0.123 | -0.221 | -0.025 | 0.014 | 0.071 |
| PC | 0.035 | -0.067 | 0.136 | 0.504 | 0.711 | 0.078 | -0.009 | 0.165 | 0.077 | 0.414 | -0.022 | -0.095 | 0.050 | 0.547 | 0.867 | -0.108 | -0.199 | -0.017 | 0.020 | 0.085 |
| Sph | 0.022 | -0.041 | 0.086 | 0.494 | 0.711 | -0.023 | -0.079 | 0.034 | 0.435 | 0.667 | 0.007 | -0.036 | 0.050 | 0.746 | 0.916 | 0.020 | -0.039 | 0.079 | 0.509 | 0.633 |
| Ubiquinone | -0.020 | -0.087 | 0.046 | 0.548 | 0.741 | 0.021 | -0.038 | 0.080 | 0.490 | 0.684 | -0.005 | -0.051 | 0.041 | 0.845 | 0.970 | -0.002 | -0.063 | 0.060 | 0.960 | 0.982 |
| PA | 0.019 | -0.053 | 0.092 | 0.602 | 0.785 | -0.010 | -0.074 | 0.053 | 0.751 | 0.805 | 0.024 | -0.023 | 0.070 | 0.317 | 0.693 | 0.057 | -0.009 | 0.123 | 0.089 | 0.204 |
| PI | -0.022 | -0.107 | 0.063 | 0.614 | 0.785 | 0.063 | -0.013 | 0.139 | 0.103 | 0.414 | -0.002 | -0.061 | 0.057 | 0.952 | 0.973 | -0.064 | -0.143 | 0.015 | 0.112 | 0.224 |
| BA | -0.014 | -0.078 | 0.051 | 0.671 | 0.835 | 0.030 | -0.028 | 0.087 | 0.311 | 0.585 | 0.025 | -0.018 | 0.068 | 0.252 | 0.649 | 0.020 | -0.039 | 0.080 | 0.505 | 0.633 |
| methyl-CE | -0.012 | -0.086 | 0.061 | 0.745 | 0.865 | -0.012 | -0.078 | 0.055 | 0.735 | 0.805 | 0.055 | 0.005 | 0.104 | 0.031 | 0.164 | 0.034 | -0.034 | 0.103 | 0.327 | 0.517 |
| C1P | 0.014 | -0.072 | 0.100 | 0.752 | 0.865 | 0.037 | -0.039 | 0.113 | 0.343 | 0.585 | 0.003 | -0.054 | 0.059 | 0.928 | 0.970 | -0.039 | -0.116 | 0.038 | 0.324 | 0.517 |
| PS | 0.012 | -0.052 | 0.076 | 0.721 | 0.865 | 0.025 | -0.032 | 0.082 | 0.391 | 0.621 | -0.009 | -0.051 | 0.033 | 0.686 | 0.916 | 0.017 | -0.042 | 0.076 | 0.567 | 0.668 |
| PE | -0.013 | -0.102 | 0.077 | 0.783 | 0.878 | 0.044 | -0.038 | 0.125 | 0.292 | 0.584 | -0.039 | -0.101 | 0.023 | 0.219 | 0.649 | -0.053 | -0.137 | 0.030 | 0.210 | 0.358 |
| TG [NL] | -0.013 | -0.117 | 0.092 | 0.810 | 0.887 | 0.070 | -0.025 | 0.164 | 0.148 | 0.447 | -0.053 | -0.128 | 0.022 | 0.169 | 0.649 | -0.116 | -0.216 | -0.016 | 0.024 | 0.085 |
| Hex3Cer | -0.008 | -0.100 | 0.083 | 0.861 | 0.922 | 0.022 | -0.059 | 0.103 | 0.593 | 0.718 | -0.026 | -0.089 | 0.037 | 0.416 | 0.780 | -0.113 | -0.195 | -0.030 | 0.008 | 0.071 |
| AC | 0.003 | -0.063 | 0.070 | 0.924 | 0.945 | 0.029 | -0.031 | 0.089 | 0.340 | 0.585 | -0.019 | -0.064 | 0.025 | 0.390 | 0.780 | -0.062 | -0.124 | 0.000 | 0.051 | 0.137 |
| PG | -0.005 | -0.109 | 0.099 | 0.922 | 0.945 | -0.057 | -0.149 | 0.036 | 0.231 | 0.531 | -0.042 | -0.111 | 0.027 | 0.236 | 0.649 | -0.037 | -0.131 | 0.058 | 0.447 | 0.604 |
| PC(P) | 0.001 | -0.089 | 0.091 | 0.981 | 0.981 | 0.016 | -0.060 | 0.093 | 0.673 | 0.786 | 0.054 | -0.005 | 0.114 | 0.074 | 0.340 | -0.037 | -0.116 | 0.042 | 0.364 | 0.523 |
