## Supplementary Table 4 for "Circulating lipid profiles are associated with cross-sectional and longitudinal changes of central biomarkers for Alzheimer’s disease"

**Supplementary Table 4. Lipid species in modules identified with WGCNA**

| Module | Number of Lipid species | Lipid species |
|---|---|---|
| M1 | 28 | PE(15-MHDA_18:1), PE(15-MHDA_18:2), PE(15-MHDA_20:4), PE(16:0_16:1), PE(16:0_18:1), PE(16:0_18:2), PE(16:0_18:3) (a), PE(16:0_18:3) (b), PE(16:0_20:3), PE(16:0_20:4), PE(16:1_18:2), PE(16:1_20:4), PE(17:0_18:1), PE(17:0_18:2), PE(17:0_20:4), PE(18:0_18:1), PE(18:0_18:2), PE(18:0_20:3) (a), PE(18:0_20:3) (b), PE(18:0_20:4), PE(18:0_22:4), PE(18:0_22:5) (n3), PE(18:0_22:5) (n6), PE(18:1_18:1), PE(18:1_18:2), PE(20:0_20:4), PE(38:5) (a), PE(38:5) (b) |
| M2 | 60 | PE(16:0_16:0), PG(34:1), PG(36:1), PG(36:2), CE(20:4), DE(20:4), DG(14:0_16:0), DG(14:0_18:2), DG(16:0_16:0), DG(16:0_16:1), DG(16:0_18:1), DG(16:1_18:1), DG(18:0_18:1), DG(18:0_18:2), DG(18:1_18:1), TG(48:0) [NL-16:0], TG(48:0) [NL-18:0], TG(48:1) [NL-16:1], TG(48:1) [NL-18:1], TG(48:2) [NL-14:0], TG(48:2) [NL-14:1], TG(48:2) [NL-16:1], TG(48:2) [NL-18:2], TG(48:3) [NL-14:0], TG(48:3) [NL-16:1], TG(48:3) [NL-18:3], TG(49:1) [NL-16:1], TG(49:1) [NL-17:1], TG(50:0) [NL-18:0], TG(50:1) [NL-14:0], TG(50:1) [NL-16:0], TG(50:1) [NL-18:1], TG(50:2) [NL-14:0], TG(50:2) [NL-16:1], TG(50:2) [NL-18:1], TG(50:2) [NL-18:2], TG(50:3) [NL-14:0], TG(50:3) [NL-14:1], TG(50:3) [NL-16:1], TG(50:3) [NL-18:2], TG(50:3) [NL-18:3], TG(50:4) [NL-14:0], TG(50:4) [NL-18:3], TG(50:4) [NL-20:4], TG(51:0) [NL-16:0], TG(51:1) [NL-17:0], TG(51:2) [NL-15:0], TG(51:2) [NL-17:0], TG(51:2) [NL-17:1], TG(52:1) [NL-18:0], TG(52:1) [NL-18:1], TG(52:2) [NL-16:0], TG(52:2) [NL-18:2], TG(52:4) [NL-18:3], TG(53:2) [NL-17:1], TG(53:2) [NL-18:1], TG(54:0) |
| M3 | 48 | LPC(15-MHDA) [sn1] [104_sn1], LPC(15-MHDA) [sn1] & LPC(17:0) [sn2], LPC(15-MHDA) [sn2], LPC(15:0) [sn1], LPC(15:0) [sn2], LPC(16:0) [sn1], LPC(16:0) [sn2], LPC(16:1) [sn1], LPC(16:1) [sn2], LPC(17:0) [sn1], LPC(17:1) (a) [sn1] [104_sn1], LPC(17:1) [sn1] (a) & LPC(17:1) [sn2] (b), LPC(17:1) [sn1] (b), LPC(17:1) [sn2] (a), LPC(18:0) [sn1], LPC(18:0) [sn2], LPC(18:1) [sn1], LPC(18:1) [sn2], LPC(19:0) (a) [sn1] [104_sn1], LPC(19:0) [sn1] (a) & LPC(19:0) [sn2] (b), LPC(19:0) [sn1] (b), LPC(19:0) [sn2] (a), LPC(19:1) (a), LPC(19:1) (b), LPC(19:1) (c), LPC(20:1) [sn1], LPC(20:1) [sn2], LPC(20:2) [sn1], LPC(20:2) [sn2], LPC(22:1) [sn1], LPC(22:1) [sn2], LPC(O-16:0), LPC(O-18:0), LPC(O-18:1), LPC(P-16:0), LPC(P-17:0) (a), LPC(P-17:0) (b), LPC(P-18:0), LPC(P-18:1), LPE(16:0) [sn1], LPE(16:0) [sn2], LPE(18:0) [sn1], LPE(18:0) [sn2], LPE(P-16:0), LPE(P-18:0), LPE(P-18:1), LPE(P- |
| M4 | 5 | PC(18:0_22:5) (n3) & PC(20:1_20:4), PC(39:5) (b), LPC(22:5) (n3) [sn1] [104_sn1], LPC(22:5) [sn1] (n3) & LPC(22:5) [sn2] (n6), LPC(22:5) [sn2] (n3) |
| M5 | 14 | FA(14:0), FA(16:0), FA(16:1), FA(17:0), FA(17:1), FA(18:0), FA(18:1), FA(18:2), FA(18:3), FA(20:2), FA(20:3), FA(20:4), FA(22:4), FA(22:5) |
| M6 | 11 | PC(15-MHDA_20:4), PC(15:0_20:4), PC(16:0_20:4), PC(16:1_20:4), PC(17:0_20:4), PC(18:0_20:4), PC(38:5) (a), TG(56:6) [NL-20:4], TG(56:7) [NL-20:4], CE(20:4) [+OH], PC(36:4) [+OH] |
| M7 | 11 | PC(20:0_20:4), LPC(20:0) [sn1], LPC(20:0) [sn2], LPC(22:0) [sn1], LPC(22:0) [sn2], LPC(24:0) [sn1], LPC(24:0) [sn2], LPC(26:0) [sn1], LPC(26:0) [sn2], PI(20:0_20:4), AC(24:0) |
| M8 | 8 | PE(P-16:0/20:4), PE(P-17:0/20:4) (b), PE(P-18:0/20:4), PE(P-18:1/20:4) (a), PE(P-19:0/20:4) (a), PE(P-19:0/20:4) (b), PE(P-20:0/20:4), PE(P-20:1/20:4) |
| M9 | 9 | SHexCer(d18:1/24:0(OH)), SM(d16:1/23:0) & SM(d17:1/22:0), SM(d18:1/20:0) & SM(d16:1/22:0), SM(d18:1/22:0) & SM(d16:1/24:0), SM(d18:1/23:0) & SM(d17:1/24:0), SM(d18:1/24:0), SM(d18:2/22:0), SM(d18:2/23:0), SM(d18:2/24:0) |
| M10 | 11 | PC(O-32:1), PC(O-32:2), PC(O-34:4), PC(O-35:4), PC(P-15:0/20:4) (a), PC(P-15:0/20:4) (b), PC(P-17:0/20:4) (a), PC(P-17:0/20:4) (b), PE(P-15:0/20:4) (a), PE(P-15:0/20:4) (b), PE(P-17:0/20:4) (a) |
| M11 | 5 | AC(18:2), AC(18:3), AC(20:3) (a), AC(20:3) (b), AC(20:4) |
| M12 | 11 | [illegible] DE(18:1), methyl-DE(18:2), dimethyl-CE(18:1), dimethyl-CE(18:2), dimethyl-CE(20:4), dimethyl-CE(22:6) |
| M13 | 11 | CE(18:2) [+OH] |
| M14 | 5 | PI(38:5) (b), PI(15-MHDA_18:2) & PI(17:0_18:2), PI(16:0_20:3) (a), PI(18:1_18:2), PI(36:2) |
| M15 | 31 | PE(O-16:0/18:2), PE(O-16:0/20:3), PE(O-16:0/20:4), PE(O-18:0/20:4), PE(O-18:0/22:5), PE(O-18:1/18:2), PE(O-34:1), PE(O-38:5) (a), PE(P-16:0/18:1), PE(P-16:0/18:2), PE(P-16:0/18:3), PE(P-16:0/20:3) (a), PE(P-16:0/20:3) (b), PE(P-16:0/22:5) (n3), PE(P-18:0/18:1), PE(P-18:0/18:2), PE(P-18:0/18:3), PE(P-18:0/20:3) (a), PE(P-18:0/20:3) (b), PE(P-18:0/22:5) (n3), PE(P-18:1/18:1) (a), PE(P-18:1/18:1) (b), PE(P-18:1/18:2) (a), PE(P-18:1/18:2) (b), PE(P-18:1/18:3), PE(P-18:1/20:3) (a), PE(P-18:1/20:3) (b), PE(P-18:1/20:4) (b), PE(P-18:1/22:5) (a), PE(P-20:0/18:1), PE(P-20:0/18:2) |

| | | |
|---|---|---|
| M16 | 19 | 50:2) [NL-18:1], TG(O-50:2) [NL-18:2], TG(O-50:3) [NL-18:2], TG(O-52:0) [NL-16:0], TG(O-52:1) [NL-16:0], TG(O-52:1) [NL-18:1], TG(O-52:2) [NL-16:0], TG(O-52:2) [NL-17:1], TG(O-52:2) [NL-18:1], TG(O-54:2) [NL-17:1], TG(O-54:2) [NL-18:1], TG(O-54:3) [NL-17:1], TG(O-54:3) [NL-18:1], TG(O-54:4) [NL-17:1], TG(O-54:4) [NL-18:2] |
| M17 | 12 | Cer(d19:1/24:0), Cer(d19:1/24:1), Cer(d19:1/26:0), Cer(d20:1/22:0), Cer(d20:1/23:0), Cer(d20:1/24:0), Cer(d20:1/24:1) |
| M18 | 5 | CE(16:2), CE(18:3), DE(18:2), deDE(18:2), deDE(20:4) |
| M19 | 14 | HexCer(d18:1/16:0), HexCer(d18:1/18:0), HexCer(d18:1/20:0), HexCer(d18:1/22:0), HexCer(d18:1/24:0), HexCer(d18:1/24:1), HexCer(d18:2/18:0), HexCer(d18:2/20:0), HexCer(d18:2/22:0), HexCer(d18:2/24:0) |
| M20 | 6 | LPI(18:1) [sn1], LPI(18:1) [sn2], LPI(18:2) [sn1], LPI(18:2) [sn2], LPI(20:4) [sn1], LPI(20:4) [sn2] |
| M21 | 12 | LPC(22:4) [sn2], LPC(22:5) [sn1] (n6), LPE(20:4) [sn1], LPE(20:4) [sn2], PI(18:0_22:4), PI(18:0_22:5) (n6) |
| M22 | 6 | (b) |
| M23 | 12 | SM(d16:1/20:0), SM(d18:2/16:0), SM(d18:2/17:0), SM(d18:2/18:0), SM(d18:2/18:1), SM(d18:2/20:0) |
| M24 | 23 | 36:5), PC(P-16:0/20:5), LPC(20:5) [sn1], LPC(20:5) [sn2], PE(16:0_20:5), PE(O-36:5), PE(P-15:0/22:6) (a), PE(P-16:0/20:5), PE(P-18:0/20:5), PE(P-18:1/20:5) (a), PE(P-18:1/20:5) (b), LPE(22:6) [sn1], LPE(22:6) [sn2], CE(20:5), DE(20:5), FA(20:5) |
| M25 | 7 | AC(15:0) (b), AC(16:0), AC(17:0) (a), AC(17:0) (b), AC(18:0), AC(24:1), AC(26:1) |
| M26 | 14 | AC(12:0), AC(12:1), AC(14:0), AC(14:1), AC(14:2), AC(16:1), AC(18:1), AC(14:0)-OH, AC(14:1)-OH, AC(16:0)-OH, AC(16:1)-OH, AC(18:0)-OH, AC(18:1)-OH, AC(20:3)-OH |
| M27 | 11 | Cer(m18:1/18:0), Cer(m18:1/20:0), Cer(m18:1/22:0), Cer(m18:1/23:0), Cer(m18:1/24:0), Cer(m18:1/24:1) |
| M28 | 7 | SM(d19:1/24:1) |
| M29 | 10 | DG(16:0_20:4), DG(18:0_20:4), DG(18:1_20:3), DG(18:1_20:4), DG(18:2_20:4), TG(52:5) [NL-20:4], TG(54:4) [NL-20:3], TG(54:5) [NL-20:4], TG(54:6) [NL-20:4], TG(56:8) [NL-20:4] |
| M30 | 26 | PC(28:0), PC(32:1), PC(32:2), LPC(14:0) [sn1], LPC(14:0) [sn2], PI(15-MHDA_18:1) & PI(17:0_18:1), PI(15-MHDA_20:4) & PI(17:0_20:4), PI(16:0_16:1), PI(16:0_20:3) (b), PI(16:0_20:4), PI(16:0/16:0), PI(18:0_18:1), PI(18:0_20:3) (b), PI(18:0_20:4), PI(34:0), PI(34:1), PI(38:5) (a), CE(14:0), CE(16:1), PC(34:2) [+OH] |
| M31 | 7 | CE(20:3) |
| M32 | 20 | S1P(d16:1), Cer(d16:1/16:0), Cer(d16:1/22:0), Cer(d16:1/23:0), Cer(d16:1/24:0), Cer(d16:1/24:1), Cer(d17:1/22:0), Cer(d17:1/23:0), Cer(d17:1/24:0), Cer(d17:1/24:1), Cer(d18:1/22:0), Cer(d18:1/23:0), Cer(d18:1/24:0), Cer(d18:1/24:1), Cer(d18:1/26:0), Cer(d18:2/22:0), Cer(d18:2/23:0), Cer(d18:2/24:0), Cer(d18:2/24:1), Cer(d18:2/26:0) |
| M33 | 30 | DG(16:0_22:5), DG(16:0_22:6), DG(18:1_20:5), DG(18:1_22:5), DG(18:1_22:6), DG(18:2_22:6), TG(52:5) [NL-20:5], TG(54:6) [NL-20:5], TG(54:6) [NL-22:6], TG(54:7) [NL-20:5], TG(54:7) [NL-22:6], TG(56:6) [NL-22:5], TG(56:7) [NL-20:5], TG(56:7) [NL-22:5], TG(56:7) [NL-22:6], TG(56:8) [NL-20:5], TG(56:8) [NL-22:6], TG(56:9) [NL-22:6], TG(58:10) [NL-22:6], TG(58:8) [NL-22:6], TG(58:9) [NL-22:6] |
| M34 | 11 | [sn2] (b), LPC(18:3) [sn1] (b), LPC(18:3) [sn2] (a), LPE(18:1) [sn1], LPE(18:1) [sn2], LPE(18:2) [sn1], LPE(18:2) [sn2], LPC(18:2) [+OH] |
| M35 | 10 | PC(O-16:0/20:4), PC(O-18:0/20:4), PC(O-38:5), PC(O-40:5), PC(P-16:0/20:4), PC(P-18:0/20:4), PC(P-18:0/22:5), PC(P-20:0/20:4), PC(P-38:5) (a), PC(P-38:5) (b) |
| M36 | 14 | DG(16:0_18:2), DG(18:1_18:2), DG(18:1_18:3), DG(18:2_18:2), TG(52:3) [NL-16:1], TG(52:3) [NL-18:2], TG(52:4) [NL-16:1], TG(52:4) [NL-18:2], TG(52:5) [NL-18:3], TG(54:3) [NL-18:1], TG(54:3) [NL-18:2], TG(54:4) [NL-18:2], TG(54:5) [NL-18:3], TG(54:6) [NL-18:3] |
| M37 | 8 | dhCer(d18:0/16:0), dhCer(d18:0/18:0), dhCer(d18:0/20:0), dhCer(d18:0/22:0), dhCer(d18:0/24:0), dhCer(d18:0/24:1), SM(41:0), SM(d18:0/22:0) |
| M38 | 10 | Hex2Cer(d18:1/22:0), Hex2Cer(d18:1/24:0), Hex2Cer(d18:1/24:1), Hex2Cer(d18:2/16:0), Hex2Cer(d18:2/24:1), GM1(d18:1/16:0) |
| M39 | 8 | 20:0) |
| M40 | 10 | Sph(d18:1), S1P(d18:0), S1P(d18:1), S1P(d18:2), PS(36:1), PS(36:2), PS(38:3), PS(38:4), LPC(20:4) [+OH], LPC(22:6) [+OH] |

| | | |
|---|---|---|
| M41 | 19 | SM(d18:0/14:0), SM(d18:1/14:0) & SM(d16:1/16:0), SM(d18:2/14:0), PC(15-MHDA_18:1), PC(17:0_18:1), PC(31:0) (a), PC(31:0) (b), PC(33:0) (a), PC(33:0) (b), PC(33:1), PC(P-16:0/14:0), CE(15:0), CE(17:0), CE(17:1) |
| M42 | 62 | Cer(d18:1/16:0), Cer(d18:2/16:0), Cer1P(d18:1/16:0), Hex3Cer(d18:1/16:0), Hex3Cer(d18:1/18:0), Hex3Cer(d18:1/20:0), Hex3Cer(d18:1/22:0), Hex3Cer(d18:1/24:0), Hex3Cer(d18:1/24:1), GM3(d18:1/16:0), GM3(d18:1/18:0), GM3(d18:1/22:0), GM3(d18:1/24:0), GM3(d18:1/24:1), SHexCer(d18:1/16:0(OH)), SHexCer(d18:1/16:0), SHexCer(d18:1/24:1(OH)), SM(35:2) (b), SM(d18:0/16:0), SM(d18:1/16:0), PC(15-MHDA_18:2), PC(15:0_20:3), PC(16:0_16:0), PC(16:0_18:0), PC(16:0_18:2), PC(16:0_18:3) (a), PC(16:1_18:2), PC(17:0_18:2), PC(17:1_18:2), PC(18:0_18:2), PC(18:1_18:1), PC(18:1_18:2), PC(18:2_18:2), PC(33:2), PC(38:2), PC(38:6) (a), PC(40:8), PC(O-16:0/16:0), PC(O-16:0/20:3), PC(O-18:0/18:1), PC(O-18:0/18:2), PC(O-18:1/18:1), PC(O-18:1/18:2), PC(O-34:1), PC(O-34:2), PC(O-36:0), PC(P-16:0/16:0), PC(P-16:0/16:1), PC(P-16:0/18:0), PC(P-16:0/18:1), PC(P-16:0/18:2), PC(P-16:0/18:3), PC(P-18:0/18:2), PC(P-18:1/18:1), PC(P-35:2) (a), PC(P-35:2) (b), PC(P-36:3), PI(18:0_20:2), CE(16:0), CE(18:1), CE(18:2), COH |
| M43 | 9 | PE(O-16:0/22:4), PE(P-16:0/22:4), PE(P-16:0/22:5) (n6), PE(P-18:0/22:4), PE(P-18:0/22:5) (n6), PE(P-18:1/22:4), PE(P-18:1/22:5) (b), CE(22:4), CE(22:5) |
| M44 | 11 | Cer(d18:1/19:0), Cer(d18:1/20:0), Cer(d18:1/21:0), Cer(d18:2/18:0), Cer(d18:2/20:0), Cer(d18:2/21:0) |
| M45 | 39 | [illegible] PC(18:1_22:6) (a), PC(18:1_22:6) (b), PC(38:7) (c), PC(39:5) (a), PC(44:12), PC(O-16:0/22:6), PC(O-18:0/22:6), PC(P-16:0/22:6), PC(P-18:0/22:6), PC(P-18:1/22:6), LPC(22:6) [sn1], LPC(22:6) [sn2], PE(O-16:0/22:6), PE(O-18:0/22:6), PE(O-18:1/22:6), PE(O-38:5) (b), PE(P-15:0/22:6) (b), PE(P-16:0/22:6), PE(P-17:0/22:6) (a), PE(P-17:0/22:6) (b), PE(P-18:0/22:6), PE(P-18:1/22:6) (a), PE(P-18:1/22:6) (b), PE(P-20:0/22:6), PE(P-20:1/22:6), PI(18:0_22:5) (n3), PI(18:0_22:6), PI(37:6), PI(38:6), PI(39:6), CE(22:6), DE(22:6), CE(22:6) [+OH] |
| M46 | 8 | SHexCer(d18:1/24:1), SM(43:2) (b), SM(43:2) (c), SM(44:2), SM(44:3), SM(d16:1/24:1), SM(d17:1/24:1), SM(d18:1/24:1) |
