## Supplementary Table 5 for "Circulating lipid profiles are associated with cross-sectional and longitudinal changes of central biomarkers for Alzheimer’s disease"

**Supplementary Table 5. Associations of lipid correlation network modules with baseline A/T/N biomarkers**

| | A: Amyloid PET (AV45) uptake | | | | | T: CSF pTau | | | | | N1: Hippocampal volume | | | | | N2: FDG uptake | | | | |
|---|---|---|---|---|---|---|---|---|---|---|---|---|---|---|---|---|---|---|---|---|
| Lipid modu | Beta | 95% CI (Lower) | 95% CI (Upper) | Pvalue | Pvalue(BH) | Beta | 95% CI (Lower) | 95% CI (Upper) | Pvalue | Pvalue(BH) | Beta | 95% CI (Lower) | 95% CI (Upper) | Pvalue | Pvalue(BH) | Beta | 95% CI (Lower) | 95% CI (Upper) | Pvalue | Pvalue(BH) |
| M39 | 0.122 | 0.053 | 0.192 | 0.001 | 0.026 | 0.053 | -0.010 | 0.116 | 0.100 | 0.229 | -0.037 | -0.088 | 0.013 | 0.149 | 0.395 | -0.110 | -0.175 | -0.045 | 0.001 | 0.042 |
| M34 | 0.112 | 0.043 | 0.180 | 0.001 | 0.033 | 0.079 | 0.018 | 0.141 | 0.011 | 0.102 | -0.014 | -0.060 | 0.033 | 0.559 | 0.857 | -0.027 | -0.092 | 0.037 | 0.403 | 0.530 |
| M3 | 0.104 | 0.033 | 0.174 | 0.004 | 0.047 | 0.048 | -0.015 | 0.111 | 0.135 | 0.239 | -0.009 | -0.061 | 0.043 | 0.723 | 0.890 | -0.065 | -0.131 | 0.000 | 0.051 | 0.148 |
| M38 | 0.118 | 0.038 | 0.198 | 0.004 | 0.047 | 0.058 | -0.012 | 0.129 | 0.104 | 0.229 | -0.036 | -0.088 | 0.017 | 0.181 | 0.438 | -0.063 | -0.137 | 0.011 | 0.097 | 0.235 |
| M37 | -0.108 | -0.188 | -0.028 | 0.008 | 0.075 | -0.049 | -0.119 | 0.021 | 0.171 | 0.271 | 0.058 | 0.005 | 0.111 | 0.031 | 0.128 | 0.032 | -0.042 | 0.106 | 0.396 | 0.530 |
| M8 | -0.090 | -0.160 | -0.020 | 0.012 | 0.090 | -0.059 | -0.121 | 0.002 | 0.060 | 0.188 | 0.068 | 0.022 | 0.114 | 0.004 | 0.035 | 0.008 | -0.056 | 0.072 | 0.808 | 0.909 |
| M19 | 0.089 | 0.015 | 0.163 | 0.018 | 0.121 | 0.024 | -0.041 | 0.090 | 0.465 | 0.578 | -0.016 | -0.065 | 0.033 | 0.515 | 0.816 | -0.018 | -0.086 | 0.051 | 0.609 | 0.737 |
| M11 | 0.066 | 0.001 | 0.132 | 0.047 | 0.271 | 0.055 | -0.004 | 0.113 | 0.068 | 0.196 | -0.024 | -0.068 | 0.020 | 0.279 | 0.569 | -0.033 | -0.094 | 0.028 | 0.286 | 0.454 |
| M4 | 0.070 | -0.001 | 0.141 | 0.055 | 0.279 | 0.070 | 0.008 | 0.133 | 0.028 | 0.143 | -0.059 | -0.107 | -0.012 | 0.015 | 0.069 | -0.033 | -0.099 | 0.032 | 0.318 | 0.488 |
| M36 | 0.083 | -0.005 | 0.170 | 0.064 | 0.292 | 0.056 | -0.024 | 0.137 | 0.169 | 0.271 | -0.028 | -0.094 | 0.039 | 0.412 | 0.702 | -0.066 | -0.150 | 0.018 | 0.121 | 0.266 |
| M9 | -0.088 | -0.188 | 0.012 | 0.085 | 0.292 | -0.001 | -0.088 | 0.087 | 0.984 | 0.984 | 0.103 | 0.036 | 0.171 | 0.003 | 0.032 | 0.069 | -0.025 | 0.163 | 0.148 | 0.310 |
| M7 | 0.068 | -0.008 | 0.144 | 0.081 | 0.292 | -0.006 | -0.074 | 0.062 | 0.859 | 0.919 | 0.028 | -0.022 | 0.078 | 0.274 | 0.569 | 0.042 | -0.027 | 0.111 | 0.236 | 0.402 |
| M32 | 0.071 | -0.010 | 0.151 | 0.085 | 0.292 | 0.056 | -0.016 | 0.127 | 0.125 | 0.239 | -0.011 | -0.065 | 0.043 | 0.682 | 0.871 | -0.042 | -0.115 | 0.032 | 0.270 | 0.443 |
| M33 | -0.063 | -0.136 | 0.010 | 0.089 | 0.292 | -0.079 | -0.143 | -0.014 | 0.018 | 0.135 | 0.030 | -0.019 | 0.078 | 0.232 | 0.533 | 0.008 | -0.059 | 0.076 | 0.810 | 0.909 |
| M42 | 0.090 | -0.022 | 0.203 | 0.115 | 0.352 | 0.083 | -0.011 | 0.178 | 0.083 | 0.214 | 0.018 | -0.064 | 0.100 | 0.666 | 0.871 | -0.125 | -0.223 | -0.028 | 0.012 | 0.079 |
| M15 | -0.056 | -0.131 | 0.019 | 0.143 | 0.410 | -0.008 | -0.074 | 0.059 | 0.821 | 0.910 | 0.079 | 0.027 | 0.130 | 0.003 | 0.032 | 0.029 | -0.039 | 0.098 | 0.402 | 0.530 |
| M16 | -0.051 | -0.123 | 0.020 | 0.157 | 0.412 | -0.050 | -0.114 | 0.014 | 0.125 | 0.239 | 0.119 | 0.070 | 0.169 | 0.000 | 0.000 | 0.017 | -0.050 | 0.083 | 0.626 | 0.739 |
| M40 | 0.046 | -0.018 | 0.109 | 0.161 | 0.412 | 0.055 | -0.003 | 0.112 | 0.061 | 0.188 | -0.016 | -0.059 | 0.028 | 0.479 | 0.787 | -0.001 | -0.060 | 0.058 | 0.976 | 0.983 |
| M18 | 0.070 | -0.040 | 0.181 | 0.211 | 0.477 | 0.112 | 0.017 | 0.207 | 0.021 | 0.135 | -0.138 | -0.205 | -0.071 | 0.000 | 0.001 | -0.100 | -0.196 | -0.005 | 0.039 | 0.139 |
| M10 | -0.046 | -0.116 | 0.025 | 0.202 | 0.477 | 0.001 | -0.060 | 0.063 | 0.966 | 0.984 | 0.058 | 0.012 | 0.105 | 0.014 | 0.069 | -0.004 | -0.068 | 0.060 | 0.895 | 0.980 |
| M45 | -0.049 | -0.126 | 0.029 | 0.218 | 0.477 | -0.105 | -0.173 | -0.038 | 0.002 | 0.053 | 0.069 | 0.017 | 0.121 | 0.009 | 0.054 | 0.002 | -0.068 | 0.072 | 0.952 | 0.983 |
| M20 | -0.039 | -0.107 | 0.029 | 0.260 | 0.543 | 0.046 | -0.014 | 0.105 | 0.135 | 0.239 | 0.007 | -0.040 | 0.054 | 0.775 | 0.891 | -0.045 | -0.108 | 0.018 | 0.160 | 0.317 |
| M21 | 0.035 | -0.040 | 0.109 | 0.361 | 0.554 | 0.113 | 0.047 | 0.178 | 0.001 | 0.036 | -0.041 | -0.094 | 0.011 | 0.123 | 0.378 | -0.101 | -0.170 | -0.031 | 0.005 | 0.077 |
| M43 | -0.037 | -0.111 | 0.037 | 0.331 | 0.554 | 0.091 | 0.025 | 0.156 | 0.007 | 0.080 | 0.000 | -0.054 | 0.054 | 0.996 | 0.996 | -0.083 | -0.152 | -0.014 | 0.018 | 0.104 |
| M12 | -0.036 | -0.111 | 0.039 | 0.345 | 0.554 | -0.032 | -0.100 | 0.036 | 0.356 | 0.468 | 0.073 | 0.023 | 0.123 | 0.005 | 0.035 | 0.070 | 0.001 | 0.140 | 0.047 | 0.146 |
| M13 | 0.053 | -0.046 | 0.151 | 0.293 | 0.554 | 0.023 | -0.062 | 0.108 | 0.594 | 0.683 | 0.011 | -0.053 | 0.075 | 0.741 | 0.890 | -0.081 | -0.170 | 0.009 | 0.077 | 0.197 |
| M27 | -0.040 | -0.125 | 0.046 | 0.361 | 0.554 | -0.026 | -0.100 | 0.048 | 0.490 | 0.593 | 0.003 | -0.053 | 0.059 | 0.925 | 0.967 | -0.038 | -0.115 | 0.039 | 0.337 | 0.493 |
| M24 | -0.037 | -0.111 | 0.037 | 0.326 | 0.554 | -0.066 | -0.133 | 0.001 | 0.053 | 0.188 | 0.013 | -0.037 | 0.062 | 0.616 | 0.864 | 0.022 | -0.047 | 0.091 | 0.534 | 0.682 |
| M28 | -0.034 | -0.106 | 0.038 | 0.351 | 0.554 | -0.002 | -0.065 | 0.062 | 0.962 | 0.984 | 0.064 | 0.016 | 0.112 | 0.009 | 0.054 | 0.019 | -0.049 | 0.087 | 0.583 | 0.725 |
| M22 | -0.041 | -0.122 | 0.040 | 0.324 | 0.554 | -0.077 | -0.150 | -0.004 | 0.038 | 0.174 | 0.014 | -0.041 | 0.069 | 0.620 | 0.864 | -0.001 | -0.076 | 0.074 | 0.983 | 0.983 |
| M14 | 0.032 | -0.046 | 0.110 | 0.417 | 0.619 | 0.102 | 0.032 | 0.171 | 0.004 | 0.064 | -0.012 | -0.067 | 0.043 | 0.676 | 0.871 | -0.077 | -0.148 | -0.005 | 0.036 | 0.136 |
| M5 | -0.024 | -0.091 | 0.043 | 0.487 | 0.700 | -0.007 | -0.067 | 0.054 | 0.831 | 0.910 | -0.003 | -0.048 | 0.041 | 0.886 | 0.948 | -0.063 | -0.125 | -0.001 | 0.048 | 0.146 |
| M26 | -0.020 | -0.085 | 0.045 | 0.549 | 0.765 | 0.030 | -0.028 | 0.089 | 0.309 | 0.431 | -0.021 | -0.065 | 0.023 | 0.339 | 0.624 | -0.080 | -0.141 | -0.019 | 0.010 | 0.077 |
| M23 | -0.023 | -0.110 | 0.064 | 0.600 | 0.812 | 0.072 | -0.002 | 0.146 | 0.057 | 0.188 | 0.005 | -0.058 | 0.068 | 0.867 | 0.948 | -0.102 | -0.179 | -0.024 | 0.010 | 0.077 |
| M29 | -0.022 | -0.113 | 0.068 | 0.628 | 0.826 | 0.038 | -0.042 | 0.118 | 0.354 | 0.468 | 0.000 | -0.063 | 0.062 | 0.990 | 0.996 | -0.081 | -0.166 | 0.004 | 0.061 | 0.166 |
| M31 | 0.019 | -0.063 | 0.101 | 0.655 | 0.837 | 0.081 | 0.010 | 0.151 | 0.026 | 0.143 | 0.042 | -0.015 | 0.100 | 0.150 | 0.395 | -0.001 | -0.077 | 0.074 | 0.971 | 0.983 |
| M41 | 0.015 | -0.068 | 0.097 | 0.724 | 0.900 | 0.054 | -0.019 | 0.127 | 0.144 | 0.246 | -0.015 | -0.070 | 0.041 | 0.604 | 0.864 | -0.050 | -0.126 | 0.026 | 0.196 | 0.347 |
| M44 | 0.012 | -0.063 | 0.087 | 0.748 | 0.905 | 0.042 | -0.025 | 0.109 | 0.219 | 0.324 | -0.024 | -0.074 | 0.027 | 0.362 | 0.641 | -0.049 | -0.119 | 0.020 | 0.165 | 0.317 |
| M35 | -0.006 | -0.086 | 0.074 | 0.887 | 0.913 | 0.055 | -0.014 | 0.125 | 0.118 | 0.239 | 0.031 | -0.026 | 0.087 | 0.284 | 0.569 | -0.098 | -0.170 | -0.026 | 0.008 | 0.077 |
| M6 | -0.010 | -0.086 | 0.065 | 0.788 | 0.913 | 0.058 | -0.008 | 0.124 | 0.084 | 0.214 | -0.008 | -0.060 | 0.044 | 0.755 | 0.890 | -0.095 | -0.165 | -0.026 | 0.007 | 0.077 |
| M2 | -0.009 | -0.116 | 0.097 | 0.862 | 0.913 | 0.062 | -0.034 | 0.159 | 0.204 | 0.313 | -0.067 | -0.142 | 0.008 | 0.081 | 0.294 | -0.116 | -0.218 | -0.015 | 0.025 | 0.114 |
| M30 | 0.006 | -0.080 | 0.092 | 0.890 | 0.913 | 0.079 | 0.003 | 0.155 | 0.042 | 0.174 | -0.046 | -0.105 | 0.013 | 0.123 | 0.378 | -0.093 | -0.173 | -0.013 | 0.023 | 0.114 |
| M25 | -0.006 | -0.073 | 0.062 | 0.868 | 0.913 | 0.023 | -0.037 | 0.084 | 0.451 | 0.576 | -0.023 | -0.068 | 0.022 | 0.319 | 0.612 | -0.069 | -0.132 | -0.007 | 0.030 | 0.127 |
| M17 | 0.006 | -0.062 | 0.075 | 0.855 | 0.913 | 0.017 | -0.044 | 0.078 | 0.585 | 0.683 | 0.005 | -0.041 | 0.051 | 0.832 | 0.934 | -0.052 | -0.115 | 0.011 | 0.106 | 0.244 |
| M1 | -0.006 | -0.093 | 0.081 | 0.893 | 0.913 | 0.066 | -0.014 | 0.145 | 0.105 | 0.229 | -0.044 | -0.105 | 0.017 | 0.155 | 0.395 | -0.057 | -0.139 | 0.025 | 0.174 | 0.320 |
| M46 | 0.002 | -0.086 | 0.089 | 0.968 | 0.968 | -0.045 | -0.121 | 0.032 | 0.251 | 0.361 | 0.050 | -0.007 | 0.108 | 0.083 | 0.294 | -0.039 | -0.121 | 0.042 | 0.343 | 0.493 |
