## Supplementary Table 6 for "Circulating lipid profiles are associated with cross-sectional and longitudinal changes of central biomarkers for Alzheimer’s disease"

**Supplementary Table 6. Associations of lipid species with longitudinal A/T/N biomarkers**

| | A: Amyloid PET (AV45) uptake | | | | | T: CSF pTau | | | | | N1: Hippocampal volume | | | | | N2: FDG uptake | | | | |
|---|---|---|---|---|---|---|---|---|---|---|---|---|---|---|---|---|---|---|---|---|
| Lipid Species | Beta | 95% CI (Lower) | 95% CI (Upper) | Pvalue | Pvalue(BH) | Beta | 95% CI (Lower) | 95% CI (Upper) | Pvalue | Pvalue(BH) | Beta | 95% CI (Lower) | 95% CI (Upper) | Pvalue | Pvalue(BH) | Beta | 95% CI (Lower) | 95% CI (Upper) | Pvalue | Pvalue(BH) |
| deDE(20:4) | -0.002 | -0.024 | 0.020 | 0.859 | 0.981 | 0.008 | -0.006 | 0.023 | 0.255 | 0.506 | -0.026 | -0.037 | -0.015 | 0.000 | 0.004 | -0.048 | -0.077 | -0.019 | 0.001 | 0.018 |
| PI(38:5) (b) | -0.017 | -0.039 | 0.005 | 0.136 | 0.903 | -0.009 | -0.023 | 0.005 | 0.222 | 0.482 | -0.024 | -0.035 | -0.013 | 0.000 | 0.005 | -0.058 | -0.084 | -0.031 | 0.000 | 0.004 |
| PC(18:1_22:6) (b) | -0.002 | -0.024 | 0.021 | 0.890 | 0.981 | -0.009 | -0.023 | 0.005 | 0.231 | 0.488 | -0.023 | -0.034 | -0.012 | 0.000 | 0.008 | -0.067 | -0.094 | -0.040 | 0.000 | 0.001 |
| deDE(18:2) | 0.000 | -0.022 | 0.023 | 0.975 | 0.995 | -0.007 | -0.021 | 0.007 | 0.314 | 0.557 | -0.022 | -0.032 | -0.011 | 0.000 | 0.013 | -0.049 | -0.077 | -0.022 | 0.000 | 0.010 |
| GM3(d18:1/22:0) | 0.000 | -0.022 | 0.021 | 0.976 | 0.995 | -0.007 | -0.021 | 0.007 | 0.336 | 0.574 | -0.021 | -0.032 | -0.011 | 0.000 | 0.018 | -0.056 | -0.083 | -0.028 | 0.000 | 0.005 |
| AC(18:2) | -0.008 | -0.030 | 0.014 | 0.496 | 0.922 | -0.007 | -0.021 | 0.007 | 0.315 | 0.557 | -0.021 | -0.031 | -0.010 | 0.000 | 0.020 | -0.026 | -0.053 | 0.001 | 0.064 | 0.184 |
| LPC(O-24:2) | -0.013 | -0.035 | 0.009 | 0.262 | 0.903 | -0.016 | -0.030 | -0.003 | 0.020 | 0.273 | -0.020 | -0.031 | -0.009 | 0.000 | 0.022 | -0.036 | -0.063 | -0.008 | 0.012 | 0.066 |
| LPC(O-22:1) | -0.004 | -0.026 | 0.018 | 0.693 | 0.964 | -0.022 | -0.036 | -0.008 | 0.002 | 0.259 | -0.020 | -0.031 | -0.009 | 0.000 | 0.022 | -0.044 | -0.071 | -0.017 | 0.002 | 0.022 |
| GM3(d18:1/16:0) | -0.001 | -0.023 | 0.021 | 0.949 | 0.995 | -0.007 | -0.021 | 0.008 | 0.352 | 0.592 | -0.020 | -0.031 | -0.009 | 0.000 | 0.022 | -0.057 | -0.084 | -0.030 | 0.000 | 0.004 |
| GM3(d18:1/20:0) | -0.007 | -0.029 | 0.015 | 0.527 | 0.922 | -0.009 | -0.023 | 0.006 | 0.240 | 0.495 | -0.020 | -0.031 | -0.009 | 0.000 | 0.030 | -0.052 | -0.080 | -0.024 | 0.000 | 0.008 |
| LPC(O-20:1) | -0.004 | -0.026 | 0.018 | 0.716 | 0.964 | -0.019 | -0.033 | -0.005 | 0.007 | 0.259 | -0.019 | -0.030 | -0.009 | 0.000 | 0.030 | -0.039 | -0.066 | -0.011 | 0.006 | 0.047 |
| AC(18:1) | 0.002 | -0.020 | 0.024 | 0.841 | 0.981 | -0.016 | -0.030 | -0.002 | 0.024 | 0.285 | -0.019 | -0.030 | -0.008 | 0.001 | 0.030 | -0.051 | -0.078 | -0.024 | 0.000 | 0.008 |
| Hex2Cer(d18:1/20:0) | 0.002 | -0.020 | 0.024 | 0.858 | 0.981 | 0.007 | -0.007 | 0.021 | 0.332 | 0.572 | -0.019 | -0.030 | -0.008 | 0.001 | 0.030 | -0.038 | -0.066 | -0.010 | 0.007 | 0.049 |
| LPC(O-24:1) | -0.011 | -0.033 | 0.011 | 0.330 | 0.922 | -0.026 | -0.040 | -0.013 | 0.000 | 0.092 | -0.018 | -0.029 | -0.008 | 0.001 | 0.043 | -0.043 | -0.070 | -0.016 | 0.002 | 0.025 |
| HexCer(d18:1/20:0) | 0.006 | -0.016 | 0.028 | 0.575 | 0.933 | -0.012 | -0.026 | 0.002 | 0.093 | 0.359 | -0.018 | -0.029 | -0.008 | 0.001 | 0.043 | -0.040 | -0.068 | -0.013 | 0.004 | 0.042 |
| PE(P-15:0/22:6) (b) | -0.005 | -0.028 | 0.017 | 0.647 | 0.960 | 0.001 | -0.013 | 0.015 | 0.881 | 0.934 | 0.018 | 0.007 | 0.029 | 0.001 | 0.051 | 0.008 | -0.019 | 0.036 | 0.551 | 0.730 |
| SM(d18:2/18:1) | -0.003 | -0.025 | 0.019 | 0.766 | 0.969 | 0.001 | -0.013 | 0.015 | 0.915 | 0.951 | -0.018 | -0.029 | -0.007 | 0.001 | 0.059 | -0.055 | -0.081 | -0.028 | 0.000 | 0.005 |
| PE(P-15:0/22:6) (a) | -0.001 | -0.024 | 0.022 | 0.929 | 0.995 | -0.006 | -0.019 | 0.008 | 0.429 | 0.661 | 0.017 | 0.006 | 0.028 | 0.002 | 0.076 | 0.012 | -0.015 | 0.039 | 0.396 | 0.611 |
| LPC(18:1) [sn1] | -0.006 | -0.028 | 0.016 | 0.574 | 0.933 | -0.016 | -0.029 | -0.002 | 0.026 | 0.285 | -0.017 | -0.028 | -0.006 | 0.002 | 0.081 | -0.037 | -0.065 | -0.010 | 0.007 | 0.049 |
| LPC(18:1) [sn2] | -0.004 | -0.026 | 0.018 | 0.724 | 0.964 | -0.017 | -0.031 | -0.004 | 0.012 | 0.273 | -0.017 | -0.028 | -0.006 | 0.002 | 0.081 | -0.041 | -0.068 | -0.014 | 0.003 | 0.037 |
| LPC(O-18:0) | -0.012 | -0.034 | 0.010 | 0.275 | 0.903 | -0.011 | -0.025 | 0.003 | 0.125 | 0.385 | -0.017 | -0.027 | -0.006 | 0.002 | 0.083 | -0.036 | -0.063 | -0.008 | 0.011 | 0.063 |
| AC(20:4) | -0.009 | -0.031 | 0.013 | 0.444 | 0.922 | -0.009 | -0.022 | 0.005 | 0.193 | 0.454 | -0.017 | -0.027 | -0.006 | 0.002 | 0.083 | -0.027 | -0.054 | -0.001 | 0.046 | 0.155 |
| GM3(d18:1/24:1) | -0.015 | -0.037 | 0.007 | 0.185 | 0.903 | -0.021 | -0.036 | -0.007 | 0.003 | 0.259 | -0.017 | -0.027 | -0.006 | 0.003 | 0.084 | -0.058 | -0.086 | -0.031 | 0.000 | 0.004 |
| LPC(20:2) [sn1] | -0.012 | -0.034 | 0.010 | 0.281 | 0.903 | -0.011 | -0.025 | 0.003 | 0.111 | 0.380 | -0.016 | -0.027 | -0.006 | 0.003 | 0.084 | -0.021 | -0.048 | 0.005 | 0.118 | 0.293 |
| Hex3Cer(d18:1/16:0) | -0.006 | -0.028 | 0.016 | 0.574 | 0.933 | -0.012 | -0.026 | 0.002 | 0.107 | 0.375 | -0.017 | -0.027 | -0.006 | 0.003 | 0.084 | -0.060 | -0.086 | -0.033 | 0.000 | 0.004 |
| AC(18:3) | 0.002 | -0.020 | 0.024 | 0.842 | 0.981 | -0.002 | -0.016 | 0.012 | 0.792 | 0.891 | -0.016 | -0.027 | -0.006 | 0.003 | 0.084 | -0.018 | -0.045 | 0.010 | 0.205 | 0.416 |
| PI(16:0_20:3) (a) | -0.019 | -0.041 | 0.003 | 0.089 | 0.903 | -0.008 | -0.022 | 0.006 | 0.279 | 0.524 | -0.016 | -0.027 | -0.005 | 0.003 | 0.090 | -0.035 | -0.062 | -0.008 | 0.010 | 0.061 |
| LPC(P-20:0) | -0.009 | -0.031 | 0.013 | 0.444 | 0.922 | -0.008 | -0.022 | 0.006 | 0.282 | 0.527 | -0.016 | -0.027 | -0.005 | 0.003 | 0.090 | -0.026 | -0.053 | 0.001 | 0.063 | 0.184 |
| LPC(O-20:0) | -0.010 | -0.032 | 0.012 | 0.374 | 0.922 | -0.017 | -0.031 | -0.003 | 0.016 | 0.273 | -0.016 | -0.027 | -0.005 | 0.004 | 0.092 | -0.038 | -0.066 | -0.011 | 0.007 | 0.048 |
| AC(20:3) (a) | -0.017 | -0.038 | 0.005 | 0.139 | 0.903 | -0.014 | -0.028 | -0.001 | 0.035 | 0.298 | -0.016 | -0.027 | -0.005 | 0.004 | 0.097 | -0.026 | -0.053 | 0.001 | 0.061 | 0.183 |
| HexCer(d18:1/18:0) | -0.011 | -0.034 | 0.011 | 0.317 | 0.922 | -0.012 | -0.026 | 0.002 | 0.081 | 0.356 | -0.016 | -0.026 | -0.005 | 0.004 | 0.097 | -0.056 | -0.083 | -0.028 | 0.000 | 0.005 |
| PI(18:1_18:2) | 0.000 | -0.022 | 0.022 | 0.973 | 0.995 | 0.006 | -0.008 | 0.019 | 0.435 | 0.666 | -0.016 | -0.027 | -0.005 | 0.004 | 0.097 | -0.013 | -0.040 | 0.014 | 0.353 | 0.572 |
| AC(20:3) (b) | -0.010 | -0.032 | 0.012 | 0.360 | 0.922 | -0.016 | -0.029 | -0.003 | 0.018 | 0.273 | -0.015 | -0.026 | -0.004 | 0.006 | 0.119 | -0.041 | -0.066 | -0.015 | 0.002 | 0.025 |
| Cer(d18:2/20:0) | -0.009 | -0.031 | 0.013 | 0.405 | 0.922 | -0.012 | -0.026 | 0.002 | 0.092 | 0.356 | -0.015 | -0.026 | -0.004 | 0.006 | 0.119 | -0.043 | -0.071 | -0.016 | 0.002 | 0.026 |
| GM3(d18:1/24:0) | 0.007 | -0.015 | 0.029 | 0.528 | 0.922 | -0.001 | -0.015 | 0.013 | 0.867 | 0.932 | -0.015 | -0.026 | -0.004 | 0.006 | 0.119 | -0.039 | -0.067 | -0.012 | 0.005 | 0.045 |
| LPC(20:1) [sn1] | 0.007 | -0.015 | 0.029 | 0.536 | 0.923 | -0.011 | -0.025 | 0.003 | 0.137 | 0.401 | -0.015 | -0.026 | -0.004 | 0.006 | 0.119 | -0.025 | -0.052 | 0.003 | 0.079 | 0.217 |
| AC(18:0) | -0.006 | -0.028 | 0.016 | 0.616 | 0.950 | -0.020 | -0.034 | -0.007 | 0.004 | 0.259 | -0.015 | -0.026 | -0.005 | 0.005 | 0.119 | -0.054 | -0.081 | -0.027 | 0.000 | 0.005 |
| LPC(O-18:1) | -0.004 | -0.025 | 0.018 | 0.744 | 0.964 | -0.012 | -0.026 | 0.002 | 0.084 | 0.356 | -0.015 | -0.026 | -0.004 | 0.006 | 0.119 | -0.026 | -0.053 | 0.001 | 0.057 | 0.173 |
| GM1(d18:1/16:0) | 0.003 | -0.020 | 0.025 | 0.823 | 0.980 | -0.018 | -0.032 | -0.003 | 0.015 | 0.273 | -0.015 | -0.026 | -0.005 | 0.005 | 0.119 | -0.060 | -0.089 | -0.032 | 0.000 | 0.004 |
| LPC(P-18:1) | -0.007 | -0.029 | 0.015 | 0.526 | 0.922 | -0.012 | -0.025 | 0.002 | 0.099 | 0.366 | -0.015 | -0.026 | -0.004 | 0.007 | 0.120 | -0.024 | -0.051 | 0.003 | 0.081 | 0.221 |
| LPC(19:1) (b) | -0.006 | -0.028 | 0.016 | 0.597 | 0.950 | -0.010 | -0.024 | 0.003 | 0.141 | 0.406 | -0.015 | -0.026 | -0.004 | 0.006 | 0.120 | -0.040 | -0.068 | -0.013 | 0.004 | 0.041 |
| DE(18:1) | 0.001 | -0.021 | 0.023 | 0.935 | 0.995 | -0.003 | -0.016 | 0.011 | 0.717 | 0.855 | -0.015 | -0.026 | -0.004 | 0.007 | 0.120 | -0.040 | -0.068 | -0.012 | 0.006 | 0.047 |
| SM(40:3) (b) | -0.013 | -0.036 | 0.009 | 0.234 | 0.903 | -0.009 | -0.023 | 0.005 | 0.216 | 0.477 | -0.015 | -0.026 | -0.004 | 0.007 | 0.127 | -0.054 | -0.081 | -0.028 | 0.000 | 0.005 |
| PE(P-15:0/20:4) (b) | -0.012 | -0.034 | 0.010 | 0.293 | 0.903 | -0.005 | -0.019 | 0.008 | 0.458 | 0.682 | 0.014 | 0.004 | 0.025 | 0.008 | 0.127 | 0.009 | -0.018 | 0.036 | 0.524 | 0.714 |
| PE(18:1_22:6) (b) | -0.008 | -0.030 | 0.014 | 0.484 | 0.922 | -0.013 | -0.027 | 0.001 | 0.071 | 0.349 | -0.015 | -0.026 | -0.004 | 0.008 | 0.127 | -0.036 | -0.063 | -0.008 | 0.012 | 0.066 |
| SM(d18:0/22:0) | 0.009 | -0.013 | 0.031 | 0.424 | 0.922 | 0.011 | -0.003 | 0.025 | 0.122 | 0.385 | 0.015 | 0.004 | 0.025 | 0.008 | 0.127 | 0.008 | -0.019 | 0.036 | 0.566 | 0.742 |
| PE(16:0_20:3) | -0.014 | -0.036 | 0.008 | 0.199 | 0.903 | -0.015 | -0.029 | -0.001 | 0.031 | 0.291 | -0.015 | -0.025 | -0.004 | 0.008 | 0.128 | -0.026 | -0.054 | 0.001 | 0.063 | 0.184 |

| | | | | | | | | | | | | | | | | | | | | |
|---|---|---|---|---|---|---|---|---|---|---|---|---|---|---|---|---|---|---|---|---|
| Hex3Cer(d18:1/20:0) | -0.003 | -0.025 | 0.019 | 0.778 | 0.973 | -0.008 | -0.022 | 0.006 | 0.263 | 0.514 | -0.015 | -0.025 | -0.004 | 0.008 | 0.128 | -0.046 | -0.073 | -0.018 | 0.001 | 0.019 |
| PC(18:1_22:6) (a) | 0.011 | -0.011 | 0.033 | 0.337 | 0.922 | -0.013 | -0.027 | 0.001 | 0.060 | 0.327 | -0.014 | -0.025 | -0.004 | 0.009 | 0.139 | -0.045 | -0.072 | -0.018 | 0.001 | 0.020 |
| TG(52:4) [NL-16:1] | -0.011 | -0.033 | 0.011 | 0.324 | 0.922 | -0.016 | -0.030 | -0.002 | 0.021 | 0.273 | -0.014 | -0.025 | -0.003 | 0.010 | 0.139 | -0.044 | -0.071 | -0.016 | 0.002 | 0.025 |
| LPC(O-22:0) | -0.005 | -0.028 | 0.017 | 0.629 | 0.954 | -0.015 | -0.029 | -0.001 | 0.040 | 0.305 | -0.014 | -0.025 | -0.004 | 0.009 | 0.139 | -0.033 | -0.061 | -0.005 | 0.020 | 0.092 |
| SM(35:2) (b) | -0.005 | -0.027 | 0.017 | 0.660 | 0.960 | -0.006 | -0.020 | 0.008 | 0.413 | 0.646 | -0.014 | -0.025 | -0.004 | 0.009 | 0.139 | -0.051 | -0.078 | -0.025 | 0.000 | 0.006 |
| AC(16:1) | 0.000 | -0.022 | 0.022 | 0.986 | 0.996 | -0.011 | -0.025 | 0.002 | 0.108 | 0.377 | -0.014 | -0.025 | -0.003 | 0.010 | 0.146 | -0.046 | -0.074 | -0.019 | 0.001 | 0.017 |
| TG(O-50:1) [NL-18:1] | -0.010 | -0.032 | 0.011 | 0.351 | 0.922 | -0.003 | -0.016 | 0.011 | 0.703 | 0.846 | 0.014 | 0.003 | 0.025 | 0.011 | 0.150 | 0.028 | 0.000 | 0.056 | 0.047 | 0.156 |
| LPE(18:2) [sn2] | -0.024 | -0.046 | -0.002 | 0.030 | 0.903 | -0.012 | -0.026 | 0.002 | 0.087 | 0.356 | -0.014 | -0.025 | -0.003 | 0.012 | 0.156 | -0.021 | -0.049 | 0.006 | 0.134 | 0.320 |
| TG(O-54:2) [NL-17:1] | -0.017 | -0.039 | 0.006 | 0.146 | 0.903 | -0.008 | -0.022 | 0.005 | 0.218 | 0.477 | 0.014 | 0.003 | 0.025 | 0.012 | 0.156 | 0.016 | -0.011 | 0.043 | 0.242 | 0.456 |
| PC(O-18:0/18:2) | -0.011 | -0.033 | 0.011 | 0.326 | 0.922 | -0.008 | -0.022 | 0.006 | 0.246 | 0.501 | -0.014 | -0.025 | -0.003 | 0.012 | 0.156 | -0.037 | -0.064 | -0.010 | 0.007 | 0.049 |
| PC(38:5) (a) | -0.004 | -0.026 | 0.018 | 0.715 | 0.964 | -0.005 | -0.019 | 0.008 | 0.439 | 0.667 | -0.014 | -0.024 | -0.003 | 0.012 | 0.156 | -0.056 | -0.083 | -0.028 | 0.000 | 0.005 |
| Cer(d18:1/20:0) | -0.021 | -0.043 | 0.000 | 0.054 | 0.903 | -0.014 | -0.028 | 0.000 | 0.053 | 0.317 | -0.014 | -0.024 | -0.003 | 0.013 | 0.166 | -0.039 | -0.066 | -0.011 | 0.006 | 0.048 |
| LPC(20:0) [sn1] | 0.013 | -0.009 | 0.034 | 0.261 | 0.903 | -0.003 | -0.018 | 0.011 | 0.633 | 0.803 | -0.014 | -0.024 | -0.003 | 0.014 | 0.167 | -0.010 | -0.038 | 0.018 | 0.472 | 0.665 |
| PS(36:2) | -0.010 | -0.032 | 0.012 | 0.379 | 0.922 | -0.016 | -0.030 | -0.002 | 0.024 | 0.285 | -0.014 | -0.024 | -0.003 | 0.014 | 0.167 | -0.018 | -0.045 | 0.009 | 0.197 | 0.409 |
| PC(38:7) (c) | -0.004 | -0.026 | 0.018 | 0.725 | 0.964 | -0.013 | -0.027 | 0.001 | 0.073 | 0.350 | -0.014 | -0.025 | -0.003 | 0.014 | 0.167 | -0.051 | -0.079 | -0.024 | 0.000 | 0.008 |
| CE(16:2) | 0.004 | -0.018 | 0.026 | 0.739 | 0.964 | -0.003 | -0.016 | 0.011 | 0.712 | 0.853 | -0.013 | -0.024 | -0.003 | 0.014 | 0.167 | -0.032 | -0.058 | -0.005 | 0.021 | 0.093 |
| LPE(18:2) [sn1] | -0.021 | -0.042 | 0.001 | 0.066 | 0.903 | -0.011 | -0.024 | 0.003 | 0.122 | 0.385 | -0.014 | -0.024 | -0.003 | 0.015 | 0.169 | -0.026 | -0.054 | 0.001 | 0.064 | 0.184 |
| PE(P-15:0/20:4) (a) | -0.020 | -0.043 | 0.002 | 0.074 | 0.903 | -0.009 | -0.023 | 0.005 | 0.201 | 0.463 | 0.013 | 0.003 | 0.024 | 0.015 | 0.169 | 0.008 | -0.018 | 0.035 | 0.548 | 0.729 |
| Hex2Cer(d18:1/16:0) | -0.006 | -0.028 | 0.016 | 0.613 | 0.950 | -0.005 | -0.019 | 0.009 | 0.471 | 0.689 | -0.013 | -0.024 | -0.003 | 0.015 | 0.169 | -0.049 | -0.075 | -0.022 | 0.000 | 0.010 |
| PC(18:1_18:2) | 0.002 | -0.020 | 0.024 | 0.884 | 0.981 | -0.008 | -0.022 | 0.005 | 0.232 | 0.488 | -0.013 | -0.024 | -0.003 | 0.015 | 0.169 | -0.025 | -0.052 | 0.002 | 0.072 | 0.201 |
| PC(O-35:4) | -0.019 | -0.040 | 0.003 | 0.092 | 0.903 | -0.002 | -0.016 | 0.012 | 0.769 | 0.881 | 0.013 | 0.003 | 0.024 | 0.015 | 0.170 | 0.007 | -0.020 | 0.034 | 0.625 | 0.790 |
| LPC(20:1) [sn2] | 0.015 | -0.007 | 0.037 | 0.177 | 0.903 | -0.005 | -0.019 | 0.009 | 0.513 | 0.724 | -0.013 | -0.024 | -0.002 | 0.016 | 0.171 | -0.026 | -0.054 | 0.001 | 0.060 | 0.179 |
| CE(22:5) | -0.003 | -0.025 | 0.019 | 0.780 | 0.973 | 0.003 | -0.011 | 0.016 | 0.700 | 0.846 | -0.013 | -0.024 | -0.002 | 0.016 | 0.171 | -0.038 | -0.065 | -0.011 | 0.006 | 0.047 |
| PE(P-17:0/22:6) (a) | -0.007 | -0.029 | 0.016 | 0.568 | 0.933 | -0.009 | -0.023 | 0.005 | 0.216 | 0.477 | 0.013 | 0.002 | 0.024 | 0.017 | 0.174 | 0.011 | -0.017 | 0.038 | 0.446 | 0.652 |
| dhCer(d18:0/24:0) | 0.000 | -0.022 | 0.023 | 0.965 | 0.995 | 0.009 | -0.005 | 0.023 | 0.227 | 0.488 | 0.013 | 0.002 | 0.024 | 0.016 | 0.174 | 0.014 | -0.014 | 0.041 | 0.326 | 0.544 |
| Hex2Cer(d18:1/22:0) | 0.004 | -0.018 | 0.026 | 0.747 | 0.964 | 0.004 | -0.010 | 0.018 | 0.622 | 0.792 | -0.013 | -0.024 | -0.002 | 0.018 | 0.181 | -0.027 | -0.054 | 0.000 | 0.051 | 0.168 |
| SM(34:3) | -0.003 | -0.025 | 0.020 | 0.817 | 0.980 | -0.001 | -0.015 | 0.013 | 0.842 | 0.925 | -0.013 | -0.024 | -0.002 | 0.018 | 0.181 | -0.050 | -0.076 | -0.023 | 0.000 | 0.008 |
| Cer(d18:2/21:0) | -0.003 | -0.025 | 0.019 | 0.810 | 0.980 | -0.009 | -0.022 | 0.005 | 0.197 | 0.458 | -0.013 | -0.024 | -0.002 | 0.018 | 0.181 | -0.040 | -0.068 | -0.012 | 0.006 | 0.047 |
| Hex3Cer(d18:1/18:0) | -0.009 | -0.031 | 0.012 | 0.397 | 0.922 | -0.007 | -0.021 | 0.006 | 0.296 | 0.540 | -0.013 | -0.024 | -0.002 | 0.019 | 0.186 | -0.044 | -0.071 | -0.017 | 0.002 | 0.022 |
| DE(18:2) | 0.009 | -0.013 | 0.032 | 0.400 | 0.922 | 0.010 | -0.003 | 0.024 | 0.126 | 0.385 | -0.013 | -0.023 | -0.002 | 0.019 | 0.186 | -0.006 | -0.034 | 0.022 | 0.676 | 0.824 |
| Hex2Cer(d18:2/16:0) | -0.006 | -0.028 | 0.016 | 0.602 | 0.950 | 0.003 | -0.011 | 0.017 | 0.687 | 0.840 | -0.013 | -0.024 | -0.002 | 0.019 | 0.186 | -0.024 | -0.051 | 0.004 | 0.093 | 0.244 |
| PE(P-17:0/20:4) (a) | -0.006 | -0.028 | 0.017 | 0.624 | 0.954 | -0.004 | -0.018 | 0.010 | 0.584 | 0.763 | 0.013 | 0.002 | 0.023 | 0.020 | 0.186 | 0.011 | -0.016 | 0.038 | 0.428 | 0.638 |
| LPC(16:1) [sn1] | -0.018 | -0.040 | 0.004 | 0.109 | 0.903 | -0.012 | -0.026 | 0.001 | 0.081 | 0.356 | -0.013 | -0.023 | -0.002 | 0.020 | 0.186 | -0.038 | -0.064 | -0.011 | 0.006 | 0.047 |
| HexCer(d16:1/20:0) | -0.002 | -0.024 | 0.020 | 0.862 | 0.981 | -0.013 | -0.027 | 0.000 | 0.056 | 0.317 | -0.013 | -0.023 | -0.002 | 0.020 | 0.186 | -0.032 | -0.059 | -0.004 | 0.024 | 0.105 |
| Cer(d18:1/16:0) | -0.013 | -0.035 | 0.009 | 0.235 | 0.903 | -0.013 | -0.027 | 0.001 | 0.066 | 0.338 | -0.013 | -0.024 | -0.002 | 0.020 | 0.186 | -0.039 | -0.067 | -0.012 | 0.005 | 0.046 |
| CE(22:4) | -0.009 | -0.031 | 0.013 | 0.411 | 0.922 | 0.000 | -0.013 | 0.014 | 0.957 | 0.976 | -0.013 | -0.023 | -0.002 | 0.021 | 0.192 | -0.038 | -0.065 | -0.011 | 0.006 | 0.048 |
| PC(16:0_18:3) (a) | -0.014 | -0.036 | 0.008 | 0.211 | 0.903 | -0.007 | -0.022 | 0.007 | 0.317 | 0.557 | -0.013 | -0.023 | -0.002 | 0.023 | 0.193 | -0.029 | -0.056 | -0.002 | 0.036 | 0.134 |
| LPC(20:2) [sn2] | -0.013 | -0.035 | 0.009 | 0.237 | 0.903 | -0.012 | -0.026 | 0.002 | 0.085 | 0.356 | -0.013 | -0.023 | -0.002 | 0.023 | 0.193 | -0.015 | -0.042 | 0.012 | 0.276 | 0.500 |
| LPC(22:5) [sn1] (n3) & LPC(22:5) [sn2] (n6) | -0.009 | -0.031 | 0.013 | 0.403 | 0.922 | -0.014 | -0.027 | 0.000 | 0.052 | 0.317 | -0.012 | -0.023 | -0.002 | 0.022 | 0.193 | -0.040 | -0.068 | -0.013 | 0.004 | 0.043 |
| PE(P-16:0/20:4) | -0.005 | -0.027 | 0.017 | 0.657 | 0.960 | 0.009 | -0.005 | 0.024 | 0.198 | 0.460 | 0.013 | 0.002 | 0.023 | 0.022 | 0.193 | -0.002 | -0.030 | 0.026 | 0.879 | 0.948 |
| CE(18:2) | -0.003 | -0.026 | 0.019 | 0.756 | 0.969 | -0.004 | -0.018 | 0.010 | 0.537 | 0.740 | -0.013 | -0.023 | -0.002 | 0.022 | 0.193 | -0.047 | -0.075 | -0.019 | 0.001 | 0.017 |
| PE(P-19:0/20:4) (a) | -0.010 | -0.032 | 0.012 | 0.363 | 0.922 | -0.007 | -0.020 | 0.007 | 0.355 | 0.595 | 0.012 | 0.002 | 0.023 | 0.023 | 0.194 | 0.011 | -0.016 | 0.038 | 0.422 | 0.635 |
| LPC(P-18:0) | -0.008 | -0.030 | 0.014 | 0.481 | 0.922 | -0.010 | -0.024 | 0.004 | 0.159 | 0.432 | -0.012 | -0.023 | -0.002 | 0.024 | 0.195 | -0.029 | -0.056 | -0.001 | 0.040 | 0.144 |
| PC(P-15:0/20:4) (b) | -0.004 | -0.026 | 0.018 | 0.716 | 0.964 | -0.003 | -0.017 | 0.011 | 0.689 | 0.840 | 0.012 | 0.002 | 0.023 | 0.023 | 0.195 | 0.018 | -0.010 | 0.045 | 0.206 | 0.417 |
| CE(20:2) | -0.014 | -0.036 | 0.008 | 0.205 | 0.903 | -0.002 | -0.016 | 0.012 | 0.780 | 0.884 | -0.012 | -0.023 | -0.002 | 0.024 | 0.197 | -0.032 | -0.059 | -0.005 | 0.019 | 0.089 |
| SM(41:0) | 0.016 | -0.005 | 0.038 | 0.141 | 0.903 | 0.000 | -0.014 | 0.014 | 0.985 | 0.989 | 0.012 | 0.002 | 0.023 | 0.025 | 0.198 | -0.002 | -0.029 | 0.025 | 0.883 | 0.948 |
| HexCer(d18:2/22:0) | 0.012 | -0.010 | 0.034 | 0.273 | 0.903 | 0.006 | -0.008 | 0.019 | 0.437 | 0.667 | -0.012 | -0.023 | -0.002 | 0.025 | 0.201 | -0.013 | -0.040 | 0.015 | 0.370 | 0.590 |
| SM(d18:2/20:0) | -0.003 | -0.025 | 0.019 | 0.782 | 0.973 | -0.001 | -0.015 | 0.013 | 0.854 | 0.929 | -0.012 | -0.023 | -0.001 | 0.026 | 0.201 | -0.055 | -0.082 | -0.028 | 0.000 | 0.005 |
| AC(14:2) | 0.003 | -0.019 | 0.025 | 0.791 | 0.973 | 0.000 | -0.014 | 0.013 | 0.946 | 0.972 | -0.012 | -0.023 | -0.001 | 0.026 | 0.201 | -0.035 | -0.063 | -0.007 | 0.015 | 0.076 |
| LPC(19:0) [sn1] (b) | -0.001 | -0.023 | 0.021 | 0.901 | 0.985 | -0.013 | -0.027 | 0.000 | 0.057 | 0.317 | -0.012 | -0.023 | -0.001 | 0.026 | 0.201 | -0.019 | -0.047 | 0.008 | 0.169 | 0.373 |

| | | | | | | | | | | | | | | | | | | | | |
|---|---|---|---|---|---|---|---|---|---|---|---|---|---|---|---|---|---|---|---|---|
| PA(36:2) | -0.006 | -0.028 | 0.017 | 0.625 | 0.954 | -0.007 | -0.021 | 0.007 | 0.302 | 0.546 | -0.012 | -0.023 | -0.001 | 0.027 | 0.206 | 0.001 | -0.027 | 0.029 | 0.933 | 0.964 |
| TG(54:4) [NL-18:2] | 0.005 | -0.016 | 0.027 | 0.623 | 0.954 | -0.008 | -0.022 | 0.006 | 0.280 | 0.524 | -0.012 | -0.023 | -0.001 | 0.028 | 0.212 | -0.020 | -0.047 | 0.008 | 0.159 | 0.359 |
| PE(P-19:0/20:4) (b) | -0.008 | -0.030 | 0.015 | 0.493 | 0.922 | -0.001 | -0.015 | 0.013 | 0.919 | 0.953 | 0.012 | 0.001 | 0.023 | 0.030 | 0.223 | 0.006 | -0.021 | 0.034 | 0.664 | 0.815 |
| PE(P-17:0/20:4) (b) | -0.009 | -0.031 | 0.013 | 0.413 | 0.922 | 0.002 | -0.012 | 0.016 | 0.751 | 0.877 | 0.012 | 0.001 | 0.023 | 0.030 | 0.225 | -0.007 | -0.034 | 0.021 | 0.632 | 0.794 |
| PE(P-18:1/22:4) | -0.012 | -0.034 | 0.010 | 0.271 | 0.903 | -0.002 | -0.016 | 0.012 | 0.781 | 0.884 | -0.012 | -0.023 | -0.001 | 0.031 | 0.226 | -0.037 | -0.063 | -0.010 | 0.006 | 0.048 |
| PE(P-16:0/18:1) | -0.006 | -0.028 | 0.016 | 0.594 | 0.950 | -0.004 | -0.018 | 0.011 | 0.624 | 0.793 | 0.012 | 0.001 | 0.023 | 0.031 | 0.226 | -0.006 | -0.034 | 0.021 | 0.660 | 0.814 |
| LPC(20:0) [sn2] | 0.016 | -0.006 | 0.038 | 0.162 | 0.903 | -0.001 | -0.015 | 0.013 | 0.890 | 0.939 | -0.012 | -0.022 | -0.001 | 0.034 | 0.242 | -0.009 | -0.036 | 0.019 | 0.541 | 0.725 |
| TG(O-50:1) [NL-15:0] | -0.011 | -0.033 | 0.010 | 0.309 | 0.907 | -0.003 | -0.016 | 0.011 | 0.703 | 0.846 | 0.012 | 0.001 | 0.022 | 0.034 | 0.242 | 0.013 | -0.015 | 0.040 | 0.359 | 0.577 |
| SM(38:3) (a) | -0.010 | -0.032 | 0.012 | 0.369 | 0.922 | -0.004 | -0.018 | 0.010 | 0.607 | 0.777 | -0.012 | -0.022 | -0.001 | 0.035 | 0.244 | -0.051 | -0.077 | -0.024 | 0.000 | 0.008 |
| SM(38:3) (b) | -0.012 | -0.034 | 0.010 | 0.299 | 0.903 | -0.012 | -0.026 | 0.002 | 0.090 | 0.356 | -0.012 | -0.022 | -0.001 | 0.037 | 0.253 | -0.049 | -0.075 | -0.023 | 0.000 | 0.008 |
| Cer(d18:1/21:0) | -0.014 | -0.036 | 0.008 | 0.225 | 0.903 | -0.008 | -0.022 | 0.006 | 0.258 | 0.509 | -0.012 | -0.022 | -0.001 | 0.036 | 0.253 | -0.046 | -0.073 | -0.018 | 0.001 | 0.018 |
| LPC(17:0) [sn1] | -0.004 | -0.026 | 0.018 | 0.709 | 0.964 | -0.013 | -0.027 | 0.001 | 0.064 | 0.338 | -0.011 | -0.022 | -0.001 | 0.037 | 0.253 | -0.033 | -0.061 | -0.006 | 0.017 | 0.085 |
| Cer(d18:2/24:1) | -0.007 | -0.029 | 0.015 | 0.517 | 0.922 | -0.017 | -0.031 | -0.004 | 0.014 | 0.273 | -0.012 | -0.022 | -0.001 | 0.037 | 0.253 | -0.031 | -0.059 | -0.004 | 0.027 | 0.110 |
| AC(12:1) | -0.005 | -0.027 | 0.017 | 0.675 | 0.964 | -0.005 | -0.018 | 0.009 | 0.501 | 0.719 | -0.011 | -0.022 | -0.001 | 0.038 | 0.253 | -0.048 | -0.076 | -0.020 | 0.001 | 0.014 |
| PE(38:5) (a) | -0.013 | -0.035 | 0.009 | 0.259 | 0.903 | -0.014 | -0.028 | 0.000 | 0.054 | 0.317 | -0.011 | -0.022 | -0.001 | 0.038 | 0.255 | -0.034 | -0.062 | -0.006 | 0.018 | 0.087 |
| TG(54:5) [NL-18:3] | -0.005 | -0.027 | 0.017 | 0.643 | 0.960 | -0.012 | -0.026 | 0.001 | 0.081 | 0.356 | -0.011 | -0.022 | -0.001 | 0.040 | 0.266 | -0.012 | -0.039 | 0.015 | 0.392 | 0.609 |
| GM3(d18:1/18:0) | -0.018 | -0.040 | 0.004 | 0.114 | 0.903 | -0.009 | -0.022 | 0.005 | 0.225 | 0.486 | -0.011 | -0.022 | 0.000 | 0.042 | 0.267 | -0.056 | -0.083 | -0.028 | 0.000 | 0.005 |
| LPE(16:0) [sn2] | -0.017 | -0.039 | 0.005 | 0.125 | 0.903 | -0.017 | -0.030 | -0.003 | 0.014 | 0.273 | -0.011 | -0.022 | 0.000 | 0.041 | 0.267 | -0.031 | -0.058 | -0.004 | 0.025 | 0.106 |
| PI(38:5) (a) | -0.016 | -0.038 | 0.006 | 0.155 | 0.903 | -0.006 | -0.019 | 0.008 | 0.431 | 0.663 | -0.011 | -0.022 | 0.000 | 0.043 | 0.267 | -0.027 | -0.055 | 0.000 | 0.053 | 0.170 |
| TG(O-50:1) [NL-16:0] | -0.013 | -0.035 | 0.008 | 0.228 | 0.903 | -0.006 | -0.019 | 0.008 | 0.419 | 0.650 | 0.011 | 0.000 | 0.022 | 0.042 | 0.267 | 0.018 | -0.010 | 0.046 | 0.211 | 0.420 |
| PC(O-40:5) | -0.008 | -0.031 | 0.014 | 0.456 | 0.922 | -0.009 | -0.023 | 0.005 | 0.218 | 0.477 | -0.011 | -0.022 | 0.000 | 0.042 | 0.267 | -0.052 | -0.079 | -0.026 | 0.000 | 0.006 |
| LPE(P-20:0) | 0.005 | -0.017 | 0.027 | 0.629 | 0.954 | -0.013 | -0.027 | 0.001 | 0.062 | 0.333 | -0.011 | -0.022 | 0.000 | 0.041 | 0.267 | -0.023 | -0.051 | 0.005 | 0.107 | 0.272 |
| PC(18:1_18:1) | -0.004 | -0.026 | 0.018 | 0.706 | 0.964 | -0.018 | -0.032 | -0.004 | 0.013 | 0.273 | -0.011 | -0.022 | 0.000 | 0.042 | 0.267 | -0.032 | -0.059 | -0.005 | 0.020 | 0.092 |
| TG(O-52:1) [NL-18:1] | -0.013 | -0.035 | 0.008 | 0.225 | 0.903 | -0.008 | -0.022 | 0.005 | 0.239 | 0.495 | 0.011 | 0.000 | 0.022 | 0.043 | 0.268 | 0.018 | -0.009 | 0.046 | 0.189 | 0.401 |
| PC(O-18:0/18:1) | -0.012 | -0.034 | 0.010 | 0.290 | 0.903 | -0.012 | -0.026 | 0.002 | 0.081 | 0.356 | -0.011 | -0.022 | 0.000 | 0.048 | 0.270 | -0.051 | -0.078 | -0.024 | 0.000 | 0.008 |
| LPC(O-16:0) | -0.012 | -0.033 | 0.010 | 0.280 | 0.903 | -0.010 | -0.024 | 0.004 | 0.153 | 0.424 | -0.011 | -0.022 | 0.000 | 0.047 | 0.270 | -0.021 | -0.049 | 0.006 | 0.124 | 0.301 |
| HexCer(d18:1/22:0) | 0.015 | -0.007 | 0.037 | 0.184 | 0.903 | 0.001 | -0.013 | 0.015 | 0.862 | 0.930 | -0.011 | -0.022 | 0.000 | 0.049 | 0.270 | -0.020 | -0.048 | 0.008 | 0.155 | 0.352 |
| LPC(16:0) [sn1] | -0.014 | -0.036 | 0.007 | 0.198 | 0.903 | -0.017 | -0.031 | -0.003 | 0.014 | 0.273 | -0.011 | -0.022 | 0.000 | 0.051 | 0.270 | -0.019 | -0.046 | 0.008 | 0.168 | 0.373 |
| PC(O-34:4) | -0.015 | -0.037 | 0.007 | 0.183 | 0.903 | -0.004 | -0.018 | 0.010 | 0.616 | 0.787 | 0.011 | 0.000 | 0.021 | 0.048 | 0.270 | 0.010 | -0.017 | 0.037 | 0.466 | 0.664 |
| LPC(18:3) [sn2] (a) | -0.012 | -0.034 | 0.010 | 0.295 | 0.903 | -0.009 | -0.023 | 0.005 | 0.220 | 0.479 | -0.011 | -0.022 | 0.000 | 0.047 | 0.270 | -0.010 | -0.037 | 0.018 | 0.481 | 0.675 |
| LPC(16:0) [sn2] | -0.015 | -0.037 | 0.007 | 0.175 | 0.903 | -0.012 | -0.026 | 0.002 | 0.089 | 0.356 | -0.011 | -0.022 | 0.000 | 0.047 | 0.270 | -0.005 | -0.032 | 0.022 | 0.698 | 0.842 |
| HexCer(d18:1/24:1) | 0.008 | -0.014 | 0.030 | 0.463 | 0.922 | -0.015 | -0.028 | -0.001 | 0.034 | 0.298 | -0.011 | -0.022 | 0.000 | 0.047 | 0.270 | -0.029 | -0.057 | -0.001 | 0.040 | 0.142 |
| TG(O-52:2) [NL-17:1] | -0.008 | -0.029 | 0.014 | 0.495 | 0.922 | -0.005 | -0.019 | 0.008 | 0.453 | 0.678 | 0.011 | 0.000 | 0.021 | 0.049 | 0.270 | 0.021 | -0.007 | 0.048 | 0.141 | 0.330 |
| PE(P-16:0/20:5) | 0.011 | -0.011 | 0.033 | 0.336 | 0.922 | -0.006 | -0.020 | 0.008 | 0.416 | 0.649 | 0.011 | 0.000 | 0.022 | 0.047 | 0.270 | 0.001 | -0.027 | 0.028 | 0.961 | 0.971 |
| LPC(22:5) (n3) [sn1] [104_sn1] | -0.006 | -0.028 | 0.016 | 0.617 | 0.950 | -0.015 | -0.029 | -0.001 | 0.032 | 0.298 | -0.011 | -0.022 | 0.000 | 0.048 | 0.270 | -0.039 | -0.067 | -0.011 | 0.006 | 0.048 |
| LPC(22:5) [sn2] (n3) | -0.005 | -0.027 | 0.017 | 0.657 | 0.960 | -0.014 | -0.028 | -0.001 | 0.042 | 0.306 | -0.011 | -0.021 | 0.000 | 0.051 | 0.270 | -0.037 | -0.065 | -0.010 | 0.008 | 0.050 |
| TG(54:6) [NL-22:6] | -0.005 | -0.027 | 0.017 | 0.652 | 0.960 | -0.011 | -0.025 | 0.003 | 0.119 | 0.385 | 0.011 | 0.000 | 0.022 | 0.051 | 0.270 | 0.013 | -0.015 | 0.041 | 0.366 | 0.586 |
| DG(18:1_22:6) | 0.004 | -0.018 | 0.026 | 0.735 | 0.964 | -0.012 | -0.026 | 0.002 | 0.086 | 0.356 | 0.011 | 0.000 | 0.022 | 0.049 | 0.270 | 0.009 | -0.019 | 0.038 | 0.508 | 0.702 |
| TG(56:7) [NL-22:6] | 0.005 | -0.017 | 0.027 | 0.668 | 0.964 | -0.014 | -0.028 | 0.000 | 0.051 | 0.317 | 0.011 | 0.000 | 0.022 | 0.047 | 0.270 | 0.007 | -0.021 | 0.035 | 0.640 | 0.801 |
| PE(P-18:1/20:4) (a) | -0.003 | -0.025 | 0.019 | 0.794 | 0.974 | 0.008 | -0.006 | 0.022 | 0.272 | 0.521 | 0.011 | 0.000 | 0.022 | 0.046 | 0.270 | -0.003 | -0.031 | 0.025 | 0.826 | 0.923 |
| AC(24:1) | 0.002 | -0.020 | 0.024 | 0.848 | 0.981 | -0.010 | -0.024 | 0.004 | 0.172 | 0.438 | -0.011 | -0.022 | 0.000 | 0.049 | 0.270 | -0.020 | -0.047 | 0.006 | 0.135 | 0.320 |
| LPC(18:2) [+OH] | 0.001 | -0.021 | 0.024 | 0.898 | 0.983 | -0.010 | -0.024 | 0.004 | 0.163 | 0.432 | -0.011 | -0.022 | 0.000 | 0.051 | 0.270 | -0.002 | -0.030 | 0.026 | 0.900 | 0.952 |
| Hex2Cer(d18:1/24:0) | 0.001 | -0.021 | 0.023 | 0.913 | 0.987 | 0.000 | -0.013 | 0.014 | 0.956 | 0.976 | -0.011 | -0.022 | 0.000 | 0.044 | 0.270 | -0.021 | -0.048 | 0.007 | 0.142 | 0.330 |
| LPE(P-18:1) | -0.001 | -0.022 | 0.021 | 0.962 | 0.995 | -0.001 | -0.016 | 0.013 | 0.856 | 0.929 | -0.011 | -0.021 | 0.000 | 0.051 | 0.270 | -0.031 | -0.058 | -0.004 | 0.026 | 0.107 |
| PE(O-36:5) | 0.001 | -0.022 | 0.023 | 0.964 | 0.995 | -0.009 | -0.023 | 0.005 | 0.212 | 0.475 | 0.011 | 0.000 | 0.021 | 0.051 | 0.270 | 0.004 | -0.024 | 0.031 | 0.793 | 0.904 |
| TG(52:4) [NL-18:2] | -0.012 | -0.034 | 0.010 | 0.276 | 0.903 | -0.007 | -0.021 | 0.007 | 0.301 | 0.546 | -0.011 | -0.021 | 0.000 | 0.053 | 0.274 | -0.023 | -0.050 | 0.005 | 0.108 | 0.274 |
| LPC(18:0) [sn1] | -0.005 | -0.027 | 0.017 | 0.649 | 0.960 | -0.009 | -0.023 | 0.004 | 0.180 | 0.450 | -0.011 | -0.021 | 0.000 | 0.052 | 0.274 | -0.027 | -0.054 | 0.001 | 0.056 | 0.173 |
| PC(16:1_18:2) | -0.013 | -0.035 | 0.009 | 0.253 | 0.903 | -0.008 | -0.022 | 0.006 | 0.267 | 0.520 | -0.011 | -0.021 | 0.000 | 0.053 | 0.275 | -0.032 | -0.059 | -0.006 | 0.018 | 0.086 |
| dhCer(d18:0/20:0) | -0.007 | -0.029 | 0.015 | 0.543 | 0.929 | 0.001 | -0.013 | 0.014 | 0.938 | 0.969 | 0.011 | 0.000 | 0.021 | 0.054 | 0.278 | -0.002 | -0.030 | 0.025 | 0.881 | 0.948 |
| AC(14:1) | 0.005 | -0.017 | 0.027 | 0.629 | 0.954 | -0.006 | -0.020 | 0.007 | 0.358 | 0.598 | -0.011 | -0.021 | 0.000 | 0.055 | 0.280 | -0.046 | -0.073 | -0.018 | 0.001 | 0.020 |

| | | | | | | | | | | | | | | | | | | | | |
|---|---|---|---|---|---|---|---|---|---|---|---|---|---|---|---|---|---|---|---|---|
| HexCer(d18:2/18:0) | 0.002 | -0.020 | 0.024 | 0.861 | 0.981 | -0.017 | -0.031 | -0.003 | 0.017 | 0.273 | -0.011 | -0.021 | 0.000 | 0.056 | 0.282 | -0.034 | -0.062 | -0.006 | 0.017 | 0.084 |
| PE(P-16:0/22:6) | 0.002 | -0.021 | 0.024 | 0.882 | 0.981 | 0.000 | -0.014 | 0.015 | 0.962 | 0.979 | 0.011 | 0.000 | 0.021 | 0.056 | 0.283 | 0.002 | -0.026 | 0.030 | 0.878 | 0.948 |
| Cer(d18:2/22:0) | -0.004 | -0.026 | 0.018 | 0.730 | 0.964 | -0.003 | -0.017 | 0.012 | 0.698 | 0.845 | -0.011 | -0.021 | 0.000 | 0.057 | 0.284 | -0.016 | -0.044 | 0.012 | 0.259 | 0.477 |
| dhCer(d18:0/22:0) | 0.001 | -0.021 | 0.023 | 0.914 | 0.987 | 0.009 | -0.005 | 0.023 | 0.213 | 0.475 | 0.010 | 0.000 | 0.021 | 0.058 | 0.287 | 0.013 | -0.014 | 0.040 | 0.358 | 0.577 |
| PE(P-18:1/20:5) (a) | 0.013 | -0.009 | 0.035 | 0.250 | 0.903 | -0.008 | -0.022 | 0.006 | 0.269 | 0.520 | 0.010 | 0.000 | 0.021 | 0.060 | 0.292 | -0.009 | -0.036 | 0.019 | 0.532 | 0.719 |
| PE(P-18:1/18:2) (b) | -0.008 | -0.030 | 0.014 | 0.474 | 0.922 | 0.001 | -0.013 | 0.015 | 0.906 | 0.944 | -0.010 | -0.021 | 0.000 | 0.060 | 0.292 | -0.034 | -0.060 | -0.007 | 0.014 | 0.072 |
| LPC(18:0) [sn2] | -0.003 | -0.025 | 0.019 | 0.781 | 0.973 | -0.009 | -0.022 | 0.005 | 0.226 | 0.487 | -0.010 | -0.021 | 0.000 | 0.061 | 0.292 | -0.025 | -0.052 | 0.003 | 0.077 | 0.215 |
| PC(P-16:0/22:6) | -0.001 | -0.023 | 0.022 | 0.961 | 0.995 | -0.007 | -0.021 | 0.007 | 0.340 | 0.578 | 0.010 | 0.000 | 0.021 | 0.060 | 0.292 | -0.001 | -0.029 | 0.026 | 0.924 | 0.964 |
| Cer1P(d18:1/16:0) | -0.004 | -0.025 | 0.018 | 0.747 | 0.964 | -0.001 | -0.015 | 0.013 | 0.898 | 0.942 | -0.010 | -0.021 | 0.000 | 0.061 | 0.293 | -0.025 | -0.053 | 0.003 | 0.078 | 0.216 |
| SM(44:3) | -0.008 | -0.030 | 0.015 | 0.500 | 0.922 | -0.008 | -0.022 | 0.006 | 0.256 | 0.507 | -0.010 | -0.021 | 0.000 | 0.062 | 0.294 | -0.039 | -0.066 | -0.012 | 0.004 | 0.042 |
| TG(52:3) [NL-16:1] | 0.003 | -0.018 | 0.025 | 0.757 | 0.969 | -0.019 | -0.033 | -0.006 | 0.006 | 0.259 | -0.010 | -0.021 | 0.001 | 0.062 | 0.294 | -0.053 | -0.080 | -0.025 | 0.000 | 0.008 |
| AC(18:1)-OH | 0.002 | -0.020 | 0.024 | 0.878 | 0.981 | -0.008 | -0.022 | 0.005 | 0.240 | 0.495 | -0.010 | -0.021 | 0.001 | 0.063 | 0.294 | -0.041 | -0.068 | -0.013 | 0.004 | 0.038 |
| PC(40:8) | -0.002 | -0.024 | 0.021 | 0.874 | 0.981 | -0.009 | -0.023 | 0.005 | 0.232 | 0.488 | -0.010 | -0.021 | 0.001 | 0.064 | 0.300 | -0.026 | -0.054 | 0.001 | 0.060 | 0.181 |
| LPE(18:1) [sn1] | -0.013 | -0.035 | 0.009 | 0.263 | 0.903 | -0.015 | -0.028 | -0.001 | 0.038 | 0.301 | -0.010 | -0.021 | 0.001 | 0.067 | 0.309 | -0.019 | -0.047 | 0.008 | 0.174 | 0.378 |
| Hex3Cer(d18:1/22:0) | 0.001 | -0.021 | 0.023 | 0.948 | 0.995 | -0.004 | -0.018 | 0.010 | 0.607 | 0.777 | -0.010 | -0.021 | 0.001 | 0.067 | 0.309 | -0.045 | -0.072 | -0.018 | 0.001 | 0.018 |
| Cer(d18:1/18:0) | -0.026 | -0.048 | -0.005 | 0.017 | 0.903 | -0.013 | -0.027 | 0.000 | 0.056 | 0.317 | -0.010 | -0.021 | 0.001 | 0.068 | 0.310 | -0.049 | -0.076 | -0.022 | 0.001 | 0.011 |
| PC(O-18:1/18:1) | 0.000 | -0.022 | 0.022 | 1.000 | 1.000 | -0.011 | -0.025 | 0.003 | 0.129 | 0.390 | -0.010 | -0.021 | 0.001 | 0.068 | 0.310 | -0.036 | -0.063 | -0.009 | 0.009 | 0.056 |
| PE(P-18:0/20:4) | -0.006 | -0.028 | 0.016 | 0.568 | 0.933 | 0.006 | -0.008 | 0.021 | 0.400 | 0.638 | 0.010 | -0.001 | 0.021 | 0.069 | 0.312 | -0.002 | -0.030 | 0.026 | 0.887 | 0.948 |
| HexCer(d16:1/22:0) | 0.009 | -0.013 | 0.032 | 0.404 | 0.922 | -0.009 | -0.023 | 0.004 | 0.186 | 0.453 | -0.010 | -0.021 | 0.001 | 0.069 | 0.312 | -0.011 | -0.039 | 0.016 | 0.417 | 0.631 |
| AC(12:0) | -0.002 | -0.024 | 0.021 | 0.891 | 0.981 | -0.007 | -0.021 | 0.007 | 0.304 | 0.546 | -0.010 | -0.021 | 0.001 | 0.070 | 0.312 | -0.050 | -0.079 | -0.022 | 0.000 | 0.010 |
| LPC(15-MHDA) [sn1] & LPC(17:0) [sn2] | -0.003 | -0.025 | 0.019 | 0.791 | 0.973 | -0.015 | -0.029 | -0.001 | 0.037 | 0.301 | -0.010 | -0.021 | 0.001 | 0.071 | 0.317 | -0.036 | -0.063 | -0.008 | 0.011 | 0.064 |
| LPE(18:0) [sn1] | -0.018 | -0.040 | 0.004 | 0.111 | 0.903 | -0.012 | -0.026 | 0.001 | 0.073 | 0.350 | -0.010 | -0.021 | 0.001 | 0.072 | 0.319 | -0.021 | -0.049 | 0.006 | 0.127 | 0.309 |
| PC(P-16:0/20:5) | 0.008 | -0.014 | 0.030 | 0.481 | 0.922 | -0.009 | -0.023 | 0.005 | 0.219 | 0.477 | 0.010 | -0.001 | 0.021 | 0.073 | 0.319 | 0.003 | -0.025 | 0.031 | 0.837 | 0.928 |
| S1P(d18:0) | -0.005 | -0.027 | 0.017 | 0.676 | 0.964 | -0.010 | -0.024 | 0.004 | 0.145 | 0.406 | -0.010 | -0.020 | 0.001 | 0.073 | 0.319 | -0.022 | -0.050 | 0.006 | 0.119 | 0.293 |
| PE(P-17:0/22:6) (b) | -0.002 | -0.024 | 0.021 | 0.879 | 0.981 | -0.002 | -0.017 | 0.012 | 0.749 | 0.877 | 0.010 | -0.001 | 0.021 | 0.073 | 0.319 | -0.003 | -0.032 | 0.025 | 0.826 | 0.923 |
| PE(P-16:0/20:3) (a) | -0.010 | -0.033 | 0.012 | 0.356 | 0.922 | -0.005 | -0.019 | 0.009 | 0.465 | 0.686 | 0.010 | -0.001 | 0.021 | 0.074 | 0.320 | -0.014 | -0.042 | 0.013 | 0.303 | 0.526 |
| AC(16:0) | -0.014 | -0.036 | 0.008 | 0.211 | 0.903 | -0.015 | -0.029 | -0.002 | 0.030 | 0.289 | -0.010 | -0.020 | 0.001 | 0.075 | 0.323 | -0.048 | -0.075 | -0.022 | 0.000 | 0.010 |
| PC(38:4) (b) | -0.016 | -0.037 | 0.006 | 0.157 | 0.903 | -0.003 | -0.017 | 0.011 | 0.640 | 0.807 | -0.010 | -0.021 | 0.001 | 0.076 | 0.323 | -0.039 | -0.066 | -0.013 | 0.004 | 0.042 |
| LPC(22:4) [sn1] | -0.011 | -0.033 | 0.010 | 0.300 | 0.903 | -0.004 | -0.018 | 0.010 | 0.566 | 0.748 | -0.010 | -0.020 | 0.001 | 0.076 | 0.323 | -0.018 | -0.045 | 0.010 | 0.208 | 0.419 |
| AC(14:0) | -0.003 | -0.025 | 0.019 | 0.791 | 0.973 | -0.013 | -0.027 | 0.000 | 0.057 | 0.317 | -0.010 | -0.020 | 0.001 | 0.076 | 0.323 | -0.046 | -0.074 | -0.019 | 0.001 | 0.017 |
| LPE(18:1) [sn2] | -0.013 | -0.035 | 0.009 | 0.263 | 0.903 | -0.013 | -0.027 | 0.000 | 0.055 | 0.317 | -0.010 | -0.021 | 0.001 | 0.077 | 0.324 | -0.020 | -0.048 | 0.007 | 0.149 | 0.340 |
| Cer(d18:2/16:0) | -0.003 | -0.025 | 0.019 | 0.770 | 0.973 | -0.006 | -0.021 | 0.008 | 0.368 | 0.603 | -0.010 | -0.021 | 0.001 | 0.078 | 0.325 | -0.033 | -0.061 | -0.006 | 0.018 | 0.086 |
| TG(O-50:3) [NL-18:2] | -0.030 | -0.052 | -0.009 | 0.006 | 0.903 | 0.010 | -0.003 | 0.024 | 0.138 | 0.403 | 0.010 | -0.001 | 0.020 | 0.078 | 0.326 | 0.020 | -0.006 | 0.047 | 0.133 | 0.318 |
| Cer(d18:1/24:1) | -0.020 | -0.042 | 0.002 | 0.070 | 0.903 | -0.019 | -0.033 | -0.006 | 0.006 | 0.259 | -0.010 | -0.020 | 0.001 | 0.079 | 0.328 | -0.040 | -0.067 | -0.012 | 0.005 | 0.044 |
| LPC(22:0) [sn2] | 0.012 | -0.009 | 0.034 | 0.266 | 0.903 | 0.005 | -0.009 | 0.018 | 0.521 | 0.727 | -0.010 | -0.020 | 0.001 | 0.080 | 0.328 | 0.004 | -0.023 | 0.031 | 0.768 | 0.892 |
| Cer(d18:2/18:0) | -0.015 | -0.037 | 0.007 | 0.181 | 0.903 | -0.010 | -0.023 | 0.004 | 0.162 | 0.432 | -0.010 | -0.020 | 0.001 | 0.081 | 0.330 | -0.044 | -0.072 | -0.017 | 0.002 | 0.023 |
| dxCA | -0.013 | -0.036 | 0.009 | 0.250 | 0.903 | 0.001 | -0.013 | 0.014 | 0.941 | 0.971 | -0.010 | -0.021 | 0.001 | 0.082 | 0.333 | -0.015 | -0.042 | 0.012 | 0.279 | 0.503 |
| PE(18:1_18:2) | -0.005 | -0.027 | 0.017 | 0.649 | 0.960 | -0.012 | -0.026 | 0.001 | 0.080 | 0.356 | -0.010 | -0.020 | 0.001 | 0.083 | 0.337 | -0.005 | -0.033 | 0.023 | 0.728 | 0.863 |
| PE(P-18:0/22:6) | 0.003 | -0.019 | 0.025 | 0.798 | 0.975 | -0.004 | -0.019 | 0.010 | 0.544 | 0.745 | 0.010 | -0.001 | 0.020 | 0.086 | 0.345 | -0.003 | -0.031 | 0.025 | 0.815 | 0.918 |
| SM(d18:2/18:0) | -0.014 | -0.036 | 0.008 | 0.208 | 0.903 | -0.005 | -0.019 | 0.008 | 0.448 | 0.673 | -0.009 | -0.020 | 0.001 | 0.087 | 0.346 | -0.050 | -0.077 | -0.023 | 0.000 | 0.008 |
| LPE(20:4) [sn2] | -0.016 | -0.038 | 0.006 | 0.164 | 0.903 | -0.010 | -0.024 | 0.004 | 0.159 | 0.432 | -0.009 | -0.020 | 0.001 | 0.090 | 0.346 | -0.043 | -0.070 | -0.015 | 0.003 | 0.030 |
| PC(16:0_20:3) (a) | -0.012 | -0.034 | 0.010 | 0.276 | 0.903 | -0.011 | -0.025 | 0.003 | 0.119 | 0.385 | -0.009 | -0.020 | 0.001 | 0.089 | 0.346 | -0.037 | -0.064 | -0.011 | 0.006 | 0.048 |
| LPC(22:0) [sn1] | 0.014 | -0.008 | 0.036 | 0.216 | 0.903 | 0.002 | -0.012 | 0.016 | 0.769 | 0.881 | -0.009 | -0.020 | 0.001 | 0.090 | 0.346 | 0.008 | -0.020 | 0.035 | 0.590 | 0.756 |
| HexCer(d16:1/18:0) | -0.009 | -0.032 | 0.013 | 0.411 | 0.922 | -0.010 | -0.023 | 0.004 | 0.162 | 0.432 | -0.009 | -0.020 | 0.001 | 0.089 | 0.346 | -0.034 | -0.062 | -0.007 | 0.014 | 0.074 |
| PE(P-18:0/20:5) | 0.010 | -0.012 | 0.033 | 0.363 | 0.922 | -0.008 | -0.023 | 0.006 | 0.246 | 0.501 | 0.009 | -0.001 | 0.020 | 0.089 | 0.346 | -0.005 | -0.032 | 0.023 | 0.748 | 0.882 |
| PC(O-36:0) | -0.006 | -0.028 | 0.016 | 0.590 | 0.948 | -0.008 | -0.022 | 0.006 | 0.274 | 0.521 | -0.009 | -0.020 | 0.001 | 0.089 | 0.346 | -0.044 | -0.071 | -0.017 | 0.001 | 0.021 |
| TG(O-52:2) [NL-18:1] | -0.005 | -0.026 | 0.017 | 0.674 | 0.964 | -0.004 | -0.018 | 0.010 | 0.554 | 0.748 | 0.009 | -0.001 | 0.020 | 0.089 | 0.346 | 0.018 | -0.010 | 0.046 | 0.203 | 0.416 |
| TG(54:6) [NL-18:3] | -0.001 | -0.023 | 0.021 | 0.897 | 0.983 | -0.005 | -0.018 | 0.009 | 0.519 | 0.727 | -0.009 | -0.020 | 0.001 | 0.087 | 0.346 | -0.002 | -0.029 | 0.026 | 0.905 | 0.954 |
| LPC(19:1) (a) | -0.005 | -0.027 | 0.018 | 0.687 | 0.964 | -0.010 | -0.024 | 0.004 | 0.148 | 0.410 | -0.009 | -0.020 | 0.001 | 0.091 | 0.347 | -0.028 | -0.056 | -0.001 | 0.044 | 0.150 |
| DG(16:0_22:6) | -0.003 | -0.026 | 0.019 | 0.762 | 0.969 | -0.011 | -0.025 | 0.003 | 0.115 | 0.382 | 0.009 | -0.002 | 0.020 | 0.092 | 0.350 | 0.009 | -0.018 | 0.037 | 0.513 | 0.706 |

| | | | | | | | | | | | | | | | | | | | | |
|---|---|---|---|---|---|---|---|---|---|---|---|---|---|---|---|---|---|---|---|---|
| DG(18:2_22:6) | 0.003 | -0.019 | 0.025 | 0.782 | 0.973 | -0.008 | -0.022 | 0.006 | 0.280 | 0.524 | 0.009 | -0.002 | 0.020 | 0.093 | 0.351 | 0.008 | -0.020 | 0.036 | 0.568 | 0.743 |
| LPC(19:0) [sn1] (a) & LPC(19:0) [sn2] (b) | 0.003 | -0.019 | 0.025 | 0.760 | 0.969 | -0.010 | -0.024 | 0.004 | 0.165 | 0.432 | -0.009 | -0.020 | 0.002 | 0.094 | 0.354 | -0.019 | -0.046 | 0.008 | 0.172 | 0.376 |
| LPC(20:4) [+OH] | 0.006 | -0.016 | 0.029 | 0.571 | 0.933 | -0.002 | -0.015 | 0.012 | 0.819 | 0.907 | -0.009 | -0.020 | 0.002 | 0.095 | 0.355 | -0.021 | -0.048 | 0.006 | 0.134 | 0.320 |
| CE(16:0) | -0.002 | -0.024 | 0.020 | 0.847 | 0.981 | -0.004 | -0.018 | 0.010 | 0.548 | 0.747 | -0.009 | -0.020 | 0.002 | 0.095 | 0.355 | -0.038 | -0.065 | -0.010 | 0.007 | 0.049 |
| PE(P-20:1/20:4) | 0.004 | -0.019 | 0.026 | 0.756 | 0.969 | 0.001 | -0.012 | 0.015 | 0.871 | 0.932 | 0.009 | -0.002 | 0.020 | 0.096 | 0.356 | 0.016 | -0.011 | 0.043 | 0.252 | 0.465 |
| HexCer(d18:2/24:0) | 0.013 | -0.009 | 0.035 | 0.243 | 0.903 | -0.003 | -0.017 | 0.011 | 0.686 | 0.840 | -0.009 | -0.020 | 0.002 | 0.097 | 0.358 | -0.012 | -0.040 | 0.015 | 0.376 | 0.592 |
| LPE(20:4) [sn1] | -0.019 | -0.041 | 0.003 | 0.095 | 0.903 | -0.010 | -0.024 | 0.004 | 0.157 | 0.432 | -0.009 | -0.020 | 0.002 | 0.109 | 0.389 | -0.038 | -0.065 | -0.010 | 0.008 | 0.053 |
| LPC(O-24:0) | -0.012 | -0.034 | 0.010 | 0.284 | 0.903 | -0.021 | -0.035 | -0.007 | 0.004 | 0.259 | -0.009 | -0.020 | 0.002 | 0.107 | 0.389 | -0.036 | -0.064 | -0.009 | 0.010 | 0.062 |
| LPE(18:0) [sn2] | -0.011 | -0.033 | 0.010 | 0.304 | 0.903 | -0.010 | -0.024 | 0.003 | 0.143 | 0.406 | -0.009 | -0.020 | 0.002 | 0.109 | 0.389 | -0.018 | -0.045 | 0.010 | 0.202 | 0.416 |
| PC(P-15:0/20:4) (a) | -0.007 | -0.029 | 0.015 | 0.529 | 0.922 | -0.007 | -0.021 | 0.007 | 0.317 | 0.557 | 0.009 | -0.002 | 0.019 | 0.108 | 0.389 | 0.021 | -0.006 | 0.048 | 0.131 | 0.317 |
| Cer(d20:1/23:0) | -0.007 | -0.029 | 0.015 | 0.527 | 0.922 | -0.006 | -0.020 | 0.008 | 0.395 | 0.634 | 0.009 | -0.002 | 0.020 | 0.110 | 0.389 | -0.014 | -0.041 | 0.013 | 0.314 | 0.537 |
| SHexCer(d18:1/24:1(OH)) | 0.005 | -0.017 | 0.027 | 0.667 | 0.964 | -0.005 | -0.019 | 0.009 | 0.493 | 0.710 | -0.009 | -0.020 | 0.002 | 0.109 | 0.389 | -0.017 | -0.046 | 0.011 | 0.239 | 0.452 |
| PC(O-16:0/22:6) | -0.002 | -0.024 | 0.020 | 0.851 | 0.981 | -0.008 | -0.022 | 0.006 | 0.285 | 0.530 | 0.009 | -0.002 | 0.020 | 0.108 | 0.389 | -0.008 | -0.036 | 0.020 | 0.573 | 0.745 |
| PC(16:1_22:6) | 0.001 | -0.021 | 0.023 | 0.942 | 0.995 | -0.012 | -0.026 | 0.002 | 0.085 | 0.356 | -0.009 | -0.020 | 0.002 | 0.106 | 0.389 | -0.038 | -0.065 | -0.011 | 0.007 | 0.048 |
| PC(38:6) (a) | 0.001 | -0.021 | 0.024 | 0.912 | 0.987 | -0.006 | -0.020 | 0.008 | 0.391 | 0.632 | -0.009 | -0.020 | 0.002 | 0.111 | 0.391 | -0.025 | -0.052 | 0.003 | 0.079 | 0.218 |
| TG(54:0) [NL-18:0] | -0.024 | -0.046 | -0.002 | 0.030 | 0.903 | -0.016 | -0.029 | -0.002 | 0.021 | 0.273 | 0.009 | -0.002 | 0.019 | 0.115 | 0.403 | 0.014 | -0.014 | 0.042 | 0.322 | 0.540 |
| Cer(d18:2/14:0) | -0.005 | -0.027 | 0.017 | 0.679 | 0.964 | -0.007 | -0.021 | 0.007 | 0.330 | 0.572 | -0.009 | -0.020 | 0.002 | 0.115 | 0.404 | -0.042 | -0.070 | -0.015 | 0.003 | 0.032 |
| Sph(d18:2) | 0.012 | -0.010 | 0.034 | 0.287 | 0.903 | -0.007 | -0.021 | 0.007 | 0.304 | 0.546 | 0.009 | -0.002 | 0.020 | 0.119 | 0.404 | 0.015 | -0.010 | 0.041 | 0.245 | 0.458 |
| TG(50:0) [NL-18:0] | -0.024 | -0.046 | -0.002 | 0.032 | 0.903 | -0.012 | -0.026 | 0.001 | 0.070 | 0.348 | 0.008 | -0.002 | 0.019 | 0.118 | 0.404 | 0.012 | -0.015 | 0.040 | 0.381 | 0.596 |
| PI(34:0) | -0.019 | -0.041 | 0.004 | 0.101 | 0.903 | -0.004 | -0.019 | 0.010 | 0.555 | 0.748 | 0.009 | -0.002 | 0.019 | 0.118 | 0.404 | -0.011 | -0.038 | 0.016 | 0.432 | 0.640 |
| TG(51:0) [NL-16:0] | -0.023 | -0.044 | -0.001 | 0.044 | 0.903 | -0.012 | -0.026 | 0.001 | 0.069 | 0.347 | 0.009 | -0.002 | 0.019 | 0.116 | 0.404 | 0.009 | -0.018 | 0.037 | 0.518 | 0.709 |
| TG(O-52:2) [NL-16:0] | -0.009 | -0.030 | 0.013 | 0.436 | 0.922 | -0.003 | -0.017 | 0.011 | 0.655 | 0.817 | 0.009 | -0.002 | 0.019 | 0.118 | 0.404 | 0.021 | -0.007 | 0.049 | 0.138 | 0.325 |
| PC(O-18:1/18:2) | -0.002 | -0.024 | 0.020 | 0.847 | 0.981 | -0.002 | -0.016 | 0.012 | 0.746 | 0.876 | -0.009 | -0.020 | 0.002 | 0.118 | 0.404 | -0.025 | -0.053 | 0.002 | 0.069 | 0.195 |
| SM(d18:1/16:0) | -0.010 | -0.031 | 0.012 | 0.396 | 0.922 | -0.007 | -0.021 | 0.007 | 0.332 | 0.572 | -0.009 | -0.020 | 0.002 | 0.119 | 0.404 | -0.050 | -0.077 | -0.023 | 0.000 | 0.008 |
| TG(O-50:2) [NL-18:2] | -0.013 | -0.035 | 0.009 | 0.247 | 0.903 | 0.002 | -0.012 | 0.016 | 0.781 | 0.884 | 0.009 | -0.002 | 0.019 | 0.121 | 0.405 | 0.025 | -0.003 | 0.053 | 0.078 | 0.216 |
| PI(16:0/16:0) | -0.019 | -0.041 | 0.003 | 0.096 | 0.903 | 0.001 | -0.013 | 0.014 | 0.944 | 0.971 | 0.009 | -0.002 | 0.019 | 0.121 | 0.405 | 0.002 | -0.025 | 0.029 | 0.901 | 0.952 |
| PC(17:1_18:2) | -0.005 | -0.027 | 0.018 | 0.689 | 0.964 | -0.013 | -0.026 | 0.001 | 0.076 | 0.356 | -0.009 | -0.019 | 0.002 | 0.120 | 0.405 | -0.029 | -0.056 | -0.003 | 0.032 | 0.127 |
| LPE(16:0) [sn1] | -0.017 | -0.039 | 0.005 | 0.129 | 0.903 | -0.017 | -0.030 | -0.003 | 0.015 | 0.273 | -0.008 | -0.019 | 0.002 | 0.122 | 0.406 | -0.027 | -0.054 | 0.001 | 0.055 | 0.173 |
| TG(56:8) [NL-22:6] | 0.000 | -0.022 | 0.023 | 0.965 | 0.995 | -0.011 | -0.025 | 0.003 | 0.132 | 0.395 | 0.009 | -0.002 | 0.019 | 0.124 | 0.411 | 0.010 | -0.018 | 0.038 | 0.468 | 0.665 |
| LPC(22:4) [sn2] | -0.012 | -0.034 | 0.009 | 0.269 | 0.903 | -0.004 | -0.018 | 0.010 | 0.560 | 0.748 | -0.008 | -0.019 | 0.002 | 0.129 | 0.426 | -0.019 | -0.046 | 0.009 | 0.181 | 0.387 |
| LPC(17:1) (a) [sn1] [104_sn1] | -0.014 | -0.036 | 0.008 | 0.229 | 0.903 | -0.019 | -0.032 | -0.005 | 0.007 | 0.259 | -0.008 | -0.019 | 0.002 | 0.131 | 0.429 | -0.030 | -0.057 | -0.003 | 0.032 | 0.127 |
| PC(16:0_18:0) | -0.010 | -0.032 | 0.012 | 0.385 | 0.922 | -0.008 | -0.022 | 0.006 | 0.247 | 0.501 | -0.008 | -0.019 | 0.002 | 0.131 | 0.429 | -0.040 | -0.067 | -0.013 | 0.003 | 0.037 |
| Cer(d17:1/16:0) | -0.004 | -0.026 | 0.019 | 0.736 | 0.964 | -0.001 | -0.015 | 0.013 | 0.880 | 0.934 | -0.008 | -0.019 | 0.003 | 0.133 | 0.432 | -0.010 | -0.038 | 0.017 | 0.469 | 0.665 |
| PI(16:0_20:3) (b) | -0.014 | -0.036 | 0.008 | 0.227 | 0.903 | -0.014 | -0.028 | 0.000 | 0.050 | 0.317 | -0.008 | -0.019 | 0.003 | 0.138 | 0.442 | -0.027 | -0.054 | 0.000 | 0.054 | 0.172 |
| TG(50:3) [NL-14:0] | -0.014 | -0.036 | 0.007 | 0.196 | 0.903 | -0.015 | -0.029 | -0.002 | 0.027 | 0.285 | -0.008 | -0.019 | 0.003 | 0.141 | 0.442 | -0.023 | -0.052 | 0.005 | 0.105 | 0.269 |
| LPC(18:3) (a) [sn1] [104_sn1] | -0.013 | -0.035 | 0.010 | 0.270 | 0.903 | -0.007 | -0.021 | 0.007 | 0.301 | 0.546 | -0.008 | -0.019 | 0.003 | 0.139 | 0.442 | -0.002 | -0.030 | 0.025 | 0.880 | 0.948 |
| TG(53:2) [NL-17:1] | -0.017 | -0.038 | 0.005 | 0.133 | 0.903 | -0.016 | -0.029 | -0.002 | 0.022 | 0.273 | 0.008 | -0.003 | 0.019 | 0.138 | 0.442 | 0.000 | -0.028 | 0.027 | 0.993 | 0.996 |
| S1P(d18:1) | -0.010 | -0.032 | 0.012 | 0.388 | 0.922 | -0.015 | -0.029 | -0.001 | 0.040 | 0.305 | -0.008 | -0.019 | 0.003 | 0.141 | 0.442 | -0.027 | -0.054 | 0.000 | 0.052 | 0.170 |
| CE(20:4) [+OH] | -0.005 | -0.027 | 0.018 | 0.686 | 0.964 | -0.004 | -0.018 | 0.010 | 0.564 | 0.748 | -0.008 | -0.019 | 0.003 | 0.139 | 0.442 | -0.020 | -0.048 | 0.008 | 0.169 | 0.373 |
| FA(20:4) | 0.001 | -0.021 | 0.023 | 0.912 | 0.987 | -0.004 | -0.018 | 0.010 | 0.583 | 0.763 | -0.008 | -0.019 | 0.003 | 0.139 | 0.442 | -0.015 | -0.042 | 0.013 | 0.299 | 0.523 |
| PA(36:3) | 0.000 | -0.021 | 0.022 | 0.968 | 0.995 | -0.007 | -0.021 | 0.008 | 0.366 | 0.602 | -0.008 | -0.019 | 0.003 | 0.140 | 0.442 | -0.008 | -0.035 | 0.020 | 0.583 | 0.753 |
| PE(O-16:0/22:6) | 0.000 | -0.022 | 0.022 | 0.997 | 0.999 | 0.001 | -0.013 | 0.016 | 0.872 | 0.932 | 0.008 | -0.003 | 0.019 | 0.137 | 0.442 | -0.001 | -0.029 | 0.027 | 0.931 | 0.964 |
| TG(O-50:2) [NL-18:1] | -0.002 | -0.024 | 0.020 | 0.842 | 0.981 | -0.010 | -0.024 | 0.004 | 0.172 | 0.438 | 0.008 | -0.003 | 0.019 | 0.143 | 0.447 | -0.001 | -0.030 | 0.027 | 0.925 | 0.964 |
| TG(54:1) [NL-18:1] | -0.021 | -0.042 | 0.001 | 0.065 | 0.903 | -0.013 | -0.027 | 0.000 | 0.055 | 0.317 | 0.008 | -0.003 | 0.019 | 0.144 | 0.448 | 0.012 | -0.016 | 0.040 | 0.404 | 0.620 |
| TG(58:8) [NL-22:6] | 0.007 | -0.015 | 0.029 | 0.559 | 0.933 | -0.011 | -0.025 | 0.003 | 0.133 | 0.395 | 0.008 | -0.003 | 0.019 | 0.146 | 0.451 | 0.004 | -0.024 | 0.032 | 0.793 | 0.904 |
| PC(O-16:0/20:3) | -0.007 | -0.029 | 0.015 | 0.518 | 0.922 | -0.008 | -0.022 | 0.006 | 0.254 | 0.506 | -0.008 | -0.019 | 0.003 | 0.150 | 0.457 | -0.039 | -0.065 | -0.012 | 0.005 | 0.044 |
| PC(18:1_20:3) | -0.009 | -0.031 | 0.013 | 0.413 | 0.922 | -0.010 | -0.024 | 0.003 | 0.142 | 0.406 | -0.008 | -0.019 | 0.003 | 0.150 | 0.457 | -0.034 | -0.061 | -0.007 | 0.014 | 0.074 |
| Cer(d17:1/24:1) | -0.010 | -0.032 | 0.012 | 0.364 | 0.922 | -0.017 | -0.031 | -0.003 | 0.018 | 0.273 | -0.008 | -0.019 | 0.003 | 0.149 | 0.457 | -0.029 | -0.057 | -0.002 | 0.038 | 0.138 |
| PE(P-18:1/18:1) (a) | 0.010 | -0.012 | 0.033 | 0.363 | 0.922 | 0.002 | -0.012 | 0.016 | 0.793 | 0.891 | 0.008 | -0.003 | 0.019 | 0.149 | 0.457 | -0.008 | -0.036 | 0.020 | 0.562 | 0.739 |
| DE(22:6) | 0.010 | -0.012 | 0.032 | 0.368 | 0.922 | 0.004 | -0.010 | 0.018 | 0.566 | 0.748 | -0.008 | -0.019 | 0.003 | 0.152 | 0.461 | -0.024 | -0.051 | 0.004 | 0.099 | 0.256 |

| | | | | | | | | | | | | | | | | | | | | |
|---|---|---|---|---|---|---|---|---|---|---|---|---|---|---|---|---|---|---|---|---|
| TG(O-52:0) [NL-16:0] | -0.023 | -0.045 | -0.002 | 0.035 | 0.903 | -0.009 | -0.022 | 0.004 | 0.190 | 0.454 | 0.008 | -0.003 | 0.018 | 0.156 | 0.463 | 0.008 | -0.019 | 0.035 | 0.552 | 0.730 |
| PC(16:1_20:4) | -0.009 | -0.031 | 0.013 | 0.421 | 0.922 | -0.003 | -0.017 | 0.011 | 0.696 | 0.845 | -0.008 | -0.019 | 0.003 | 0.157 | 0.463 | -0.032 | -0.060 | -0.005 | 0.023 | 0.099 |
| PC(P-16:0/20:4) | -0.010 | -0.032 | 0.012 | 0.389 | 0.922 | 0.001 | -0.013 | 0.015 | 0.845 | 0.927 | 0.008 | -0.003 | 0.019 | 0.157 | 0.463 | -0.002 | -0.030 | 0.025 | 0.860 | 0.937 |
| dhCer(d18:0/16:0) | -0.008 | -0.030 | 0.014 | 0.479 | 0.922 | -0.003 | -0.017 | 0.011 | 0.651 | 0.815 | 0.008 | -0.003 | 0.018 | 0.154 | 0.463 | 0.001 | -0.026 | 0.029 | 0.932 | 0.964 |
| PE(P-18:1/22:6) (b) | 0.004 | -0.018 | 0.026 | 0.734 | 0.964 | -0.006 | -0.020 | 0.009 | 0.435 | 0.666 | -0.008 | -0.019 | 0.003 | 0.155 | 0.463 | -0.036 | -0.062 | -0.009 | 0.008 | 0.053 |
| PC(18:2_18:2) | -0.004 | -0.026 | 0.018 | 0.742 | 0.964 | -0.001 | -0.015 | 0.013 | 0.855 | 0.929 | -0.008 | -0.019 | 0.003 | 0.154 | 0.463 | -0.003 | -0.031 | 0.024 | 0.815 | 0.918 |
| PC(P-17:0/20:4) (b) | -0.003 | -0.025 | 0.019 | 0.812 | 0.980 | -0.002 | -0.016 | 0.012 | 0.778 | 0.884 | 0.008 | -0.003 | 0.018 | 0.155 | 0.463 | -0.001 | -0.028 | 0.026 | 0.942 | 0.966 |
| LPC(18:2) [sn1] | -0.008 | -0.030 | 0.014 | 0.476 | 0.922 | -0.008 | -0.022 | 0.005 | 0.241 | 0.495 | -0.008 | -0.019 | 0.003 | 0.159 | 0.466 | -0.014 | -0.042 | 0.014 | 0.330 | 0.547 |
| PIP1(38:4) | -0.006 | -0.028 | 0.016 | 0.601 | 0.950 | -0.007 | -0.021 | 0.006 | 0.290 | 0.536 | -0.008 | -0.018 | 0.003 | 0.159 | 0.466 | -0.035 | -0.062 | -0.008 | 0.011 | 0.062 |
| TG(50:3) [NL-14:1] | -0.012 | -0.034 | 0.010 | 0.278 | 0.903 | -0.017 | -0.031 | -0.004 | 0.012 | 0.273 | -0.008 | -0.019 | 0.003 | 0.160 | 0.466 | -0.033 | -0.061 | -0.005 | 0.023 | 0.101 |
| CE(18:2) [+OH] | -0.008 | -0.030 | 0.014 | 0.468 | 0.922 | -0.012 | -0.026 | 0.002 | 0.089 | 0.356 | -0.008 | -0.019 | 0.003 | 0.162 | 0.472 | -0.017 | -0.045 | 0.010 | 0.225 | 0.434 |
| PC(P-18:1/18:1) | -0.003 | -0.025 | 0.019 | 0.779 | 0.973 | -0.008 | -0.022 | 0.005 | 0.236 | 0.493 | -0.008 | -0.019 | 0.003 | 0.164 | 0.474 | -0.035 | -0.062 | -0.008 | 0.012 | 0.066 |
| PI(18:0_20:4) | -0.019 | -0.041 | 0.002 | 0.077 | 0.903 | -0.014 | -0.027 | 0.000 | 0.055 | 0.317 | -0.008 | -0.018 | 0.003 | 0.165 | 0.475 | -0.040 | -0.067 | -0.013 | 0.004 | 0.038 |
| TG(O-52:1) [NL-16:0] | -0.015 | -0.036 | 0.007 | 0.177 | 0.903 | -0.007 | -0.020 | 0.007 | 0.337 | 0.574 | 0.008 | -0.003 | 0.018 | 0.165 | 0.475 | 0.009 | -0.018 | 0.036 | 0.529 | 0.718 |
| PE(P-18:1/22:6) (a) | 0.012 | -0.010 | 0.034 | 0.286 | 0.903 | -0.003 | -0.018 | 0.011 | 0.682 | 0.840 | 0.008 | -0.003 | 0.018 | 0.167 | 0.477 | -0.009 | -0.037 | 0.019 | 0.536 | 0.720 |
| TG(51:1) [NL-17:0] | -0.024 | -0.045 | -0.002 | 0.033 | 0.903 | -0.015 | -0.029 | -0.001 | 0.031 | 0.291 | 0.007 | -0.003 | 0.018 | 0.169 | 0.482 | -0.001 | -0.029 | 0.027 | 0.954 | 0.969 |
| SM(40:3) (a) | -0.010 | -0.032 | 0.012 | 0.389 | 0.922 | -0.007 | -0.021 | 0.006 | 0.293 | 0.536 | -0.008 | -0.018 | 0.003 | 0.172 | 0.484 | -0.045 | -0.071 | -0.018 | 0.001 | 0.018 |
| SHexCer(d18:1/16:0(OH)) | -0.007 | -0.029 | 0.015 | 0.533 | 0.923 | 0.001 | -0.013 | 0.015 | 0.889 | 0.939 | -0.008 | -0.018 | 0.003 | 0.172 | 0.484 | -0.032 | -0.060 | -0.004 | 0.025 | 0.106 |
| DG(18:2_18:2) | -0.006 | -0.028 | 0.016 | 0.576 | 0.933 | -0.002 | -0.016 | 0.012 | 0.779 | 0.884 | -0.008 | -0.018 | 0.003 | 0.171 | 0.484 | -0.012 | -0.039 | 0.015 | 0.394 | 0.609 |
| LPC(18:3) [sn1] (a) & LPC(18:3) [sn2] (b) | -0.013 | -0.035 | 0.010 | 0.261 | 0.903 | -0.004 | -0.018 | 0.010 | 0.561 | 0.748 | -0.007 | -0.018 | 0.003 | 0.174 | 0.487 | -0.001 | -0.028 | 0.026 | 0.941 | 0.966 |
| TG(52:3) [NL-18:2] | -0.010 | -0.031 | 0.011 | 0.367 | 0.922 | -0.012 | -0.026 | 0.002 | 0.092 | 0.356 | -0.007 | -0.018 | 0.003 | 0.174 | 0.487 | -0.027 | -0.055 | 0.001 | 0.057 | 0.174 |
| TG(52:4) [NL-18:3] | -0.019 | -0.041 | 0.002 | 0.081 | 0.903 | -0.015 | -0.029 | -0.002 | 0.028 | 0.285 | -0.007 | -0.018 | 0.003 | 0.180 | 0.487 | -0.022 | -0.050 | 0.006 | 0.123 | 0.300 |
| TG(52:1) [NL-18:0] | -0.018 | -0.040 | 0.004 | 0.108 | 0.903 | -0.013 | -0.027 | 0.000 | 0.057 | 0.317 | 0.007 | -0.003 | 0.018 | 0.179 | 0.487 | 0.006 | -0.021 | 0.034 | 0.657 | 0.814 |
| PS(40:5) | 0.017 | -0.005 | 0.039 | 0.131 | 0.903 | -0.013 | -0.027 | 0.000 | 0.058 | 0.319 | -0.007 | -0.018 | 0.003 | 0.179 | 0.487 | -0.004 | -0.032 | 0.024 | 0.778 | 0.894 |
| PE(O-16:0/20:4) | -0.008 | -0.030 | 0.014 | 0.452 | 0.922 | 0.002 | -0.012 | 0.016 | 0.742 | 0.874 | 0.007 | -0.003 | 0.018 | 0.179 | 0.487 | -0.002 | -0.030 | 0.026 | 0.886 | 0.948 |
| PI(36:2) | -0.007 | -0.028 | 0.015 | 0.547 | 0.929 | 0.000 | -0.015 | 0.014 | 0.948 | 0.972 | -0.007 | -0.018 | 0.003 | 0.178 | 0.487 | -0.018 | -0.045 | 0.009 | 0.191 | 0.402 |
| PE(P-18:1/18:1) (b) | -0.006 | -0.029 | 0.016 | 0.576 | 0.933 | 0.004 | -0.010 | 0.018 | 0.587 | 0.764 | -0.007 | -0.018 | 0.003 | 0.178 | 0.487 | -0.032 | -0.058 | -0.005 | 0.019 | 0.090 |
| TG(54:7) [NL-22:6] | -0.005 | -0.027 | 0.018 | 0.689 | 0.964 | -0.015 | -0.029 | -0.001 | 0.037 | 0.301 | 0.008 | -0.003 | 0.018 | 0.176 | 0.487 | 0.004 | -0.024 | 0.032 | 0.767 | 0.892 |
| AC(22:5) | 0.003 | -0.019 | 0.025 | 0.798 | 0.975 | -0.015 | -0.029 | -0.002 | 0.029 | 0.289 | -0.007 | -0.018 | 0.003 | 0.177 | 0.487 | -0.011 | -0.038 | 0.016 | 0.417 | 0.631 |
| PE(15-MHDA_18:1) | -0.015 | -0.037 | 0.008 | 0.203 | 0.903 | -0.014 | -0.028 | 0.000 | 0.045 | 0.306 | 0.007 | -0.004 | 0.018 | 0.187 | 0.499 | -0.019 | -0.047 | 0.009 | 0.188 | 0.401 |
| PE(16:1_18:2) | -0.021 | -0.043 | 0.001 | 0.063 | 0.903 | -0.013 | -0.026 | 0.001 | 0.073 | 0.350 | -0.007 | -0.018 | 0.004 | 0.189 | 0.499 | -0.017 | -0.044 | 0.010 | 0.205 | 0.416 |
| LPC(18:3) [sn1] (b) | -0.016 | -0.038 | 0.007 | 0.173 | 0.903 | -0.008 | -0.022 | 0.006 | 0.263 | 0.514 | -0.007 | -0.018 | 0.004 | 0.189 | 0.499 | 0.002 | -0.026 | 0.029 | 0.914 | 0.959 |
| LPC(P-16:0) | -0.010 | -0.032 | 0.012 | 0.369 | 0.922 | -0.007 | -0.021 | 0.007 | 0.324 | 0.563 | -0.007 | -0.018 | 0.004 | 0.187 | 0.499 | -0.015 | -0.042 | 0.012 | 0.284 | 0.509 |
| PE(P-16:0/20:3) (b) | -0.009 | -0.031 | 0.013 | 0.426 | 0.922 | -0.001 | -0.015 | 0.013 | 0.879 | 0.934 | 0.007 | -0.004 | 0.018 | 0.186 | 0.499 | -0.010 | -0.038 | 0.017 | 0.456 | 0.660 |
| PC(O-36:5) | 0.010 | -0.012 | 0.032 | 0.381 | 0.922 | -0.009 | -0.023 | 0.005 | 0.194 | 0.454 | 0.007 | -0.004 | 0.018 | 0.187 | 0.499 | -0.002 | -0.029 | 0.026 | 0.910 | 0.956 |
| PE(O-38:5) (a) | -0.007 | -0.029 | 0.016 | 0.560 | 0.933 | 0.007 | -0.007 | 0.021 | 0.309 | 0.550 | 0.007 | -0.004 | 0.018 | 0.188 | 0.499 | -0.009 | -0.037 | 0.019 | 0.529 | 0.718 |
| PE(O-38:5) (b) | -0.002 | -0.025 | 0.020 | 0.826 | 0.981 | -0.004 | -0.018 | 0.010 | 0.586 | 0.763 | 0.007 | -0.003 | 0.018 | 0.186 | 0.499 | -0.013 | -0.040 | 0.014 | 0.343 | 0.563 |
| HexCer(d18:1/24:0) | 0.012 | -0.010 | 0.034 | 0.285 | 0.903 | -0.001 | -0.015 | 0.012 | 0.859 | 0.929 | -0.007 | -0.018 | 0.004 | 0.191 | 0.501 | -0.014 | -0.042 | 0.013 | 0.304 | 0.527 |
| Cer(d18:2/23:0) | -0.009 | -0.031 | 0.013 | 0.431 | 0.922 | -0.005 | -0.019 | 0.009 | 0.512 | 0.724 | -0.007 | -0.018 | 0.004 | 0.191 | 0.501 | -0.022 | -0.050 | 0.005 | 0.110 | 0.276 |
| TG(O-50:1) [NL-17:1] | -0.007 | -0.029 | 0.014 | 0.512 | 0.922 | -0.006 | -0.020 | 0.008 | 0.393 | 0.633 | 0.007 | -0.004 | 0.018 | 0.194 | 0.505 | 0.019 | -0.008 | 0.047 | 0.173 | 0.378 |
| PE(16:1_20:4) | -0.022 | -0.044 | 0.000 | 0.053 | 0.903 | -0.010 | -0.024 | 0.003 | 0.142 | 0.406 | -0.007 | -0.018 | 0.004 | 0.194 | 0.505 | -0.014 | -0.041 | 0.013 | 0.306 | 0.527 |
| Cer(m18:1/23:0) | -0.016 | -0.038 | 0.006 | 0.166 | 0.903 | -0.008 | -0.022 | 0.006 | 0.251 | 0.503 | 0.007 | -0.004 | 0.018 | 0.196 | 0.509 | 0.001 | -0.026 | 0.029 | 0.925 | 0.964 |
| dhCer(d18:0/24:1) | -0.009 | -0.031 | 0.013 | 0.422 | 0.922 | -0.006 | -0.020 | 0.007 | 0.363 | 0.601 | 0.007 | -0.004 | 0.018 | 0.197 | 0.509 | -0.011 | -0.038 | 0.017 | 0.447 | 0.652 |
| PE(P-18:0/18:1) | 0.000 | -0.022 | 0.022 | 0.985 | 0.996 | 0.002 | -0.012 | 0.016 | 0.805 | 0.898 | 0.007 | -0.004 | 0.018 | 0.198 | 0.510 | -0.004 | -0.032 | 0.023 | 0.761 | 0.891 |
| PC(20:0_20:4) | 0.008 | -0.014 | 0.030 | 0.495 | 0.922 | 0.009 | -0.005 | 0.022 | 0.215 | 0.476 | -0.007 | -0.018 | 0.004 | 0.200 | 0.511 | -0.018 | -0.046 | 0.009 | 0.191 | 0.402 |
| SM(d18:2/22:0) | 0.004 | -0.018 | 0.026 | 0.728 | 0.964 | 0.004 | -0.009 | 0.018 | 0.534 | 0.738 | -0.007 | -0.018 | 0.004 | 0.201 | 0.511 | -0.029 | -0.055 | -0.002 | 0.037 | 0.134 |
| Hex2Cer(d16:1/16:0) | -0.002 | -0.024 | 0.021 | 0.883 | 0.981 | -0.002 | -0.016 | 0.012 | 0.775 | 0.884 | -0.007 | -0.018 | 0.004 | 0.200 | 0.511 | -0.020 | -0.047 | 0.008 | 0.162 | 0.362 |
| Cer(d16:1/20:0) | -0.015 | -0.037 | 0.007 | 0.187 | 0.903 | -0.013 | -0.026 | 0.001 | 0.071 | 0.349 | -0.007 | -0.018 | 0.004 | 0.202 | 0.512 | -0.026 | -0.054 | 0.001 | 0.064 | 0.184 |
| PC(O-34:1) | -0.009 | -0.031 | 0.013 | 0.445 | 0.922 | -0.012 | -0.026 | 0.002 | 0.097 | 0.361 | -0.007 | -0.018 | 0.004 | 0.202 | 0.512 | -0.035 | -0.062 | -0.008 | 0.012 | 0.067 |
| PC(O-16:0/16:0) | -0.016 | -0.038 | 0.006 | 0.149 | 0.903 | -0.011 | -0.025 | 0.003 | 0.114 | 0.382 | -0.007 | -0.018 | 0.004 | 0.205 | 0.516 | -0.033 | -0.060 | -0.006 | 0.017 | 0.085 |

| | | | | | | | | | | | | | | | | | | | | |
|---|---|---|---|---|---|---|---|---|---|---|---|---|---|---|---|---|---|---|---|---|
| PI(18:0_20:3) (a) | -0.022 | -0.044 | 0.000 | 0.054 | 0.903 | -0.016 | -0.030 | -0.003 | 0.021 | 0.273 | -0.007 | -0.018 | 0.004 | 0.209 | 0.524 | -0.035 | -0.063 | -0.008 | 0.013 | 0.068 |
| PE(16:0_18:3) (a) | -0.019 | -0.041 | 0.003 | 0.097 | 0.903 | -0.007 | -0.021 | 0.007 | 0.312 | 0.555 | -0.007 | -0.018 | 0.004 | 0.209 | 0.524 | -0.011 | -0.038 | 0.016 | 0.423 | 0.635 |
| SHexCer(d18:1/16:0) | -0.009 | -0.031 | 0.013 | 0.442 | 0.922 | -0.011 | -0.025 | 0.003 | 0.124 | 0.385 | -0.007 | -0.018 | 0.004 | 0.212 | 0.528 | -0.025 | -0.052 | 0.003 | 0.078 | 0.217 |
| PI(18:0_20:2) | -0.020 | -0.041 | 0.002 | 0.073 | 0.903 | -0.017 | -0.032 | -0.003 | 0.016 | 0.273 | -0.007 | -0.018 | 0.004 | 0.212 | 0.528 | -0.027 | -0.053 | 0.000 | 0.047 | 0.157 |
| TG(50:4) [NL-14:0] | -0.014 | -0.036 | 0.008 | 0.213 | 0.903 | -0.009 | -0.023 | 0.005 | 0.192 | 0.454 | -0.007 | -0.018 | 0.004 | 0.213 | 0.529 | -0.005 | -0.032 | 0.023 | 0.750 | 0.882 |
| FA(18:3) | 0.001 | -0.021 | 0.023 | 0.937 | 0.995 | 0.003 | -0.011 | 0.017 | 0.701 | 0.846 | -0.007 | -0.018 | 0.004 | 0.215 | 0.531 | -0.009 | -0.037 | 0.019 | 0.545 | 0.728 |
| Cer(d19:1/20:0) | -0.001 | -0.023 | 0.021 | 0.959 | 0.995 | -0.008 | -0.022 | 0.006 | 0.278 | 0.524 | -0.007 | -0.018 | 0.004 | 0.216 | 0.532 | -0.039 | -0.068 | -0.011 | 0.006 | 0.048 |
| TG(52:5) [NL-18:3] | -0.012 | -0.034 | 0.010 | 0.280 | 0.903 | -0.006 | -0.020 | 0.008 | 0.396 | 0.634 | -0.007 | -0.017 | 0.004 | 0.221 | 0.539 | -0.007 | -0.035 | 0.020 | 0.605 | 0.768 |
| PE(O-18:0/20:4) | -0.008 | -0.030 | 0.014 | 0.463 | 0.922 | 0.005 | -0.009 | 0.019 | 0.495 | 0.711 | 0.007 | -0.004 | 0.017 | 0.221 | 0.539 | 0.008 | -0.019 | 0.035 | 0.570 | 0.745 |
| AC(22:5)-OH | 0.003 | -0.019 | 0.025 | 0.808 | 0.980 | -0.007 | -0.020 | 0.007 | 0.350 | 0.592 | 0.007 | -0.004 | 0.018 | 0.220 | 0.539 | 0.005 | -0.022 | 0.033 | 0.703 | 0.845 |
| Hex3Cer(d18:1/24:0) | -0.001 | -0.023 | 0.021 | 0.946 | 0.995 | -0.004 | -0.018 | 0.010 | 0.565 | 0.748 | -0.007 | -0.017 | 0.004 | 0.220 | 0.539 | -0.028 | -0.055 | -0.001 | 0.045 | 0.153 |
| LPC(16:1) [sn2] | -0.014 | -0.036 | 0.008 | 0.217 | 0.903 | -0.010 | -0.023 | 0.004 | 0.166 | 0.432 | -0.007 | -0.017 | 0.004 | 0.222 | 0.539 | -0.026 | -0.053 | 0.001 | 0.056 | 0.173 |
| Cer(d18:1/22:0) | -0.012 | -0.034 | 0.010 | 0.290 | 0.903 | -0.001 | -0.015 | 0.013 | 0.885 | 0.938 | -0.007 | -0.017 | 0.004 | 0.229 | 0.551 | -0.018 | -0.045 | 0.009 | 0.189 | 0.401 |
| SHexCer(d18:1/24:0(OH)) | -0.011 | -0.034 | 0.011 | 0.323 | 0.922 | 0.004 | -0.010 | 0.018 | 0.563 | 0.748 | -0.007 | -0.017 | 0.004 | 0.229 | 0.551 | -0.023 | -0.051 | 0.006 | 0.116 | 0.289 |
| CE(20:4) | 0.008 | -0.014 | 0.030 | 0.482 | 0.922 | 0.014 | 0.000 | 0.028 | 0.044 | 0.306 | -0.007 | -0.017 | 0.004 | 0.231 | 0.552 | -0.018 | -0.046 | 0.010 | 0.209 | 0.419 |
| CE(18:3) | 0.003 | -0.019 | 0.025 | 0.794 | 0.974 | 0.004 | -0.009 | 0.018 | 0.539 | 0.742 | -0.007 | -0.017 | 0.004 | 0.230 | 0.552 | -0.016 | -0.044 | 0.011 | 0.239 | 0.452 |
| LPC(15-MHDA) [sn1] [104_sn1] | -0.007 | -0.029 | 0.015 | 0.546 | 0.929 | -0.014 | -0.028 | -0.001 | 0.041 | 0.306 | -0.007 | -0.017 | 0.004 | 0.232 | 0.552 | -0.031 | -0.059 | -0.003 | 0.028 | 0.112 |
| PC(P-16:0/18:0) | -0.012 | -0.034 | 0.010 | 0.286 | 0.903 | -0.006 | -0.020 | 0.008 | 0.372 | 0.608 | -0.007 | -0.017 | 0.004 | 0.233 | 0.555 | -0.037 | -0.063 | -0.010 | 0.007 | 0.049 |
| PE(O-18:0/22:6) | 0.000 | -0.023 | 0.022 | 0.987 | 0.996 | -0.001 | -0.016 | 0.013 | 0.847 | 0.927 | 0.007 | -0.004 | 0.017 | 0.235 | 0.557 | 0.002 | -0.026 | 0.030 | 0.896 | 0.952 |
| Cer(d17:1/20:0) | -0.014 | -0.036 | 0.008 | 0.214 | 0.903 | -0.014 | -0.028 | 0.000 | 0.053 | 0.317 | -0.006 | -0.017 | 0.004 | 0.240 | 0.562 | -0.035 | -0.063 | -0.007 | 0.014 | 0.074 |
| PG(34:1) | -0.015 | -0.037 | 0.007 | 0.188 | 0.903 | -0.020 | -0.034 | -0.007 | 0.004 | 0.259 | -0.006 | -0.017 | 0.004 | 0.240 | 0.562 | -0.017 | -0.045 | 0.011 | 0.225 | 0.434 |
| LPC(17:1) [sn1] (a) & LPC(17:1) [sn2] (b) | -0.008 | -0.031 | 0.014 | 0.454 | 0.922 | -0.018 | -0.031 | -0.004 | 0.010 | 0.270 | -0.006 | -0.017 | 0.004 | 0.240 | 0.562 | -0.028 | -0.055 | -0.001 | 0.043 | 0.149 |
| DE(16:0) | 0.009 | -0.013 | 0.031 | 0.418 | 0.922 | 0.016 | 0.003 | 0.030 | 0.021 | 0.273 | -0.006 | -0.017 | 0.004 | 0.238 | 0.562 | -0.017 | -0.043 | 0.010 | 0.210 | 0.420 |
| PE(P-18:1/22:5) (a) | 0.004 | -0.018 | 0.026 | 0.730 | 0.964 | 0.000 | -0.015 | 0.014 | 0.966 | 0.980 | -0.007 | -0.017 | 0.004 | 0.244 | 0.565 | -0.049 | -0.076 | -0.022 | 0.000 | 0.009 |
| SM(43:1) | 0.004 | -0.018 | 0.026 | 0.708 | 0.964 | -0.004 | -0.018 | 0.010 | 0.588 | 0.764 | 0.006 | -0.004 | 0.017 | 0.244 | 0.565 | -0.018 | -0.045 | 0.009 | 0.195 | 0.408 |
| PE(P-20:0/18:1) | -0.004 | -0.026 | 0.018 | 0.744 | 0.964 | -0.005 | -0.020 | 0.009 | 0.445 | 0.672 | 0.006 | -0.004 | 0.017 | 0.244 | 0.565 | -0.002 | -0.030 | 0.025 | 0.872 | 0.944 |
| CE(22:6) [+OH] | -0.001 | -0.023 | 0.022 | 0.956 | 0.995 | -0.011 | -0.024 | 0.003 | 0.120 | 0.385 | -0.007 | -0.018 | 0.005 | 0.244 | 0.565 | -0.015 | -0.043 | 0.013 | 0.287 | 0.509 |
| TG(52:1) [NL-18:1] | -0.020 | -0.042 | 0.002 | 0.074 | 0.903 | -0.014 | -0.027 | 0.000 | 0.051 | 0.317 | 0.006 | -0.004 | 0.017 | 0.250 | 0.565 | 0.004 | -0.024 | 0.032 | 0.789 | 0.904 |
| TG(O-54:2) [NL-18:1] | -0.007 | -0.029 | 0.015 | 0.515 | 0.922 | -0.004 | -0.018 | 0.009 | 0.542 | 0.743 | 0.006 | -0.004 | 0.017 | 0.246 | 0.565 | 0.005 | -0.022 | 0.033 | 0.706 | 0.845 |
| SM(d18:2/17:0) | -0.003 | -0.025 | 0.019 | 0.766 | 0.969 | -0.003 | -0.017 | 0.011 | 0.668 | 0.829 | -0.006 | -0.017 | 0.004 | 0.247 | 0.565 | -0.048 | -0.074 | -0.021 | 0.001 | 0.012 |
| CE(24:1) | -0.003 | -0.026 | 0.019 | 0.786 | 0.973 | -0.002 | -0.015 | 0.012 | 0.814 | 0.903 | -0.006 | -0.017 | 0.004 | 0.249 | 0.565 | -0.027 | -0.055 | 0.000 | 0.047 | 0.156 |
| PI(20:0_20:4) | -0.003 | -0.025 | 0.019 | 0.791 | 0.973 | 0.001 | -0.013 | 0.015 | 0.899 | 0.942 | -0.006 | -0.017 | 0.004 | 0.247 | 0.565 | -0.010 | -0.039 | 0.018 | 0.478 | 0.672 |
| AC(16:1)-OH | -0.003 | -0.025 | 0.019 | 0.813 | 0.980 | -0.010 | -0.024 | 0.004 | 0.145 | 0.406 | -0.006 | -0.017 | 0.004 | 0.250 | 0.565 | -0.036 | -0.063 | -0.008 | 0.011 | 0.063 |
| LPC(19:0) (a) [sn1] [104_sn1] | 0.002 | -0.021 | 0.024 | 0.889 | 0.981 | -0.009 | -0.022 | 0.005 | 0.231 | 0.488 | -0.006 | -0.017 | 0.004 | 0.249 | 0.565 | -0.015 | -0.042 | 0.013 | 0.294 | 0.518 |
| Hex3Cer(d18:1/24:1) | -0.005 | -0.028 | 0.017 | 0.629 | 0.954 | -0.007 | -0.022 | 0.007 | 0.307 | 0.549 | -0.006 | -0.017 | 0.004 | 0.252 | 0.568 | -0.040 | -0.068 | -0.012 | 0.005 | 0.044 |
| Sph(d18:1) | 0.006 | -0.016 | 0.028 | 0.609 | 0.950 | -0.006 | -0.020 | 0.009 | 0.449 | 0.673 | -0.006 | -0.017 | 0.005 | 0.253 | 0.568 | 0.006 | -0.022 | 0.034 | 0.681 | 0.827 |
| DG(18:1_20:5) | -0.004 | -0.026 | 0.018 | 0.729 | 0.964 | -0.011 | -0.025 | 0.003 | 0.122 | 0.385 | 0.006 | -0.005 | 0.017 | 0.254 | 0.569 | -0.002 | -0.030 | 0.026 | 0.894 | 0.952 |
| PC(P-17:0/20:4) (a) | -0.002 | -0.024 | 0.020 | 0.871 | 0.981 | -0.007 | -0.020 | 0.007 | 0.357 | 0.598 | 0.006 | -0.004 | 0.017 | 0.255 | 0.569 | 0.002 | -0.025 | 0.029 | 0.888 | 0.948 |
| PE(P-16:0/18:2) | -0.012 | -0.034 | 0.011 | 0.306 | 0.903 | 0.002 | -0.012 | 0.016 | 0.804 | 0.898 | 0.006 | -0.005 | 0.017 | 0.258 | 0.572 | -0.003 | -0.031 | 0.024 | 0.812 | 0.917 |
| TG(52:5) [NL-20:5] | -0.007 | -0.029 | 0.015 | 0.545 | 0.929 | -0.009 | -0.023 | 0.004 | 0.179 | 0.450 | 0.006 | -0.005 | 0.017 | 0.257 | 0.572 | 0.005 | -0.023 | 0.033 | 0.720 | 0.854 |
| TG(58:10) [NL-22:6] | 0.006 | -0.016 | 0.028 | 0.609 | 0.950 | -0.006 | -0.020 | 0.008 | 0.367 | 0.603 | 0.006 | -0.005 | 0.017 | 0.262 | 0.581 | 0.020 | -0.007 | 0.048 | 0.151 | 0.345 |
| LPC(22:5) [sn1] (n6) | -0.012 | -0.034 | 0.009 | 0.270 | 0.903 | -0.003 | -0.017 | 0.011 | 0.672 | 0.831 | -0.006 | -0.017 | 0.005 | 0.263 | 0.581 | -0.011 | -0.038 | 0.016 | 0.428 | 0.638 |
| SM(d18:1/23:0) & SM(d17:1/24:0) | -0.004 | -0.026 | 0.018 | 0.738 | 0.964 | 0.000 | -0.014 | 0.013 | 0.948 | 0.972 | 0.006 | -0.005 | 0.017 | 0.264 | 0.582 | -0.016 | -0.042 | 0.011 | 0.249 | 0.464 |
| HexCer(d18:1/16:0) | 0.011 | -0.012 | 0.033 | 0.349 | 0.922 | -0.004 | -0.018 | 0.010 | 0.594 | 0.767 | -0.006 | -0.017 | 0.005 | 0.267 | 0.586 | -0.037 | -0.065 | -0.010 | 0.008 | 0.051 |
| LPC(18:2) [sn2] | -0.002 | -0.025 | 0.020 | 0.839 | 0.981 | -0.003 | -0.017 | 0.011 | 0.650 | 0.814 | -0.006 | -0.017 | 0.005 | 0.268 | 0.587 | -0.006 | -0.034 | 0.022 | 0.668 | 0.818 |
| S1P(d18:2) | -0.009 | -0.031 | 0.013 | 0.429 | 0.922 | 0.002 | -0.011 | 0.016 | 0.733 | 0.870 | -0.006 | -0.017 | 0.005 | 0.270 | 0.588 | -0.017 | -0.044 | 0.010 | 0.214 | 0.422 |
| PE(17:0_18:2) | -0.011 | -0.033 | 0.011 | 0.320 | 0.922 | -0.016 | -0.029 | -0.002 | 0.026 | 0.285 | -0.006 | -0.017 | 0.005 | 0.271 | 0.588 | -0.013 | -0.040 | 0.015 | 0.367 | 0.586 |
| PC(18:0_18:2) | -0.007 | -0.029 | 0.015 | 0.530 | 0.922 | -0.009 | -0.023 | 0.005 | 0.206 | 0.467 | -0.006 | -0.017 | 0.005 | 0.271 | 0.588 | -0.036 | -0.064 | -0.009 | 0.010 | 0.061 |

| | | | | | | | | | | | | | | | | | | | | |
|---|---|---|---|---|---|---|---|---|---|---|---|---|---|---|---|---|---|---|---|---|
| PC(16:0_18:3) (b) | -0.021 | -0.043 | 0.001 | 0.065 | 0.903 | -0.009 | -0.023 | 0.005 | 0.203 | 0.465 | -0.006 | -0.017 | 0.005 | 0.273 | 0.588 | -0.035 | -0.062 | -0.007 | 0.013 | 0.070 |
| TG(48:0) [NL-16:0] | -0.023 | -0.045 | -0.001 | 0.041 | 0.903 | -0.005 | -0.019 | 0.009 | 0.471 | 0.689 | 0.006 | -0.005 | 0.017 | 0.273 | 0.588 | 0.013 | -0.014 | 0.041 | 0.337 | 0.554 |
| LPC(24:0) [sn1] | 0.011 | -0.011 | 0.033 | 0.321 | 0.922 | 0.002 | -0.012 | 0.016 | 0.749 | 0.877 | -0.006 | -0.017 | 0.005 | 0.273 | 0.588 | 0.005 | -0.023 | 0.033 | 0.710 | 0.847 |
| PC(18:0_22:5) (n3) & PC(20:1_20:4) | -0.007 | -0.029 | 0.015 | 0.520 | 0.922 | -0.010 | -0.024 | 0.003 | 0.145 | 0.406 | -0.006 | -0.017 | 0.005 | 0.275 | 0.589 | -0.055 | -0.082 | -0.027 | 0.000 | 0.006 |
| LPC(P-17:0) (a) | -0.003 | -0.025 | 0.019 | 0.766 | 0.969 | -0.010 | -0.024 | 0.003 | 0.142 | 0.406 | -0.006 | -0.017 | 0.005 | 0.275 | 0.589 | -0.020 | -0.047 | 0.008 | 0.159 | 0.358 |
| AC(13:0) | 0.011 | -0.011 | 0.033 | 0.326 | 0.922 | -0.012 | -0.025 | 0.002 | 0.097 | 0.361 | 0.006 | -0.005 | 0.017 | 0.277 | 0.590 | 0.010 | -0.018 | 0.038 | 0.484 | 0.676 |
| LPC(15-MHDA) [sn2] | -0.006 | -0.028 | 0.017 | 0.614 | 0.950 | -0.014 | -0.028 | 0.000 | 0.051 | 0.317 | -0.006 | -0.017 | 0.005 | 0.279 | 0.590 | -0.030 | -0.058 | -0.002 | 0.034 | 0.132 |
| CE(20:3) | 0.003 | -0.019 | 0.025 | 0.813 | 0.980 | -0.002 | -0.015 | 0.012 | 0.817 | 0.905 | -0.006 | -0.017 | 0.005 | 0.279 | 0.590 | -0.039 | -0.067 | -0.011 | 0.006 | 0.048 |
| SHexCer(d18:1/24:1) | -0.001 | -0.023 | 0.021 | 0.910 | 0.987 | -0.006 | -0.020 | 0.007 | 0.364 | 0.602 | -0.006 | -0.017 | 0.005 | 0.279 | 0.590 | -0.024 | -0.052 | 0.003 | 0.081 | 0.220 |
| TG(48:0) [NL-18:0] | -0.021 | -0.043 | 0.001 | 0.062 | 0.903 | -0.009 | -0.022 | 0.004 | 0.190 | 0.454 | 0.006 | -0.005 | 0.017 | 0.284 | 0.594 | 0.010 | -0.018 | 0.037 | 0.502 | 0.699 |
| Cer(d16:1/24:1) | -0.011 | -0.033 | 0.011 | 0.334 | 0.922 | -0.019 | -0.033 | -0.005 | 0.008 | 0.265 | -0.006 | -0.017 | 0.005 | 0.284 | 0.594 | -0.024 | -0.052 | 0.004 | 0.088 | 0.236 |
| TG(56:7) [NL-20:5] | 0.006 | -0.016 | 0.028 | 0.615 | 0.950 | -0.010 | -0.024 | 0.004 | 0.158 | 0.432 | 0.006 | -0.005 | 0.017 | 0.282 | 0.594 | -0.003 | -0.031 | 0.025 | 0.827 | 0.923 |
| PC(35:5) | 0.004 | -0.018 | 0.026 | 0.723 | 0.964 | -0.008 | -0.022 | 0.005 | 0.236 | 0.493 | 0.006 | -0.005 | 0.017 | 0.284 | 0.594 | -0.012 | -0.040 | 0.016 | 0.400 | 0.617 |
| TG(50:3) [NL-18:2] | -0.018 | -0.039 | 0.004 | 0.112 | 0.903 | -0.014 | -0.028 | -0.001 | 0.040 | 0.306 | -0.006 | -0.016 | 0.005 | 0.290 | 0.597 | -0.029 | -0.057 | -0.001 | 0.045 | 0.153 |
| PE(16:0_18:3) (b) | -0.026 | -0.048 | -0.004 | 0.022 | 0.903 | -0.008 | -0.022 | 0.005 | 0.224 | 0.485 | -0.006 | -0.017 | 0.005 | 0.289 | 0.597 | -0.010 | -0.037 | 0.017 | 0.460 | 0.663 |
| LPC(19:1) (c) | -0.006 | -0.029 | 0.016 | 0.589 | 0.948 | -0.011 | -0.025 | 0.002 | 0.096 | 0.361 | -0.006 | -0.017 | 0.005 | 0.290 | 0.597 | -0.029 | -0.056 | -0.002 | 0.039 | 0.139 |
| FA(17:1) | -0.004 | -0.027 | 0.019 | 0.738 | 0.964 | 0.004 | -0.010 | 0.018 | 0.560 | 0.748 | 0.006 | -0.005 | 0.017 | 0.288 | 0.597 | 0.013 | -0.014 | 0.040 | 0.335 | 0.553 |
| LPI(18:2) [sn2] | -0.004 | -0.027 | 0.018 | 0.691 | 0.964 | 0.006 | -0.008 | 0.020 | 0.373 | 0.608 | -0.006 | -0.017 | 0.005 | 0.289 | 0.597 | -0.010 | -0.037 | 0.017 | 0.461 | 0.664 |
| Hex2Cer(d18:2/24:1) | 0.002 | -0.021 | 0.024 | 0.886 | 0.981 | 0.000 | -0.014 | 0.014 | 0.998 | 0.998 | -0.006 | -0.017 | 0.005 | 0.288 | 0.597 | -0.024 | -0.052 | 0.004 | 0.091 | 0.241 |
| PE(P-16:0/22:5) (n3) | -0.015 | -0.037 | 0.007 | 0.189 | 0.903 | 0.006 | -0.009 | 0.020 | 0.445 | 0.672 | 0.006 | -0.005 | 0.017 | 0.293 | 0.598 | -0.024 | -0.052 | 0.003 | 0.086 | 0.230 |
| TG(58:9) [NL-22:6] | 0.008 | -0.014 | 0.030 | 0.472 | 0.922 | -0.010 | -0.024 | 0.004 | 0.161 | 0.432 | 0.006 | -0.005 | 0.017 | 0.292 | 0.598 | 0.013 | -0.015 | 0.041 | 0.359 | 0.577 |
| PE(18:0_22:6) | -0.005 | -0.027 | 0.017 | 0.652 | 0.960 | -0.011 | -0.025 | 0.002 | 0.109 | 0.379 | 0.006 | -0.005 | 0.017 | 0.294 | 0.598 | -0.003 | -0.031 | 0.024 | 0.804 | 0.910 |
| PE(O-34:1) | 0.001 | -0.021 | 0.024 | 0.908 | 0.987 | 0.005 | -0.009 | 0.019 | 0.506 | 0.722 | 0.006 | -0.005 | 0.017 | 0.294 | 0.598 | -0.003 | -0.030 | 0.025 | 0.850 | 0.933 |
| Cer(d17:1/18:0) | -0.013 | -0.036 | 0.009 | 0.238 | 0.903 | -0.008 | -0.022 | 0.006 | 0.248 | 0.501 | -0.006 | -0.016 | 0.005 | 0.295 | 0.598 | -0.037 | -0.065 | -0.010 | 0.008 | 0.053 |
| PE(16:0_16:0) | -0.023 | -0.045 | -0.002 | 0.036 | 0.903 | -0.008 | -0.021 | 0.006 | 0.249 | 0.501 | -0.006 | -0.016 | 0.005 | 0.298 | 0.598 | -0.008 | -0.035 | 0.019 | 0.552 | 0.730 |
| Cer(d20:1/22:0) | -0.009 | -0.030 | 0.013 | 0.440 | 0.922 | 0.003 | -0.011 | 0.017 | 0.673 | 0.831 | 0.006 | -0.005 | 0.016 | 0.296 | 0.598 | -0.007 | -0.034 | 0.020 | 0.601 | 0.765 |
| Hex2Cer(d18:1/24:1) | -0.005 | -0.027 | 0.017 | 0.654 | 0.960 | -0.008 | -0.022 | 0.006 | 0.239 | 0.495 | -0.006 | -0.017 | 0.005 | 0.298 | 0.598 | -0.024 | -0.051 | 0.004 | 0.096 | 0.250 |
| PC(39:5) (b) | -0.002 | -0.024 | 0.021 | 0.863 | 0.981 | -0.015 | -0.028 | -0.001 | 0.033 | 0.298 | -0.006 | -0.016 | 0.005 | 0.297 | 0.598 | -0.051 | -0.078 | -0.024 | 0.000 | 0.008 |
| HexCer(d18:2/20:0) | 0.000 | -0.022 | 0.022 | 0.993 | 0.997 | -0.006 | -0.020 | 0.008 | 0.411 | 0.646 | -0.006 | -0.017 | 0.005 | 0.299 | 0.598 | -0.018 | -0.046 | 0.010 | 0.204 | 0.416 |
| Cer(m18:1/24:0) | -0.017 | -0.039 | 0.005 | 0.126 | 0.903 | -0.006 | -0.020 | 0.008 | 0.405 | 0.643 | 0.006 | -0.005 | 0.016 | 0.304 | 0.605 | 0.003 | -0.024 | 0.031 | 0.819 | 0.920 |
| TG(54:3) [NL-18:1] | 0.006 | -0.016 | 0.027 | 0.616 | 0.950 | -0.014 | -0.028 | 0.000 | 0.048 | 0.317 | -0.006 | -0.016 | 0.005 | 0.304 | 0.605 | -0.017 | -0.045 | 0.010 | 0.216 | 0.425 |
| TG(56:9) [NL-22:6] | 0.003 | -0.019 | 0.025 | 0.783 | 0.973 | -0.009 | -0.023 | 0.005 | 0.186 | 0.453 | 0.006 | -0.005 | 0.017 | 0.306 | 0.605 | 0.014 | -0.014 | 0.041 | 0.337 | 0.554 |
| PE(18:1_22:6) (a) | -0.003 | -0.024 | 0.019 | 0.820 | 0.980 | -0.014 | -0.028 | 0.000 | 0.044 | 0.306 | -0.006 | -0.016 | 0.005 | 0.305 | 0.605 | -0.010 | -0.038 | 0.018 | 0.471 | 0.665 |
| CE(18:1) | 0.000 | -0.022 | 0.022 | 0.976 | 0.995 | -0.015 | -0.029 | -0.002 | 0.030 | 0.289 | -0.006 | -0.017 | 0.005 | 0.307 | 0.605 | -0.032 | -0.059 | -0.004 | 0.025 | 0.105 |
| LPC(26:0) [sn1] | 0.000 | -0.023 | 0.022 | 0.973 | 0.995 | -0.009 | -0.023 | 0.005 | 0.214 | 0.476 | -0.006 | -0.017 | 0.005 | 0.308 | 0.605 | -0.029 | -0.056 | -0.001 | 0.042 | 0.147 |
| FA(18:2) | 0.000 | -0.022 | 0.022 | 0.998 | 0.999 | 0.005 | -0.009 | 0.018 | 0.511 | 0.724 | -0.006 | -0.017 | 0.005 | 0.307 | 0.605 | -0.007 | -0.034 | 0.021 | 0.636 | 0.798 |
| TG(50:3) [NL-16:1] | -0.021 | -0.043 | 0.001 | 0.059 | 0.903 | -0.017 | -0.031 | -0.004 | 0.014 | 0.273 | -0.006 | -0.016 | 0.005 | 0.309 | 0.606 | -0.030 | -0.058 | -0.003 | 0.032 | 0.126 |
| DG(18:0_18:1) | -0.017 | -0.038 | 0.005 | 0.136 | 0.903 | -0.015 | -0.029 | -0.002 | 0.026 | 0.285 | 0.006 | -0.005 | 0.016 | 0.311 | 0.609 | -0.003 | -0.031 | 0.025 | 0.853 | 0.933 |
| PE(P-18:0/20:3) (a) | -0.009 | -0.031 | 0.014 | 0.449 | 0.922 | -0.004 | -0.018 | 0.010 | 0.566 | 0.748 | 0.006 | -0.005 | 0.016 | 0.313 | 0.610 | -0.015 | -0.042 | 0.012 | 0.275 | 0.499 |
| PS(38:3) | -0.002 | -0.024 | 0.020 | 0.880 | 0.981 | -0.001 | -0.015 | 0.012 | 0.857 | 0.929 | 0.006 | -0.005 | 0.016 | 0.314 | 0.611 | 0.018 | -0.009 | 0.046 | 0.181 | 0.387 |
| SM(d18:1/18:0) & SM(d16:1/20:0) | -0.017 | -0.039 | 0.005 | 0.129 | 0.903 | -0.008 | -0.021 | 0.006 | 0.276 | 0.524 | -0.006 | -0.016 | 0.005 | 0.316 | 0.613 | -0.047 | -0.075 | -0.020 | 0.001 | 0.014 |
| PE(P-20:0/18:2) | -0.003 | -0.025 | 0.019 | 0.813 | 0.980 | 0.001 | -0.013 | 0.015 | 0.936 | 0.968 | -0.006 | -0.016 | 0.005 | 0.317 | 0.613 | -0.014 | -0.042 | 0.013 | 0.297 | 0.521 |
| PI(16:0_16:1) | -0.017 | -0.039 | 0.005 | 0.135 | 0.903 | -0.001 | -0.015 | 0.013 | 0.914 | 0.951 | 0.005 | -0.005 | 0.016 | 0.321 | 0.616 | -0.004 | -0.031 | 0.023 | 0.774 | 0.893 |
| Cer(m18:1/22:0) | -0.014 | -0.036 | 0.008 | 0.219 | 0.903 | -0.008 | -0.022 | 0.006 | 0.291 | 0.536 | 0.005 | -0.005 | 0.016 | 0.320 | 0.616 | 0.004 | -0.024 | 0.031 | 0.796 | 0.905 |
| PE(P-20:1/22:6) | 0.004 | -0.018 | 0.027 | 0.696 | 0.964 | -0.010 | -0.023 | 0.004 | 0.180 | 0.450 | 0.005 | -0.005 | 0.016 | 0.321 | 0.616 | 0.007 | -0.020 | 0.035 | 0.606 | 0.769 |
| SM(d18:1/20:0) & SM(d16:1/22:0) | 0.000 | -0.022 | 0.022 | 0.989 | 0.996 | -0.003 | -0.017 | 0.011 | 0.675 | 0.833 | -0.005 | -0.016 | 0.005 | 0.322 | 0.616 | -0.026 | -0.053 | 0.001 | 0.063 | 0.184 |
| TG(48:3) [NL-16:1] | -0.021 | -0.043 | 0.001 | 0.066 | 0.903 | -0.014 | -0.028 | -0.001 | 0.041 | 0.306 | -0.005 | -0.016 | 0.005 | 0.324 | 0.620 | -0.017 | -0.045 | 0.010 | 0.214 | 0.422 |
| AC(24:0) | 0.008 | -0.014 | 0.030 | 0.503 | 0.922 | 0.009 | -0.005 | 0.023 | 0.191 | 0.454 | 0.005 | -0.005 | 0.016 | 0.326 | 0.620 | 0.014 | -0.013 | 0.042 | 0.301 | 0.524 |

| | | | | | | | | | | | | | | | | | | | | |
|---|---|---|---|---|---|---|---|---|---|---|---|---|---|---|---|---|---|---|---|---|
| LPC(17:1) [sn2] (a) | -0.015 | -0.037 | 0.007 | 0.193 | 0.903 | -0.018 | -0.032 | -0.005 | 0.008 | 0.263 | -0.005 | -0.016 | 0.005 | 0.328 | 0.621 | -0.027 | -0.054 | 0.001 | 0.055 | 0.173 |
| TG(49:1) [NL-17:1] | -0.019 | -0.041 | 0.003 | 0.087 | 0.903 | -0.011 | -0.024 | 0.003 | 0.117 | 0.385 | 0.005 | -0.005 | 0.016 | 0.327 | 0.621 | 0.000 | -0.027 | 0.027 | 0.997 | 0.997 |
| AC(26:0) | -0.002 | -0.024 | 0.020 | 0.890 | 0.981 | 0.005 | -0.009 | 0.019 | 0.516 | 0.727 | 0.005 | -0.005 | 0.016 | 0.329 | 0.621 | 0.003 | -0.025 | 0.030 | 0.836 | 0.928 |
| PC(O-32:1) | -0.010 | -0.032 | 0.012 | 0.391 | 0.922 | -0.006 | -0.020 | 0.008 | 0.373 | 0.608 | 0.005 | -0.005 | 0.016 | 0.330 | 0.623 | -0.022 | -0.050 | 0.005 | 0.116 | 0.289 |
| COH | -0.005 | -0.027 | 0.017 | 0.644 | 0.960 | -0.003 | -0.017 | 0.011 | 0.666 | 0.829 | -0.005 | -0.016 | 0.006 | 0.332 | 0.625 | -0.040 | -0.067 | -0.013 | 0.004 | 0.038 |
| dhCer(d18:0/18:0) | -0.026 | -0.048 | -0.004 | 0.022 | 0.903 | 0.002 | -0.011 | 0.016 | 0.735 | 0.870 | 0.005 | -0.005 | 0.016 | 0.338 | 0.634 | -0.036 | -0.063 | -0.009 | 0.009 | 0.054 |
| Cer(d18:2/24:0) | 0.001 | -0.021 | 0.023 | 0.944 | 0.995 | 0.001 | -0.013 | 0.016 | 0.864 | 0.930 | -0.005 | -0.016 | 0.006 | 0.339 | 0.636 | -0.008 | -0.036 | 0.020 | 0.565 | 0.742 |
| DG(18:1_18:3) | -0.011 | -0.033 | 0.011 | 0.343 | 0.922 | -0.009 | -0.023 | 0.005 | 0.194 | 0.454 | -0.005 | -0.016 | 0.006 | 0.343 | 0.641 | -0.013 | -0.040 | 0.015 | 0.372 | 0.590 |
| PC(P-18:0/18:2) | -0.006 | -0.028 | 0.016 | 0.572 | 0.933 | -0.004 | -0.018 | 0.010 | 0.552 | 0.748 | -0.005 | -0.016 | 0.006 | 0.344 | 0.642 | -0.021 | -0.047 | 0.006 | 0.133 | 0.318 |
| TG(54:6) [NL-20:5] | 0.000 | -0.023 | 0.022 | 0.976 | 0.995 | -0.013 | -0.027 | 0.001 | 0.066 | 0.338 | 0.005 | -0.006 | 0.016 | 0.346 | 0.643 | 0.002 | -0.026 | 0.030 | 0.901 | 0.952 |
| DG(16:0_16:0) | -0.014 | -0.036 | 0.008 | 0.211 | 0.903 | -0.012 | -0.026 | 0.001 | 0.082 | 0.356 | 0.005 | -0.006 | 0.016 | 0.350 | 0.648 | 0.014 | -0.014 | 0.041 | 0.324 | 0.541 |
| Cer(d18:1/14:0) | -0.001 | -0.023 | 0.021 | 0.933 | 0.995 | -0.005 | -0.020 | 0.009 | 0.464 | 0.686 | -0.005 | -0.016 | 0.006 | 0.350 | 0.648 | -0.029 | -0.057 | -0.001 | 0.046 | 0.155 |
| methyl-CE(18:2) | 0.013 | -0.009 | 0.035 | 0.249 | 0.903 | 0.003 | -0.011 | 0.017 | 0.694 | 0.845 | -0.005 | -0.016 | 0.006 | 0.353 | 0.649 | -0.011 | -0.039 | 0.016 | 0.421 | 0.634 |
| TG(50:4) [NL-18:3] | -0.022 | -0.044 | 0.000 | 0.047 | 0.903 | -0.012 | -0.026 | 0.001 | 0.076 | 0.356 | -0.005 | -0.016 | 0.006 | 0.352 | 0.649 | -0.011 | -0.039 | 0.017 | 0.429 | 0.638 |
| TG(54:3) [NL-18:2] | -0.006 | -0.027 | 0.016 | 0.602 | 0.950 | -0.011 | -0.025 | 0.003 | 0.117 | 0.385 | -0.005 | -0.016 | 0.006 | 0.354 | 0.649 | 0.001 | -0.027 | 0.029 | 0.945 | 0.966 |
| FA(16:1) | 0.003 | -0.019 | 0.025 | 0.777 | 0.973 | -0.002 | -0.016 | 0.012 | 0.764 | 0.881 | -0.005 | -0.016 | 0.006 | 0.355 | 0.650 | -0.014 | -0.042 | 0.014 | 0.316 | 0.537 |
| PC(38:2) | -0.009 | -0.031 | 0.013 | 0.423 | 0.922 | -0.005 | -0.019 | 0.009 | 0.478 | 0.696 | -0.005 | -0.016 | 0.006 | 0.358 | 0.654 | -0.029 | -0.056 | -0.002 | 0.033 | 0.127 |
| Cer(m18:0/24:0) | -0.014 | -0.036 | 0.008 | 0.204 | 0.903 | -0.003 | -0.017 | 0.011 | 0.719 | 0.856 | 0.005 | -0.006 | 0.016 | 0.360 | 0.656 | 0.004 | -0.024 | 0.031 | 0.791 | 0.904 |
| PC(16:0_20:5) | 0.007 | -0.015 | 0.029 | 0.521 | 0.922 | -0.012 | -0.025 | 0.002 | 0.105 | 0.373 | 0.005 | -0.006 | 0.016 | 0.361 | 0.656 | -0.011 | -0.039 | 0.017 | 0.440 | 0.644 |
| PC(17:0_18:2) | -0.008 | -0.030 | 0.013 | 0.455 | 0.922 | -0.011 | -0.025 | 0.003 | 0.122 | 0.385 | -0.005 | -0.016 | 0.006 | 0.364 | 0.660 | -0.034 | -0.061 | -0.007 | 0.014 | 0.074 |
| AC(17:0) (b) | -0.007 | -0.029 | 0.016 | 0.555 | 0.933 | -0.015 | -0.028 | -0.001 | 0.036 | 0.301 | -0.005 | -0.016 | 0.006 | 0.365 | 0.660 | -0.034 | -0.061 | -0.008 | 0.012 | 0.068 |
| PE(P-18:1/20:4) (b) | -0.006 | -0.028 | 0.016 | 0.583 | 0.940 | 0.002 | -0.012 | 0.016 | 0.755 | 0.879 | -0.005 | -0.016 | 0.006 | 0.366 | 0.661 | -0.031 | -0.058 | -0.005 | 0.022 | 0.097 |
| TG(48:1) [NL-16:1] | -0.021 | -0.043 | 0.001 | 0.058 | 0.903 | -0.009 | -0.023 | 0.004 | 0.186 | 0.453 | 0.005 | -0.006 | 0.016 | 0.368 | 0.661 | 0.004 | -0.023 | 0.032 | 0.768 | 0.892 |
| PE(P-18:1/20:3) (a) | -0.009 | -0.031 | 0.013 | 0.432 | 0.922 | -0.003 | -0.017 | 0.011 | 0.634 | 0.803 | 0.005 | -0.006 | 0.016 | 0.368 | 0.661 | -0.019 | -0.045 | 0.008 | 0.175 | 0.378 |
| SM(41:1) (a) | 0.001 | -0.021 | 0.023 | 0.911 | 0.987 | -0.004 | -0.018 | 0.010 | 0.558 | 0.748 | 0.005 | -0.006 | 0.016 | 0.369 | 0.661 | -0.017 | -0.044 | 0.010 | 0.211 | 0.420 |
| PC(38:5) (b) | 0.002 | -0.021 | 0.024 | 0.892 | 0.981 | -0.012 | -0.026 | 0.002 | 0.091 | 0.356 | -0.005 | -0.016 | 0.006 | 0.370 | 0.661 | -0.047 | -0.074 | -0.020 | 0.001 | 0.014 |
| AC(15:0) (a) | 0.008 | -0.014 | 0.030 | 0.473 | 0.922 | -0.015 | -0.029 | -0.001 | 0.031 | 0.291 | 0.005 | -0.006 | 0.016 | 0.371 | 0.662 | -0.004 | -0.032 | 0.023 | 0.749 | 0.882 |
| LPC(22:1) [sn1] | 0.011 | -0.011 | 0.033 | 0.331 | 0.922 | -0.011 | -0.024 | 0.003 | 0.136 | 0.399 | -0.005 | -0.016 | 0.006 | 0.374 | 0.665 | -0.009 | -0.036 | 0.019 | 0.543 | 0.726 |
| Cer(m18:1/18:0) | -0.022 | -0.044 | -0.001 | 0.045 | 0.903 | -0.009 | -0.023 | 0.004 | 0.180 | 0.450 | 0.005 | -0.006 | 0.016 | 0.375 | 0.666 | -0.016 | -0.043 | 0.011 | 0.251 | 0.465 |
| PC(31:0) (a) | -0.017 | -0.040 | 0.005 | 0.129 | 0.903 | -0.009 | -0.022 | 0.005 | 0.212 | 0.475 | 0.005 | -0.006 | 0.016 | 0.379 | 0.671 | -0.020 | -0.048 | 0.007 | 0.145 | 0.335 |
| PE(P-18:0/22:5) (n3) | -0.009 | -0.031 | 0.013 | 0.427 | 0.922 | 0.004 | -0.010 | 0.019 | 0.540 | 0.743 | 0.005 | -0.006 | 0.016 | 0.380 | 0.671 | -0.030 | -0.057 | -0.002 | 0.035 | 0.132 |
| SM(44:1) | 0.012 | -0.011 | 0.034 | 0.303 | 0.903 | -0.005 | -0.019 | 0.009 | 0.486 | 0.704 | 0.005 | -0.006 | 0.016 | 0.391 | 0.680 | -0.024 | -0.051 | 0.004 | 0.093 | 0.244 |
| Cer(m18:0/23:0) | -0.015 | -0.037 | 0.008 | 0.198 | 0.903 | -0.006 | -0.020 | 0.008 | 0.399 | 0.638 | 0.005 | -0.006 | 0.015 | 0.393 | 0.680 | -0.011 | -0.039 | 0.016 | 0.427 | 0.638 |
| DG(18:1_18:2) | -0.008 | -0.030 | 0.014 | 0.478 | 0.922 | -0.010 | -0.024 | 0.003 | 0.147 | 0.408 | -0.005 | -0.015 | 0.006 | 0.388 | 0.680 | -0.017 | -0.045 | 0.010 | 0.222 | 0.430 |
| PE(O-18:1/22:6) | 0.009 | -0.013 | 0.031 | 0.415 | 0.922 | -0.001 | -0.015 | 0.013 | 0.864 | 0.930 | 0.005 | -0.006 | 0.015 | 0.393 | 0.680 | -0.004 | -0.032 | 0.023 | 0.762 | 0.891 |
| CE(15:0) | 0.007 | -0.015 | 0.029 | 0.534 | 0.923 | -0.006 | -0.019 | 0.008 | 0.413 | 0.646 | -0.005 | -0.015 | 0.006 | 0.388 | 0.680 | -0.037 | -0.065 | -0.009 | 0.009 | 0.055 |
| PE(P-18:0/20:3) (b) | -0.006 | -0.028 | 0.016 | 0.577 | 0.933 | 0.000 | -0.014 | 0.015 | 0.981 | 0.989 | 0.005 | -0.006 | 0.016 | 0.392 | 0.680 | -0.018 | -0.045 | 0.010 | 0.204 | 0.416 |
| PC(P-16:0/18:1) | -0.004 | -0.026 | 0.018 | 0.708 | 0.964 | -0.008 | -0.022 | 0.006 | 0.248 | 0.501 | -0.005 | -0.016 | 0.006 | 0.391 | 0.680 | -0.032 | -0.059 | -0.005 | 0.022 | 0.098 |
| TG(56:7) [NL-20:4] | -0.004 | -0.026 | 0.018 | 0.723 | 0.964 | 0.005 | -0.009 | 0.019 | 0.520 | 0.727 | -0.005 | -0.015 | 0.006 | 0.392 | 0.680 | -0.003 | -0.031 | 0.025 | 0.860 | 0.937 |
| CE(22:0) | -0.003 | -0.025 | 0.020 | 0.818 | 0.980 | 0.010 | -0.004 | 0.023 | 0.162 | 0.432 | -0.005 | -0.016 | 0.006 | 0.392 | 0.680 | -0.009 | -0.035 | 0.018 | 0.507 | 0.702 |
| Cer(d18:1/23:0) | -0.012 | -0.034 | 0.010 | 0.275 | 0.903 | -0.007 | -0.021 | 0.007 | 0.353 | 0.592 | -0.005 | -0.015 | 0.006 | 0.395 | 0.680 | -0.023 | -0.050 | 0.004 | 0.094 | 0.246 |
| DE(20:4) | 0.016 | -0.006 | 0.038 | 0.150 | 0.903 | 0.019 | 0.005 | 0.032 | 0.007 | 0.263 | -0.005 | -0.015 | 0.006 | 0.395 | 0.680 | 0.001 | -0.027 | 0.029 | 0.945 | 0.966 |
| PI(15-MHDA_18:2) & PI(17:0_18:2) | -0.014 | -0.036 | 0.008 | 0.216 | 0.903 | 0.004 | -0.010 | 0.018 | 0.599 | 0.771 | -0.005 | -0.015 | 0.006 | 0.398 | 0.684 | -0.021 | -0.047 | 0.006 | 0.130 | 0.315 |
| Cer(d19:1/18:0) | -0.006 | -0.028 | 0.016 | 0.568 | 0.933 | -0.009 | -0.023 | 0.005 | 0.211 | 0.475 | -0.005 | -0.015 | 0.006 | 0.400 | 0.685 | -0.043 | -0.070 | -0.016 | 0.002 | 0.024 |
| DE(20:5) | 0.013 | -0.010 | 0.035 | 0.268 | 0.903 | -0.004 | -0.018 | 0.010 | 0.555 | 0.748 | 0.005 | -0.006 | 0.015 | 0.402 | 0.687 | -0.008 | -0.035 | 0.020 | 0.590 | 0.756 |
| PE(38:5) (b) | -0.010 | -0.032 | 0.013 | 0.399 | 0.922 | -0.016 | -0.030 | -0.003 | 0.019 | 0.273 | -0.005 | -0.015 | 0.006 | 0.402 | 0.687 | -0.024 | -0.051 | 0.004 | 0.088 | 0.236 |
| TG(54:5) [NL-20:4] | -0.015 | -0.037 | 0.007 | 0.178 | 0.903 | -0.007 | -0.021 | 0.006 | 0.285 | 0.530 | 0.004 | -0.006 | 0.015 | 0.417 | 0.708 | -0.001 | -0.029 | 0.027 | 0.932 | 0.964 |
| DG(18:1_20:3) | -0.011 | -0.033 | 0.011 | 0.324 | 0.922 | -0.011 | -0.025 | 0.002 | 0.106 | 0.373 | 0.004 | -0.006 | 0.015 | 0.417 | 0.708 | -0.014 | -0.042 | 0.015 | 0.346 | 0.564 |
| PG(36:1) | -0.020 | -0.042 | 0.002 | 0.069 | 0.903 | -0.014 | -0.028 | -0.001 | 0.038 | 0.301 | 0.004 | -0.006 | 0.015 | 0.418 | 0.709 | 0.010 | -0.017 | 0.038 | 0.466 | 0.664 |
| DG(14:0_18:2) | -0.017 | -0.040 | 0.005 | 0.121 | 0.903 | -0.009 | -0.023 | 0.004 | 0.176 | 0.446 | -0.004 | -0.015 | 0.006 | 0.420 | 0.709 | -0.014 | -0.042 | 0.014 | 0.315 | 0.537 |

| | | | | | | | | | | | | | | | | | | | | |
|---|---|---|---|---|---|---|---|---|---|---|---|---|---|---|---|---|---|---|---|---|
| FA(20:2) | -0.009 | -0.031 | 0.014 | 0.449 | 0.922 | 0.000 | -0.014 | 0.014 | 0.982 | 0.989 | -0.005 | -0.016 | 0.007 | 0.421 | 0.709 | -0.010 | -0.038 | 0.017 | 0.462 | 0.664 |
| PC(P-16:0/18:3) | -0.009 | -0.031 | 0.013 | 0.423 | 0.922 | -0.005 | -0.019 | 0.009 | 0.448 | 0.673 | -0.004 | -0.015 | 0.006 | 0.423 | 0.709 | -0.008 | -0.035 | 0.020 | 0.583 | 0.753 |
| CE(20:1) | 0.005 | -0.017 | 0.027 | 0.649 | 0.960 | -0.001 | -0.015 | 0.013 | 0.860 | 0.929 | -0.004 | -0.015 | 0.006 | 0.422 | 0.709 | -0.026 | -0.054 | 0.002 | 0.066 | 0.189 |
| PC(34:5) | 0.001 | -0.022 | 0.023 | 0.951 | 0.995 | -0.006 | -0.020 | 0.008 | 0.406 | 0.643 | 0.004 | -0.006 | 0.015 | 0.422 | 0.709 | -0.004 | -0.032 | 0.024 | 0.770 | 0.892 |
| AC(24:1)-OH | 0.015 | -0.007 | 0.037 | 0.172 | 0.903 | -0.012 | -0.026 | 0.001 | 0.071 | 0.350 | -0.004 | -0.015 | 0.006 | 0.424 | 0.710 | 0.007 | -0.020 | 0.034 | 0.613 | 0.775 |
| TG(O-54:3) [NL-17:1] | -0.007 | -0.029 | 0.014 | 0.501 | 0.922 | -0.005 | -0.019 | 0.009 | 0.471 | 0.689 | 0.004 | -0.006 | 0.015 | 0.426 | 0.711 | 0.003 | -0.025 | 0.031 | 0.836 | 0.928 |
| CE(24:0) | -0.001 | -0.023 | 0.022 | 0.944 | 0.995 | 0.011 | -0.003 | 0.024 | 0.127 | 0.385 | -0.004 | -0.015 | 0.006 | 0.427 | 0.711 | -0.010 | -0.037 | 0.017 | 0.463 | 0.664 |
| PC(16:0_20:3) (b) | -0.016 | -0.038 | 0.007 | 0.168 | 0.903 | -0.011 | -0.025 | 0.002 | 0.101 | 0.368 | -0.004 | -0.015 | 0.006 | 0.429 | 0.712 | -0.044 | -0.071 | -0.017 | 0.002 | 0.022 |
| Ubiquinone | 0.007 | -0.015 | 0.029 | 0.511 | 0.922 | -0.004 | -0.018 | 0.010 | 0.577 | 0.759 | -0.004 | -0.015 | 0.006 | 0.431 | 0.713 | -0.010 | -0.038 | 0.017 | 0.464 | 0.664 |
| LPI(18:0) [sn1] | -0.010 | -0.032 | 0.012 | 0.360 | 0.922 | -0.002 | -0.017 | 0.013 | 0.812 | 0.902 | 0.004 | -0.007 | 0.015 | 0.431 | 0.713 | -0.001 | -0.029 | 0.027 | 0.941 | 0.966 |
| PC(16:0_16:0) | -0.014 | -0.036 | 0.008 | 0.222 | 0.903 | -0.009 | -0.023 | 0.005 | 0.203 | 0.464 | -0.004 | -0.015 | 0.007 | 0.435 | 0.715 | -0.029 | -0.056 | -0.002 | 0.035 | 0.133 |
| LPE(P-16:0) | -0.009 | -0.031 | 0.013 | 0.426 | 0.922 | -0.006 | -0.020 | 0.009 | 0.438 | 0.667 | -0.004 | -0.015 | 0.006 | 0.434 | 0.715 | -0.016 | -0.044 | 0.012 | 0.275 | 0.499 |
| PE(18:0_18:1) | -0.007 | -0.029 | 0.015 | 0.548 | 0.929 | -0.012 | -0.025 | 0.002 | 0.095 | 0.360 | 0.004 | -0.006 | 0.015 | 0.434 | 0.715 | 0.015 | -0.012 | 0.043 | 0.280 | 0.505 |
| TG(50:1) [NL-18:1] | -0.022 | -0.043 | 0.000 | 0.052 | 0.903 | -0.011 | -0.025 | 0.003 | 0.122 | 0.385 | 0.004 | -0.006 | 0.015 | 0.438 | 0.719 | -0.002 | -0.030 | 0.026 | 0.884 | 0.948 |
| PC(17:0_18:1) | -0.015 | -0.036 | 0.007 | 0.193 | 0.903 | -0.018 | -0.032 | -0.004 | 0.010 | 0.270 | -0.004 | -0.015 | 0.007 | 0.441 | 0.720 | -0.048 | -0.075 | -0.021 | 0.001 | 0.012 |
| PE(16:0_18:2) | -0.017 | -0.038 | 0.005 | 0.133 | 0.903 | -0.012 | -0.025 | 0.002 | 0.104 | 0.372 | -0.004 | -0.015 | 0.007 | 0.441 | 0.720 | -0.011 | -0.039 | 0.016 | 0.425 | 0.636 |
| PE(P-18:1/22:5) (b) | -0.004 | -0.026 | 0.018 | 0.722 | 0.964 | 0.004 | -0.010 | 0.018 | 0.549 | 0.747 | -0.004 | -0.015 | 0.007 | 0.444 | 0.723 | -0.005 | -0.032 | 0.021 | 0.698 | 0.842 |
| SM(d18:0/16:0) | 0.007 | -0.015 | 0.029 | 0.547 | 0.929 | -0.002 | -0.016 | 0.012 | 0.810 | 0.902 | -0.004 | -0.015 | 0.007 | 0.445 | 0.723 | -0.028 | -0.055 | -0.001 | 0.042 | 0.147 |
| SM(d18:1/24:0) | 0.003 | -0.019 | 0.025 | 0.805 | 0.980 | 0.006 | -0.008 | 0.020 | 0.418 | 0.649 | 0.004 | -0.007 | 0.015 | 0.446 | 0.723 | -0.008 | -0.035 | 0.019 | 0.575 | 0.746 |
| Cer(m18:0/22:0) | -0.014 | -0.036 | 0.008 | 0.225 | 0.903 | -0.003 | -0.017 | 0.011 | 0.640 | 0.807 | 0.004 | -0.007 | 0.015 | 0.450 | 0.727 | -0.007 | -0.034 | 0.021 | 0.629 | 0.793 |
| PC(O-32:2) | -0.013 | -0.035 | 0.009 | 0.253 | 0.903 | -0.006 | -0.020 | 0.008 | 0.423 | 0.655 | 0.004 | -0.007 | 0.015 | 0.450 | 0.727 | -0.003 | -0.031 | 0.024 | 0.802 | 0.910 |
| PC(O-16:0/20:4) | -0.007 | -0.029 | 0.014 | 0.512 | 0.922 | 0.003 | -0.011 | 0.017 | 0.633 | 0.803 | 0.004 | -0.007 | 0.015 | 0.452 | 0.727 | -0.016 | -0.043 | 0.012 | 0.262 | 0.482 |
| Cer(d18:2/26:0) | 0.004 | -0.019 | 0.026 | 0.745 | 0.964 | 0.002 | -0.012 | 0.016 | 0.804 | 0.898 | -0.004 | -0.015 | 0.007 | 0.452 | 0.727 | -0.022 | -0.050 | 0.005 | 0.112 | 0.278 |
| Cer(d16:1/18:0) | -0.012 | -0.034 | 0.009 | 0.267 | 0.903 | -0.014 | -0.027 | 0.000 | 0.052 | 0.317 | -0.004 | -0.015 | 0.007 | 0.455 | 0.727 | -0.033 | -0.061 | -0.006 | 0.018 | 0.087 |
| AC(18:0)-OH | 0.000 | -0.022 | 0.022 | 0.989 | 0.996 | -0.008 | -0.021 | 0.006 | 0.278 | 0.524 | -0.004 | -0.015 | 0.007 | 0.454 | 0.727 | -0.032 | -0.060 | -0.005 | 0.021 | 0.093 |
| TG(50:1) [NL-16:0] | -0.019 | -0.041 | 0.002 | 0.082 | 0.903 | -0.009 | -0.023 | 0.004 | 0.181 | 0.450 | 0.004 | -0.007 | 0.015 | 0.458 | 0.731 | 0.001 | -0.027 | 0.029 | 0.964 | 0.973 |
| AC(26:1) | 0.001 | -0.021 | 0.023 | 0.949 | 0.995 | -0.001 | -0.015 | 0.013 | 0.857 | 0.929 | -0.004 | -0.015 | 0.007 | 0.460 | 0.733 | -0.016 | -0.043 | 0.011 | 0.237 | 0.450 |
| PC(18:0_20:3) | -0.010 | -0.032 | 0.011 | 0.347 | 0.922 | -0.006 | -0.019 | 0.008 | 0.418 | 0.649 | 0.004 | -0.007 | 0.015 | 0.462 | 0.734 | -0.020 | -0.047 | 0.007 | 0.145 | 0.335 |
| Cer(d17:1/22:0) | -0.007 | -0.029 | 0.015 | 0.524 | 0.922 | -0.002 | -0.016 | 0.012 | 0.753 | 0.879 | -0.004 | -0.015 | 0.007 | 0.468 | 0.741 | -0.016 | -0.044 | 0.011 | 0.245 | 0.458 |
| LPC(20:4) [sn1] | -0.005 | -0.027 | 0.017 | 0.645 | 0.960 | -0.002 | -0.016 | 0.012 | 0.730 | 0.868 | -0.004 | -0.015 | 0.007 | 0.471 | 0.741 | -0.014 | -0.042 | 0.014 | 0.323 | 0.541 |
| SM(d18:2/16:0) | -0.002 | -0.024 | 0.020 | 0.871 | 0.981 | 0.005 | -0.009 | 0.019 | 0.462 | 0.686 | -0.004 | -0.015 | 0.007 | 0.470 | 0.741 | -0.027 | -0.053 | 0.000 | 0.053 | 0.171 |
| PC(18:0_22:6) | 0.001 | -0.021 | 0.024 | 0.895 | 0.983 | -0.008 | -0.022 | 0.006 | 0.251 | 0.503 | 0.004 | -0.007 | 0.015 | 0.469 | 0.741 | -0.025 | -0.053 | 0.003 | 0.083 | 0.224 |
| PE(17:0_22:6) | 0.001 | -0.021 | 0.023 | 0.920 | 0.990 | -0.014 | -0.028 | -0.001 | 0.038 | 0.302 | 0.004 | -0.007 | 0.015 | 0.467 | 0.741 | -0.004 | -0.031 | 0.024 | 0.793 | 0.904 |
| PC(P-16:0/16:1) | -0.009 | -0.031 | 0.014 | 0.449 | 0.922 | -0.013 | -0.027 | 0.001 | 0.066 | 0.338 | -0.004 | -0.015 | 0.007 | 0.475 | 0.746 | -0.029 | -0.056 | -0.002 | 0.034 | 0.132 |
| CE(22:1) | 0.011 | -0.011 | 0.033 | 0.342 | 0.922 | 0.001 | -0.013 | 0.014 | 0.942 | 0.971 | 0.004 | -0.007 | 0.015 | 0.476 | 0.746 | -0.013 | -0.041 | 0.014 | 0.335 | 0.553 |
| PC(15:0_20:3) | -0.013 | -0.035 | 0.009 | 0.265 | 0.903 | -0.008 | -0.021 | 0.006 | 0.269 | 0.520 | -0.004 | -0.015 | 0.007 | 0.478 | 0.747 | -0.032 | -0.059 | -0.006 | 0.018 | 0.087 |
| Cer(d16:1/16:0) | -0.003 | -0.025 | 0.020 | 0.822 | 0.980 | -0.008 | -0.022 | 0.006 | 0.252 | 0.503 | -0.004 | -0.015 | 0.007 | 0.480 | 0.749 | -0.014 | -0.042 | 0.013 | 0.306 | 0.527 |
| PC(O-18:0/22:6) | -0.008 | -0.030 | 0.015 | 0.504 | 0.922 | -0.015 | -0.029 | -0.001 | 0.043 | 0.306 | -0.004 | -0.015 | 0.007 | 0.484 | 0.752 | -0.037 | -0.065 | -0.010 | 0.007 | 0.049 |
| PC(O-38:5) | 0.004 | -0.018 | 0.026 | 0.723 | 0.964 | 0.004 | -0.009 | 0.018 | 0.528 | 0.735 | -0.004 | -0.015 | 0.007 | 0.483 | 0.752 | -0.030 | -0.056 | -0.003 | 0.032 | 0.126 |
| LPC(P-17:0) (b) | -0.011 | -0.033 | 0.011 | 0.330 | 0.922 | -0.007 | -0.021 | 0.007 | 0.307 | 0.549 | -0.004 | -0.014 | 0.007 | 0.487 | 0.756 | -0.018 | -0.045 | 0.009 | 0.195 | 0.408 |
| TG(O-54:3) [NL-18:1] | 0.001 | -0.021 | 0.023 | 0.946 | 0.995 | -0.002 | -0.016 | 0.012 | 0.766 | 0.881 | 0.004 | -0.007 | 0.014 | 0.490 | 0.758 | 0.006 | -0.021 | 0.034 | 0.647 | 0.807 |
| TG(48:3) [NL-14:0] | -0.020 | -0.042 | 0.002 | 0.077 | 0.903 | -0.012 | -0.026 | 0.002 | 0.083 | 0.356 | -0.004 | -0.014 | 0.007 | 0.494 | 0.759 | -0.011 | -0.039 | 0.016 | 0.420 | 0.634 |
| LPC(24:0) [sn2] | 0.013 | -0.009 | 0.035 | 0.241 | 0.903 | 0.002 | -0.012 | 0.016 | 0.739 | 0.873 | -0.004 | -0.015 | 0.007 | 0.495 | 0.759 | 0.009 | -0.019 | 0.036 | 0.549 | 0.729 |
| LPC(22:1) [sn2] | 0.015 | -0.006 | 0.037 | 0.164 | 0.903 | -0.008 | -0.022 | 0.005 | 0.232 | 0.488 | -0.004 | -0.015 | 0.007 | 0.493 | 0.759 | -0.005 | -0.032 | 0.023 | 0.740 | 0.874 |
| SM(d18:2/14:0) | 0.001 | -0.021 | 0.023 | 0.914 | 0.987 | -0.001 | -0.015 | 0.013 | 0.899 | 0.942 | -0.004 | -0.015 | 0.007 | 0.495 | 0.759 | -0.036 | -0.063 | -0.009 | 0.008 | 0.053 |
| PE(15-MHDA_18:2) | -0.012 | -0.034 | 0.010 | 0.292 | 0.903 | -0.017 | -0.031 | -0.003 | 0.017 | 0.273 | -0.004 | -0.015 | 0.007 | 0.496 | 0.760 | -0.030 | -0.057 | -0.002 | 0.036 | 0.134 |
| LPC(20:5) [sn2] | 0.008 | -0.014 | 0.030 | 0.482 | 0.922 | -0.011 | -0.025 | 0.003 | 0.121 | 0.385 | 0.004 | -0.007 | 0.015 | 0.498 | 0.761 | -0.005 | -0.033 | 0.022 | 0.713 | 0.847 |
| Cer(d18:1/19:0) | -0.018 | -0.040 | 0.005 | 0.124 | 0.903 | -0.008 | -0.021 | 0.006 | 0.273 | 0.521 | -0.004 | -0.015 | 0.007 | 0.500 | 0.762 | -0.028 | -0.055 | 0.000 | 0.049 | 0.161 |
| PC(33:0) (b) | -0.012 | -0.034 | 0.010 | 0.285 | 0.903 | -0.009 | -0.022 | 0.005 | 0.231 | 0.488 | -0.004 | -0.014 | 0.007 | 0.505 | 0.767 | -0.047 | -0.074 | -0.019 | 0.001 | 0.016 |
| DG(16:0_18:2) | -0.014 | -0.036 | 0.008 | 0.205 | 0.903 | -0.008 | -0.022 | 0.005 | 0.230 | 0.488 | -0.004 | -0.014 | 0.007 | 0.510 | 0.767 | -0.009 | -0.036 | 0.019 | 0.534 | 0.719 |
| DG(18:1_20:4) | -0.016 | -0.038 | 0.006 | 0.155 | 0.903 | -0.006 | -0.020 | 0.008 | 0.386 | 0.626 | 0.004 | -0.007 | 0.014 | 0.509 | 0.767 | -0.003 | -0.031 | 0.025 | 0.835 | 0.928 |

| | | | | | | | | | | | | | | | | | | | | |
|---|---|---|---|---|---|---|---|---|---|---|---|---|---|---|---|---|---|---|---|---|
| PC(P-16:0/14:0) | -0.007 | -0.029 | 0.016 | 0.557 | 0.933 | -0.007 | -0.021 | 0.007 | 0.337 | 0.574 | 0.004 | -0.007 | 0.014 | 0.509 | 0.767 | -0.030 | -0.057 | -0.002 | 0.035 | 0.132 |
| SM(d18:2/24:0) | 0.007 | -0.015 | 0.029 | 0.554 | 0.933 | 0.004 | -0.010 | 0.018 | 0.555 | 0.748 | -0.004 | -0.015 | 0.007 | 0.508 | 0.767 | -0.019 | -0.046 | 0.008 | 0.161 | 0.360 |
| PC(36:4) [+OH] | -0.005 | -0.027 | 0.017 | 0.633 | 0.958 | 0.005 | -0.008 | 0.019 | 0.432 | 0.663 | 0.004 | -0.007 | 0.014 | 0.508 | 0.767 | -0.003 | -0.031 | 0.024 | 0.803 | 0.910 |
| methyl-CE(18:0) | -0.002 | -0.025 | 0.021 | 0.856 | 0.981 | 0.003 | -0.011 | 0.016 | 0.698 | 0.845 | -0.004 | -0.015 | 0.007 | 0.511 | 0.767 | -0.013 | -0.040 | 0.014 | 0.344 | 0.563 |
| SM(d16:1/19:0) | -0.001 | -0.024 | 0.021 | 0.928 | 0.995 | -0.009 | -0.023 | 0.005 | 0.190 | 0.454 | -0.004 | -0.014 | 0.007 | 0.508 | 0.767 | -0.044 | -0.071 | -0.016 | 0.002 | 0.024 |
| FA(22:5) | -0.003 | -0.025 | 0.019 | 0.806 | 0.980 | 0.002 | -0.011 | 0.016 | 0.742 | 0.874 | 0.004 | -0.007 | 0.015 | 0.512 | 0.767 | 0.004 | -0.024 | 0.031 | 0.795 | 0.905 |
| TG(54:6) [NL-20:4] | -0.014 | -0.036 | 0.008 | 0.217 | 0.903 | -0.002 | -0.016 | 0.012 | 0.778 | 0.884 | -0.004 | -0.014 | 0.007 | 0.513 | 0.768 | -0.006 | -0.034 | 0.022 | 0.675 | 0.823 |
| AC(22:6) | 0.010 | -0.012 | 0.032 | 0.376 | 0.922 | -0.009 | -0.023 | 0.005 | 0.212 | 0.475 | 0.004 | -0.007 | 0.014 | 0.514 | 0.768 | 0.001 | -0.027 | 0.028 | 0.954 | 0.969 |
| TG(48:1) [NL-18:1] | -0.019 | -0.041 | 0.003 | 0.083 | 0.903 | -0.012 | -0.026 | 0.001 | 0.081 | 0.356 | 0.004 | -0.007 | 0.014 | 0.518 | 0.771 | -0.003 | -0.031 | 0.025 | 0.847 | 0.933 |
| PC(P-18:0/22:6) | -0.002 | -0.025 | 0.020 | 0.830 | 0.981 | -0.010 | -0.024 | 0.004 | 0.171 | 0.438 | 0.004 | -0.007 | 0.014 | 0.519 | 0.771 | -0.015 | -0.042 | 0.012 | 0.281 | 0.505 |
| PE(P-20:0/20:4) | -0.001 | -0.023 | 0.021 | 0.939 | 0.995 | 0.001 | -0.013 | 0.015 | 0.877 | 0.934 | 0.003 | -0.007 | 0.014 | 0.528 | 0.782 | -0.022 | -0.049 | 0.006 | 0.131 | 0.315 |
| PC(P-38:5) (b) | 0.001 | -0.021 | 0.023 | 0.958 | 0.995 | -0.011 | -0.025 | 0.003 | 0.126 | 0.385 | -0.003 | -0.014 | 0.007 | 0.528 | 0.782 | -0.028 | -0.055 | -0.002 | 0.038 | 0.138 |
| PI(18:0_22:5) (n3) | -0.013 | -0.035 | 0.009 | 0.232 | 0.903 | -0.010 | -0.024 | 0.004 | 0.164 | 0.432 | -0.003 | -0.014 | 0.007 | 0.529 | 0.782 | -0.035 | -0.062 | -0.008 | 0.013 | 0.069 |
| Cer(m18:0/24:1) | -0.019 | -0.040 | 0.003 | 0.096 | 0.903 | -0.013 | -0.027 | 0.000 | 0.059 | 0.323 | 0.003 | -0.007 | 0.014 | 0.537 | 0.791 | -0.017 | -0.044 | 0.010 | 0.230 | 0.439 |
| PC(P-16:0/16:0) | -0.012 | -0.034 | 0.010 | 0.305 | 0.903 | -0.010 | -0.024 | 0.003 | 0.144 | 0.406 | -0.003 | -0.014 | 0.007 | 0.538 | 0.791 | -0.014 | -0.042 | 0.013 | 0.306 | 0.527 |
| Cer(d19:1/26:0) | 0.001 | -0.022 | 0.024 | 0.928 | 0.995 | -0.001 | -0.016 | 0.014 | 0.892 | 0.940 | 0.003 | -0.007 | 0.014 | 0.537 | 0.791 | -0.004 | -0.032 | 0.024 | 0.777 | 0.894 |
| TG(54:4) [NL-20:3] | -0.014 | -0.035 | 0.008 | 0.220 | 0.903 | -0.014 | -0.028 | 0.000 | 0.046 | 0.306 | 0.003 | -0.007 | 0.014 | 0.540 | 0.791 | -0.012 | -0.040 | 0.016 | 0.405 | 0.620 |
| PE(16:0_16:1) | -0.021 | -0.043 | 0.001 | 0.066 | 0.903 | -0.009 | -0.023 | 0.004 | 0.187 | 0.454 | 0.003 | -0.007 | 0.014 | 0.544 | 0.791 | -0.012 | -0.040 | 0.015 | 0.381 | 0.596 |
| PE(P-16:0/22:4) | -0.023 | -0.044 | -0.001 | 0.044 | 0.903 | 0.008 | -0.006 | 0.022 | 0.269 | 0.520 | 0.003 | -0.007 | 0.014 | 0.543 | 0.791 | -0.008 | -0.035 | 0.020 | 0.574 | 0.745 |
| AC(16:0)-OH | -0.007 | -0.029 | 0.015 | 0.520 | 0.922 | -0.009 | -0.022 | 0.005 | 0.213 | 0.475 | -0.003 | -0.014 | 0.007 | 0.543 | 0.791 | -0.033 | -0.060 | -0.006 | 0.017 | 0.084 |
| PE(O-16:0/18:2) | -0.009 | -0.031 | 0.014 | 0.451 | 0.922 | 0.003 | -0.011 | 0.017 | 0.644 | 0.809 | 0.003 | -0.007 | 0.014 | 0.541 | 0.791 | -0.006 | -0.033 | 0.021 | 0.659 | 0.814 |
| Hex2Cer(d16:1/24:1) | -0.003 | -0.025 | 0.020 | 0.814 | 0.980 | -0.004 | -0.018 | 0.009 | 0.530 | 0.735 | -0.003 | -0.014 | 0.007 | 0.545 | 0.792 | -0.006 | -0.034 | 0.021 | 0.658 | 0.814 |
| PC(18:0_22:5) (n6) | -0.010 | -0.032 | 0.012 | 0.386 | 0.922 | 0.002 | -0.012 | 0.016 | 0.787 | 0.889 | -0.003 | -0.014 | 0.007 | 0.547 | 0.792 | -0.014 | -0.041 | 0.013 | 0.298 | 0.522 |
| DG(18:1_22:5) | -0.006 | -0.028 | 0.016 | 0.611 | 0.950 | -0.016 | -0.029 | -0.002 | 0.025 | 0.285 | 0.003 | -0.007 | 0.014 | 0.548 | 0.792 | -0.014 | -0.042 | 0.014 | 0.315 | 0.537 |
| TG(56:6) [NL-22:5] | -0.008 | -0.029 | 0.014 | 0.501 | 0.922 | -0.019 | -0.033 | -0.006 | 0.005 | 0.259 | 0.003 | -0.007 | 0.014 | 0.550 | 0.794 | -0.019 | -0.047 | 0.009 | 0.178 | 0.383 |
| Cer(d16:1/22:0) | -0.005 | -0.027 | 0.017 | 0.666 | 0.964 | -0.007 | -0.022 | 0.007 | 0.295 | 0.539 | -0.003 | -0.014 | 0.008 | 0.552 | 0.795 | 0.005 | -0.022 | 0.033 | 0.702 | 0.845 |
| SM(44:2) | -0.002 | -0.024 | 0.020 | 0.872 | 0.981 | -0.012 | -0.026 | 0.002 | 0.087 | 0.356 | 0.003 | -0.007 | 0.014 | 0.555 | 0.797 | -0.031 | -0.058 | -0.004 | 0.025 | 0.106 |
| FA(22:4) | -0.004 | -0.026 | 0.018 | 0.718 | 0.964 | 0.004 | -0.010 | 0.018 | 0.545 | 0.745 | -0.003 | -0.014 | 0.008 | 0.558 | 0.801 | -0.012 | -0.040 | 0.015 | 0.381 | 0.596 |
| LPC(20:5) [sn1] | 0.008 | -0.015 | 0.030 | 0.497 | 0.922 | -0.012 | -0.026 | 0.002 | 0.102 | 0.368 | 0.003 | -0.008 | 0.014 | 0.565 | 0.810 | -0.004 | -0.032 | 0.024 | 0.777 | 0.894 |
| SM(43:2) (c) | 0.002 | -0.020 | 0.024 | 0.865 | 0.981 | -0.009 | -0.023 | 0.004 | 0.170 | 0.437 | 0.003 | -0.008 | 0.014 | 0.566 | 0.810 | -0.023 | -0.050 | 0.003 | 0.084 | 0.226 |
| PC(O-34:2) | -0.006 | -0.028 | 0.016 | 0.612 | 0.950 | 0.003 | -0.011 | 0.017 | 0.687 | 0.840 | -0.003 | -0.014 | 0.008 | 0.568 | 0.810 | -0.014 | -0.041 | 0.013 | 0.317 | 0.537 |
| TG(O-54:4) [NL-17:1] | -0.015 | -0.036 | 0.007 | 0.190 | 0.903 | -0.001 | -0.016 | 0.013 | 0.836 | 0.919 | 0.003 | -0.008 | 0.014 | 0.573 | 0.813 | 0.018 | -0.010 | 0.046 | 0.208 | 0.419 |
| PC(40:7) (a) | -0.010 | -0.032 | 0.012 | 0.386 | 0.922 | -0.005 | -0.018 | 0.009 | 0.521 | 0.727 | -0.003 | -0.014 | 0.008 | 0.574 | 0.813 | -0.022 | -0.050 | 0.005 | 0.109 | 0.276 |
| FA(22:6) | 0.003 | -0.019 | 0.025 | 0.816 | 0.980 | -0.006 | -0.020 | 0.007 | 0.361 | 0.599 | 0.003 | -0.008 | 0.014 | 0.573 | 0.813 | 0.003 | -0.025 | 0.031 | 0.837 | 0.928 |
| CA | -0.001 | -0.023 | 0.021 | 0.917 | 0.988 | 0.002 | -0.012 | 0.016 | 0.760 | 0.881 | 0.003 | -0.008 | 0.014 | 0.574 | 0.813 | 0.009 | -0.018 | 0.037 | 0.507 | 0.702 |
| TG(54:2) [NL-18:0] | -0.012 | -0.033 | 0.010 | 0.289 | 0.903 | -0.014 | -0.028 | 0.000 | 0.044 | 0.306 | 0.003 | -0.008 | 0.014 | 0.580 | 0.817 | -0.001 | -0.029 | 0.027 | 0.927 | 0.964 |
| CE(18:0) | -0.006 | -0.028 | 0.016 | 0.604 | 0.950 | -0.002 | -0.016 | 0.012 | 0.793 | 0.891 | -0.003 | -0.014 | 0.008 | 0.580 | 0.817 | -0.041 | -0.069 | -0.014 | 0.003 | 0.035 |
| CE(24:4) | 0.002 | -0.021 | 0.024 | 0.879 | 0.981 | 0.010 | -0.004 | 0.023 | 0.163 | 0.432 | -0.003 | -0.014 | 0.008 | 0.578 | 0.817 | -0.011 | -0.039 | 0.016 | 0.408 | 0.623 |
| LPC(22:6) [sn1] | 0.001 | -0.022 | 0.023 | 0.955 | 0.995 | -0.015 | -0.029 | -0.001 | 0.034 | 0.298 | -0.003 | -0.014 | 0.008 | 0.581 | 0.817 | -0.020 | -0.048 | 0.008 | 0.155 | 0.352 |
| PC(15:0_20:4) | -0.006 | -0.027 | 0.016 | 0.613 | 0.950 | 0.000 | -0.014 | 0.014 | 0.984 | 0.989 | 0.003 | -0.008 | 0.014 | 0.583 | 0.817 | -0.030 | -0.057 | -0.002 | 0.035 | 0.133 |
| TG(51:2) [NL-15:0] | -0.012 | -0.034 | 0.010 | 0.296 | 0.903 | -0.019 | -0.033 | -0.006 | 0.006 | 0.259 | -0.003 | -0.014 | 0.008 | 0.586 | 0.820 | -0.028 | -0.056 | 0.000 | 0.053 | 0.170 |
| PE(36:0) | -0.016 | -0.038 | 0.005 | 0.140 | 0.903 | -0.009 | -0.023 | 0.004 | 0.185 | 0.453 | -0.003 | -0.014 | 0.008 | 0.588 | 0.820 | -0.003 | -0.031 | 0.025 | 0.840 | 0.928 |
| PC(P-18:0/20:4) | -0.009 | -0.031 | 0.012 | 0.398 | 0.922 | 0.002 | -0.012 | 0.016 | 0.745 | 0.875 | 0.003 | -0.008 | 0.014 | 0.588 | 0.820 | -0.014 | -0.041 | 0.013 | 0.320 | 0.539 |
| PI(18:0_18:1) | -0.008 | -0.030 | 0.014 | 0.456 | 0.922 | -0.012 | -0.027 | 0.002 | 0.083 | 0.356 | -0.003 | -0.014 | 0.008 | 0.591 | 0.821 | -0.020 | -0.047 | 0.007 | 0.147 | 0.337 |
| TG(53:2) [NL-18:1] | -0.011 | -0.033 | 0.011 | 0.339 | 0.922 | -0.020 | -0.033 | -0.006 | 0.004 | 0.259 | 0.003 | -0.008 | 0.014 | 0.592 | 0.821 | -0.014 | -0.042 | 0.014 | 0.317 | 0.537 |
| SM(d18:1/17:0) & SM(d17:1/18:0) | -0.002 | -0.024 | 0.021 | 0.882 | 0.981 | -0.005 | -0.019 | 0.009 | 0.510 | 0.724 | -0.003 | -0.014 | 0.008 | 0.592 | 0.821 | -0.048 | -0.075 | -0.021 | 0.001 | 0.013 |
| LPC(20:4) [sn2] | -0.005 | -0.027 | 0.017 | 0.659 | 0.960 | -0.002 | -0.016 | 0.012 | 0.827 | 0.913 | -0.003 | -0.014 | 0.008 | 0.595 | 0.823 | -0.015 | -0.043 | 0.013 | 0.285 | 0.509 |
| PC(33:0) (a) | -0.015 | -0.037 | 0.007 | 0.184 | 0.903 | -0.012 | -0.026 | 0.002 | 0.092 | 0.356 | 0.003 | -0.008 | 0.014 | 0.599 | 0.825 | -0.034 | -0.061 | -0.006 | 0.016 | 0.080 |
| Cer(m18:1/24:1) | -0.018 | -0.040 | 0.004 | 0.113 | 0.903 | -0.012 | -0.026 | 0.002 | 0.082 | 0.356 | 0.003 | -0.008 | 0.014 | 0.599 | 0.825 | -0.004 | -0.031 | 0.023 | 0.774 | 0.893 |
| SHexCer(d18:1/24:0) | 0.002 | -0.020 | 0.025 | 0.839 | 0.981 | -0.015 | -0.028 | -0.001 | 0.041 | 0.306 | 0.003 | -0.008 | 0.014 | 0.600 | 0.825 | -0.009 | -0.037 | 0.020 | 0.548 | 0.729 |

| | | | | | | | | | | | | | | | | | | | | |
|---|---|---|---|---|---|---|---|---|---|---|---|---|---|---|---|---|---|---|---|---|
| AC(14:1)-OH | 0.001 | -0.021 | 0.023 | 0.946 | 0.995 | -0.002 | -0.015 | 0.011 | 0.770 | 0.881 | -0.003 | -0.014 | 0.008 | 0.600 | 0.825 | -0.024 | -0.052 | 0.004 | 0.091 | 0.241 |
| TG(56:8) [NL-20:4] | -0.012 | -0.034 | 0.010 | 0.295 | 0.903 | 0.006 | -0.008 | 0.020 | 0.425 | 0.657 | -0.003 | -0.014 | 0.008 | 0.604 | 0.828 | 0.009 | -0.019 | 0.037 | 0.530 | 0.718 |
| LPE(P-18:0) | -0.008 | -0.030 | 0.014 | 0.490 | 0.922 | -0.004 | -0.018 | 0.010 | 0.590 | 0.764 | -0.003 | -0.014 | 0.008 | 0.603 | 0.828 | -0.020 | -0.047 | 0.008 | 0.159 | 0.358 |
| PC(32:2) | -0.019 | -0.041 | 0.003 | 0.092 | 0.903 | -0.004 | -0.018 | 0.010 | 0.599 | 0.771 | -0.003 | -0.014 | 0.008 | 0.617 | 0.830 | -0.013 | -0.040 | 0.014 | 0.358 | 0.577 |
| DG(16:0_20:4) | -0.020 | -0.042 | 0.002 | 0.070 | 0.903 | -0.006 | -0.019 | 0.008 | 0.404 | 0.643 | 0.003 | -0.008 | 0.013 | 0.618 | 0.830 | 0.003 | -0.024 | 0.031 | 0.818 | 0.920 |
| PA(34:1) | -0.012 | -0.034 | 0.010 | 0.299 | 0.903 | -0.020 | -0.034 | -0.006 | 0.005 | 0.259 | -0.003 | -0.014 | 0.008 | 0.619 | 0.830 | -0.002 | -0.029 | 0.026 | 0.897 | 0.952 |
| PC(16:0_18:2) | -0.007 | -0.029 | 0.015 | 0.520 | 0.922 | -0.012 | -0.026 | 0.002 | 0.104 | 0.372 | -0.003 | -0.014 | 0.008 | 0.619 | 0.830 | -0.034 | -0.061 | -0.006 | 0.016 | 0.080 |
| AC(20:3)-OH | -0.007 | -0.029 | 0.015 | 0.523 | 0.922 | 0.004 | -0.010 | 0.018 | 0.583 | 0.763 | -0.003 | -0.014 | 0.008 | 0.614 | 0.830 | -0.017 | -0.043 | 0.010 | 0.219 | 0.429 |
| Cer(d19:1/24:0) | 0.008 | -0.014 | 0.031 | 0.465 | 0.922 | -0.001 | -0.015 | 0.013 | 0.870 | 0.932 | 0.003 | -0.008 | 0.014 | 0.613 | 0.830 | -0.010 | -0.037 | 0.018 | 0.483 | 0.676 |
| PE(P-18:0/22:5) (n6) | -0.007 | -0.029 | 0.015 | 0.515 | 0.922 | 0.007 | -0.007 | 0.021 | 0.336 | 0.574 | 0.003 | -0.008 | 0.013 | 0.610 | 0.830 | 0.009 | -0.018 | 0.035 | 0.520 | 0.709 |
| TG(54:7) [NL-20:5] | -0.007 | -0.030 | 0.015 | 0.523 | 0.922 | -0.009 | -0.023 | 0.004 | 0.185 | 0.453 | 0.003 | -0.008 | 0.014 | 0.614 | 0.830 | 0.005 | -0.022 | 0.033 | 0.706 | 0.845 |
| LPC(19:0) [sn2] (a) | 0.005 | -0.017 | 0.027 | 0.651 | 0.960 | -0.007 | -0.021 | 0.007 | 0.316 | 0.557 | -0.003 | -0.014 | 0.008 | 0.610 | 0.830 | -0.008 | -0.035 | 0.020 | 0.588 | 0.756 |
| PC(17:0_22:6) | 0.005 | -0.018 | 0.027 | 0.690 | 0.964 | -0.014 | -0.029 | 0.000 | 0.046 | 0.306 | -0.003 | -0.014 | 0.008 | 0.609 | 0.830 | -0.039 | -0.067 | -0.012 | 0.005 | 0.047 |
| SM(d19:1/24:1) | 0.005 | -0.018 | 0.027 | 0.676 | 0.964 | -0.012 | -0.026 | 0.002 | 0.097 | 0.361 | 0.003 | -0.008 | 0.013 | 0.616 | 0.830 | -0.026 | -0.054 | 0.001 | 0.063 | 0.184 |
| PC(15-MHDA_22:6) | 0.002 | -0.020 | 0.025 | 0.845 | 0.981 | -0.015 | -0.029 | -0.001 | 0.037 | 0.301 | 0.003 | -0.008 | 0.014 | 0.620 | 0.830 | -0.037 | -0.065 | -0.009 | 0.010 | 0.059 |
| SM(37:1) | 0.000 | -0.023 | 0.022 | 0.975 | 0.995 | -0.006 | -0.020 | 0.008 | 0.411 | 0.646 | -0.003 | -0.013 | 0.008 | 0.612 | 0.830 | -0.042 | -0.069 | -0.014 | 0.003 | 0.035 |
| LPC(14:0) [sn2] | -0.012 | -0.034 | 0.011 | 0.311 | 0.910 | -0.010 | -0.023 | 0.004 | 0.167 | 0.433 | -0.003 | -0.013 | 0.008 | 0.622 | 0.831 | -0.017 | -0.045 | 0.010 | 0.220 | 0.429 |
| PC(14:0_16:0) | -0.017 | -0.039 | 0.005 | 0.123 | 0.903 | -0.009 | -0.023 | 0.004 | 0.184 | 0.453 | -0.003 | -0.013 | 0.008 | 0.626 | 0.831 | -0.026 | -0.054 | 0.001 | 0.058 | 0.177 |
| PI(34:1) | -0.012 | -0.034 | 0.010 | 0.296 | 0.903 | -0.005 | -0.019 | 0.009 | 0.505 | 0.722 | 0.003 | -0.008 | 0.014 | 0.630 | 0.831 | -0.008 | -0.035 | 0.020 | 0.587 | 0.756 |
| methyl-DE(18:2) | 0.015 | -0.007 | 0.037 | 0.188 | 0.903 | 0.004 | -0.010 | 0.018 | 0.584 | 0.763 | -0.003 | -0.014 | 0.008 | 0.625 | 0.831 | 0.000 | -0.027 | 0.028 | 0.974 | 0.982 |
| LPC(15:0) [sn1] | -0.011 | -0.033 | 0.012 | 0.348 | 0.922 | -0.011 | -0.025 | 0.003 | 0.114 | 0.382 | -0.003 | -0.013 | 0.008 | 0.629 | 0.831 | -0.027 | -0.054 | 0.001 | 0.060 | 0.179 |
| PC(28:0) | -0.009 | -0.031 | 0.013 | 0.418 | 0.922 | -0.007 | -0.021 | 0.007 | 0.318 | 0.557 | -0.003 | -0.013 | 0.008 | 0.630 | 0.831 | -0.026 | -0.053 | 0.001 | 0.061 | 0.183 |
| HexCer(d16:1/24:0) | 0.009 | -0.013 | 0.031 | 0.424 | 0.922 | -0.002 | -0.016 | 0.012 | 0.766 | 0.881 | -0.003 | -0.014 | 0.008 | 0.627 | 0.831 | 0.006 | -0.022 | 0.033 | 0.694 | 0.840 |
| PS(40:6) | 0.008 | -0.014 | 0.031 | 0.456 | 0.922 | -0.017 | -0.031 | -0.003 | 0.020 | 0.273 | 0.003 | -0.008 | 0.014 | 0.630 | 0.831 | 0.003 | -0.025 | 0.031 | 0.842 | 0.928 |
| PC(P-18:0/22:5) | -0.010 | -0.032 | 0.013 | 0.401 | 0.922 | -0.008 | -0.022 | 0.006 | 0.259 | 0.509 | -0.003 | -0.013 | 0.008 | 0.638 | 0.835 | -0.038 | -0.065 | -0.012 | 0.005 | 0.044 |
| PS(38:4) | 0.009 | -0.014 | 0.031 | 0.440 | 0.922 | -0.010 | -0.024 | 0.003 | 0.132 | 0.395 | -0.003 | -0.013 | 0.008 | 0.637 | 0.835 | -0.010 | -0.038 | 0.017 | 0.453 | 0.657 |
| FA(18:0) | -0.005 | -0.027 | 0.018 | 0.680 | 0.964 | -0.002 | -0.015 | 0.011 | 0.766 | 0.881 | 0.003 | -0.008 | 0.013 | 0.637 | 0.835 | 0.008 | -0.017 | 0.034 | 0.535 | 0.719 |
| CE(20:0) | -0.003 | -0.026 | 0.019 | 0.764 | 0.969 | 0.007 | -0.006 | 0.021 | 0.303 | 0.546 | -0.003 | -0.013 | 0.008 | 0.635 | 0.835 | -0.012 | -0.039 | 0.015 | 0.380 | 0.596 |
| S1P(d16:1) | 0.000 | -0.023 | 0.022 | 0.980 | 0.995 | -0.006 | -0.020 | 0.008 | 0.384 | 0.624 | -0.003 | -0.013 | 0.008 | 0.639 | 0.835 | -0.010 | -0.038 | 0.018 | 0.471 | 0.665 |
| PI(37:6) | 0.005 | -0.017 | 0.028 | 0.638 | 0.960 | -0.010 | -0.024 | 0.004 | 0.177 | 0.447 | 0.003 | -0.008 | 0.013 | 0.642 | 0.837 | -0.038 | -0.067 | -0.010 | 0.007 | 0.049 |
| FA(16:0) | -0.004 | -0.026 | 0.019 | 0.753 | 0.969 | 0.003 | -0.011 | 0.016 | 0.717 | 0.855 | 0.003 | -0.008 | 0.013 | 0.644 | 0.837 | 0.000 | -0.027 | 0.027 | 0.988 | 0.992 |
| PE(P-18:1/18:3) | 0.000 | -0.022 | 0.023 | 0.979 | 0.995 | -0.002 | -0.016 | 0.012 | 0.765 | 0.881 | -0.003 | -0.013 | 0.008 | 0.644 | 0.837 | -0.005 | -0.032 | 0.022 | 0.737 | 0.872 |
| CE(22:6) | 0.008 | -0.014 | 0.030 | 0.494 | 0.922 | -0.003 | -0.016 | 0.011 | 0.715 | 0.855 | -0.002 | -0.013 | 0.008 | 0.650 | 0.841 | -0.029 | -0.057 | -0.001 | 0.042 | 0.147 |
| PS(36:1) | -0.007 | -0.029 | 0.015 | 0.548 | 0.929 | -0.014 | -0.028 | 0.000 | 0.045 | 0.306 | 0.003 | -0.008 | 0.013 | 0.650 | 0.841 | -0.001 | -0.029 | 0.026 | 0.928 | 0.964 |
| SM(d17:1/14:0) | -0.004 | -0.027 | 0.018 | 0.709 | 0.964 | -0.005 | -0.019 | 0.009 | 0.466 | 0.686 | -0.002 | -0.013 | 0.008 | 0.649 | 0.841 | -0.033 | -0.060 | -0.005 | 0.021 | 0.093 |
| LPC(20:3) [sn1] | -0.012 | -0.034 | 0.010 | 0.291 | 0.903 | -0.012 | -0.025 | 0.002 | 0.092 | 0.356 | -0.002 | -0.013 | 0.008 | 0.653 | 0.842 | -0.019 | -0.046 | 0.009 | 0.180 | 0.386 |
| CE(24:6) | 0.004 | -0.018 | 0.026 | 0.740 | 0.964 | -0.005 | -0.019 | 0.009 | 0.480 | 0.697 | -0.003 | -0.014 | 0.008 | 0.652 | 0.842 | -0.006 | -0.034 | 0.021 | 0.656 | 0.814 |
| PC(O-18:0/20:4) | -0.009 | -0.031 | 0.013 | 0.403 | 0.922 | 0.000 | -0.014 | 0.014 | 0.997 | 0.998 | -0.002 | -0.013 | 0.008 | 0.658 | 0.846 | -0.033 | -0.060 | -0.006 | 0.016 | 0.080 |
| PC(18:0_18:1) | -0.010 | -0.032 | 0.012 | 0.364 | 0.922 | -0.015 | -0.029 | -0.001 | 0.034 | 0.298 | -0.002 | -0.013 | 0.008 | 0.658 | 0.846 | -0.032 | -0.060 | -0.005 | 0.022 | 0.097 |
| PE(18:0_22:5) (n3) | -0.012 | -0.034 | 0.009 | 0.262 | 0.903 | -0.014 | -0.027 | 0.000 | 0.052 | 0.317 | -0.002 | -0.013 | 0.008 | 0.662 | 0.847 | -0.021 | -0.048 | 0.007 | 0.141 | 0.330 |
| TG(50:1) [NL-14:0] | -0.016 | -0.038 | 0.006 | 0.163 | 0.903 | -0.010 | -0.024 | 0.003 | 0.134 | 0.396 | 0.002 | -0.008 | 0.013 | 0.663 | 0.847 | 0.003 | -0.026 | 0.031 | 0.862 | 0.937 |
| Cer(d18:1/24:0) | -0.002 | -0.024 | 0.020 | 0.881 | 0.981 | 0.003 | -0.011 | 0.017 | 0.670 | 0.831 | -0.002 | -0.013 | 0.008 | 0.661 | 0.847 | -0.007 | -0.034 | 0.021 | 0.639 | 0.800 |
| Cer(m18:0/20:0) | -0.018 | -0.040 | 0.004 | 0.105 | 0.903 | -0.013 | -0.027 | 0.001 | 0.062 | 0.332 | 0.002 | -0.008 | 0.013 | 0.666 | 0.850 | -0.012 | -0.040 | 0.015 | 0.373 | 0.590 |
| LPC(17:1) [sn1] (b) | -0.012 | -0.035 | 0.011 | 0.300 | 0.903 | -0.012 | -0.026 | 0.002 | 0.084 | 0.356 | -0.002 | -0.013 | 0.008 | 0.675 | 0.853 | -0.031 | -0.059 | -0.003 | 0.029 | 0.118 |
| PE(P-18:0/22:4) | -0.013 | -0.035 | 0.009 | 0.241 | 0.903 | 0.008 | -0.006 | 0.022 | 0.274 | 0.521 | -0.002 | -0.013 | 0.008 | 0.675 | 0.853 | -0.015 | -0.042 | 0.012 | 0.287 | 0.509 |
| TG(48:2) [NL-14:0] | -0.019 | -0.041 | 0.003 | 0.092 | 0.903 | -0.013 | -0.026 | 0.001 | 0.069 | 0.347 | -0.002 | -0.013 | 0.008 | 0.674 | 0.853 | -0.009 | -0.037 | 0.019 | 0.519 | 0.709 |
| PE(17:0_20:4) | -0.010 | -0.032 | 0.012 | 0.372 | 0.922 | -0.013 | -0.027 | 0.001 | 0.072 | 0.350 | -0.002 | -0.013 | 0.008 | 0.676 | 0.853 | -0.029 | -0.057 | -0.001 | 0.042 | 0.147 |
| SM(d17:1/16:0) | -0.004 | -0.026 | 0.018 | 0.738 | 0.964 | 0.000 | -0.014 | 0.014 | 0.957 | 0.976 | -0.002 | -0.013 | 0.009 | 0.675 | 0.853 | -0.028 | -0.056 | -0.001 | 0.044 | 0.150 |
| LPC(22:6) [+OH] | 0.002 | -0.021 | 0.024 | 0.883 | 0.981 | -0.005 | -0.020 | 0.009 | 0.445 | 0.672 | -0.002 | -0.013 | 0.009 | 0.677 | 0.853 | -0.001 | -0.029 | 0.027 | 0.943 | 0.966 |
| LPI(20:4) [sn1] | -0.001 | -0.023 | 0.021 | 0.961 | 0.995 | -0.002 | -0.016 | 0.013 | 0.826 | 0.913 | 0.002 | -0.008 | 0.013 | 0.677 | 0.853 | 0.006 | -0.021 | 0.033 | 0.669 | 0.818 |

| | | | | | | | | | | | | | | | | | | | | |
|---|---|---|---|---|---|---|---|---|---|---|---|---|---|---|---|---|---|---|---|---|
| SM(d16:1/23:0) & SM(d17:1/22:0) | 0.000 | -0.022 | 0.022 | 0.990 | 0.996 | -0.001 | -0.015 | 0.013 | 0.904 | 0.943 | 0.002 | -0.008 | 0.013 | 0.670 | 0.853 | -0.014 | -0.041 | 0.013 | 0.318 | 0.538 |
| DG(16:1_18:1) | -0.018 | -0.040 | 0.004 | 0.107 | 0.903 | -0.016 | -0.030 | -0.003 | 0.020 | 0.273 | -0.002 | -0.013 | 0.008 | 0.681 | 0.855 | -0.029 | -0.056 | -0.001 | 0.042 | 0.147 |
| CE(20:5) | 0.015 | -0.007 | 0.037 | 0.192 | 0.903 | 0.001 | -0.013 | 0.015 | 0.875 | 0.934 | 0.002 | -0.008 | 0.013 | 0.681 | 0.855 | -0.010 | -0.038 | 0.018 | 0.475 | 0.668 |
| methyl-DE(18:1) | 0.016 | -0.006 | 0.038 | 0.159 | 0.903 | -0.001 | -0.015 | 0.013 | 0.852 | 0.929 | 0.002 | -0.009 | 0.013 | 0.684 | 0.855 | 0.003 | -0.025 | 0.030 | 0.839 | 0.928 |
| dimethyl-CE(18:1) | 0.011 | -0.011 | 0.033 | 0.329 | 0.922 | -0.005 | -0.019 | 0.009 | 0.466 | 0.686 | 0.002 | -0.009 | 0.013 | 0.683 | 0.855 | -0.016 | -0.044 | 0.012 | 0.260 | 0.479 |
| PE(O-18:1/18:2) | -0.004 | -0.027 | 0.018 | 0.699 | 0.964 | 0.008 | -0.006 | 0.022 | 0.290 | 0.536 | 0.002 | -0.009 | 0.013 | 0.685 | 0.855 | -0.007 | -0.035 | 0.020 | 0.603 | 0.767 |
| PE(18:0_18:2) | -0.013 | -0.035 | 0.009 | 0.243 | 0.903 | -0.012 | -0.026 | 0.002 | 0.095 | 0.360 | -0.002 | -0.013 | 0.009 | 0.689 | 0.858 | 0.000 | -0.027 | 0.028 | 0.979 | 0.985 |
| methyl-CE(20:4) | 0.013 | -0.010 | 0.035 | 0.268 | 0.903 | 0.009 | -0.006 | 0.023 | 0.233 | 0.490 | 0.002 | -0.009 | 0.013 | 0.694 | 0.863 | -0.006 | -0.033 | 0.022 | 0.685 | 0.830 |
| PE(15-MHDA_20:4) | -0.015 | -0.037 | 0.007 | 0.176 | 0.903 | -0.010 | -0.024 | 0.004 | 0.145 | 0.406 | 0.002 | -0.009 | 0.013 | 0.695 | 0.864 | -0.020 | -0.048 | 0.007 | 0.146 | 0.337 |
| PI(38:6) | -0.006 | -0.028 | 0.016 | 0.601 | 0.950 | -0.012 | -0.026 | 0.002 | 0.092 | 0.356 | -0.002 | -0.013 | 0.009 | 0.697 | 0.864 | -0.017 | -0.045 | 0.010 | 0.220 | 0.430 |
| CE(17:1) | -0.005 | -0.027 | 0.017 | 0.652 | 0.960 | -0.008 | -0.022 | 0.005 | 0.241 | 0.495 | -0.002 | -0.013 | 0.009 | 0.704 | 0.872 | -0.038 | -0.066 | -0.011 | 0.007 | 0.048 |
| PC(18:0_22:4) | -0.015 | -0.037 | 0.006 | 0.162 | 0.903 | 0.001 | -0.013 | 0.015 | 0.903 | 0.943 | -0.002 | -0.013 | 0.009 | 0.707 | 0.873 | -0.019 | -0.046 | 0.008 | 0.164 | 0.366 |
| LPE(22:6) [sn1] | -0.011 | -0.033 | 0.012 | 0.346 | 0.922 | -0.012 | -0.026 | 0.001 | 0.082 | 0.356 | -0.002 | -0.013 | 0.009 | 0.707 | 0.873 | -0.012 | -0.040 | 0.015 | 0.372 | 0.590 |
| PC(33:2) | -0.010 | -0.032 | 0.012 | 0.383 | 0.922 | -0.007 | -0.021 | 0.007 | 0.347 | 0.589 | -0.002 | -0.013 | 0.009 | 0.710 | 0.873 | -0.031 | -0.058 | -0.004 | 0.026 | 0.107 |
| PE(18:1_18:1) | 0.000 | -0.022 | 0.022 | 0.988 | 0.996 | -0.013 | -0.027 | 0.000 | 0.059 | 0.323 | -0.002 | -0.013 | 0.009 | 0.710 | 0.873 | 0.009 | -0.019 | 0.037 | 0.512 | 0.706 |
| LPC(26:0) [sn2] | 0.006 | -0.016 | 0.029 | 0.569 | 0.933 | -0.007 | -0.021 | 0.007 | 0.322 | 0.560 | -0.002 | -0.013 | 0.009 | 0.715 | 0.878 | -0.009 | -0.037 | 0.018 | 0.517 | 0.709 |
| PE(P-18:1/20:5) (b) | 0.008 | -0.014 | 0.030 | 0.475 | 0.922 | -0.008 | -0.022 | 0.006 | 0.292 | 0.536 | 0.002 | -0.009 | 0.013 | 0.722 | 0.885 | -0.022 | -0.049 | 0.006 | 0.119 | 0.293 |
| DG(18:0_20:4) | -0.017 | -0.039 | 0.005 | 0.124 | 0.903 | -0.013 | -0.027 | 0.000 | 0.053 | 0.317 | 0.002 | -0.009 | 0.013 | 0.726 | 0.888 | -0.001 | -0.029 | 0.027 | 0.948 | 0.968 |
| TG(52:5) [NL-20:4] | -0.020 | -0.042 | 0.002 | 0.076 | 0.903 | -0.006 | -0.020 | 0.007 | 0.351 | 0.592 | 0.002 | -0.009 | 0.012 | 0.739 | 0.892 | -0.002 | -0.030 | 0.026 | 0.901 | 0.952 |
| LPI(18:1) [sn2] | 0.012 | -0.010 | 0.034 | 0.287 | 0.903 | 0.001 | -0.013 | 0.014 | 0.903 | 0.943 | -0.002 | -0.013 | 0.009 | 0.734 | 0.892 | -0.002 | -0.028 | 0.025 | 0.908 | 0.955 |
| CE(14:0) | -0.009 | -0.031 | 0.014 | 0.450 | 0.922 | -0.013 | -0.027 | 0.001 | 0.065 | 0.338 | -0.002 | -0.013 | 0.009 | 0.738 | 0.892 | -0.021 | -0.049 | 0.007 | 0.137 | 0.323 |
| DG(16:0_22:5) | -0.011 | -0.033 | 0.011 | 0.333 | 0.922 | -0.015 | -0.028 | -0.002 | 0.028 | 0.285 | -0.002 | -0.013 | 0.009 | 0.735 | 0.892 | -0.017 | -0.044 | 0.011 | 0.229 | 0.439 |
| FA(20:5) | 0.010 | -0.012 | 0.032 | 0.371 | 0.922 | -0.013 | -0.027 | 0.001 | 0.066 | 0.338 | 0.002 | -0.009 | 0.013 | 0.738 | 0.892 | -0.001 | -0.028 | 0.027 | 0.957 | 0.970 |
| Cer(d17:1/23:0) | -0.007 | -0.029 | 0.015 | 0.536 | 0.923 | -0.005 | -0.020 | 0.009 | 0.450 | 0.674 | -0.002 | -0.013 | 0.009 | 0.739 | 0.892 | -0.017 | -0.044 | 0.010 | 0.225 | 0.434 |
| PC(36:6) (a) | 0.004 | -0.019 | 0.026 | 0.744 | 0.964 | -0.009 | -0.023 | 0.005 | 0.199 | 0.460 | -0.002 | -0.013 | 0.009 | 0.740 | 0.892 | -0.019 | -0.046 | 0.008 | 0.175 | 0.378 |
| PC(15:0_22:6) | 0.001 | -0.021 | 0.023 | 0.942 | 0.995 | -0.012 | -0.026 | 0.002 | 0.100 | 0.366 | 0.002 | -0.009 | 0.013 | 0.732 | 0.892 | -0.026 | -0.055 | 0.002 | 0.065 | 0.186 |
| FA(18:1) | -0.001 | -0.023 | 0.022 | 0.944 | 0.995 | 0.001 | -0.013 | 0.015 | 0.872 | 0.932 | -0.002 | -0.013 | 0.009 | 0.740 | 0.892 | -0.017 | -0.045 | 0.011 | 0.231 | 0.440 |
| Cer(d18:1/26:0) | -0.012 | -0.035 | 0.010 | 0.286 | 0.903 | 0.005 | -0.009 | 0.019 | 0.503 | 0.720 | -0.002 | -0.013 | 0.009 | 0.743 | 0.893 | -0.015 | -0.043 | 0.012 | 0.270 | 0.495 |
| PE(15-MHDA_22:6) | -0.007 | -0.030 | 0.015 | 0.520 | 0.922 | -0.014 | -0.028 | 0.000 | 0.044 | 0.306 | 0.002 | -0.009 | 0.013 | 0.742 | 0.893 | -0.014 | -0.042 | 0.014 | 0.327 | 0.545 |
| LPI(18:0) [sn2] | -0.010 | -0.032 | 0.012 | 0.371 | 0.922 | -0.005 | -0.019 | 0.010 | 0.530 | 0.735 | -0.002 | -0.013 | 0.009 | 0.745 | 0.894 | -0.002 | -0.029 | 0.026 | 0.896 | 0.952 |
| TG(48:2) [NL-18:2] | -0.022 | -0.043 | 0.000 | 0.051 | 0.903 | -0.010 | -0.024 | 0.003 | 0.142 | 0.406 | -0.002 | -0.012 | 0.009 | 0.748 | 0.896 | -0.005 | -0.034 | 0.023 | 0.707 | 0.845 |
| PI(18:0_22:4) | -0.026 | -0.047 | -0.004 | 0.020 | 0.903 | -0.012 | -0.026 | 0.002 | 0.103 | 0.371 | -0.002 | -0.013 | 0.009 | 0.750 | 0.897 | -0.019 | -0.046 | 0.008 | 0.167 | 0.371 |
| TG(56:8) [NL-20:5] | 0.003 | -0.019 | 0.025 | 0.785 | 0.973 | -0.007 | -0.021 | 0.007 | 0.331 | 0.572 | 0.002 | -0.009 | 0.013 | 0.751 | 0.898 | 0.003 | -0.025 | 0.030 | 0.853 | 0.933 |
| PE(P-18:1/20:3) (b) | -0.012 | -0.034 | 0.010 | 0.304 | 0.903 | -0.004 | -0.018 | 0.010 | 0.576 | 0.758 | -0.002 | -0.013 | 0.009 | 0.753 | 0.898 | -0.031 | -0.057 | -0.004 | 0.025 | 0.106 |
| PC(44:12) | 0.009 | -0.013 | 0.031 | 0.413 | 0.922 | -0.008 | -0.022 | 0.006 | 0.273 | 0.521 | 0.002 | -0.009 | 0.013 | 0.758 | 0.902 | 0.003 | -0.026 | 0.031 | 0.851 | 0.933 |
| TG(54:2) [NL-20:1] | -0.009 | -0.031 | 0.013 | 0.406 | 0.922 | -0.017 | -0.031 | -0.004 | 0.014 | 0.273 | 0.002 | -0.009 | 0.012 | 0.765 | 0.909 | -0.007 | -0.035 | 0.021 | 0.632 | 0.794 |
| PE(O-18:0/22:5) | -0.008 | -0.030 | 0.014 | 0.491 | 0.922 | 0.003 | -0.011 | 0.017 | 0.646 | 0.811 | 0.002 | -0.009 | 0.012 | 0.771 | 0.915 | -0.020 | -0.047 | 0.007 | 0.141 | 0.330 |
| PA(40:6) | 0.000 | -0.021 | 0.022 | 0.984 | 0.996 | -0.007 | -0.022 | 0.007 | 0.318 | 0.557 | -0.002 | -0.012 | 0.009 | 0.774 | 0.917 | -0.007 | -0.036 | 0.023 | 0.657 | 0.814 |
| LPC(15:0) [sn2] | -0.011 | -0.033 | 0.011 | 0.335 | 0.922 | -0.011 | -0.025 | 0.002 | 0.100 | 0.366 | -0.002 | -0.012 | 0.009 | 0.777 | 0.919 | -0.027 | -0.055 | 0.001 | 0.055 | 0.173 |
| LPI(20:4) [sn2] | -0.010 | -0.032 | 0.012 | 0.363 | 0.922 | -0.003 | -0.017 | 0.011 | 0.684 | 0.840 | 0.002 | -0.009 | 0.012 | 0.779 | 0.921 | -0.010 | -0.037 | 0.018 | 0.486 | 0.678 |
| FA(17:0) | -0.005 | -0.027 | 0.018 | 0.694 | 0.964 | -0.006 | -0.021 | 0.009 | 0.409 | 0.645 | -0.002 | -0.013 | 0.010 | 0.781 | 0.921 | 0.001 | -0.027 | 0.029 | 0.958 | 0.970 |
| TG(50:4) [NL-20:4] | -0.020 | -0.042 | 0.002 | 0.075 | 0.903 | -0.008 | -0.021 | 0.006 | 0.273 | 0.521 | 0.001 | -0.009 | 0.012 | 0.783 | 0.921 | 0.006 | -0.022 | 0.034 | 0.682 | 0.827 |
| SM(d18:2/23:0) | 0.002 | -0.020 | 0.024 | 0.854 | 0.981 | 0.000 | -0.014 | 0.014 | 0.996 | 0.998 | -0.002 | -0.012 | 0.009 | 0.783 | 0.921 | -0.024 | -0.051 | 0.003 | 0.081 | 0.221 |
| PC(16:0_22:6) | 0.002 | -0.020 | 0.024 | 0.867 | 0.981 | -0.013 | -0.027 | 0.002 | 0.084 | 0.356 | 0.002 | -0.009 | 0.012 | 0.786 | 0.922 | -0.023 | -0.052 | 0.005 | 0.102 | 0.261 |
| Cer(d19:1/16:0) | 0.001 | -0.021 | 0.024 | 0.902 | 0.985 | -0.006 | -0.020 | 0.008 | 0.390 | 0.631 | -0.001 | -0.012 | 0.009 | 0.789 | 0.925 | -0.016 | -0.044 | 0.012 | 0.272 | 0.498 |
| TG(51:2) [NL-17:1] | -0.017 | -0.039 | 0.005 | 0.126 | 0.903 | -0.019 | -0.033 | -0.005 | 0.006 | 0.259 | 0.001 | -0.009 | 0.012 | 0.792 | 0.925 | -0.023 | -0.051 | 0.004 | 0.099 | 0.256 |
| CE(24:5) | 0.003 | -0.019 | 0.025 | 0.777 | 0.973 | -0.001 | -0.015 | 0.012 | 0.832 | 0.917 | -0.001 | -0.012 | 0.009 | 0.791 | 0.925 | -0.013 | -0.041 | 0.014 | 0.352 | 0.572 |
| Cer(d19:1/22:0) | 0.004 | -0.018 | 0.027 | 0.708 | 0.964 | -0.004 | -0.018 | 0.009 | 0.532 | 0.737 | -0.001 | -0.012 | 0.009 | 0.794 | 0.926 | -0.027 | -0.055 | 0.000 | 0.048 | 0.157 |
| TG(48:2) [NL-16:1] | -0.021 | -0.044 | 0.001 | 0.057 | 0.903 | -0.015 | -0.028 | -0.001 | 0.036 | 0.301 | -0.001 | -0.012 | 0.009 | 0.795 | 0.927 | -0.016 | -0.044 | 0.011 | 0.243 | 0.457 |
| DG(14:0_16:0) | -0.015 | -0.037 | 0.007 | 0.186 | 0.903 | -0.011 | -0.024 | 0.003 | 0.121 | 0.385 | 0.001 | -0.009 | 0.012 | 0.797 | 0.927 | 0.006 | -0.021 | 0.033 | 0.661 | 0.814 |

| | | | | | | | | | | | | | | | | | | | | |
|---|---|---|---|---|---|---|---|---|---|---|---|---|---|---|---|---|---|---|---|---|
| Cer(d19:1/24:1) | -0.002 | -0.025 | 0.020 | 0.857 | 0.981 | -0.017 | -0.031 | -0.003 | 0.018 | 0.273 | -0.001 | -0.012 | 0.009 | 0.801 | 0.930 | -0.030 | -0.057 | -0.002 | 0.035 | 0.132 |
| PE(O-16:0/22:4) | -0.016 | -0.038 | 0.006 | 0.163 | 0.903 | 0.009 | -0.005 | 0.023 | 0.194 | 0.454 | 0.001 | -0.009 | 0.012 | 0.807 | 0.932 | -0.006 | -0.033 | 0.021 | 0.669 | 0.818 |
| SM(d16:1/24:1) | -0.004 | -0.027 | 0.018 | 0.694 | 0.964 | -0.012 | -0.026 | 0.002 | 0.089 | 0.356 | 0.001 | -0.009 | 0.012 | 0.805 | 0.932 | -0.027 | -0.055 | 0.001 | 0.057 | 0.173 |
| PE(P-20:0/22:6) | 0.004 | -0.019 | 0.026 | 0.747 | 0.964 | -0.012 | -0.026 | 0.003 | 0.112 | 0.382 | 0.001 | -0.010 | 0.012 | 0.806 | 0.932 | -0.026 | -0.054 | 0.002 | 0.069 | 0.195 |
| TG(52:2) [NL-18:2] | -0.025 | -0.046 | -0.004 | 0.023 | 0.903 | -0.007 | -0.021 | 0.006 | 0.293 | 0.536 | 0.001 | -0.009 | 0.012 | 0.810 | 0.934 | 0.005 | -0.022 | 0.033 | 0.712 | 0.847 |
| PE(16:0_22:6) | -0.002 | -0.024 | 0.020 | 0.848 | 0.981 | -0.012 | -0.026 | 0.001 | 0.075 | 0.355 | 0.001 | -0.009 | 0.012 | 0.809 | 0.934 | -0.012 | -0.040 | 0.016 | 0.406 | 0.620 |
| dimethyl-CE(18:2) | 0.012 | -0.010 | 0.034 | 0.287 | 0.903 | 0.002 | -0.012 | 0.016 | 0.771 | 0.881 | -0.001 | -0.012 | 0.010 | 0.814 | 0.935 | -0.011 | -0.038 | 0.017 | 0.456 | 0.660 |
| SM(43:2) (b) | -0.007 | -0.029 | 0.015 | 0.561 | 0.933 | -0.015 | -0.029 | -0.001 | 0.034 | 0.298 | 0.001 | -0.009 | 0.012 | 0.813 | 0.935 | -0.038 | -0.065 | -0.011 | 0.007 | 0.048 |
| Cer(d20:1/24:1) | -0.012 | -0.034 | 0.010 | 0.273 | 0.903 | -0.012 | -0.026 | 0.002 | 0.089 | 0.356 | 0.001 | -0.010 | 0.012 | 0.817 | 0.937 | -0.023 | -0.050 | 0.005 | 0.107 | 0.272 |
| PC(31:0) (b) | -0.008 | -0.030 | 0.014 | 0.491 | 0.922 | -0.007 | -0.021 | 0.007 | 0.320 | 0.559 | 0.001 | -0.009 | 0.012 | 0.821 | 0.940 | -0.036 | -0.063 | -0.008 | 0.011 | 0.064 |
| CE(16:1) | -0.008 | -0.030 | 0.015 | 0.505 | 0.922 | -0.011 | -0.024 | 0.003 | 0.127 | 0.385 | -0.001 | -0.012 | 0.010 | 0.824 | 0.940 | -0.018 | -0.045 | 0.010 | 0.209 | 0.419 |
| TG(50:2) [NL-14:0] | -0.002 | -0.024 | 0.020 | 0.853 | 0.981 | -0.017 | -0.031 | -0.003 | 0.015 | 0.273 | -0.001 | -0.012 | 0.010 | 0.825 | 0.940 | -0.016 | -0.045 | 0.013 | 0.276 | 0.500 |
| TG(56:6) [NL-20:4] | -0.002 | -0.024 | 0.020 | 0.850 | 0.981 | 0.002 | -0.012 | 0.016 | 0.807 | 0.899 | 0.001 | -0.010 | 0.012 | 0.823 | 0.940 | -0.004 | -0.032 | 0.024 | 0.771 | 0.892 |
| PE(P-18:0/18:2) | -0.008 | -0.030 | 0.014 | 0.500 | 0.922 | 0.004 | -0.010 | 0.018 | 0.570 | 0.752 | -0.001 | -0.012 | 0.010 | 0.827 | 0.941 | -0.008 | -0.035 | 0.019 | 0.571 | 0.745 |
| FA(14:0) | 0.008 | -0.014 | 0.030 | 0.460 | 0.922 | 0.001 | -0.013 | 0.015 | 0.891 | 0.939 | -0.001 | -0.012 | 0.010 | 0.829 | 0.942 | -0.012 | -0.040 | 0.016 | 0.401 | 0.617 |
| PI(18:0_20:3) (b) | -0.016 | -0.038 | 0.007 | 0.170 | 0.903 | -0.026 | -0.040 | -0.012 | 0.000 | 0.092 | -0.001 | -0.012 | 0.010 | 0.832 | 0.943 | -0.038 | -0.066 | -0.011 | 0.007 | 0.048 |
| PI(15-MHDA_18:1) & PI(17:0_18:1) | -0.013 | -0.035 | 0.009 | 0.259 | 0.903 | -0.011 | -0.025 | 0.003 | 0.134 | 0.396 | -0.001 | -0.012 | 0.010 | 0.831 | 0.943 | -0.015 | -0.042 | 0.013 | 0.296 | 0.520 |
| dimethyl-CE(20:4) | 0.012 | -0.011 | 0.034 | 0.305 | 0.903 | 0.005 | -0.009 | 0.019 | 0.458 | 0.682 | -0.001 | -0.012 | 0.010 | 0.837 | 0.944 | -0.017 | -0.045 | 0.010 | 0.214 | 0.422 |
| Cer(m18:1/20:0) | -0.021 | -0.043 | 0.001 | 0.064 | 0.903 | -0.010 | -0.023 | 0.004 | 0.161 | 0.432 | 0.001 | -0.010 | 0.012 | 0.838 | 0.944 | -0.011 | -0.038 | 0.016 | 0.432 | 0.640 |
| TG(48:3) [NL-18:3] | -0.020 | -0.042 | 0.002 | 0.071 | 0.903 | -0.009 | -0.023 | 0.004 | 0.182 | 0.452 | -0.001 | -0.012 | 0.010 | 0.837 | 0.944 | 0.002 | -0.026 | 0.030 | 0.870 | 0.943 |
| LPC(22:6) [sn2] | 0.003 | -0.019 | 0.026 | 0.765 | 0.969 | -0.014 | -0.028 | 0.000 | 0.045 | 0.306 | -0.001 | -0.012 | 0.010 | 0.837 | 0.944 | -0.014 | -0.042 | 0.014 | 0.321 | 0.540 |
| PC(34:2) [+OH] | -0.013 | -0.035 | 0.009 | 0.248 | 0.903 | -0.002 | -0.016 | 0.011 | 0.766 | 0.881 | 0.001 | -0.010 | 0.012 | 0.844 | 0.948 | 0.004 | -0.023 | 0.032 | 0.762 | 0.891 |
| PC(P-18:1/22:6) | 0.002 | -0.020 | 0.024 | 0.871 | 0.981 | -0.012 | -0.026 | 0.002 | 0.088 | 0.356 | -0.001 | -0.012 | 0.010 | 0.843 | 0.948 | -0.028 | -0.055 | -0.001 | 0.044 | 0.150 |
| PE(20:0_20:4) | -0.016 | -0.038 | 0.007 | 0.167 | 0.903 | -0.005 | -0.018 | 0.009 | 0.512 | 0.724 | 0.001 | -0.010 | 0.012 | 0.851 | 0.954 | 0.011 | -0.016 | 0.038 | 0.439 | 0.644 |
| PC(32:1) | -0.020 | -0.042 | 0.002 | 0.075 | 0.903 | -0.011 | -0.025 | 0.003 | 0.113 | 0.382 | -0.001 | -0.012 | 0.010 | 0.860 | 0.962 | -0.027 | -0.054 | 0.001 | 0.056 | 0.173 |
| DG(18:2_20:4) | -0.015 | -0.037 | 0.007 | 0.183 | 0.903 | 0.002 | -0.012 | 0.016 | 0.790 | 0.891 | -0.001 | -0.012 | 0.010 | 0.862 | 0.962 | -0.001 | -0.029 | 0.026 | 0.920 | 0.963 |
| LPE(22:6) [sn2] | -0.010 | -0.032 | 0.013 | 0.402 | 0.922 | -0.012 | -0.025 | 0.002 | 0.088 | 0.356 | -0.001 | -0.012 | 0.010 | 0.862 | 0.962 | -0.011 | -0.039 | 0.016 | 0.412 | 0.628 |
| SM(d18:1/24:1) | -0.012 | -0.034 | 0.010 | 0.298 | 0.903 | -0.011 | -0.025 | 0.003 | 0.125 | 0.385 | -0.001 | -0.012 | 0.010 | 0.884 | 0.964 | -0.029 | -0.056 | -0.001 | 0.041 | 0.146 |
| SM(37:2) | -0.012 | -0.034 | 0.010 | 0.292 | 0.903 | 0.000 | -0.015 | 0.014 | 0.950 | 0.972 | 0.001 | -0.010 | 0.012 | 0.883 | 0.964 | -0.016 | -0.043 | 0.011 | 0.243 | 0.456 |
| PI(16:0_20:4) | -0.024 | -0.046 | -0.002 | 0.033 | 0.903 | -0.004 | -0.018 | 0.010 | 0.554 | 0.748 | -0.001 | -0.012 | 0.010 | 0.878 | 0.964 | -0.015 | -0.042 | 0.013 | 0.294 | 0.518 |
| TG(51:2) [NL-17:0] | -0.019 | -0.041 | 0.002 | 0.082 | 0.903 | -0.016 | -0.029 | -0.002 | 0.023 | 0.285 | 0.001 | -0.010 | 0.011 | 0.895 | 0.964 | -0.012 | -0.040 | 0.016 | 0.392 | 0.609 |
| DG(16:0_16:1) | -0.020 | -0.043 | 0.002 | 0.071 | 0.903 | -0.011 | -0.025 | 0.002 | 0.110 | 0.380 | -0.001 | -0.011 | 0.010 | 0.895 | 0.964 | -0.011 | -0.038 | 0.016 | 0.434 | 0.641 |
| methyl-CE(22:6) | 0.018 | -0.005 | 0.040 | 0.120 | 0.903 | 0.000 | -0.013 | 0.014 | 0.973 | 0.983 | -0.001 | -0.011 | 0.010 | 0.892 | 0.964 | -0.009 | -0.037 | 0.019 | 0.535 | 0.719 |
| Cer(d16:1/23:0) | -0.012 | -0.034 | 0.010 | 0.291 | 0.903 | -0.011 | -0.025 | 0.003 | 0.114 | 0.382 | -0.001 | -0.012 | 0.010 | 0.887 | 0.964 | -0.007 | -0.035 | 0.020 | 0.601 | 0.765 |
| dimethyl-CE(22:6) | 0.014 | -0.009 | 0.036 | 0.231 | 0.903 | 0.000 | -0.014 | 0.014 | 0.963 | 0.979 | 0.001 | -0.010 | 0.012 | 0.891 | 0.964 | -0.005 | -0.033 | 0.022 | 0.708 | 0.845 |
| PG(36:2) | -0.019 | -0.041 | 0.004 | 0.102 | 0.903 | -0.009 | -0.023 | 0.004 | 0.166 | 0.432 | -0.001 | -0.012 | 0.010 | 0.883 | 0.964 | 0.004 | -0.024 | 0.032 | 0.770 | 0.892 |
| PC(16:0_18:1) | -0.011 | -0.032 | 0.011 | 0.348 | 0.922 | -0.012 | -0.026 | 0.002 | 0.095 | 0.360 | -0.001 | -0.012 | 0.010 | 0.888 | 0.964 | -0.039 | -0.067 | -0.011 | 0.006 | 0.048 |
| CE(17:0) | -0.009 | -0.031 | 0.013 | 0.437 | 0.922 | -0.006 | -0.020 | 0.007 | 0.359 | 0.598 | -0.001 | -0.012 | 0.010 | 0.869 | 0.964 | -0.038 | -0.065 | -0.011 | 0.007 | 0.048 |
| PC(15-MHDA_18:2) | -0.009 | -0.031 | 0.013 | 0.413 | 0.922 | -0.013 | -0.027 | 0.001 | 0.065 | 0.338 | -0.001 | -0.012 | 0.010 | 0.889 | 0.964 | -0.031 | -0.058 | -0.004 | 0.027 | 0.111 |
| TG(52:2) [NL-16:0] | -0.008 | -0.030 | 0.014 | 0.466 | 0.922 | -0.020 | -0.033 | -0.006 | 0.006 | 0.259 | -0.001 | -0.012 | 0.010 | 0.892 | 0.964 | -0.030 | -0.058 | -0.002 | 0.036 | 0.134 |
| PC(P-35:2) (b) | -0.008 | -0.030 | 0.014 | 0.481 | 0.922 | -0.005 | -0.018 | 0.009 | 0.509 | 0.724 | -0.001 | -0.012 | 0.010 | 0.878 | 0.964 | -0.010 | -0.037 | 0.016 | 0.447 | 0.652 |
| AC(17:0) (a) | -0.006 | -0.028 | 0.016 | 0.612 | 0.950 | -0.017 | -0.031 | -0.004 | 0.012 | 0.273 | -0.001 | -0.012 | 0.010 | 0.878 | 0.964 | -0.031 | -0.057 | -0.004 | 0.025 | 0.106 |
| PE(P-18:0/18:3) | -0.006 | -0.028 | 0.017 | 0.627 | 0.954 | -0.002 | -0.016 | 0.012 | 0.768 | 0.881 | -0.001 | -0.012 | 0.010 | 0.888 | 0.964 | -0.007 | -0.035 | 0.022 | 0.651 | 0.811 |
| SM(d17:1/24:1) | -0.005 | -0.027 | 0.017 | 0.653 | 0.960 | -0.014 | -0.028 | 0.000 | 0.046 | 0.306 | -0.001 | -0.011 | 0.010 | 0.893 | 0.964 | -0.033 | -0.060 | -0.005 | 0.020 | 0.091 |
| PE(16:0_20:5) | -0.004 | -0.026 | 0.018 | 0.714 | 0.964 | -0.016 | -0.030 | -0.002 | 0.022 | 0.273 | 0.001 | -0.010 | 0.012 | 0.886 | 0.964 | -0.009 | -0.037 | 0.018 | 0.508 | 0.702 |
| TG(O-54:4) [NL-18:2] | -0.004 | -0.026 | 0.018 | 0.732 | 0.964 | 0.011 | -0.003 | 0.024 | 0.130 | 0.390 | 0.001 | -0.010 | 0.012 | 0.877 | 0.964 | 0.008 | -0.020 | 0.035 | 0.574 | 0.745 |
| FA(20:3) | -0.004 | -0.026 | 0.018 | 0.707 | 0.964 | 0.000 | -0.014 | 0.014 | 0.972 | 0.983 | -0.001 | -0.012 | 0.010 | 0.891 | 0.964 | -0.003 | -0.030 | 0.025 | 0.853 | 0.933 |
| PC(14:0_22:6) | -0.003 | -0.026 | 0.019 | 0.786 | 0.973 | -0.011 | -0.025 | 0.002 | 0.106 | 0.373 | -0.001 | -0.012 | 0.010 | 0.892 | 0.964 | -0.018 | -0.046 | 0.009 | 0.197 | 0.409 |
| PC(16:0_20:4) | 0.002 | -0.020 | 0.024 | 0.864 | 0.981 | -0.001 | -0.015 | 0.013 | 0.898 | 0.942 | -0.001 | -0.012 | 0.010 | 0.889 | 0.964 | -0.032 | -0.060 | -0.003 | 0.030 | 0.120 |
| PE(P-18:1/18:2) (a) | -0.002 | -0.025 | 0.020 | 0.839 | 0.981 | 0.005 | -0.009 | 0.019 | 0.459 | 0.682 | 0.001 | -0.010 | 0.012 | 0.887 | 0.964 | -0.014 | -0.041 | 0.014 | 0.332 | 0.550 |

| | | | | | | | | | | | | | | | | | | | | |
|---|---|---|---|---|---|---|---|---|---|---|---|---|---|---|---|---|---|---|---|---|
| LPI(18:1) [sn1] | 0.002 | -0.020 | 0.024 | 0.881 | 0.981 | -0.014 | -0.028 | 0.000 | 0.054 | 0.317 | -0.001 | -0.012 | 0.010 | 0.869 | 0.964 | 0.012 | -0.015 | 0.039 | 0.383 | 0.598 |
| TG(48:2) [NL-14:1] | -0.020 | -0.042 | 0.003 | 0.083 | 0.903 | -0.017 | -0.030 | -0.003 | 0.017 | 0.273 | -0.001 | -0.011 | 0.010 | 0.903 | 0.969 | -0.017 | -0.045 | 0.010 | 0.222 | 0.430 |
| PC(P-38:5) (a) | -0.004 | -0.026 | 0.018 | 0.727 | 0.964 | -0.003 | -0.017 | 0.011 | 0.643 | 0.809 | -0.001 | -0.012 | 0.010 | 0.902 | 0.969 | -0.025 | -0.052 | 0.002 | 0.071 | 0.200 |
| TG(56:7) [NL-22:5] | -0.005 | -0.027 | 0.017 | 0.669 | 0.964 | -0.011 | -0.025 | 0.003 | 0.113 | 0.382 | -0.001 | -0.011 | 0.010 | 0.903 | 0.969 | -0.015 | -0.043 | 0.013 | 0.288 | 0.509 |
| PC(33:1) | -0.017 | -0.039 | 0.005 | 0.139 | 0.903 | -0.016 | -0.029 | -0.002 | 0.028 | 0.285 | -0.001 | -0.012 | 0.010 | 0.906 | 0.971 | -0.039 | -0.067 | -0.012 | 0.005 | 0.044 |
| PI(15-MHDA_20:4) & PI(17:0_20:4) | -0.025 | -0.047 | -0.004 | 0.023 | 0.903 | -0.009 | -0.023 | 0.005 | 0.192 | 0.454 | -0.001 | -0.011 | 0.010 | 0.910 | 0.972 | -0.029 | -0.057 | -0.002 | 0.037 | 0.136 |
| PC(P-16:0/18:2) | -0.010 | -0.032 | 0.012 | 0.387 | 0.922 | -0.002 | -0.016 | 0.012 | 0.800 | 0.897 | 0.001 | -0.010 | 0.011 | 0.909 | 0.972 | -0.012 | -0.039 | 0.015 | 0.388 | 0.604 |
| LPC(20:3) [sn2] | -0.012 | -0.033 | 0.010 | 0.299 | 0.903 | -0.011 | -0.024 | 0.003 | 0.124 | 0.385 | -0.001 | -0.011 | 0.010 | 0.921 | 0.975 | -0.013 | -0.040 | 0.014 | 0.345 | 0.564 |
| PE(18:0_22:5) (n6) | -0.013 | -0.034 | 0.009 | 0.258 | 0.903 | -0.005 | -0.019 | 0.009 | 0.488 | 0.705 | -0.001 | -0.011 | 0.010 | 0.922 | 0.975 | 0.001 | -0.026 | 0.028 | 0.945 | 0.966 |
| TG(49:1) [NL-16:1] | -0.019 | -0.041 | 0.003 | 0.096 | 0.903 | -0.006 | -0.019 | 0.008 | 0.407 | 0.643 | -0.001 | -0.011 | 0.010 | 0.923 | 0.975 | 0.001 | -0.026 | 0.028 | 0.950 | 0.968 |
| AC(14:0)-OH | -0.006 | -0.029 | 0.016 | 0.565 | 0.933 | -0.006 | -0.020 | 0.007 | 0.366 | 0.602 | -0.001 | -0.011 | 0.010 | 0.920 | 0.975 | -0.019 | -0.046 | 0.008 | 0.171 | 0.376 |
| PC(P-35:2) (a) | -0.005 | -0.027 | 0.017 | 0.657 | 0.960 | -0.010 | -0.024 | 0.004 | 0.168 | 0.433 | 0.001 | -0.010 | 0.011 | 0.917 | 0.975 | -0.007 | -0.035 | 0.020 | 0.593 | 0.758 |
| AC(15:0) (b) | -0.004 | -0.026 | 0.018 | 0.714 | 0.964 | -0.010 | -0.023 | 0.004 | 0.163 | 0.432 | 0.001 | -0.010 | 0.011 | 0.920 | 0.975 | -0.032 | -0.059 | -0.005 | 0.019 | 0.089 |
| SM(d18:1/22:0) & SM(d16:1/24:0) | 0.004 | -0.018 | 0.026 | 0.733 | 0.964 | 0.006 | -0.008 | 0.020 | 0.395 | 0.634 | 0.001 | -0.010 | 0.011 | 0.917 | 0.975 | -0.017 | -0.043 | 0.010 | 0.229 | 0.439 |
| SM(d18:0/14:0) | 0.002 | -0.020 | 0.024 | 0.864 | 0.981 | -0.004 | -0.017 | 0.010 | 0.600 | 0.771 | -0.001 | -0.011 | 0.010 | 0.921 | 0.975 | -0.030 | -0.057 | -0.002 | 0.035 | 0.132 |
| PE(18:0_22:4) | -0.018 | -0.039 | 0.004 | 0.114 | 0.903 | -0.004 | -0.018 | 0.010 | 0.590 | 0.764 | 0.001 | -0.010 | 0.011 | 0.926 | 0.976 | -0.002 | -0.030 | 0.025 | 0.868 | 0.942 |
| PA(36:4) | -0.002 | -0.024 | 0.020 | 0.879 | 0.981 | -0.003 | -0.017 | 0.011 | 0.696 | 0.845 | -0.001 | -0.011 | 0.010 | 0.925 | 0.976 | 0.024 | -0.004 | 0.051 | 0.091 | 0.241 |
| PC(39:5) (a) | 0.009 | -0.013 | 0.032 | 0.415 | 0.922 | -0.009 | -0.023 | 0.005 | 0.195 | 0.455 | 0.000 | -0.010 | 0.011 | 0.928 | 0.976 | -0.042 | -0.070 | -0.014 | 0.004 | 0.038 |
| Cer(d17:1/24:0) | -0.005 | -0.028 | 0.017 | 0.636 | 0.960 | 0.002 | -0.012 | 0.017 | 0.735 | 0.870 | 0.000 | -0.011 | 0.010 | 0.929 | 0.976 | -0.003 | -0.031 | 0.024 | 0.821 | 0.921 |
| PC(P-36:3) | -0.007 | -0.028 | 0.015 | 0.559 | 0.933 | -0.005 | -0.019 | 0.009 | 0.479 | 0.696 | 0.000 | -0.011 | 0.010 | 0.933 | 0.979 | -0.022 | -0.049 | 0.005 | 0.111 | 0.277 |
| TG(50:2) [NL-16:1] | -0.020 | -0.041 | 0.002 | 0.079 | 0.903 | -0.017 | -0.030 | -0.003 | 0.015 | 0.273 | 0.000 | -0.011 | 0.010 | 0.939 | 0.980 | -0.027 | -0.055 | 0.001 | 0.056 | 0.173 |
| PE(18:0_20:3) (a) | -0.013 | -0.035 | 0.009 | 0.242 | 0.903 | -0.012 | -0.026 | 0.002 | 0.083 | 0.356 | 0.000 | -0.011 | 0.010 | 0.938 | 0.980 | -0.011 | -0.038 | 0.016 | 0.438 | 0.644 |
| TG(50:3) [NL-18:3] | -0.024 | -0.045 | -0.002 | 0.034 | 0.903 | -0.010 | -0.023 | 0.004 | 0.166 | 0.432 | 0.000 | -0.010 | 0.011 | 0.936 | 0.980 | 0.002 | -0.026 | 0.029 | 0.906 | 0.954 |
| PI(18:0_22:5) (n6) | -0.009 | -0.031 | 0.012 | 0.397 | 0.922 | -0.019 | -0.033 | -0.005 | 0.007 | 0.259 | 0.000 | -0.011 | 0.010 | 0.937 | 0.980 | -0.006 | -0.033 | 0.021 | 0.662 | 0.814 |
| PE(O-16:0/20:3) | -0.017 | -0.039 | 0.005 | 0.127 | 0.903 | -0.005 | -0.019 | 0.009 | 0.477 | 0.696 | 0.000 | -0.011 | 0.010 | 0.941 | 0.980 | -0.016 | -0.042 | 0.011 | 0.249 | 0.464 |
| PE(17:0_18:1) | -0.009 | -0.031 | 0.013 | 0.441 | 0.922 | -0.016 | -0.030 | -0.003 | 0.018 | 0.273 | 0.000 | -0.010 | 0.011 | 0.944 | 0.982 | -0.008 | -0.036 | 0.019 | 0.556 | 0.734 |
| TG(50:2) [NL-18:1] | -0.019 | -0.041 | 0.003 | 0.084 | 0.903 | -0.019 | -0.033 | -0.005 | 0.006 | 0.259 | 0.000 | -0.011 | 0.010 | 0.952 | 0.984 | -0.027 | -0.055 | 0.002 | 0.065 | 0.186 |
| LPC(14:0) [sn1] | -0.015 | -0.037 | 0.008 | 0.203 | 0.903 | -0.009 | -0.023 | 0.005 | 0.205 | 0.466 | 0.000 | -0.011 | 0.010 | 0.951 | 0.984 | -0.013 | -0.040 | 0.014 | 0.353 | 0.572 |
| PC(14:0_20:4) | -0.013 | -0.036 | 0.009 | 0.243 | 0.903 | 0.000 | -0.014 | 0.014 | 0.985 | 0.989 | 0.000 | -0.011 | 0.011 | 0.966 | 0.984 | -0.011 | -0.038 | 0.017 | 0.449 | 0.653 |
| PE(16:0_18:1) | -0.014 | -0.036 | 0.008 | 0.224 | 0.903 | -0.012 | -0.026 | 0.002 | 0.089 | 0.356 | 0.000 | -0.011 | 0.011 | 0.965 | 0.984 | -0.010 | -0.038 | 0.017 | 0.466 | 0.664 |
| DG(16:0_18:1) | -0.015 | -0.037 | 0.007 | 0.170 | 0.903 | -0.015 | -0.029 | -0.002 | 0.026 | 0.285 | 0.000 | -0.010 | 0.011 | 0.963 | 0.984 | -0.009 | -0.037 | 0.019 | 0.519 | 0.709 |
| DG(18:0_18:2) | -0.015 | -0.037 | 0.007 | 0.180 | 0.903 | -0.011 | -0.024 | 0.003 | 0.116 | 0.384 | 0.000 | -0.011 | 0.010 | 0.965 | 0.984 | 0.002 | -0.025 | 0.030 | 0.862 | 0.937 |
| PA(36:1) | 0.012 | -0.010 | 0.034 | 0.276 | 0.903 | -0.019 | -0.033 | -0.005 | 0.010 | 0.270 | 0.000 | -0.011 | 0.011 | 0.963 | 0.984 | -0.001 | -0.031 | 0.029 | 0.941 | 0.966 |
| PC(15-MHDA_18:1) | -0.011 | -0.033 | 0.012 | 0.353 | 0.922 | -0.019 | -0.032 | -0.005 | 0.009 | 0.268 | 0.000 | -0.011 | 0.011 | 0.963 | 0.984 | -0.039 | -0.066 | -0.011 | 0.006 | 0.048 |
| Cer(d20:1/24:0) | -0.008 | -0.030 | 0.014 | 0.498 | 0.922 | -0.004 | -0.018 | 0.010 | 0.563 | 0.748 | 0.000 | -0.011 | 0.011 | 0.962 | 0.984 | -0.012 | -0.040 | 0.015 | 0.374 | 0.592 |
| methyl-CE(18:1) | 0.009 | -0.013 | 0.032 | 0.414 | 0.922 | 0.000 | -0.014 | 0.014 | 0.970 | 0.983 | 0.000 | -0.010 | 0.011 | 0.952 | 0.984 | -0.012 | -0.039 | 0.016 | 0.414 | 0.629 |
| Cer(d16:1/24:0) | -0.007 | -0.030 | 0.015 | 0.514 | 0.922 | -0.005 | -0.019 | 0.009 | 0.519 | 0.727 | 0.000 | -0.011 | 0.011 | 0.954 | 0.984 | 0.001 | -0.026 | 0.029 | 0.918 | 0.962 |
| PI(39:6) | -0.007 | -0.029 | 0.015 | 0.551 | 0.931 | -0.015 | -0.029 | -0.002 | 0.030 | 0.289 | 0.000 | -0.010 | 0.011 | 0.954 | 0.984 | -0.014 | -0.042 | 0.014 | 0.317 | 0.537 |
| SM(d18:1/14:0) & SM(d16:1/16:0) | -0.006 | -0.028 | 0.016 | 0.596 | 0.950 | -0.003 | -0.017 | 0.011 | 0.707 | 0.849 | 0.000 | -0.011 | 0.011 | 0.957 | 0.984 | -0.027 | -0.054 | 0.001 | 0.056 | 0.173 |
| AC(20:5) | 0.004 | -0.018 | 0.026 | 0.712 | 0.964 | -0.009 | -0.023 | 0.004 | 0.189 | 0.454 | 0.000 | -0.011 | 0.011 | 0.967 | 0.984 | -0.004 | -0.031 | 0.023 | 0.762 | 0.891 |
| PC(15-MHDA_20:4) | -0.003 | -0.025 | 0.019 | 0.819 | 0.980 | -0.007 | -0.021 | 0.007 | 0.333 | 0.572 | 0.000 | -0.011 | 0.010 | 0.950 | 0.984 | -0.040 | -0.067 | -0.013 | 0.004 | 0.042 |
| PC(17:0_20:4) | -0.001 | -0.023 | 0.021 | 0.945 | 0.995 | -0.004 | -0.018 | 0.010 | 0.617 | 0.787 | 0.000 | -0.010 | 0.011 | 0.961 | 0.984 | -0.035 | -0.063 | -0.008 | 0.013 | 0.069 |
| PE(18:0_20:4) | -0.011 | -0.033 | 0.010 | 0.304 | 0.903 | -0.008 | -0.022 | 0.006 | 0.248 | 0.501 | 0.000 | -0.011 | 0.011 | 0.977 | 0.990 | -0.011 | -0.039 | 0.017 | 0.435 | 0.641 |
| PC(18:2_20:5) | 0.008 | -0.014 | 0.031 | 0.470 | 0.922 | -0.009 | -0.023 | 0.005 | 0.201 | 0.463 | 0.000 | -0.011 | 0.011 | 0.977 | 0.990 | -0.001 | -0.028 | 0.027 | 0.958 | 0.970 |
| LPI(18:2) [sn1] | 0.000 | -0.022 | 0.021 | 0.969 | 0.995 | 0.008 | -0.006 | 0.022 | 0.263 | 0.514 | 0.000 | -0.011 | 0.011 | 0.977 | 0.990 | 0.015 | -0.012 | 0.042 | 0.286 | 0.509 |
| PE(18:0_20:3) (b) | -0.018 | -0.040 | 0.004 | 0.103 | 0.903 | -0.020 | -0.034 | -0.007 | 0.004 | 0.259 | 0.000 | -0.011 | 0.011 | 0.979 | 0.990 | -0.019 | -0.047 | 0.009 | 0.177 | 0.382 |
| PC(18:0_20:4) | -0.002 | -0.024 | 0.020 | 0.861 | 0.981 | 0.003 | -0.011 | 0.017 | 0.687 | 0.840 | 0.000 | -0.011 | 0.011 | 0.979 | 0.990 | -0.023 | -0.051 | 0.005 | 0.106 | 0.271 |
| PI(18:0_22:6) | -0.009 | -0.031 | 0.013 | 0.442 | 0.922 | -0.016 | -0.030 | -0.002 | 0.026 | 0.285 | 0.000 | -0.011 | 0.011 | 0.981 | 0.990 | -0.018 | -0.046 | 0.010 | 0.200 | 0.416 |

| | | | | | | | | | | | | | | | | | | | | |
|---|---|---|---|---|---|---|---|---|---|---|---|---|---|---|---|---|---|---|---|---|
| DG(18:1_18:1) | -0.008 | -0.029 | 0.014 | 0.503 | 0.922 | -0.018 | -0.031 | -0.004 | 0.011 | 0.273 | 0.000 | -0.011 | 0.011 | 0.986 | 0.994 | -0.018 | -0.046 | 0.010 | 0.204 | 0.416 |
| PE(16:0_20:4) | -0.013 | -0.035 | 0.008 | 0.230 | 0.903 | -0.011 | -0.025 | 0.003 | 0.123 | 0.385 | 0.000 | -0.011 | 0.011 | 0.995 | 0.998 | -0.018 | -0.046 | 0.010 | 0.205 | 0.416 |
| TG(50:2) [NL-18:2] | -0.021 | -0.043 | 0.000 | 0.051 | 0.903 | -0.005 | -0.019 | 0.009 | 0.489 | 0.706 | 0.000 | -0.011 | 0.010 | 0.994 | 0.998 | 0.000 | -0.027 | 0.028 | 0.980 | 0.985 |
| PC(P-20:0/20:4) | -0.008 | -0.030 | 0.014 | 0.492 | 0.922 | -0.001 | -0.014 | 0.013 | 0.920 | 0.953 | 0.000 | -0.011 | 0.011 | 0.996 | 0.998 | -0.031 | -0.058 | -0.004 | 0.024 | 0.105 |
| PE(P-16:0/22:5) (n6) | -0.009 | -0.031 | 0.013 | 0.423 | 0.922 | 0.003 | -0.011 | 0.017 | 0.667 | 0.829 | 0.000 | -0.011 | 0.011 | 0.998 | 0.998 | 0.000 | -0.026 | 0.026 | 0.995 | 0.996 |
| Cer(d19:1/23:0) | 0.004 | -0.019 | 0.026 | 0.741 | 0.964 | -0.008 | -0.022 | 0.006 | 0.256 | 0.507 | 0.000 | -0.011 | 0.011 | 0.996 | 0.998 | -0.026 | -0.053 | 0.001 | 0.063 | 0.184 |
| PE(P-16:0/18:3) | -0.004 | -0.026 | 0.018 | 0.718 | 0.964 | 0.001 | -0.013 | 0.016 | 0.855 | 0.929 | 0.000 | -0.011 | 0.011 | 0.997 | 0.998 | 0.008 | -0.020 | 0.036 | 0.588 | 0.756 |
