## Supplementary Table 7 for "Circulating lipid profiles are associated with cross-sectional and longitudinal changes of central biomarkers for Alzheimer’s disease"

Supplementary Table 7. Associations of lipid classes with longitudinal A/T/N biomarkers

| | A: Amyloid PET (AV45) uptake | | | | | T: CSF pTau | | | | | N1: Hippocampal volume | | | | | N2: FDG uptake | | | | |
|---|---|---|---|---|---|---|---|---|---|---|---|---|---|---|---|---|---|---|---|---|
| Lipid classes | Beta | 95% CI (Lower) | 95% CI (Upper) | Pvalue | Pvalue(BH) | Beta | 95% CI (Lower) | 95% CI (Upper) | Pvalue | Pvalue(BH) | Beta | 95% CI (Lower) | 95% CI (Upper) | Pvalue | Pvalue(BH) | Beta | 95% CI (Lower) | 95% CI (Upper) | Pvalue | Pvalue(BH) |
| GM3 | -0.007 | -0.029 | 0.015 | 0.543 | 0.901 | -0.011 | -0.026 | 0.003 | 0.116 | 0.382 | -0.022 | -0.032 | -0.011 | 0.000 | 0.002 | -0.065 | -0.092 | -0.038 | 0.000 | 0.000 |
| GM1 | 0.003 | -0.020 | 0.025 | 0.823 | 0.901 | -0.018 | -0.032 | -0.003 | 0.015 | 0.221 | -0.015 | -0.026 | -0.005 | 0.005 | 0.063 | -0.060 | -0.089 | -0.032 | 0.000 | 0.001 |
| deDE | -0.001 | -0.024 | 0.022 | 0.929 | 0.929 | 0.000 | -0.014 | 0.015 | 0.952 | 0.973 | -0.027 | -0.038 | -0.016 | 0.000 | 0.000 | -0.057 | -0.086 | -0.029 | 0.000 | 0.001 |
| Hex3Cer | -0.005 | -0.027 | 0.017 | 0.681 | 0.901 | -0.008 | -0.022 | 0.006 | 0.261 | 0.512 | -0.013 | -0.024 | -0.002 | 0.021 | 0.138 | -0.050 | -0.077 | -0.023 | 0.000 | 0.004 |
| PC | -0.011 | -0.033 | 0.012 | 0.352 | 0.901 | -0.013 | -0.027 | 0.001 | 0.063 | 0.241 | -0.006 | -0.017 | 0.005 | 0.296 | 0.524 | -0.045 | -0.072 | -0.018 | 0.001 | 0.011 |
| AC | -0.003 | -0.025 | 0.019 | 0.798 | 0.901 | -0.014 | -0.028 | -0.001 | 0.041 | 0.224 | -0.013 | -0.023 | -0.002 | 0.020 | 0.138 | -0.044 | -0.070 | -0.017 | 0.002 | 0.012 |
| SM | -0.003 | -0.025 | 0.019 | 0.763 | 0.901 | -0.006 | -0.019 | 0.008 | 0.437 | 0.669 | -0.004 | -0.015 | 0.007 | 0.466 | 0.650 | -0.042 | -0.069 | -0.016 | 0.002 | 0.013 |
| LPC(O) | -0.010 | -0.032 | 0.012 | 0.365 | 0.901 | -0.020 | -0.033 | -0.006 | 0.006 | 0.221 | -0.019 | -0.029 | -0.008 | 0.001 | 0.011 | -0.041 | -0.068 | -0.013 | 0.004 | 0.019 |
| COH | -0.005 | -0.027 | 0.017 | 0.644 | 0.901 | -0.003 | -0.017 | 0.011 | 0.666 | 0.901 | -0.005 | -0.016 | 0.006 | 0.332 | 0.566 | -0.040 | -0.067 | -0.013 | 0.004 | 0.019 |
| CE | -0.002 | -0.024 | 0.021 | 0.895 | 0.919 | -0.002 | -0.015 | 0.012 | 0.798 | 0.973 | -0.008 | -0.019 | 0.003 | 0.139 | 0.343 | -0.039 | -0.066 | -0.012 | 0.005 | 0.025 |
| PC(O) | -0.010 | -0.032 | 0.012 | 0.360 | 0.901 | -0.008 | -0.022 | 0.006 | 0.274 | 0.512 | -0.005 | -0.015 | 0.006 | 0.408 | 0.608 | -0.036 | -0.062 | -0.009 | 0.009 | 0.038 |
| PIP1 | -0.006 | -0.028 | 0.016 | 0.601 | 0.901 | -0.007 | -0.021 | 0.006 | 0.290 | 0.512 | -0.008 | -0.018 | 0.003 | 0.159 | 0.366 | -0.035 | -0.062 | -0.008 | 0.011 | 0.040 |
| Cer(d) | -0.011 | -0.033 | 0.011 | 0.342 | 0.901 | -0.011 | -0.025 | 0.003 | 0.134 | 0.412 | -0.007 | -0.018 | 0.004 | 0.208 | 0.415 | -0.034 | -0.061 | -0.006 | 0.016 | 0.058 |
| Hex2Cer | -0.002 | -0.024 | 0.020 | 0.877 | 0.919 | -0.001 | -0.015 | 0.013 | 0.881 | 0.973 | -0.012 | -0.023 | -0.002 | 0.024 | 0.138 | -0.032 | -0.059 | -0.005 | 0.022 | 0.068 |
| AC-OH | -0.003 | -0.025 | 0.019 | 0.802 | 0.901 | -0.007 | -0.021 | 0.006 | 0.300 | 0.512 | -0.005 | -0.015 | 0.006 | 0.409 | 0.608 | -0.032 | -0.060 | -0.005 | 0.022 | 0.068 |
| HexCer | 0.008 | -0.014 | 0.030 | 0.467 | 0.901 | -0.008 | -0.022 | 0.005 | 0.230 | 0.495 | -0.013 | -0.024 | -0.003 | 0.016 | 0.138 | -0.031 | -0.058 | -0.003 | 0.028 | 0.079 |
| LPE | -0.020 | -0.042 | 0.002 | 0.078 | 0.901 | -0.016 | -0.029 | -0.002 | 0.024 | 0.221 | -0.012 | -0.023 | -0.001 | 0.030 | 0.153 | -0.030 | -0.058 | -0.003 | 0.029 | 0.079 |
| PI | -0.020 | -0.042 | 0.002 | 0.077 | 0.901 | -0.012 | -0.026 | 0.002 | 0.099 | 0.351 | -0.005 | -0.016 | 0.006 | 0.354 | 0.582 | -0.029 | -0.056 | -0.002 | 0.035 | 0.088 |
| LPC | -0.007 | -0.029 | 0.015 | 0.557 | 0.901 | -0.014 | -0.027 | 0.000 | 0.050 | 0.232 | -0.011 | -0.022 | -0.001 | 0.041 | 0.170 | -0.028 | -0.055 | 0.000 | 0.048 | 0.114 |
| SHexCer | -0.005 | -0.027 | 0.018 | 0.679 | 0.901 | -0.007 | -0.021 | 0.007 | 0.346 | 0.562 | -0.007 | -0.018 | 0.004 | 0.200 | 0.415 | -0.028 | -0.056 | 0.000 | 0.050 | 0.114 |
| LPE(P) | -0.004 | -0.026 | 0.018 | 0.717 | 0.901 | -0.007 | -0.021 | 0.008 | 0.355 | 0.562 | -0.008 | -0.019 | 0.003 | 0.139 | 0.343 | -0.026 | -0.053 | 0.002 | 0.067 | 0.147 |
| C1P | -0.004 | -0.025 | 0.018 | 0.747 | 0.901 | -0.001 | -0.015 | 0.013 | 0.898 | 0.973 | -0.010 | -0.021 | 0.000 | 0.061 | 0.216 | -0.025 | -0.053 | 0.003 | 0.078 | 0.163 |
| LPC(P) | -0.009 | -0.031 | 0.013 | 0.424 | 0.901 | -0.010 | -0.024 | 0.004 | 0.162 | 0.467 | -0.011 | -0.022 | 0.000 | 0.048 | 0.182 | -0.024 | -0.051 | 0.003 | 0.087 | 0.174 |
| S1P | -0.007 | -0.029 | 0.015 | 0.514 | 0.901 | -0.008 | -0.022 | 0.006 | 0.237 | 0.495 | -0.008 | -0.019 | 0.003 | 0.142 | 0.343 | -0.023 | -0.050 | 0.004 | 0.098 | 0.187 |
| PC(P) | -0.007 | -0.029 | 0.015 | 0.534 | 0.901 | -0.008 | -0.022 | 0.006 | 0.280 | 0.512 | 0.001 | -0.010 | 0.012 | 0.852 | 0.945 | -0.020 | -0.047 | 0.007 | 0.152 | 0.279 |
| DE | 0.013 | -0.009 | 0.035 | 0.240 | 0.901 | 0.009 | -0.004 | 0.023 | 0.178 | 0.482 | -0.009 | -0.020 | 0.002 | 0.094 | 0.281 | -0.020 | -0.048 | 0.008 | 0.159 | 0.282 |
| TG(O) [NL] | -0.012 | -0.034 | 0.009 | 0.264 | 0.901 | -0.004 | -0.018 | 0.009 | 0.544 | 0.807 | 0.010 | -0.001 | 0.020 | 0.079 | 0.260 | 0.017 | -0.011 | 0.044 | 0.240 | 0.409 |
| OxSpecies | -0.005 | -0.027 | 0.018 | 0.692 | 0.901 | -0.009 | -0.023 | 0.005 | 0.203 | 0.491 | -0.009 | -0.020 | 0.002 | 0.098 | 0.281 | -0.016 | -0.044 | 0.012 | 0.250 | 0.411 |
| PE | -0.016 | -0.037 | 0.006 | 0.166 | 0.901 | -0.015 | -0.029 | -0.001 | 0.032 | 0.224 | -0.003 | -0.014 | 0.008 | 0.582 | 0.765 | -0.015 | -0.043 | 0.012 | 0.282 | 0.448 |
| Sph | 0.011 | -0.011 | 0.033 | 0.317 | 0.901 | -0.009 | -0.022 | 0.005 | 0.229 | 0.495 | 0.001 | -0.010 | 0.012 | 0.791 | 0.933 | 0.014 | -0.013 | 0.041 | 0.308 | 0.472 |
| PE(P) | -0.007 | -0.029 | 0.015 | 0.528 | 0.901 | 0.000 | -0.015 | 0.014 | 0.951 | 0.973 | 0.007 | -0.004 | 0.018 | 0.201 | 0.415 | -0.013 | -0.041 | 0.014 | 0.329 | 0.474 |
| dimethyl-CE | 0.013 | -0.009 | 0.036 | 0.241 | 0.901 | 0.001 | -0.013 | 0.015 | 0.923 | 0.973 | 0.000 | -0.011 | 0.011 | 0.981 | 0.981 | -0.014 | -0.041 | 0.014 | 0.327 | 0.474 |
| methyl-CE | 0.012 | -0.010 | 0.035 | 0.279 | 0.901 | 0.003 | -0.011 | 0.017 | 0.646 | 0.900 | -0.002 | -0.012 | 0.009 | 0.763 | 0.923 | -0.012 | -0.040 | 0.016 | 0.392 | 0.547 |
| Ubiquinone | 0.007 | -0.015 | 0.029 | 0.511 | 0.901 | -0.004 | -0.018 | 0.010 | 0.577 | 0.830 | -0.004 | -0.015 | 0.006 | 0.431 | 0.620 | -0.010 | -0.038 | 0.017 | 0.464 | 0.628 |
| TG [NL] | -0.019 | -0.041 | 0.002 | 0.083 | 0.901 | -0.016 | -0.029 | -0.002 | 0.023 | 0.221 | 0.001 | -0.010 | 0.011 | 0.896 | 0.958 | -0.009 | -0.037 | 0.019 | 0.539 | 0.708 |
| DG | -0.015 | -0.037 | 0.007 | 0.188 | 0.901 | -0.013 | -0.027 | 0.000 | 0.059 | 0.241 | 0.001 | -0.009 | 0.012 | 0.829 | 0.945 | -0.008 | -0.035 | 0.020 | 0.592 | 0.757 |
| PE(O) | -0.006 | -0.029 | 0.016 | 0.571 | 0.901 | 0.002 | -0.012 | 0.016 | 0.754 | 0.964 | 0.006 | -0.004 | 0.017 | 0.237 | 0.454 | -0.007 | -0.034 | 0.020 | 0.625 | 0.777 |
| Cer(m) | -0.018 | -0.040 | 0.004 | 0.102 | 0.901 | -0.009 | -0.023 | 0.005 | 0.199 | 0.491 | 0.005 | -0.006 | 0.015 | 0.401 | 0.608 | -0.006 | -0.034 | 0.021 | 0.646 | 0.782 |
| FFA | -0.001 | -0.024 | 0.021 | 0.899 | 0.919 | 0.000 | -0.014 | 0.014 | 1.000 | 1.000 | -0.002 | -0.013 | 0.009 | 0.762 | 0.923 | -0.006 | -0.033 | 0.022 | 0.675 | 0.797 |
| dhCer | -0.009 | -0.032 | 0.013 | 0.403 | 0.901 | 0.002 | -0.011 | 0.016 | 0.753 | 0.964 | 0.011 | 0.001 | 0.022 | 0.038 | 0.170 | -0.003 | -0.031 | 0.024 | 0.806 | 0.904 |
| BA | -0.008 | -0.031 | 0.014 | 0.461 | 0.901 | 0.002 | -0.012 | 0.015 | 0.820 | 0.973 | -0.004 | -0.015 | 0.007 | 0.497 | 0.672 | -0.003 | -0.031 | 0.024 | 0.814 | 0.904 |
| PS | 0.003 | -0.019 | 0.025 | 0.796 | 0.901 | -0.015 | -0.028 | -0.001 | 0.035 | 0.224 | -0.003 | -0.014 | 0.008 | 0.614 | 0.785 | -0.003 | -0.031 | 0.024 | 0.826 | 0.904 |
| PA | -0.003 | -0.025 | 0.019 | 0.809 | 0.901 | -0.014 | -0.028 | 0.000 | 0.044 | 0.224 | -0.006 | -0.017 | 0.005 | 0.258 | 0.474 | 0.002 | -0.025 | 0.030 | 0.860 | 0.920 |
| LPI | -0.003 | -0.025 | 0.018 | 0.759 | 0.901 | -0.001 | -0.016 | 0.013 | 0.843 | 0.973 | 0.000 | -0.011 | 0.010 | 0.943 | 0.981 | 0.002 | -0.025 | 0.029 | 0.890 | 0.921 |
| methyl-DE | 0.016 | -0.006 | 0.039 | 0.151 | 0.901 | 0.001 | -0.013 | 0.015 | 0.852 | 0.973 | 0.000 | -0.011 | 0.011 | 0.965 | 0.981 | 0.002 | -0.026 | 0.029 | 0.901 | 0.921 |
| PG | -0.020 | -0.042 | 0.002 | 0.080 | 0.901 | -0.016 | -0.029 | -0.002 | 0.021 | 0.221 | -0.001 | -0.012 | 0.010 | 0.863 | 0.945 | -0.001 | -0.029 | 0.027 | 0.956 | 0.956 |
