## Supplementary Table 8 for "Circulating lipid profiles are associated with cross-sectional and longitudinal changes of central biomarkers for Alzheimer’s disease"

**Supplementary Table 8. Associations of lipid correlation network modules with longitudinal A/T/N biomarkers**

| | A: Amyloid PET (AV45) uptake | | | | | T: CSF pTau | | | | | N1: Hippocampal volume | | | | | N2: FDG uptake | | | | |
|---|---|---|---|---|---|---|---|---|---|---|---|---|---|---|---|---|---|---|---|---|
| Lipid modules | Beta | 95% CI (Lower) | 95% CI (Upper) | Pvalue | Pvalue(BH) | Beta | 95% CI (Lower) | 95% CI (Upper) | Pvalue | Pvalue(BH) | Beta | 95% CI (Lower) | 95% CI (Upper) | Pvalue | Pvalue(BH) | Beta | 95% CI (Lower) | 95% CI (Upper) | Pvalue | Pvalue(BH) |
| M23 | -0.010 | -0.032 | 0.012 | 0.378 | 0.900 | -0.004 | -0.018 | 0.009 | 0.536 | 0.782 | -0.011 | -0.022 | 0.000 | 0.041 | 0.147 | -0.053 | -0.079 | -0.026 | 0.000 | 0.005 |
| M4 | -0.007 | -0.029 | 0.015 | 0.557 | 0.900 | -0.015 | -0.029 | -0.001 | 0.030 | 0.270 | -0.011 | -0.021 | 0.000 | 0.055 | 0.168 | -0.049 | -0.076 | -0.021 | 0.001 | 0.012 |
| M42 | -0.009 | -0.031 | 0.013 | 0.417 | 0.900 | -0.011 | -0.025 | 0.003 | 0.129 | 0.340 | -0.011 | -0.022 | 0.000 | 0.041 | 0.147 | -0.045 | -0.071 | -0.018 | 0.001 | 0.014 |
| M44 | -0.017 | -0.039 | 0.004 | 0.119 | 0.900 | -0.013 | -0.027 | 0.001 | 0.061 | 0.309 | -0.011 | -0.022 | 0.000 | 0.048 | 0.159 | -0.046 | -0.073 | -0.018 | 0.001 | 0.014 |
| M39 | -0.010 | -0.032 | 0.012 | 0.387 | 0.900 | -0.021 | -0.034 | -0.007 | 0.004 | 0.164 | -0.019 | -0.030 | -0.008 | 0.001 | 0.008 | -0.042 | -0.070 | -0.015 | 0.002 | 0.014 |
| M26 | -0.001 | -0.024 | 0.021 | 0.894 | 0.935 | -0.008 | -0.022 | 0.005 | 0.233 | 0.465 | -0.010 | -0.020 | 0.001 | 0.074 | 0.213 | -0.044 | -0.071 | -0.016 | 0.002 | 0.014 |
| M25 | -0.006 | -0.028 | 0.016 | 0.578 | 0.900 | -0.016 | -0.030 | -0.002 | 0.024 | 0.270 | -0.008 | -0.019 | 0.003 | 0.150 | 0.346 | -0.042 | -0.069 | -0.015 | 0.002 | 0.014 |
| M41 | -0.009 | -0.031 | 0.013 | 0.428 | 0.900 | -0.010 | -0.023 | 0.004 | 0.171 | 0.414 | -0.002 | -0.013 | 0.009 | 0.714 | 0.906 | -0.043 | -0.070 | -0.015 | 0.002 | 0.014 |
| M18 | 0.005 | -0.018 | 0.027 | 0.688 | 0.935 | 0.003 | -0.010 | 0.017 | 0.627 | 0.849 | -0.019 | -0.030 | -0.009 | 0.000 | 0.008 | -0.037 | -0.064 | -0.009 | 0.009 | 0.040 |
| M38 | -0.001 | -0.024 | 0.021 | 0.901 | 0.935 | -0.003 | -0.017 | 0.011 | 0.696 | 0.873 | -0.013 | -0.024 | -0.002 | 0.016 | 0.113 | -0.036 | -0.064 | -0.009 | 0.010 | 0.040 |
| M28 | 0.003 | -0.019 | 0.025 | 0.792 | 0.935 | -0.008 | -0.022 | 0.006 | 0.276 | 0.497 | 0.002 | -0.009 | 0.012 | 0.760 | 0.906 | -0.037 | -0.064 | -0.009 | 0.009 | 0.040 |
| M46 | -0.005 | -0.028 | 0.017 | 0.627 | 0.931 | -0.013 | -0.026 | 0.001 | 0.068 | 0.312 | -0.001 | -0.012 | 0.010 | 0.863 | 0.906 | -0.035 | -0.063 | -0.008 | 0.011 | 0.041 |
| M11 | -0.010 | -0.032 | 0.012 | 0.380 | 0.900 | -0.012 | -0.025 | 0.002 | 0.091 | 0.340 | -0.020 | -0.031 | -0.009 | 0.000 | 0.008 | -0.032 | -0.059 | -0.006 | 0.018 | 0.060 |
| M14 | -0.013 | -0.035 | 0.009 | 0.243 | 0.900 | -0.002 | -0.016 | 0.012 | 0.804 | 0.902 | -0.016 | -0.026 | -0.005 | 0.005 | 0.056 | -0.032 | -0.059 | -0.006 | 0.018 | 0.060 |
| M6 | -0.004 | -0.026 | 0.018 | 0.723 | 0.935 | -0.001 | -0.015 | 0.013 | 0.863 | 0.922 | -0.003 | -0.014 | 0.008 | 0.581 | 0.835 | -0.033 | -0.061 | -0.005 | 0.020 | 0.063 |
| M19 | 0.008 | -0.014 | 0.030 | 0.467 | 0.900 | -0.008 | -0.022 | 0.005 | 0.230 | 0.465 | -0.013 | -0.024 | -0.003 | 0.016 | 0.113 | -0.031 | -0.058 | -0.003 | 0.028 | 0.075 |
| M35 | -0.007 | -0.029 | 0.015 | 0.531 | 0.900 | -0.002 | -0.016 | 0.012 | 0.761 | 0.898 | -0.001 | -0.012 | 0.010 | 0.867 | 0.906 | -0.030 | -0.057 | -0.004 | 0.027 | 0.075 |
| M3 | -0.008 | -0.030 | 0.014 | 0.485 | 0.900 | -0.015 | -0.028 | -0.001 | 0.038 | 0.270 | -0.012 | -0.023 | -0.001 | 0.031 | 0.144 | -0.030 | -0.057 | -0.003 | 0.030 | 0.077 |
| M31 | -0.010 | -0.032 | 0.011 | 0.351 | 0.900 | -0.009 | -0.023 | 0.004 | 0.188 | 0.433 | -0.004 | -0.015 | 0.007 | 0.456 | 0.724 | -0.030 | -0.057 | -0.003 | 0.032 | 0.079 |
| M30 | -0.020 | -0.042 | 0.003 | 0.083 | 0.900 | -0.011 | -0.025 | 0.003 | 0.119 | 0.340 | -0.002 | -0.013 | 0.009 | 0.749 | 0.906 | -0.027 | -0.054 | 0.001 | 0.055 | 0.125 |
| M17 | -0.002 | -0.025 | 0.020 | 0.831 | 0.935 | -0.008 | -0.022 | 0.006 | 0.278 | 0.497 | 0.000 | -0.010 | 0.011 | 0.936 | 0.936 | -0.027 | -0.054 | 0.001 | 0.057 | 0.125 |
| M21 | -0.015 | -0.037 | 0.006 | 0.163 | 0.900 | -0.006 | -0.020 | 0.008 | 0.412 | 0.654 | -0.007 | -0.018 | 0.004 | 0.192 | 0.420 | -0.026 | -0.053 | 0.001 | 0.060 | 0.126 |
| M45 | 0.001 | -0.021 | 0.024 | 0.896 | 0.935 | -0.011 | -0.026 | 0.003 | 0.120 | 0.340 | 0.002 | -0.009 | 0.013 | 0.743 | 0.906 | -0.025 | -0.053 | 0.003 | 0.077 | 0.154 |
| M13 | -0.002 | -0.024 | 0.021 | 0.876 | 0.935 | 0.003 | -0.011 | 0.016 | 0.708 | 0.873 | -0.005 | -0.016 | 0.006 | 0.359 | 0.636 | -0.023 | -0.050 | 0.004 | 0.089 | 0.171 |
| M9 | 0.001 | -0.021 | 0.023 | 0.914 | 0.935 | 0.003 | -0.011 | 0.016 | 0.721 | 0.873 | -0.001 | -0.012 | 0.010 | 0.824 | 0.906 | -0.022 | -0.049 | 0.005 | 0.109 | 0.201 |
| M36 | -0.007 | -0.029 | 0.014 | 0.509 | 0.900 | -0.011 | -0.025 | 0.003 | 0.116 | 0.340 | -0.009 | -0.020 | 0.001 | 0.088 | 0.226 | -0.020 | -0.048 | 0.008 | 0.155 | 0.274 |
| M32 | -0.009 | -0.031 | 0.013 | 0.430 | 0.900 | -0.008 | -0.022 | 0.006 | 0.281 | 0.497 | -0.006 | -0.017 | 0.005 | 0.291 | 0.558 | -0.020 | -0.047 | 0.008 | 0.163 | 0.278 |
| M15 | -0.008 | -0.030 | 0.014 | 0.481 | 0.900 | 0.001 | -0.013 | 0.015 | 0.857 | 0.922 | 0.003 | -0.008 | 0.014 | 0.546 | 0.835 | -0.017 | -0.045 | 0.010 | 0.207 | 0.340 |
| M43 | -0.013 | -0.035 | 0.009 | 0.258 | 0.900 | 0.005 | -0.008 | 0.019 | 0.448 | 0.688 | -0.004 | -0.015 | 0.007 | 0.442 | 0.724 | -0.016 | -0.043 | 0.010 | 0.227 | 0.359 |
| M16 | -0.012 | -0.034 | 0.009 | 0.264 | 0.900 | -0.004 | -0.018 | 0.009 | 0.544 | 0.782 | 0.010 | -0.001 | 0.020 | 0.079 | 0.214 | 0.017 | -0.011 | 0.044 | 0.240 | 0.368 |
| M40 | -0.003 | -0.025 | 0.019 | 0.789 | 0.935 | -0.011 | -0.025 | 0.003 | 0.113 | 0.340 | -0.007 | -0.018 | 0.004 | 0.213 | 0.446 | -0.014 | -0.042 | 0.013 | 0.300 | 0.435 |
| M1 | -0.016 | -0.038 | 0.006 | 0.143 | 0.900 | -0.014 | -0.028 | 0.000 | 0.045 | 0.270 | -0.003 | -0.014 | 0.008 | 0.572 | 0.835 | -0.015 | -0.042 | 0.013 | 0.302 | 0.435 |
| M22 | -0.004 | -0.026 | 0.018 | 0.711 | 0.935 | -0.014 | -0.028 | -0.001 | 0.041 | 0.270 | -0.001 | -0.012 | 0.010 | 0.842 | 0.906 | -0.014 | -0.042 | 0.014 | 0.330 | 0.461 |
| M34 | -0.014 | -0.037 | 0.008 | 0.202 | 0.900 | -0.010 | -0.024 | 0.003 | 0.133 | 0.340 | -0.011 | -0.022 | 0.000 | 0.042 | 0.147 | -0.013 | -0.040 | 0.015 | 0.356 | 0.482 |
| M24 | 0.007 | -0.016 | 0.029 | 0.563 | 0.900 | -0.011 | -0.025 | 0.003 | 0.125 | 0.340 | 0.005 | -0.006 | 0.016 | 0.341 | 0.627 | -0.010 | -0.038 | 0.018 | 0.477 | 0.610 |
| M12 | 0.014 | -0.008 | 0.037 | 0.212 | 0.900 | 0.002 | -0.012 | 0.016 | 0.784 | 0.901 | -0.001 | -0.012 | 0.010 | 0.899 | 0.919 | -0.010 | -0.038 | 0.017 | 0.467 | 0.610 |
| M10 | -0.012 | -0.034 | 0.010 | 0.291 | 0.900 | -0.006 | -0.020 | 0.008 | 0.395 | 0.649 | 0.013 | 0.002 | 0.023 | 0.020 | 0.113 | 0.008 | -0.019 | 0.035 | 0.544 | 0.676 |
| M27 | -0.018 | -0.040 | 0.004 | 0.102 | 0.900 | -0.009 | -0.023 | 0.005 | 0.199 | 0.436 | 0.005 | -0.006 | 0.015 | 0.401 | 0.684 | -0.006 | -0.034 | 0.021 | 0.646 | 0.747 |
| M2 | -0.021 | -0.043 | 0.001 | 0.057 | 0.900 | -0.015 | -0.029 | -0.002 | 0.026 | 0.270 | 0.001 | -0.010 | 0.012 | 0.843 | 0.906 | -0.007 | -0.035 | 0.021 | 0.617 | 0.747 |
| M5 | -0.002 | -0.024 | 0.020 | 0.852 | 0.935 | 0.001 | -0.013 | 0.015 | 0.894 | 0.922 | -0.002 | -0.013 | 0.009 | 0.711 | 0.906 | -0.006 | -0.034 | 0.021 | 0.649 | 0.747 |
| M7 | 0.012 | -0.010 | 0.034 | 0.296 | 0.900 | 0.001 | -0.013 | 0.015 | 0.902 | 0.922 | -0.008 | -0.019 | 0.003 | 0.133 | 0.323 | -0.005 | -0.032 | 0.023 | 0.748 | 0.839 |
| M33 | 0.001 | -0.021 | 0.024 | 0.909 | 0.935 | -0.014 | -0.028 | 0.000 | 0.047 | 0.270 | 0.007 | -0.004 | 0.018 | 0.229 | 0.458 | 0.003 | -0.024 | 0.031 | 0.808 | 0.871 |
| M29 | -0.017 | -0.039 | 0.005 | 0.125 | 0.900 | -0.007 | -0.020 | 0.007 | 0.341 | 0.582 | 0.002 | -0.009 | 0.012 | 0.755 | 0.906 | -0.003 | -0.031 | 0.025 | 0.831 | 0.871 |
| M20 | -0.001 | -0.023 | 0.021 | 0.946 | 0.946 | -0.001 | -0.015 | 0.013 | 0.937 | 0.937 | -0.001 | -0.012 | 0.010 | 0.864 | 0.906 | 0.003 | -0.024 | 0.030 | 0.833 | 0.871 |
| M37 | -0.003 | -0.025 | 0.019 | 0.794 | 0.935 | 0.004 | -0.010 | 0.017 | 0.610 | 0.849 | 0.013 | 0.002 | 0.024 | 0.018 | 0.113 | -0.001 | -0.028 | 0.026 | 0.929 | 0.949 |
| M8 | -0.006 | -0.028 | 0.016 | 0.587 | 0.900 | 0.003 | -0.011 | 0.017 | 0.651 | 0.856 | 0.012 | 0.002 | 0.023 | 0.024 | 0.121 | 0.000 | -0.028 | 0.027 | 0.984 | 0.984 |
